# Mitigation of COVID-19 using social distancing of the elderly in Brazil: The vertical quarantine effects in hospitalizations and deaths

**DOI:** 10.1101/2021.01.12.21249495

**Authors:** Vito Ribeiro Venturieri, Matheus Silva Gonçalves, Vinícius Rios Fuck

## Abstract

Governments and epidemiologists have been proposing several mitigation strategies based on non-pharmaceutical interventions to reduce COVID-19 cases, hospitalizations, and deaths. In this work, we quantitatively compare the effects of elderly population (60 years old or more) selective isolation with a no isolation scenario using an adapted Susceptible - Exposed - Infectious - Removed (SEIR) compartmental model. For these simulated scenarios, we estimate the number of hospitalizations and deaths for different Brazilian cities, including those due to the lack of hospital beds. Our simulations show that, for São Paulo City, the isolation of the elderly would reduce demand for hospital beds by 9% and deaths by 16% compared to the no intervention scenario. Other Brazilian cities follow the same pattern, with median reductions of deaths ranging from 12-18%. We conclude that the social distancing of the elderly would be marginally effective and would not avoid health system collapse in several Brazilian cities.

## 1. Introduction

A novel coronavirus, named SARS-CoV-2 [1], was identified in Wuhan, China, in late 2019 [2]. The COVID-19 disease caused by this virus can result several health complications, including acute respiratory distress syndrome and death [3].

The high dispersion power combined with a high proportion of patients needing critical care when infected by the SARS-COV-2 are challenging health systems worldwide. In order to control the virus spreading, several countries adopted social distancing, quarantine and isolation policies, such as the closure of stores, schools and universities, public transport reduction and prohibition of the mass gathering events [4]. Many studies endorse these policies as an effective way to combat the disease, attributing a reduction in the effective reproduction number and the total reported number of deaths [4, 5, 6, 7, 8]. For instance, Flaxman *et al*. [4] indicated that thousands of deaths were averted in Europe due to non-pharmaceutical interventions and Dehning et al. [5] noticed a clear relationship between governmental interventions and disease spreading in Germany.

In Brazil, one strategy raised by the government suggested isolating only the risk group (e.g. elderly people). This strategy was named “Isolamento vertical” [9], or “Quarentena Vertical” [10] which means vertical isolation or vertical quarantine. This strategy was formulated based on the hypothesis that isolating only the risk group would reduce the deaths and hospitalisation up to avoid overcrowding in the health system. It was hypothesised that such intervention could reduce some of the expected economic impacts due to broader isolation.

Additionally, on 4 October 2020, various scientists signed a declaration called “Great Barrington Declaration” [11]. The document recommends a focused protection of the elderly and people from other risk groups (e.g. chronic heart failure) in order to reduce the potential social and economic harms of more strict measures to mitigate COVID-19. Although based on some logic (protecting those under higher risk), this declaration does not cite any scientific paper quantifying the proposed approach’s possible effects.

Guiding scientific studies can influence millions of lives by supporting government bodies in COVID-19 related actions. At the current stage of the pandemic, many authors have modeled the effect of non-pharmaceutical measures to help decision-makers to reduce disease spreading [12, 5, 8, 13, 6]. In order to evaluate the consequences of only isolating the elderly group in Brazil, we present an adapted susceptible - exposed - infectious - removed (SEIR) model, by separating those aged 60 or more, which represent the risk group, and the remaining population. The model includes estimations for ward and intensive care unit (ICU) bed demands for COVID-19 patients, and estimates death counts for COVID-19, accounting for age-specific infection fatalities and the loss of lives due to overwhelmed health systems. Intending to provide a clear comparison threshold, we also evaluate the counterfactual no intervention case. By comparing these two distinct scenarios, we aim to effectively assess the suitability of the vertical isolation strategy for reducing the load on health systems and prevent deaths, providing valuable information for decision-makers.

## 2. Methods

### Formulation

In order to model the Brazilian epidemic growth and the respective health system load, we employed a two-step approach. In the first phase, we used a classical SEIR model [14] to model the transmission within the population. Then, we obtained the required beds (ward and ICU) and the number of deaths as a function of newly infected individuals at each time step. Compartmental models such as the proposed in the present work were widely applied to model COVID-19 disease on large populations, mainly to assess the effect of non-pharmaceutical interventions in controlling the disease [15, 16, 8, 5]. The classical SEIR compartments utilized for modeling the disease dynamic are:

- *S*: susceptible individuals,
- *E*: exposed (infected but not yet capable of infecting other people),
- *I*: infectious individuals,
- *R*: removed individuals (not able to infect or be infected anymore).

Our model subdivided the compartments into two groups to distinguish transmission rates for the elderly and young individuals. Throughout the text the subscript ()_*y*_ represents persons aged up to 59 years old (young) and the subscript ()_*e*_ indicates those aged 60 and over (elderly). Thus, this results in 8 compartments, represented by the following ordinary differential equations (ODEs):

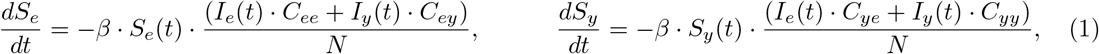

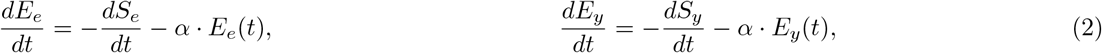

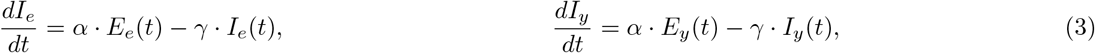

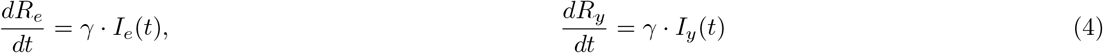

where *β, γ* and *α* stand for the transmission, recovery and incubation rates, respectively. For the whole population, we assume constant values for such rates. Besides, *N* is the total number of individuals and *C*_*ij*_ represents the contact rate between age groups *i* and *j*, where *i* and *j* can be assigned to the index *e* or *y*, as previously discussed. It is worth mentioning that we kept the population size constant throughout the process. Such hypothesis was made for convenience and can hold due to a balanced population dynamics for relatively short time lengths [17].

The parameters *γ* and *α* are calculated as [18]:

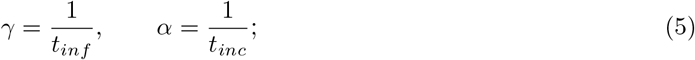

where *t*_*inf*_ and *t*_*inc*_ are the infectious and the incubation periods respectively.

We also modeled the health care system demand for ICU and ward beds for each age group and the number of deaths. During the simulation, a fraction of newly infected individuals is assigned to different outcomes (need for ICU or ward; and death or survival). In order to model the time interval between being infected and requiring hospital care, intermediate compartments were created. Furthermore, to enhance the death count estimation, the rate of individuals entering the ward and ICU compartments was divided accordingly to the outcome.

The previously exposed model is presented in a flowchart in Figure 1, depicting a general framework for an age group *i* (e.g. by switching the subscripts *i* by *e* or *y* we have the diagram for the elderly or young population respectively). It can be noticed that the infection dynamics is determined by a straightforward age-structured-SEIR embedded in a larger model. Strictly speaking, the number of newly infected individuals by age group, obtained from the SEIR model, are the inputs for the process of estimating the health care system demand and deaths. The SEIR model does not depend on the larger model (e.g. hospitalizations and deaths might be already counted as removed individuals in the SEIR model). Thus, the sum of all individuals in the *S, E, I* and *R* compartments will always be equal to *N*. The additional employed compartments are defined as:

**Figure 1:**
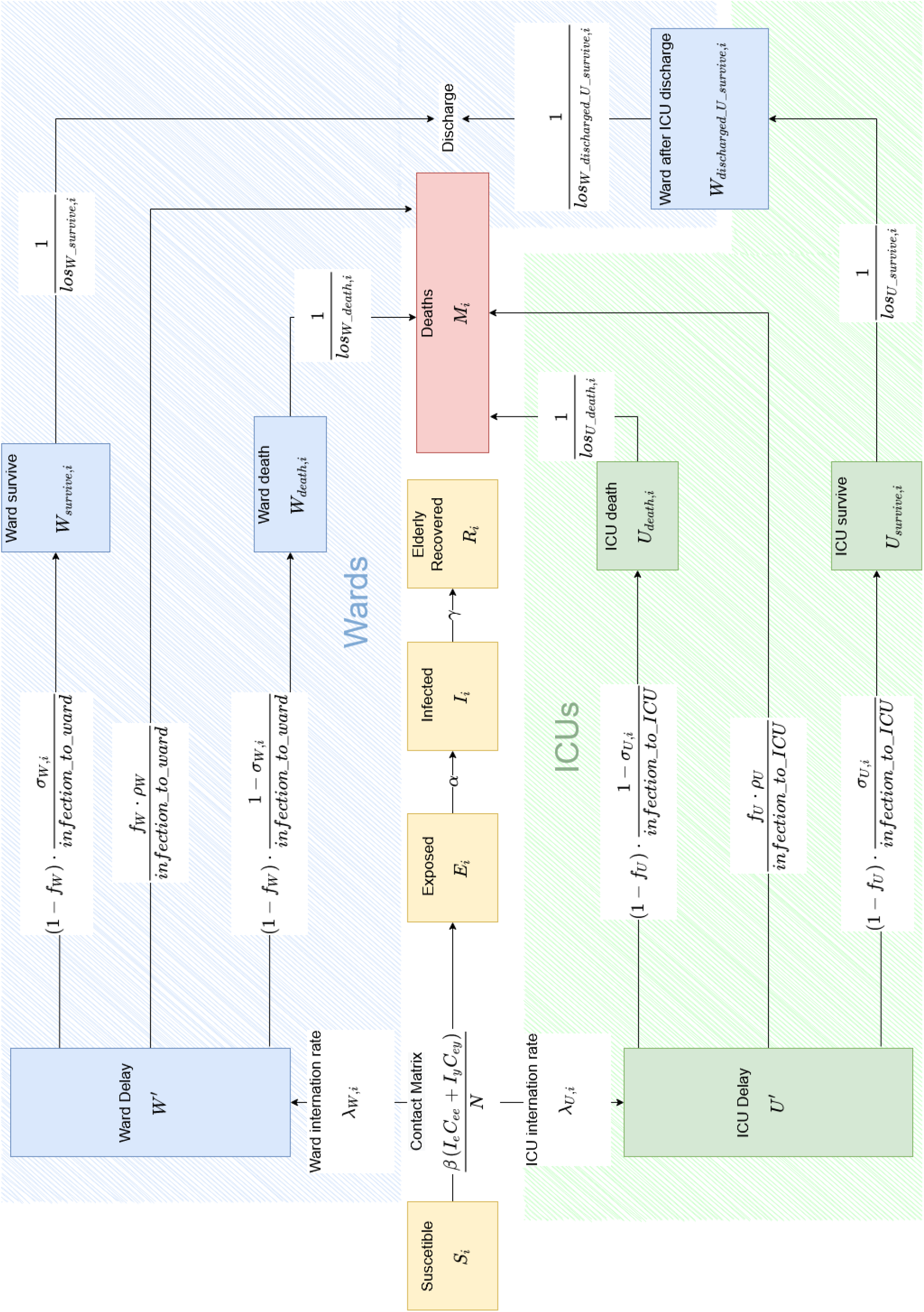
Model flowchart for an specific age group *i*. Contacts between individuals in age group *i* and the other age group *j* are represented by the subscripts _*i*_ and _*j*_ in the contact matrix.

- *W*′: recently infected individuals that are not yet in the hospital, but will require a ward bed soon
- *U*′: recently infected individuals that are not yet in the hospital, but will require an ICU bed soon
- *W*_*survive*_: individuals under ward care that will survive
- *W*_*death*_ individuals under ward care that will die
- *U*_*survive*_: individuals under ICU care that will survive
- *U*_*death*_: individuals under ICU care that will die
- *W*_*discharged*_*_*_*U_ survive*_: individuals discharged from ICU that will survive
- *D*: deceased people

The simulation starts with the number of newly exposed individuals at time step *t*. The parameters *λ*_*W,i*_ and *λ*_*U,i*_ are defined as the probability of an infected individual from the age-group *i* to require a ward or ICU bed, respectively. Following exposure, there is a time interval before hospitalization; therefore, they are firstly assigned in temporary compartments (*W*′ and *U*′). The exit rate from such compartments is respectively proportional to the *infection_to_ward* or *infection_to_ICU* parameters, related to the mean time between exposure and hospitalization. The following equations depict this process:

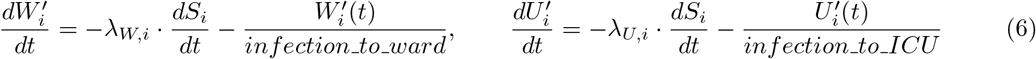

As previously stated, we created two compartments per hospitalization type, related to the two possible outcomes: survival and death. Thus, the natural sequence for those leaving *H*′ and *U*′ is one of the following compartments: *W*_*survive*_, *W*_*death*_, *U*_*survive*_ or *U*_*death*_. Additionally, for hospitalizations, we incorporated the effect of an eventual health system overloading. We constrained the total number of hospitalized individuals at time *t* to the available number of wards or ICU beds within the Brazilian health system (regardless of the age group). We employed a logistic function to model a possible shortage of beds:

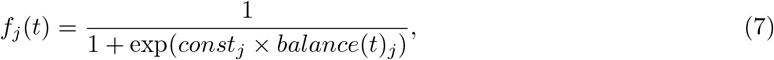

where *f*_*j*_ is the output of the logistic function, *balance*_*j*_ is the total number of available beds and *const*_*j*_ is a constant that calibrates the system to operate above its maximum capacity for each type of care bed *j* (ICU or ward). In the present work, we used −0.01 and −0.06 for *const*_*W*_ and *const*_*U*_ respectively. These values are high enough to impose the desired restriction without introducing numerical issues due to the evaluation of large exponential numbers. The number of available beds *balance*_*W*_ and *balance*_*U*_ are calculated by summing up the contribution of all compartments related to each hospitalization bed location (ICU or ward), considering both age groups:

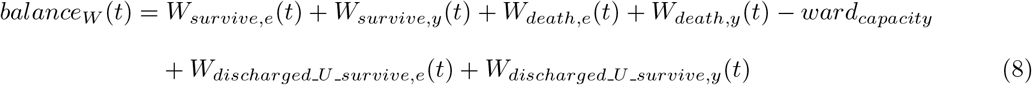

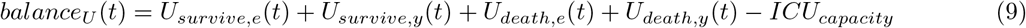

The logistic function is bounded by 0 and 1. The 0 value is returned when the system is not under high load, whereas 1 occurs when the system is under high load. Then, one could multiply the rate of individuals entering the survive and death compartments to 1 minus this logistic function. If no beds are available and the hospital still admitted a small amount of patients over its capacity, the rate of individuals entering those compartments will approximate to zero. On the other hand, if the health system is not under high strain, individuals’ rate will not be affected by this function. For intermediate cases, the ward and ICU compartments could accommodate a fraction of the individuals who require care. Thus, this strategy enables to model the health system overwhelming through a continuous function. The model admits a small level of hospital admissions above its capacity, which might be realistic under a pandemic (e.g using transport beds placed in hallways as ward beds). The occupation of ward and ICU beds are defined by the following equations:

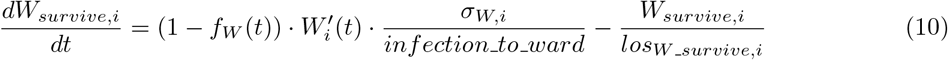

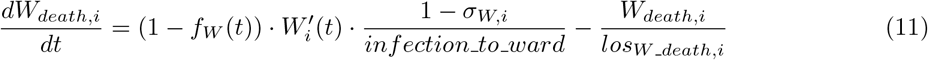

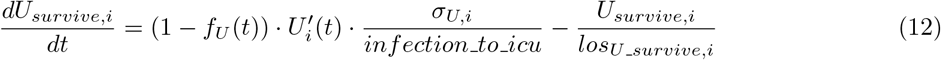

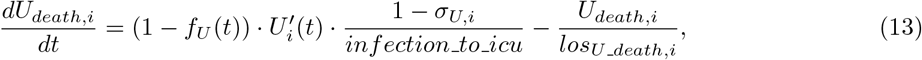

where *los*_*j*_*_*_*k,i*_ represent the average length of stay in a *j* (ward or ICU) bed, for a patient from the age *I* and *k* (death or survive) outcome groups.

In addition, *σ*_*W,i*_ and *σ*_*U,i*_ are the proportions of individuals that will survive in each group of required bed (ward only or ICU). Similarly, (1 − *σ*_*W,i*_) and (1 − *σ*_*U,i*_) are the analogous proportions of those ones who will die.

The individuals that did not receive the needed care are transferred to the death compartment *D*, with probability *ρ*_*W*_ or *ρ*_*U*_ accordingly to each required bed type (ward or ICU respectively). This compartment also receives individuals leaving the *W*_*death*_ and *U*_*death*_ compartments. Entries occurring due to lack of beds because of health system overwhelm are assumed to be instantaneous whereas those from *W*_*death*_ and *U*_*death*_ occur at a specific rate.

This rate is proportional to the average length of stay at each compartment (*los*_*W*_*_*_*death,i*_ and *los*_*U*_*_*_*death,i*_). The following equation describes the death compartment dynamic:

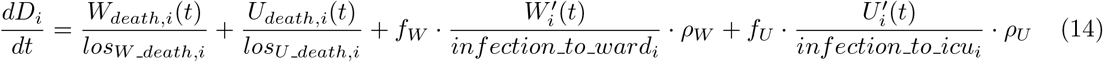

People in the *U*_*survive*_ leave the compartment to *W*_*discharged*_*_*_*U*_*_*_*survive*_ at a rate proportional to *los*_*U*_*_*_*survive,i*_, the average number of days from ICU admission to ICU discharge. They remain in this compartment for an average time of *los*_*W*_*_*_*discharged U*_*_*_*survive*_ before being discharged:

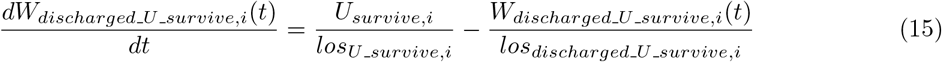

Finally, people at *W*_*survive*_ leave this compartment at a rate proportional to *los*_*W*_*_*_*survive,i*_, the average number of stay for this group of patients, representing the patient discharge to home.

### Brazilian demographic parameters and contact patterns

In our work, we defined two age groups: the young and elderly. The former corresponds to the individuals aged less than 60 years and the latter those aged 60 years or over. We retrieved the demographic data regarding Brazilian population from the Brazilian Institute of Geography and Statistics - IBGE [19], resulting in a total population (*N*) of 211755692. The proportion of young (*p*_*y*_) and elderly (*p*_*e*_) individuals within the Brazilian population are 85.75% and 14.25%, respectively.

Both age groups defined in the present study have distinct general social behavior. Young people have higher contact with other individuals when compared to elder people. In order to account for distinct contact patterns throughout age groups, we employed a contact matrix separating young and elderly citizens. As no empirical survey was conducted for Brazil, the matrix adopted here is derived based on the work of Prem *et al*. [20], where contact matrices for several countries were estimated based on POLYMOD information [21] together with Health and Demographic Surveys and socio-demographic factors.

### Healthcare System parameters

The total number of non-critical care hospitalization beds (ward beds), and critical care beds (intensive care unit beds) that could be eligible to receive COVID-19 patients were obtained from the Ministry of Health of Brazil [22]. For each simulated city, we considered that only 50% of beds could receive patients, as patients with other health conditions would occupy the remaining beds.

### COVID parameters

We adopted the range reported by Li *et al*. [23] for the basic reproduction number (*r*_0_), with mean 2.2 (95% confidence interval [CI], 1.4 to 3.9). In order to assess uncertainty, we sampled different values of a log-normal distribution assuming its 2.5% and 97.5% percentiles as the 95% confidence intervals of the *r*_0_ estimated by Li *et al*. [23]. We scaled the value of *β* to match the desired value of *r*_0_ with the unmitigated contact matrix Prem *et al*. [15].

We adopted the mean incubation period (*α*^−1^) and infectious period (*γ*^−1^) reported by Hao *et al*. [24], *2*.9 and 2.9 days respectively.

The average length of stay in ward and ICUs for each outcome and age group were obtained from 238681 hospitalized COVID-19 cases from Brazil [25] and can be found at Table 2.

We adopted 5 days as the mean time from infection to hospitalization, based on the European Centre for Disease Prevention and Control (ECDC) data [26].

As almost all COVID-19 deaths occurred in Brazil were from hospitalized patients registered in the national SIVEP-SRAG database [25, 27], we calculated the hospitalization rates for ICUs and wards as a proportion of those whom have died. By combining the expected infection fatality rate (IFR) and the proportion of hospitalized patients whom have died, we can estimate the total hospitalization probability for every infected individual from each age group. The infection fatality rate (IFR) used to estimate these values is based on Silva *et al*. [28] seroprevalence study, which has found a IFR of 0.34% for São Luís island. As this place has a human development index and age pyramid very similar to

Brazil, we chose São Luís as a proxy for the IFR. For the situation that an individual needs a hospitalization bed, but there is not one available, a probability of death is assigned. To define this probability, we applied the values reported by Walker *et al*. [16], based on multiple clinical opinions. Thus, we assumed that the probability of death of an individual that needs an ICU bed but does not receive it (p_U_) is 90%. For ward beds (p_W_), this probability is 60%.

### Simulation conditions

The simulation starts at day 0, with 300 exposed, 200 infected and 0 removed individuals as initial conditions for the system. The number of susceptible individuals at day 0 is calculated by the city population minus the number of exposed and infected individuals at day 0. The simulation ends at day 250. As the present study’s goal is to compare distinct scenarios, day 0 does not necessarily matches any date of specific events from the current pandemic.

The distinction between unmitigated scenario and vertical isolation is done by the contact matrix. The results of Prem *et al*. [20] are divided by the places where the contact occurs (at work, school, home or others). For the unmitigated case, the full matrix is considered for both young and elderly, without modification. For the vertical isolation case, the contacts for elderly are reduced to only those ones performed at home.

Simultaneously, the contact patterns for the young are adjusted for accounting on this new behavior of the elderly. In other words, a young individual could meet an elderly one just at home, but would find another young with the same frequency as in the unmitigated scenario.

A total of 1000 runs were performed for assessing uncertainty of each evaluated scenario.

The tables 1, 2 and 3 summarize the parameters used in the simulations as well as its main data sources.

**Table 1:**
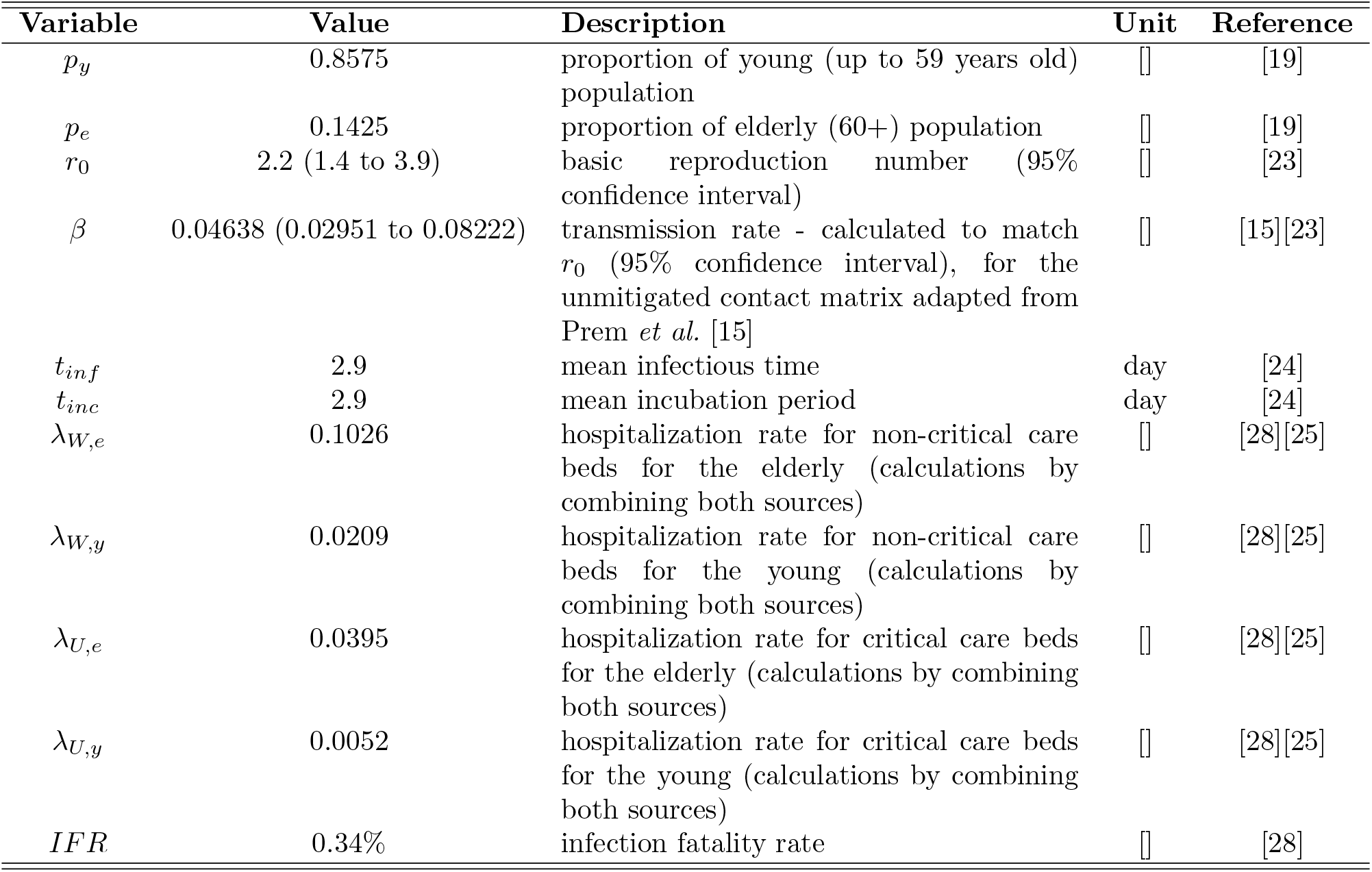
Parameters used in the simulations 1/3

**Table 2:** Parameters used in the simulations 2/3

| Variable | Value | Description | Unit | Reference |
| --- | --- | --- | --- | --- |
| $losU\_survive,e$ | 9.847628 | average length of stay of an elder patient in an ICU bed who will survive | day | [25] |
| $losU\_survive,y$ | 8.736200 | average length of stay of a young patient in an ICU bed who will survive | day | [25] |
| $losU\_death,e$ | 9.868753 | average length of stay of an elder patient in an ICU bed who will die | day | [25] |
| $losU\_death,y$ | 10.720939 | average length of stay of a young patient in an ICU bed who will die | day | [25] |
| $losW\_survive,e$ | 9.394910 | average length of stay of an elder patient in a ward bed who will survive | day | [25] |
| $losW\_survive,y$ | 7.494861 | average length of stay of a young patient in a ward bed who will survive | day | [25] |
| $losW\_death,e$ | 9.201503 | average length of stay of an elder patient in a ward bed who will die | day | [25] |
| $losW\_death,y$ | 9.475104 | average length of stay of a young patient in a ward bed who will die | day | [25] |
| $losdischarged\_U\_survive,e$ | 6.572384 | average length of stay of an elder patient that was discharged from the ICU to a non-critical care hospitalization bed who will survive | day | [25] |
| $losdischarged\_U\_survive,y$ | 4.594393 | average length of stay of a young patient that was discharged from the ICU to a non-critical care hospitalization bed who will survive | day | [25] |

**Table 3:** Parameters used in the simulations 3/3

| Variable | Value | Description | Unit | Reference |
| --- | --- | --- | --- | --- |
| $\sigma_{W,e}$ | 0.606622 | proportion elderly COVID-19 patients in ward beds that will survive | [] | [25] |
| $\sigma_{W,y}$ | 0.892248 | proportion young COVID-19 patients in ward beds that will survive | [] | [25] |
| $\sigma_{U,e}$ | 0.259130 | proportion elderly COVID-19 patients in ICU beds that will survive | [] | [25] |
| $\sigma_{U,y}$ | 0.566919 | proportion young COVID-19 patients in ICU beds that will survive | [] | [25] |
| $\rho_W$ | 0.6 | death rate for patients that needed a ward bed but could not get one because the health system was overwhelmed | [] | [16] |
| $\rho_U$ | 0.9 | death rate for patients that needed an ICU bed but could not get one because the health system was overwhelmed | [] | [16] |
| $ward_{capacity}$ | $\epsilon \cdot 0.5$ | Number of ward beds available for COVID-19 patients, where $\epsilon$ is the total number of ward beds for the simulated city | bed | [22] |
| $ICU_{capacity}$ | $\theta \cdot 0.5$ | Number of ICU beds available for COVID-19 patients, where $\theta$ is the total number of ICU beds for the simulated city | bed | [22] |

## 3. Results

We run 1000 simulations for all state capitals of Brazil, accounting for their hospital bed capacity and population. We evaluated the effects of a hypothetical scenario where either no intervention was adopted, or the vertical quarantine was adopted during the entire simulated period. Many of these cities may have implemented distinct COVID-19 mitigation strategies from those that we analysed. Since some of these strategies are out of our assumptions, our simulations may not precisely match the current observed epidemic behavior.

Figure 2 presents the number of individuals that recovered or died (removed individuals) at the end of the simulation period, for each simulation scenario, accounting for the 95% uncertainty interval due to uncertainty on model parameters. It is noteworthy that all the box plots within this work represent the whiskers *at* 1.5 interquartil range. For São Paulo, when compared to no isolation, the vertical quarantine resulted in a 4% reduction in the total number of removed people at the end of the epidemic. However, in the elderly group the reduction was considerable, with a 53% reduction on median values. Figure 3 and 4 depict the total number of required beds for COVID-19 infected patients for each scenario through time for São Paulo. Not only São Paulo, but all the simulated cities had at some point median hospitalization demands that were above the installed capacity for ICUs (see Supplementary Material). This scenario also occurred for ward beds in some cities, yet in a lesser extent. When compared to no isolation, the vertical quarantine scenario reduced the demand of hospital beds slightly; however, the reduction was not enough to avoid a healthcare system overwhelm, especially for ICUs.

**Figure 2:**
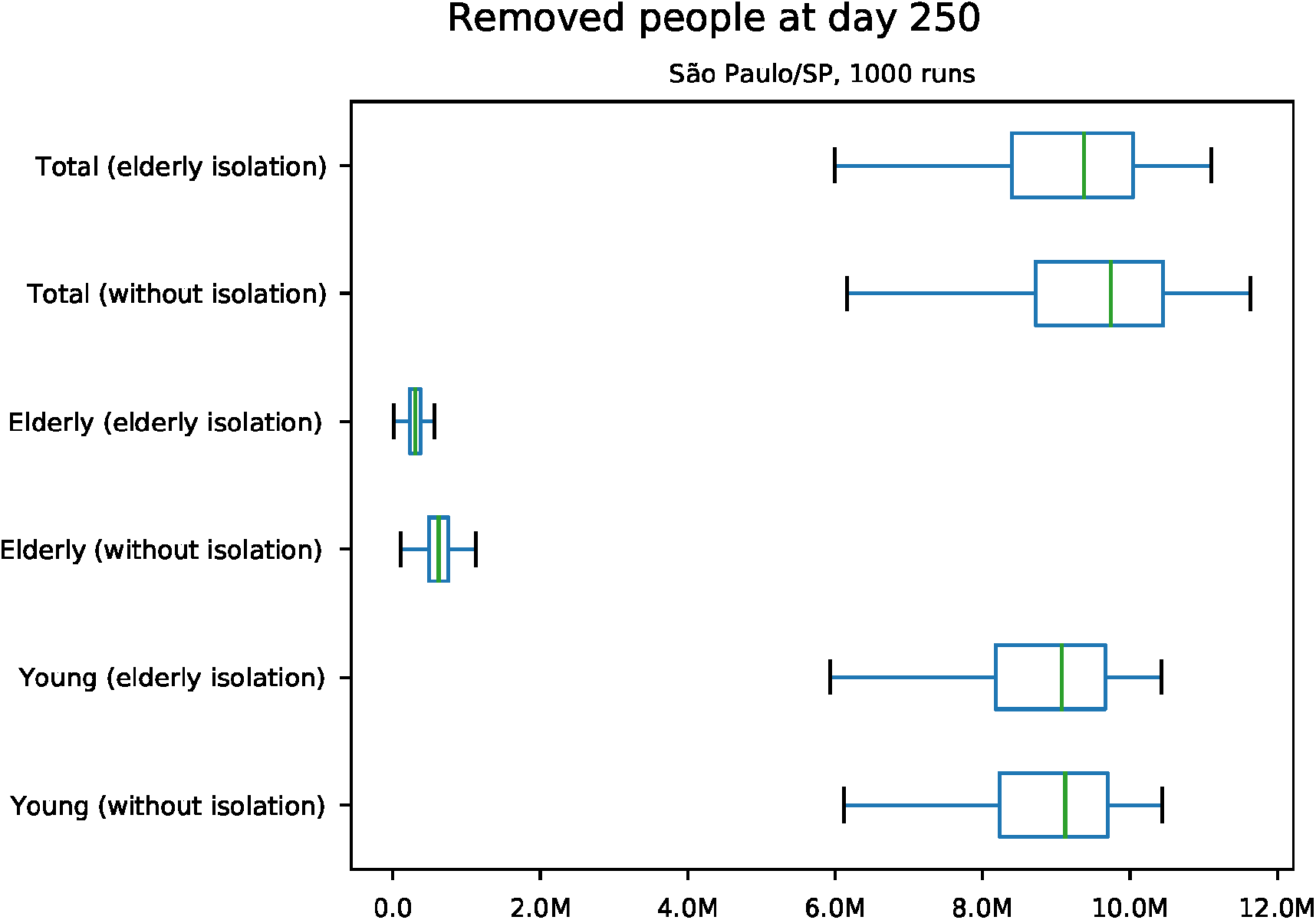
Predicted number of removed individuals at day 250, for São Paulo/SP

**Figure 3:**
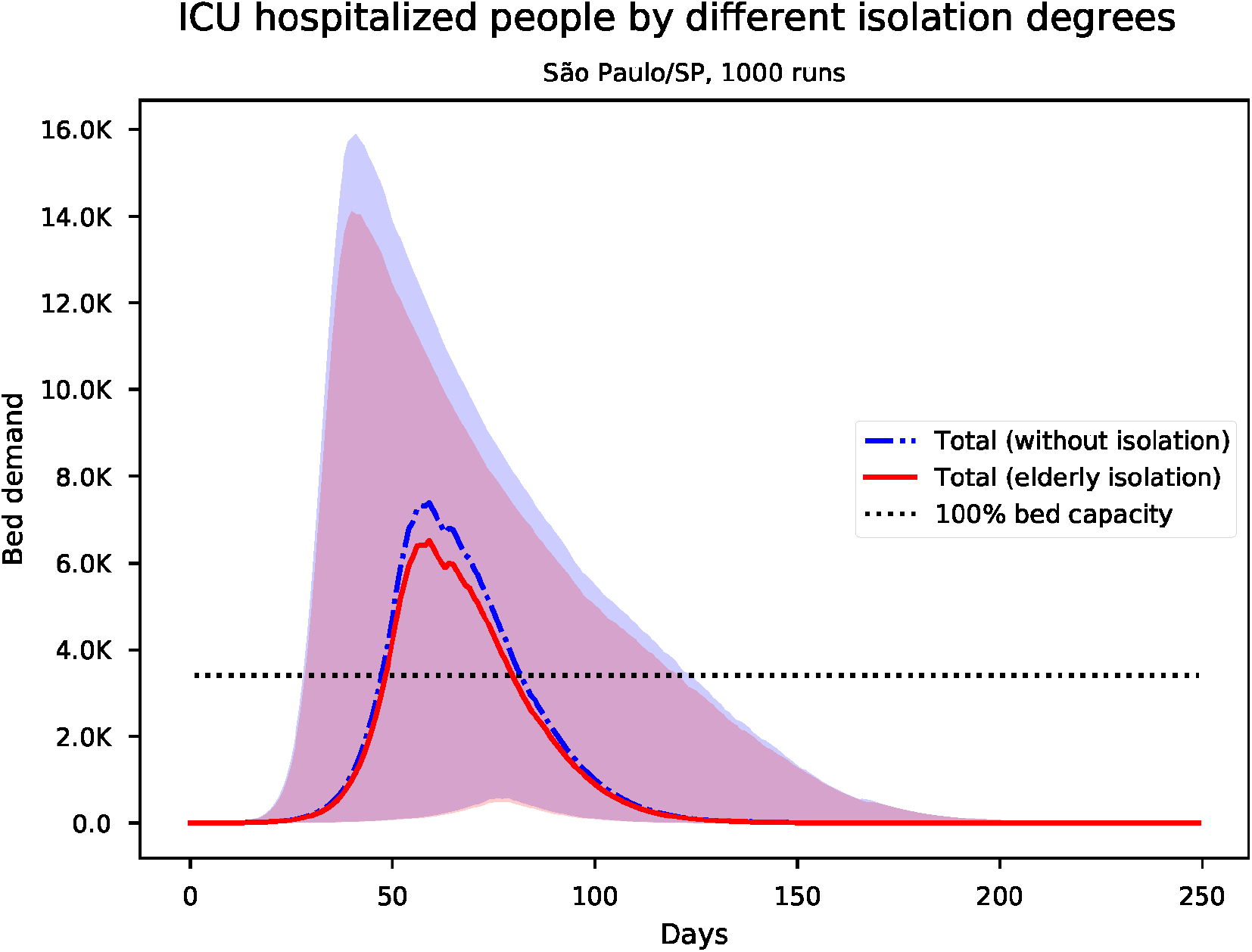
Predicted number of needed ICU beds for São Paulo/SP

**Figure 4:**
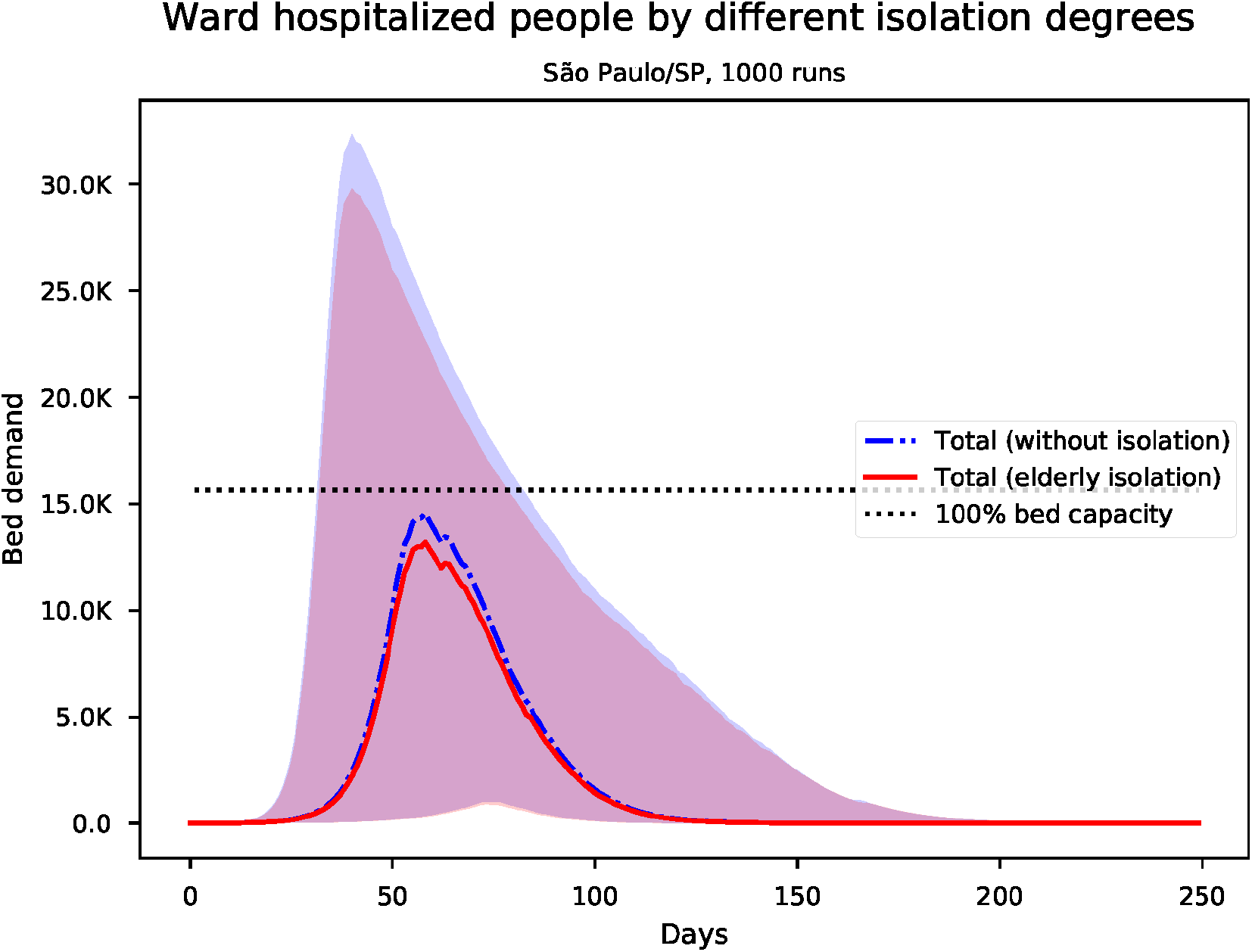
Predicted number of needed ward beds for São Paulo/SP

For São Paulo, when compared to no isolation, the vertical quarantine achieves some reduction in the total hospitalization demand (9% on number of bed-days median), even with a substantial reduction in hospitalization demand for elder patients (52% on number of bed-days median). The reduction on the number of ward and ICU beds by age group can be seen in figures 6 and 5. Despite young individuals have a proportionally lower hospitalization rate compared to elderly people, as the younger population comprises about 86% of the Brazilian population, the contribution of all young people cases surpasses the hospitalization of elder patients on both scenarios.

**Figure 5:**
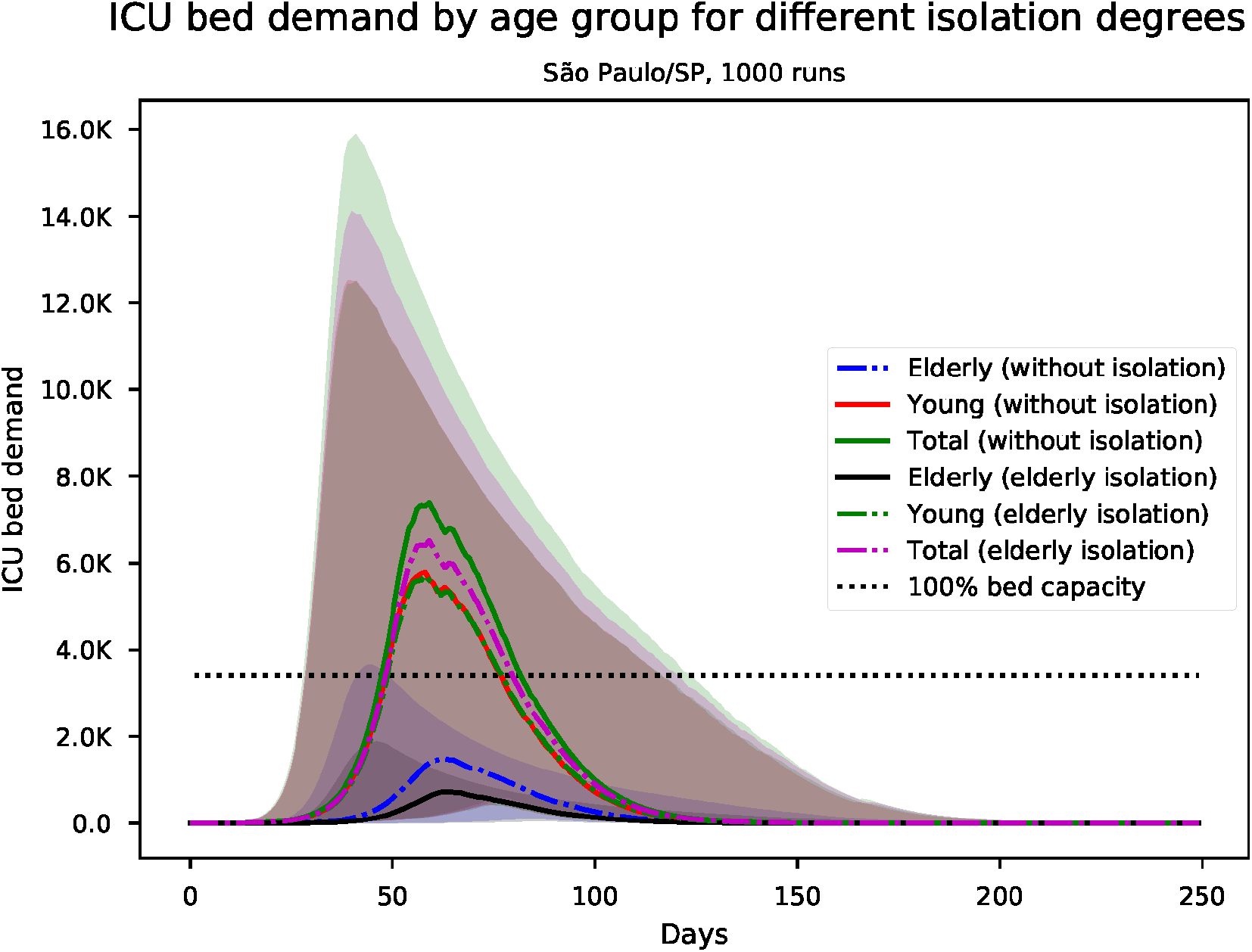
Predicted number of needed ICU beds, by age group for São Paulo/SP

**Figure 6:**
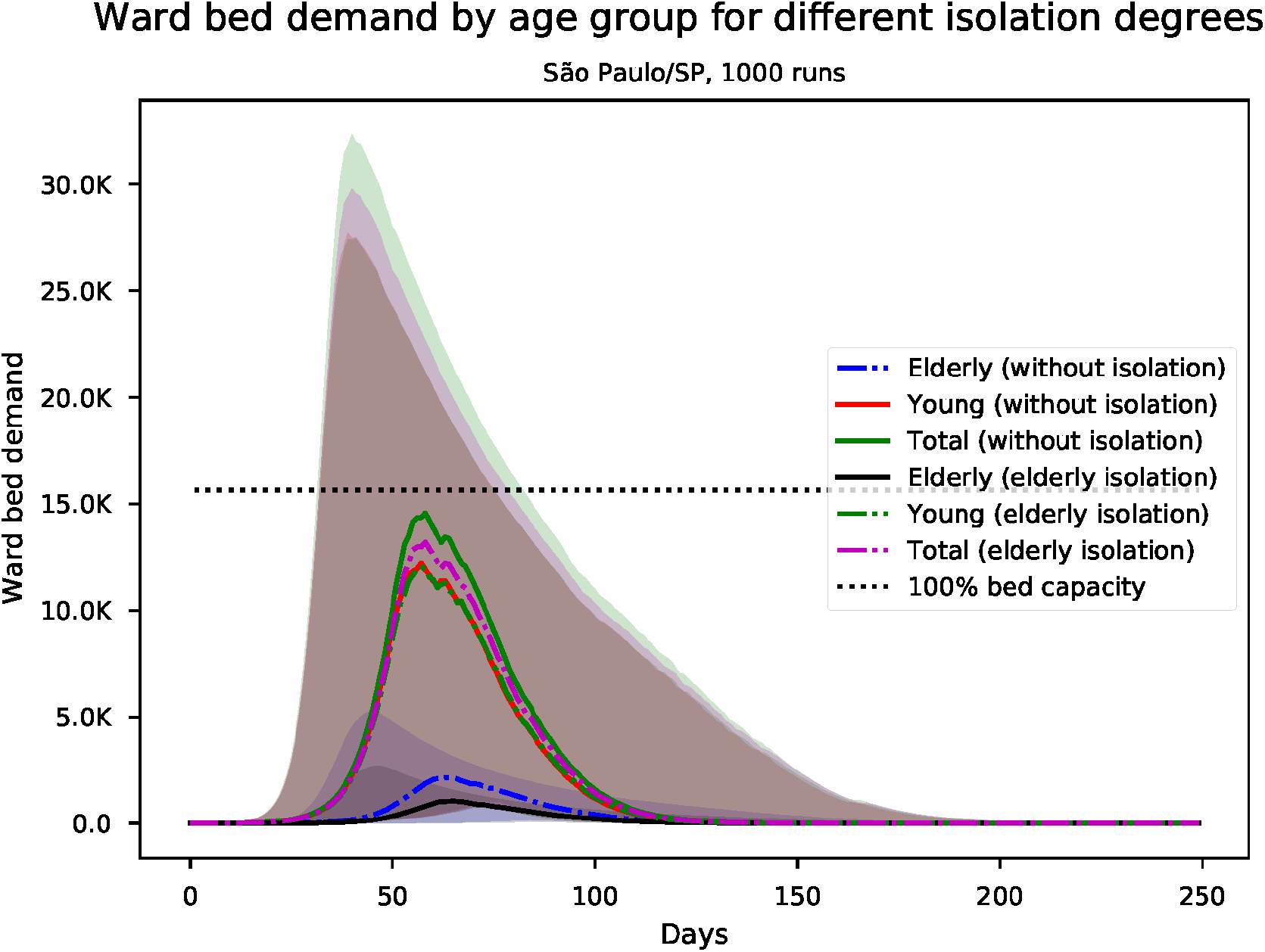
Predicted number of needed ward beds, by age group for São Paulo/SP

The vertical quarantine produced analogous effects on deaths. Figure 7 shows the estimated number of (cumulative) deaths at the end of the simulation. Although the vertical quarantine reduced 53% of the deaths in the elderly population compared to no isolation, the overall reduction of deaths was 16%. This reduction is mainly due to a decrease in elderly people cases, which have higher admission rates and infection fatality rates than young people. Additionally, there is a small reduction in bed demand over capacity in vertical quarantine scenario, contributing to a reduction in deaths. However, in both scenarios, the health system collapse is still responsible for a large number of deaths. The lack of available beds increases the fatality ratio for both, the elderly and young population. Overall, the vertical isolation strategy just marginally reduced the total death count. The number of beds required above capacity for ward and ICU beds (dotted black lines in figure 3 and 4) is also strongly related to deaths. Some variation in the proportion of deaths over city population may vary according to the proportional number of available beds in each city. For the other simulated cities, the median reduction of deaths ranged from 12% to 18%. Results of the simulations for other Brazilian state capitals can be found in the Supplementary Material.

**Figure 7:**
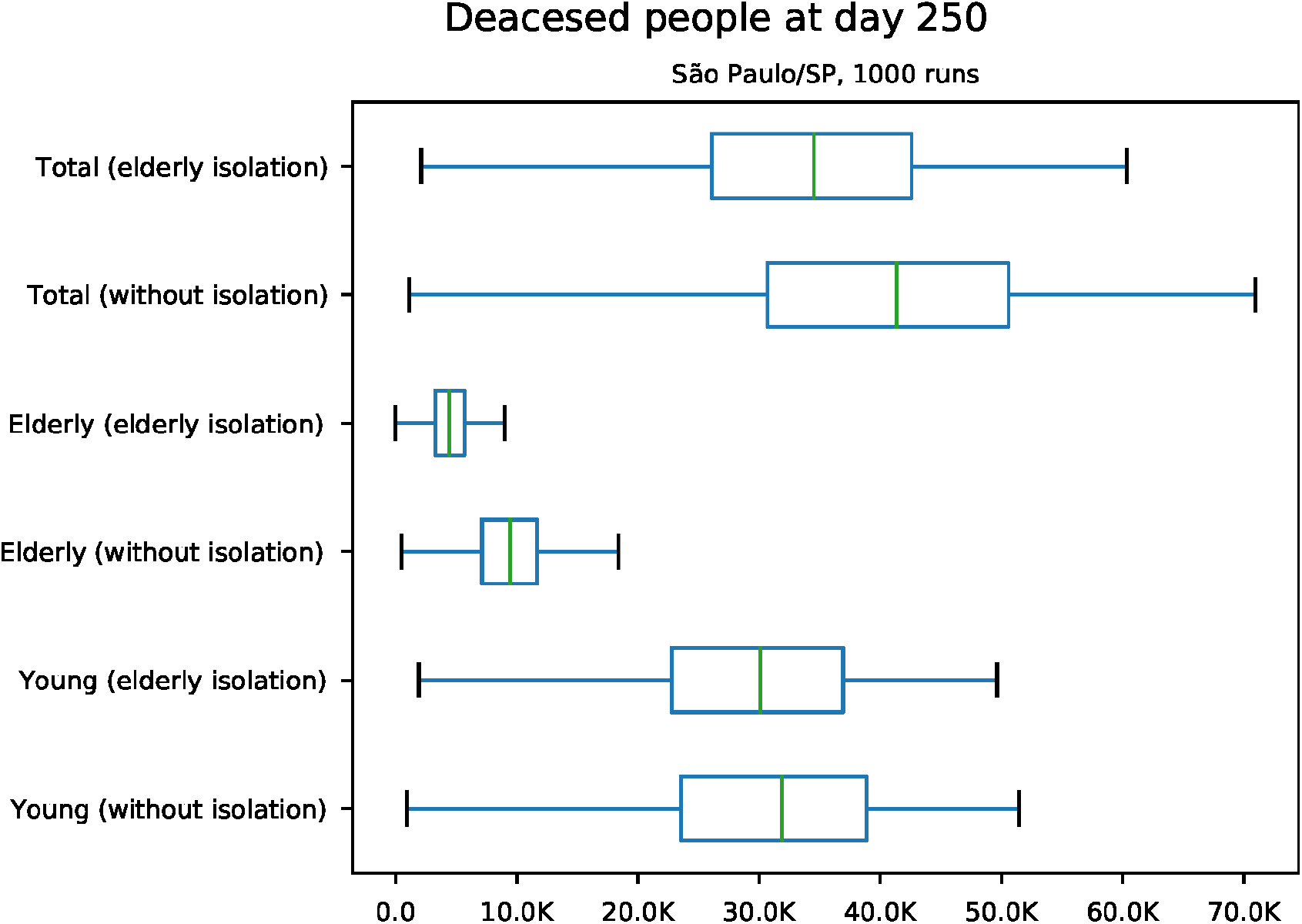
Predicted (cumulative) number of deaths at day 250 for São Paulo/SP

## 4. Discussion

Our simulations provide a better understanding of the possible impacts of adopting a vertical quarantine in the Brazilian context. When compared to no isolation, the vertical quarantine strategy might reduce overall COVID-19 cases, mortality and hospital demand by a small factor. However, the sole adoption of vertical quarantine does not seem to be enough to avoid a healthcare system collapse by exceeding the supply of available beds to receive patients with COVID-19, especially ICU beds. This collapse is responsible for a huge fraction of the total death counts. If standard critical care were available, part of these deaths would be avoided. Thus, other strategies might also be required in order to reduce the excessive amount of hospitalizations.

Duczmal *et al*. [29] also modeled the vertical quarantine for Belo Horizonte city in Brazil, finding that it would result in only a small reduction of COVID-19 cases, in accordance with our results. Duczmal and colleagues did not modeled the effects on hospitalizations or deaths.

Broader isolation strategies should be more suitable for controlling epidemic growth and reducing health system load. In this context, Walker *et al*. [16] suggested that a 75% reduction in contacts in all age groups would bring the effective reproduction number *R*_*t*_ to less than 1 for a basic reproduction number *r*_0_ = 3.0. This approach would produce a suppression in cases and reduce the number of deaths and health system strain as long as it stays in place. However, as the authors point out, this strategy might be challenging due to the high informal labor level in low-income and low medium income countries [30]. Even Brazil being considered an upper-middle-income economy by the World Bank [31], pursuing this strategy would be challenging.

In order to control the epidemic, more suitable strategies could be followed apart from age group based on isolation or indiscriminate isolation. Lu *et al*. [32] list some successful examples available worldwide. Strategies like contact tracing, quarantine of confirmed and suspected cases and eventually broader cluster isolation strategies could be done as the number of cases grows up.

### Study limitations

In our model, we considered only the age to distinguish risk groups. More sophisticated models could add more factors e.g., considering those with heart or respiratory diseases. By increasing the number of people isolated by the vertical quarantine strategy, it would be probable more effective, reducing death as well as hospitalization numbers. However, using a broader inclusion criterion for the risk group is likely to include the economically active population in the isolation group, which could have considerably different economic and social impacts.

Our model assumes a homogeneous distribution of the citizens within each age group and no contacts between individuals from different cities. It does not consider the spatial dynamics, seasonality, effects of treatments or vaccine, healthcare system networks and heterogeneous mixture of the population, except those determined by the age contact patterns. Such phenomena could alter the dissemination dynamics within the population.

Our model assumes a standard population pyramid based on the Brazilian population pyramid for all simulated cities. However, the population pyramid for the simulated cities can vary significantly, changing the expected number of deaths for each city, since there is a strong positive relationship between age and COVID-19 infection fatality ratio [33]. Further studies should address this issue by using different population pyramids accordingly to each city.

## 5. Conclusion

The vertical quarantine strategy would result in a small decrease in COVID-19 deaths and hospitalizations compared to the no isolation strategy. Our model shows that this reduction in hospitalizations would probably not be enough to prevent a health system overwhelming in Brazilian state capitals. Thus, a considerable number of deaths could occur due to the lack of medical care. Hence, the vertical quarantine strategy would not be able to control the pandemic alone, however, it could be considered an element of a broader health policy.

## Data Availability

The code used for generating the results is openly available in Zenodo at https://doi.org/10.5281/zenodo.4393737

