## Supplementary Material for "Mitigation of COVID-19 using social distancing of the elderly in Brazil: The vertical quarantine effects in hospitalizations and deaths"

### Results for Brazilian state capitals

Results for São Paulo are in the main text.

*S1. Results for Aracaju*

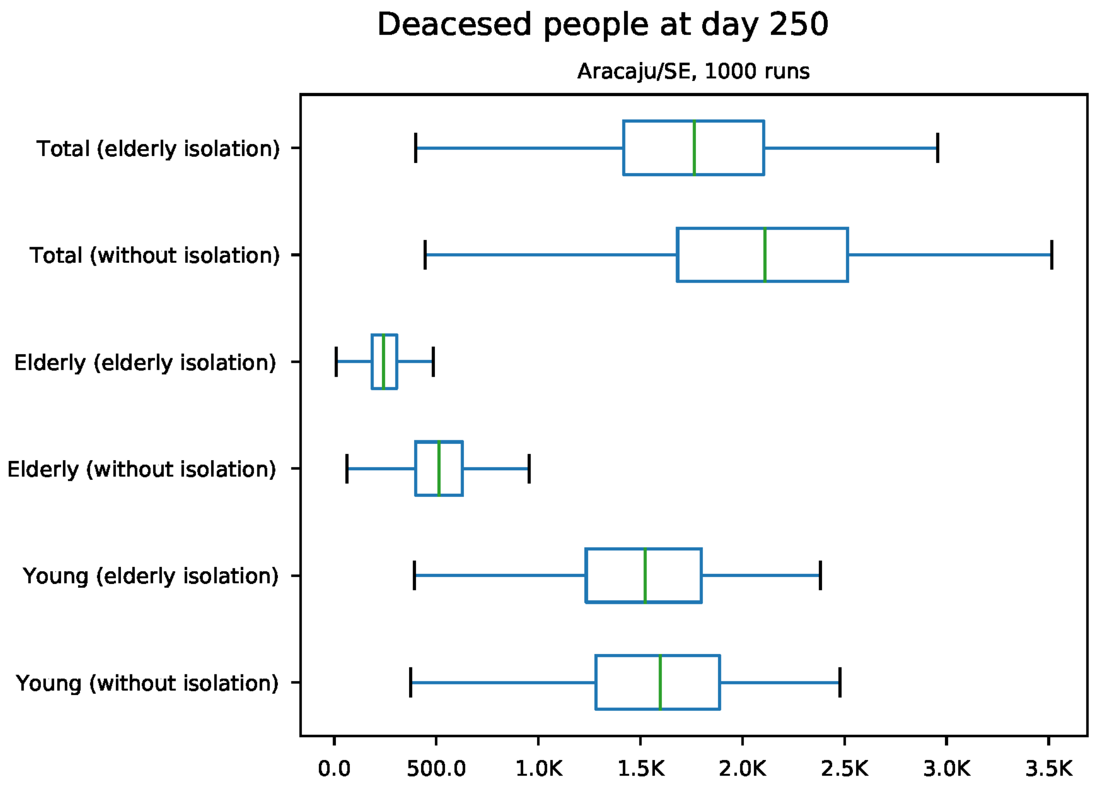


Figure S1: Predicted number of death at day 250 for Aracaju

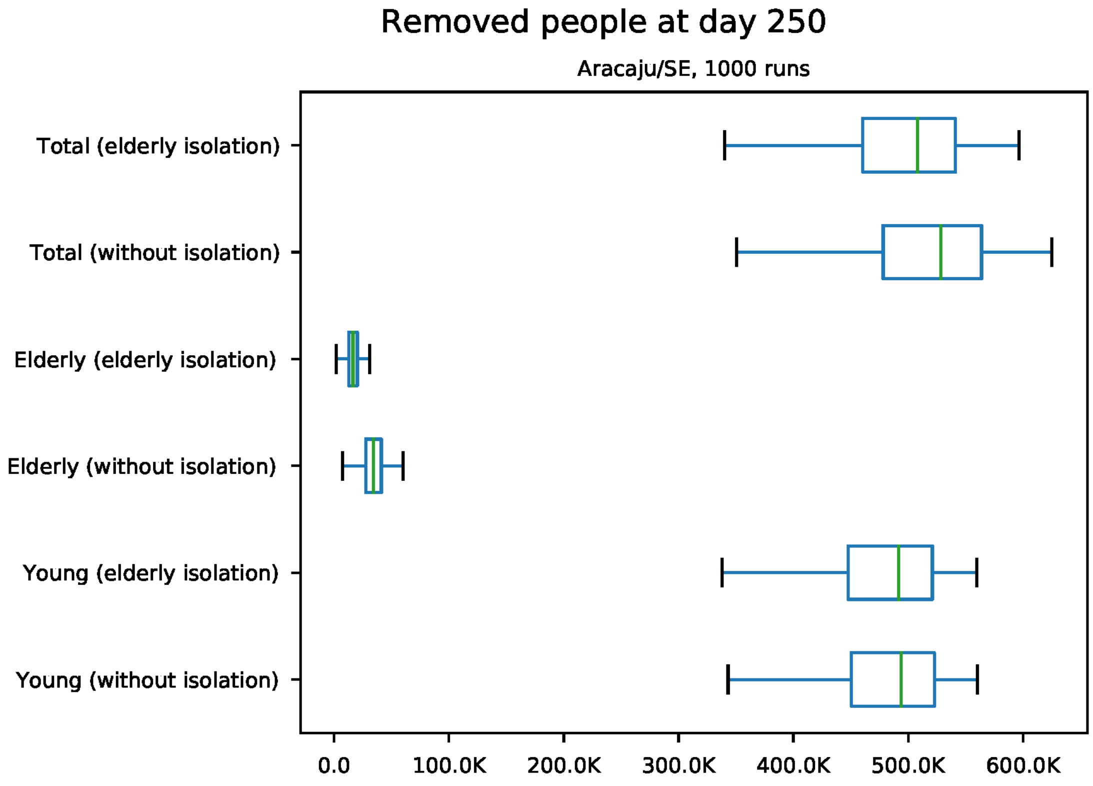


Figure S2: Predicted number of removed individuals at day 250 for Aracaju

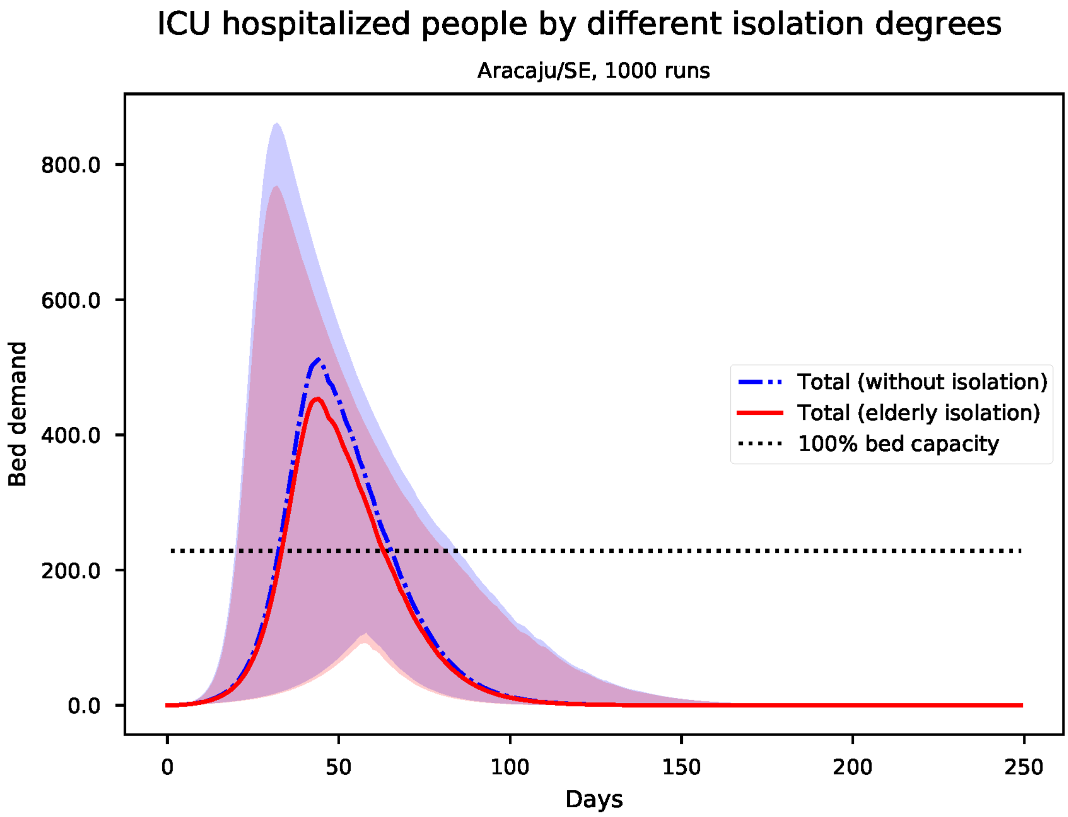


Figure S3: Predicted number of needed ICU beds for Aracaju

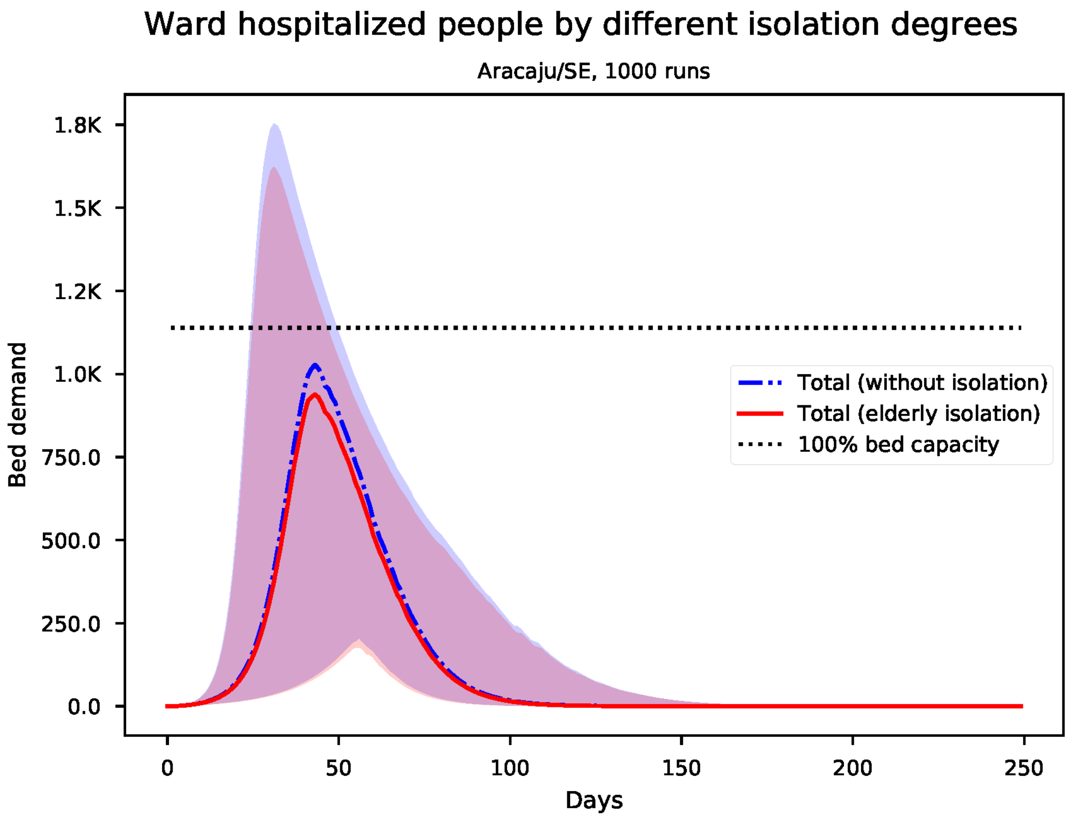


Figure S4: Predicted number of needed ward beds for Aracaju

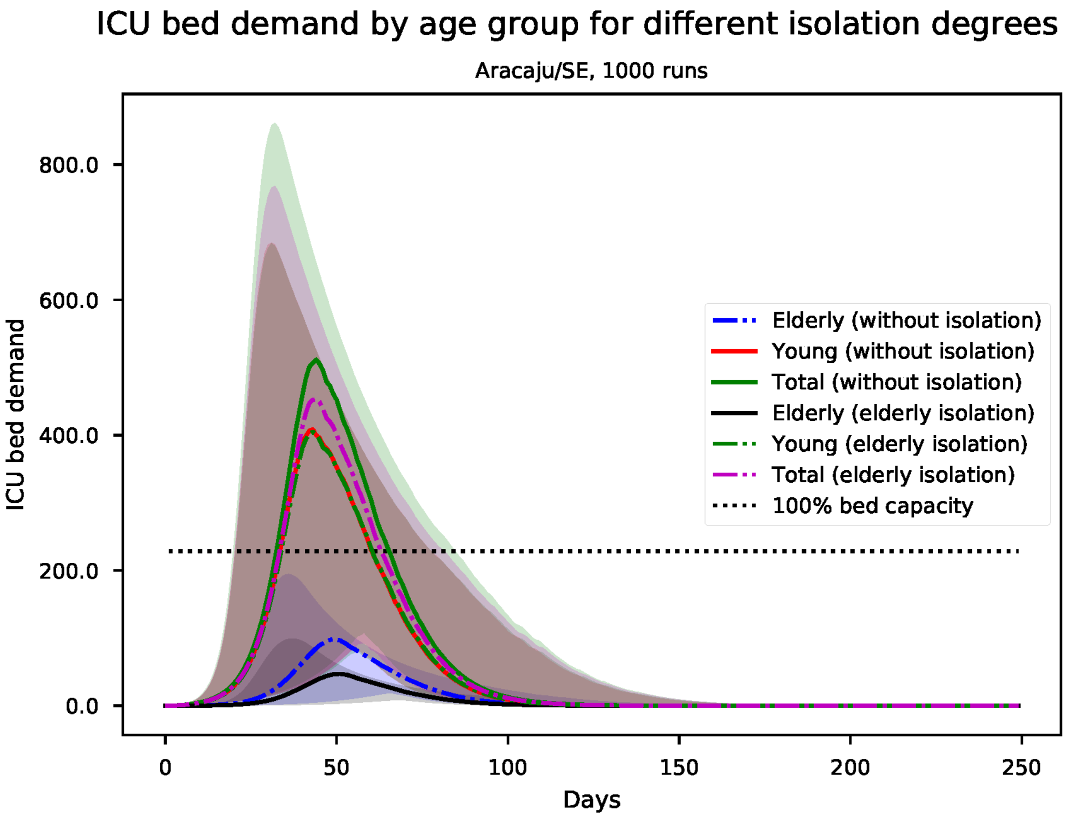


Figure S5: Predicted number of needed ICU beds, by age group for Aracaju

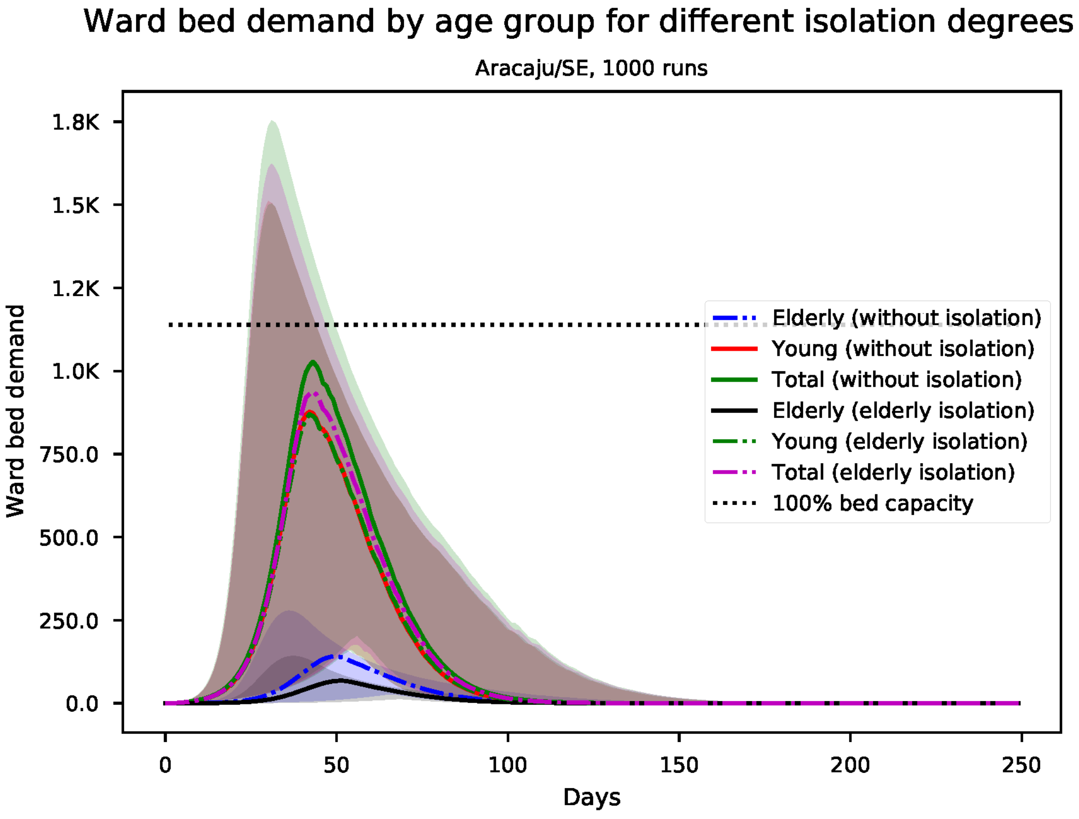


Figure S6: Predicted number of needed ward beds, by age group for Aracaju

*S2. Results for Belo Horizonte*

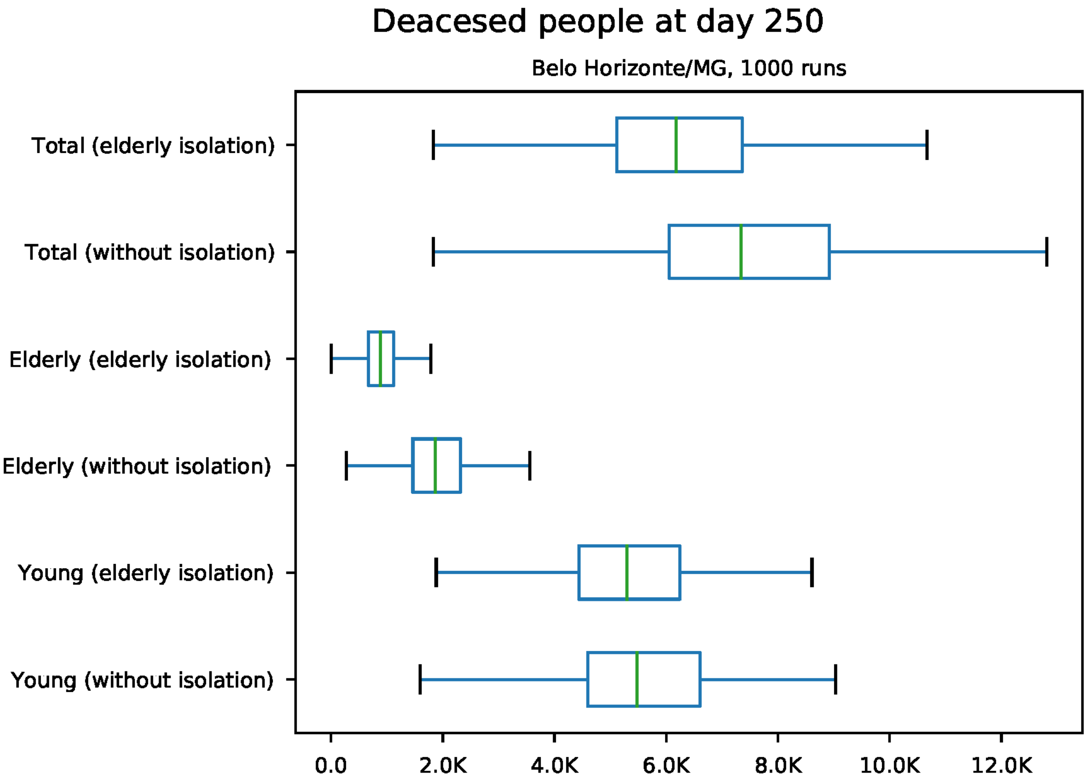


Figure S7: Predicted number of death at day 250 for Belo Horizonte

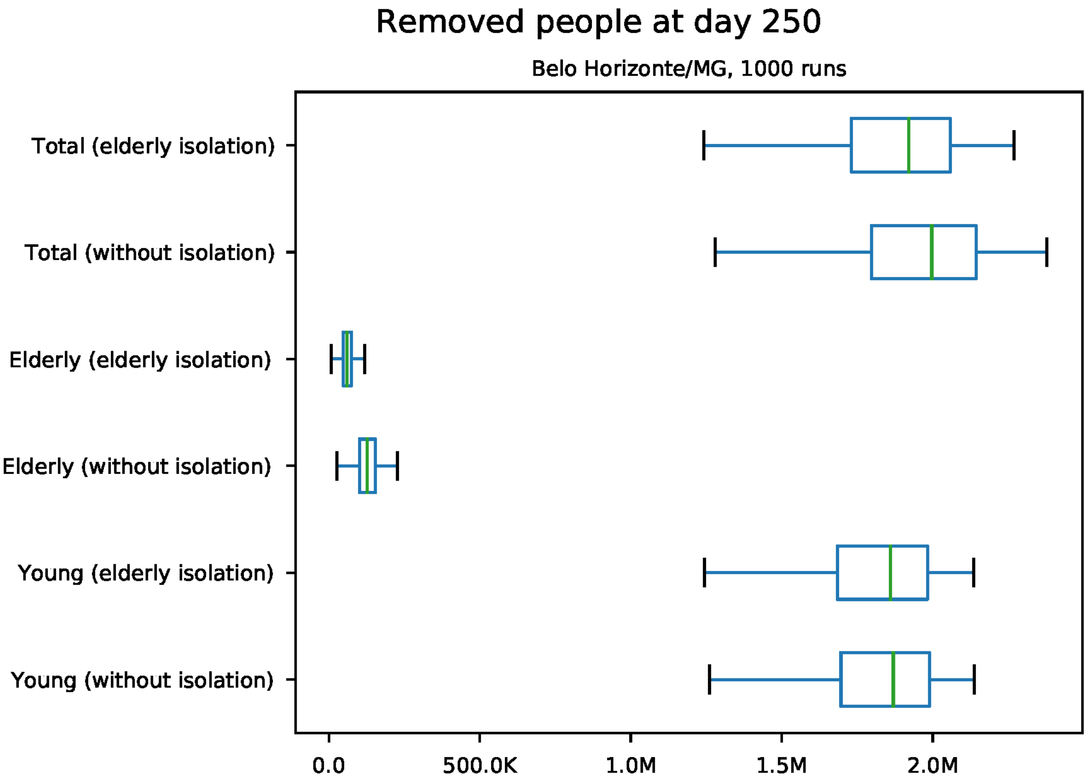


Figure S8: Predicted number of removed individuals at day 250 for Belo Horizonte

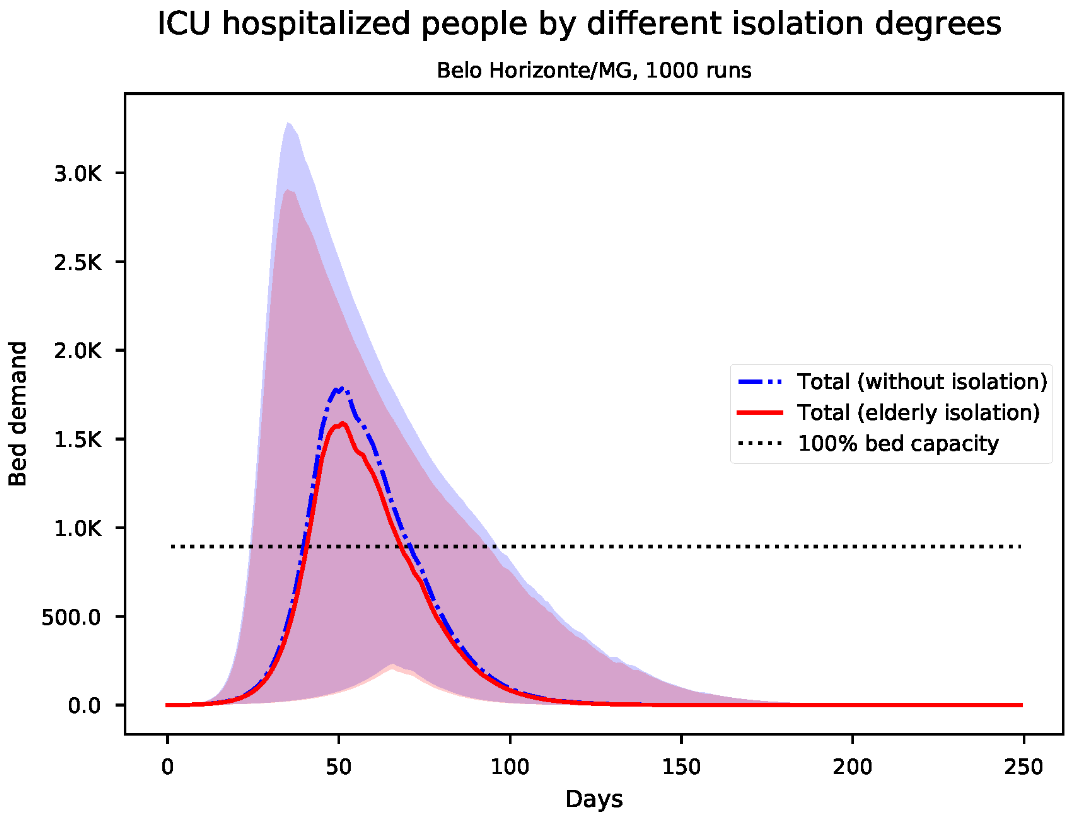


Figure S9: Predicted number of needed ICU beds for Belo Horizonte

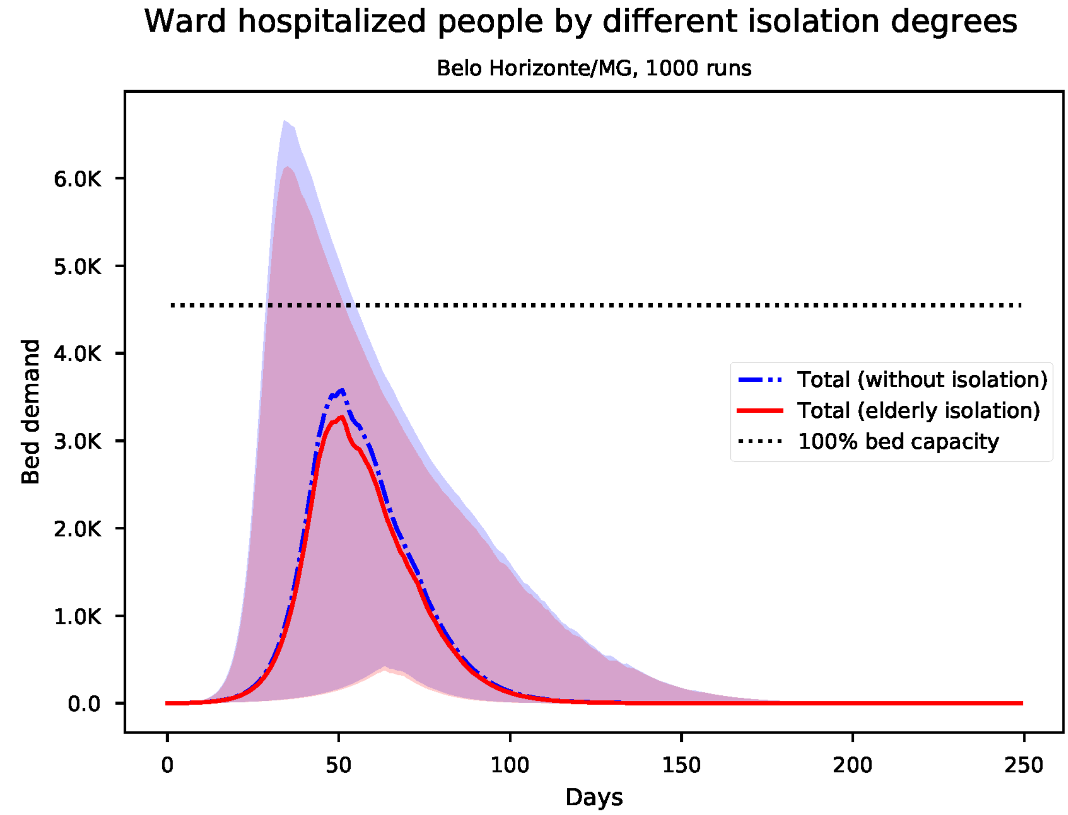


Figure S10: Predicted number of needed ward beds for Belo Horizonte

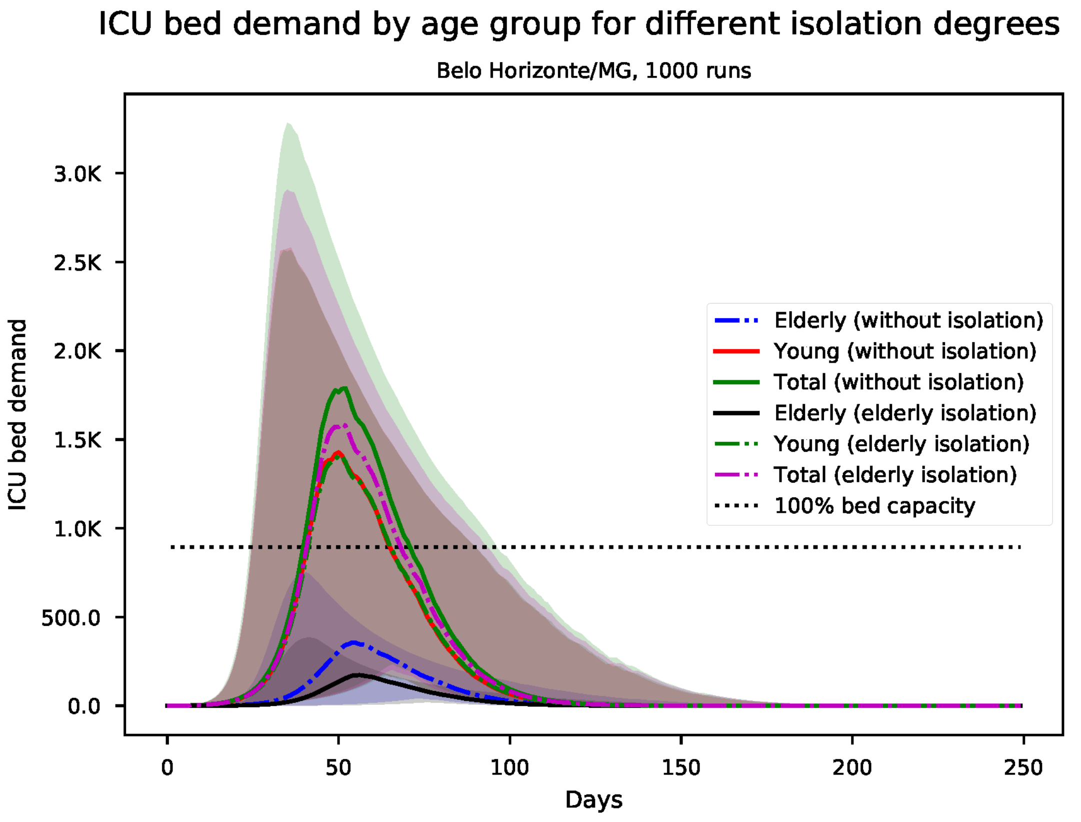


Figure S11: Predicted number of needed ICU beds, by age group for Belo Horizonte

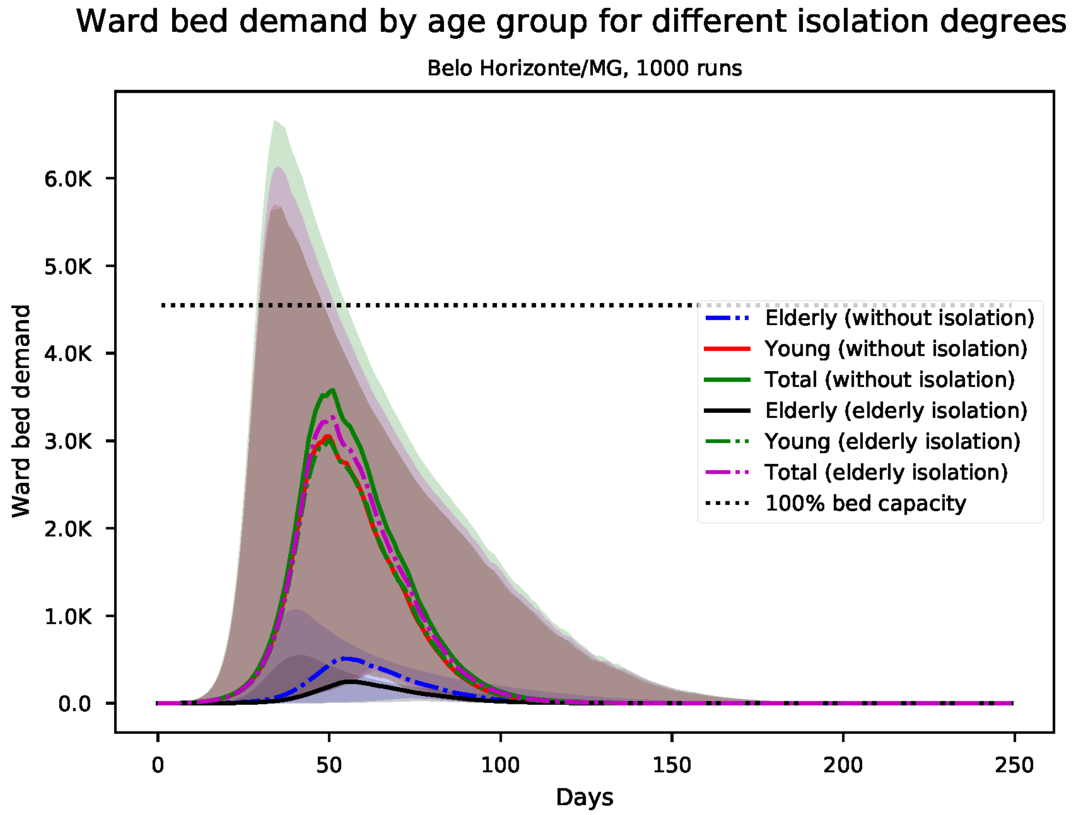


Figure S12: Predicted number of needed ward beds, by age group for Belo Horizonte

*S3. Results for Belém*

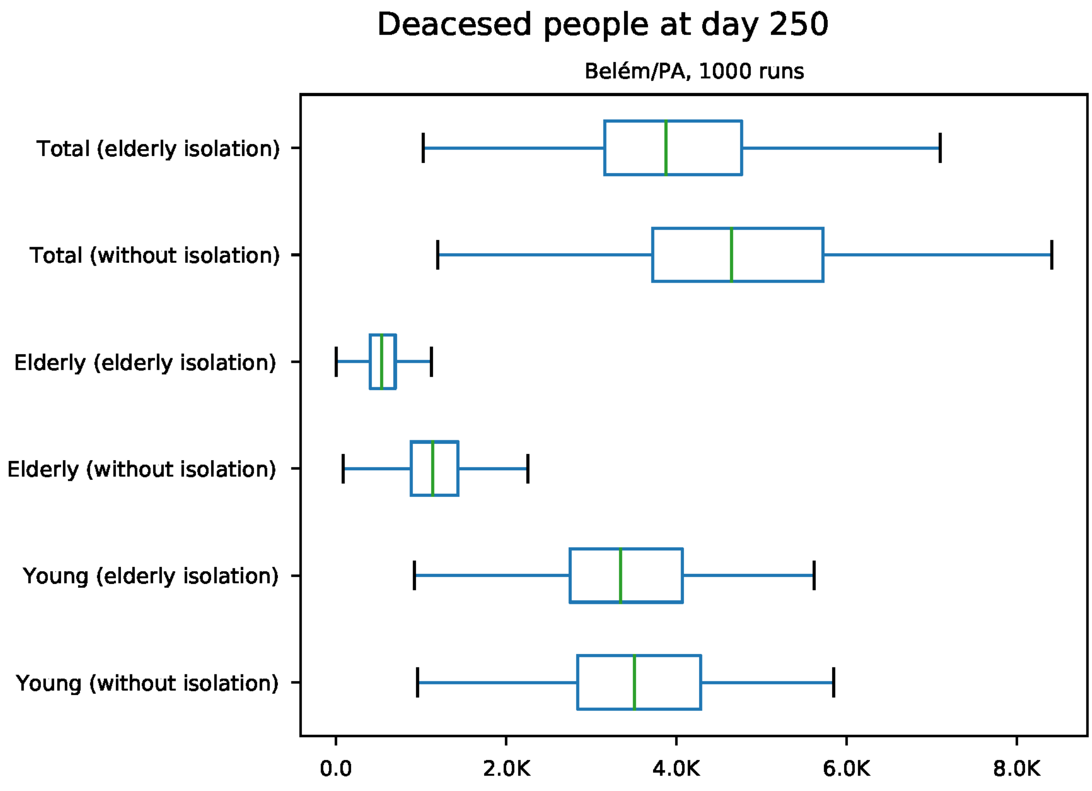


Figure S13: Predicted number of death at day 250 for Belém

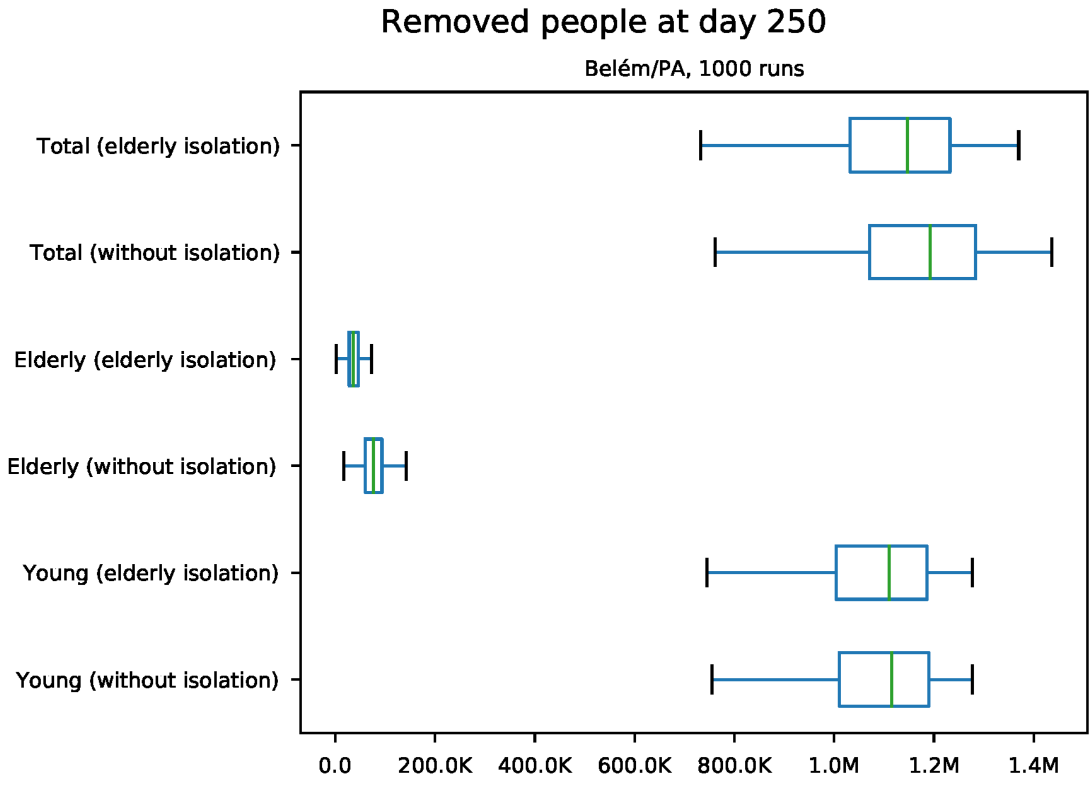


Figure S14: Predicted number of removed individuals at day 250 for Belém

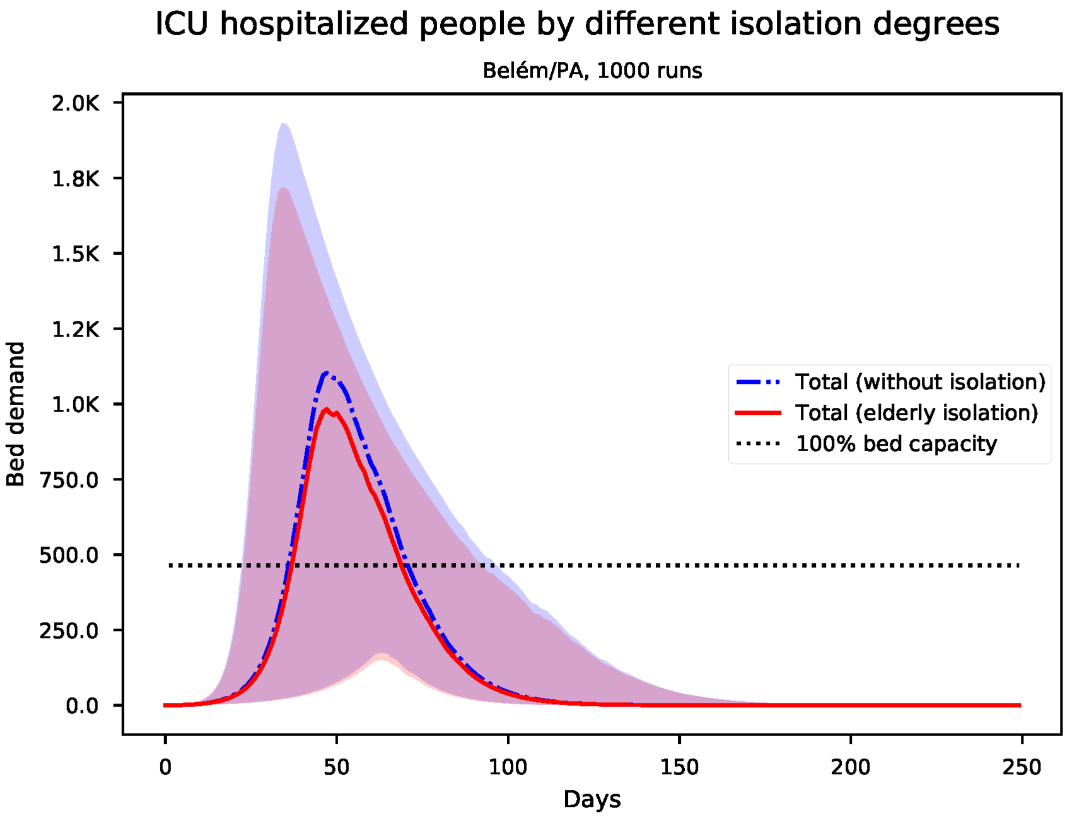


Figure S15: Predicted number of needed ICU beds for Belém

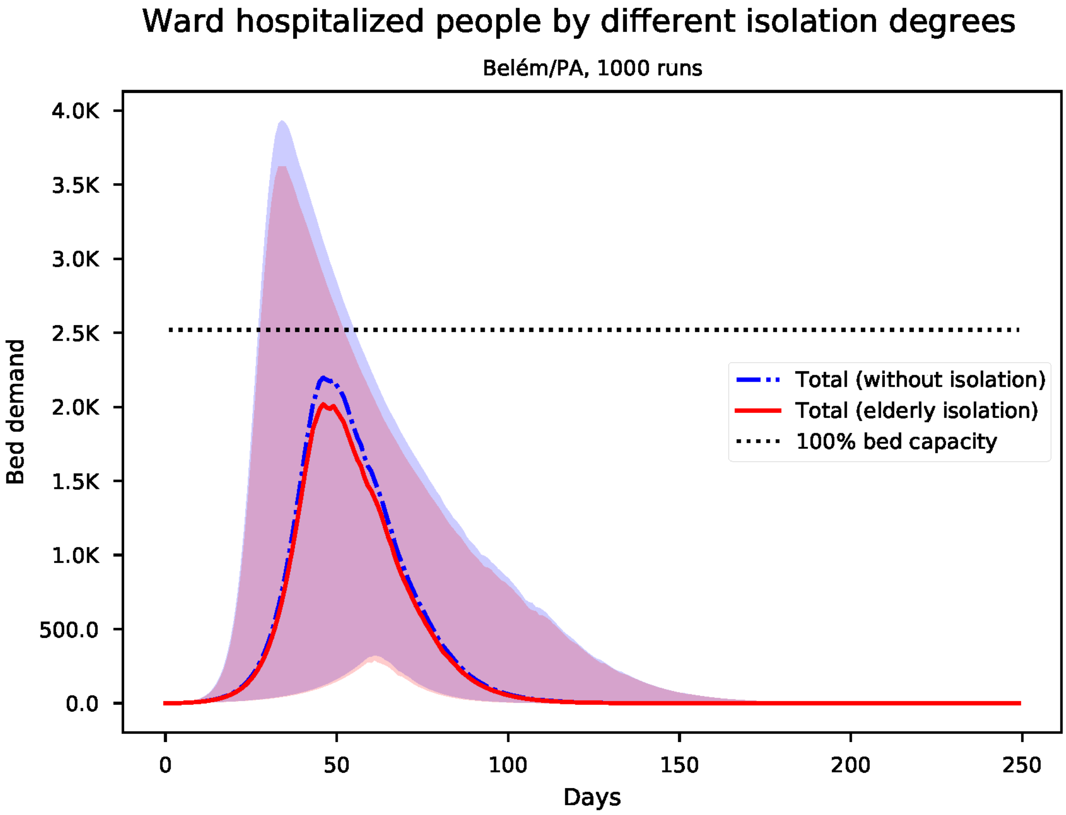


Figure S16: Predicted number of needed ward beds for Belém

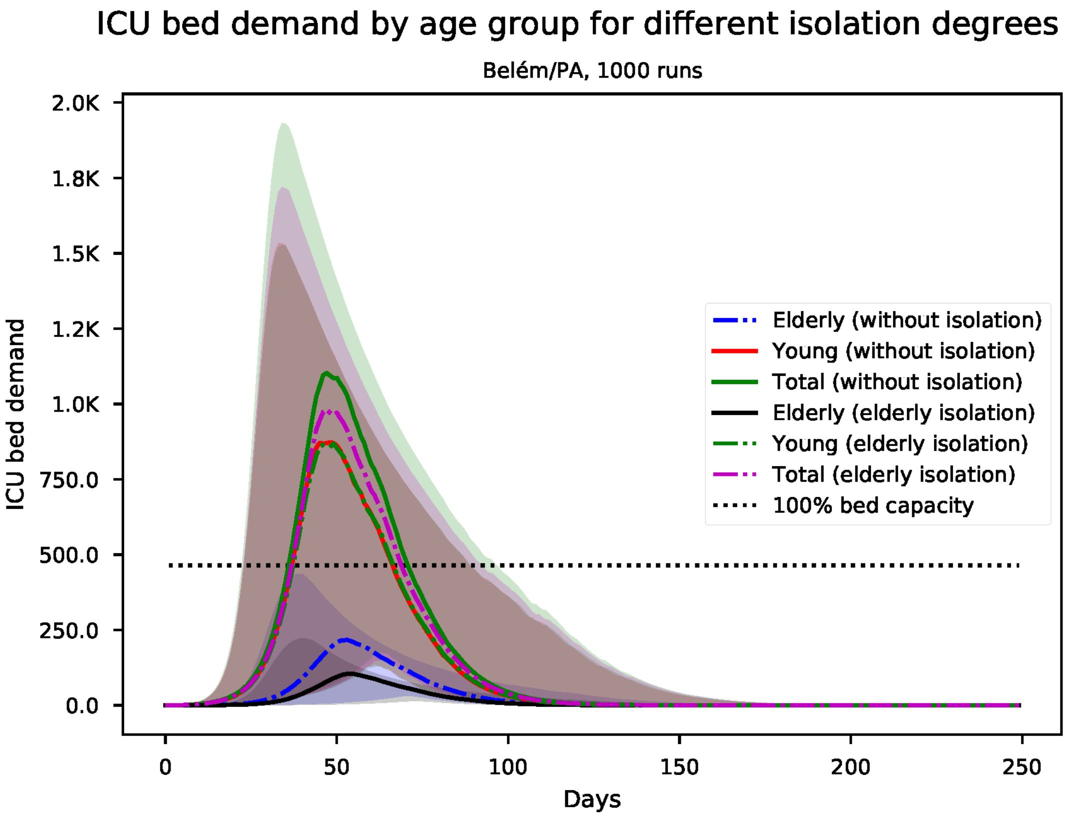


Figure S17: Predicted number of needed ICU beds, by age group for Belém

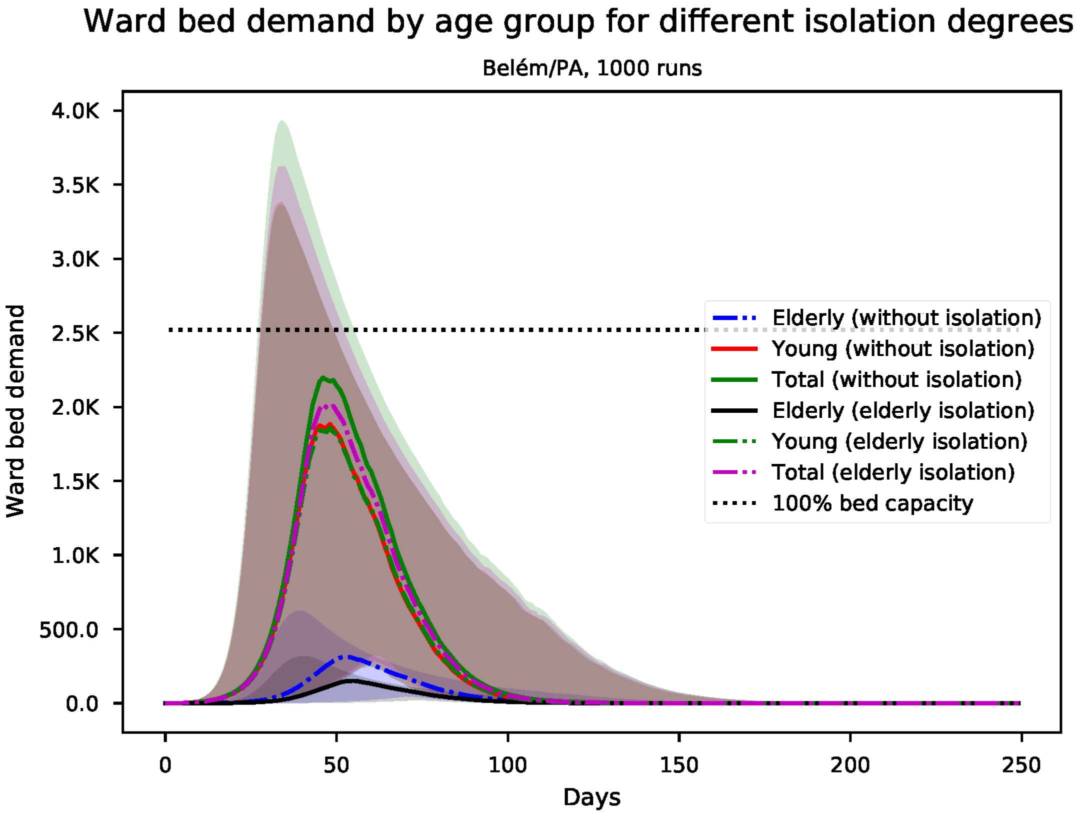


Figure S18: Predicted number of needed ward beds, by age group for Belém

*S4. Results for Boa Vista*

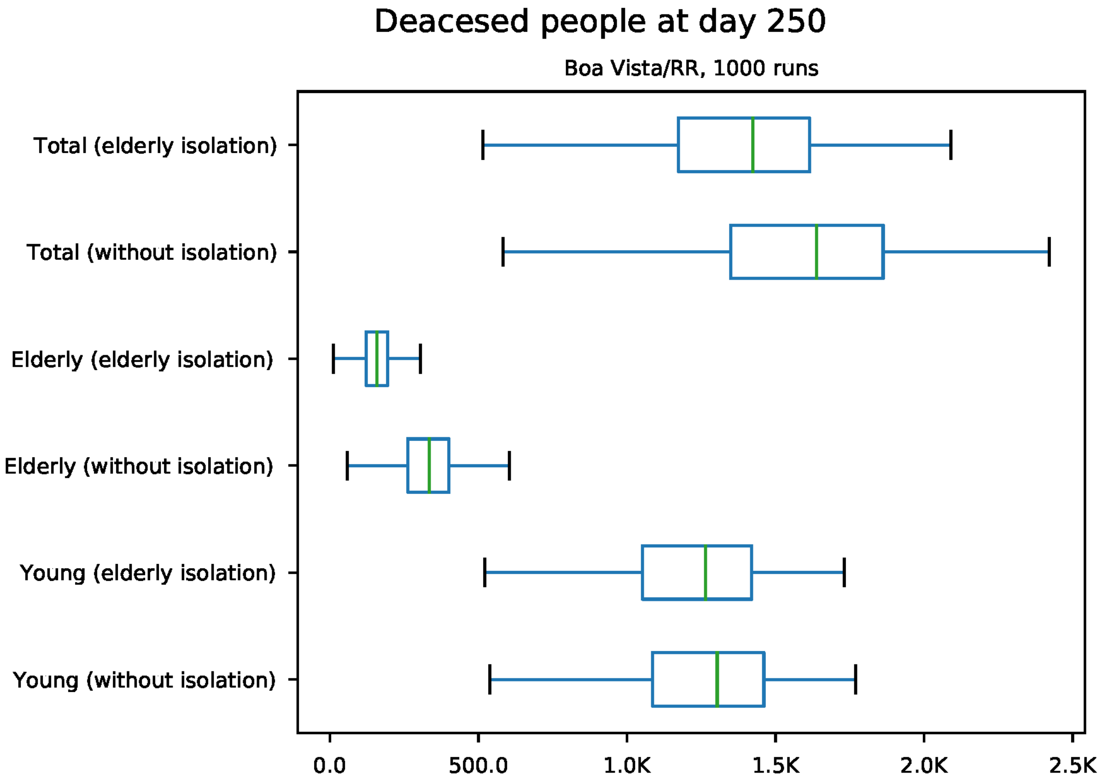


Figure S19: Predicted number of death at day 250 for Boa Vista

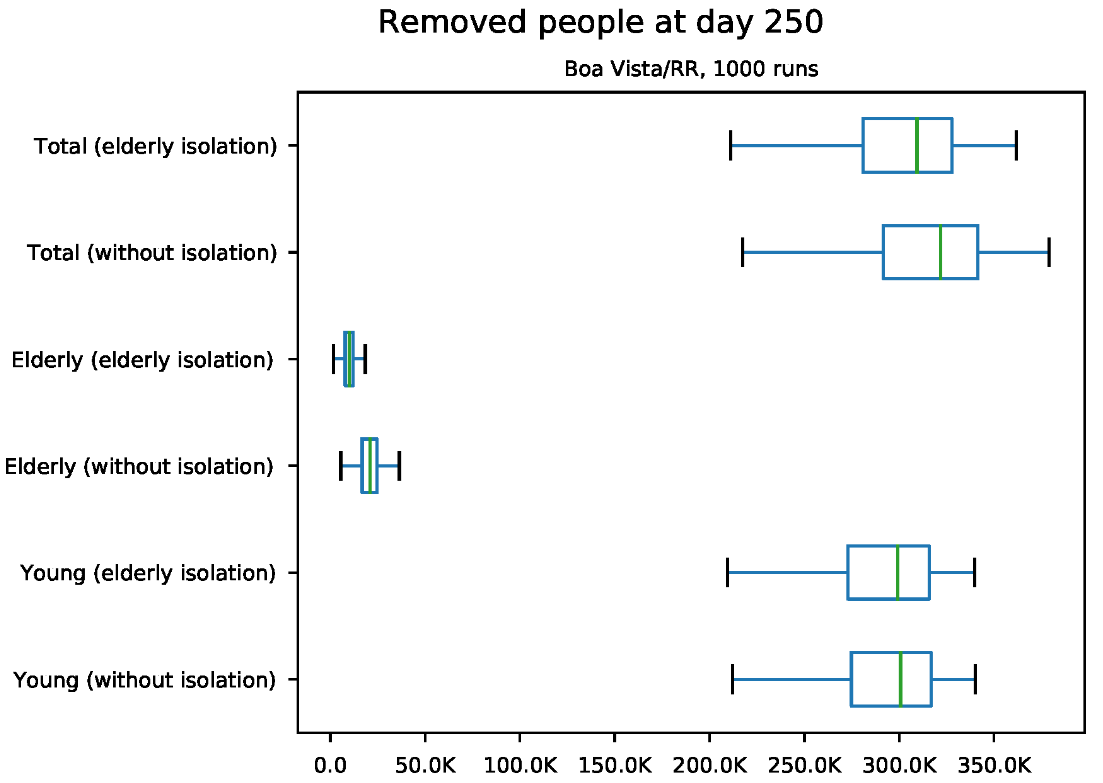


Figure S20: Predicted number of removed individuals at day 250 for Boa Vista

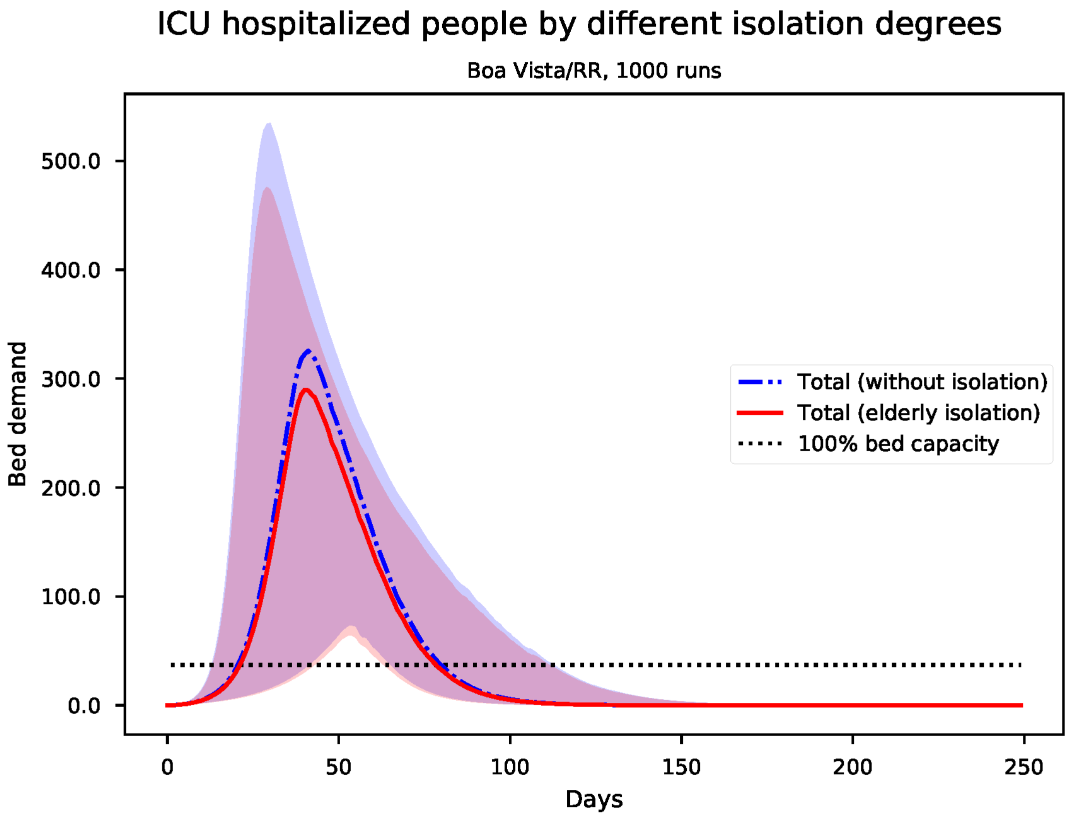


Figure S21: Predicted number of needed ICU beds for Boa Vista

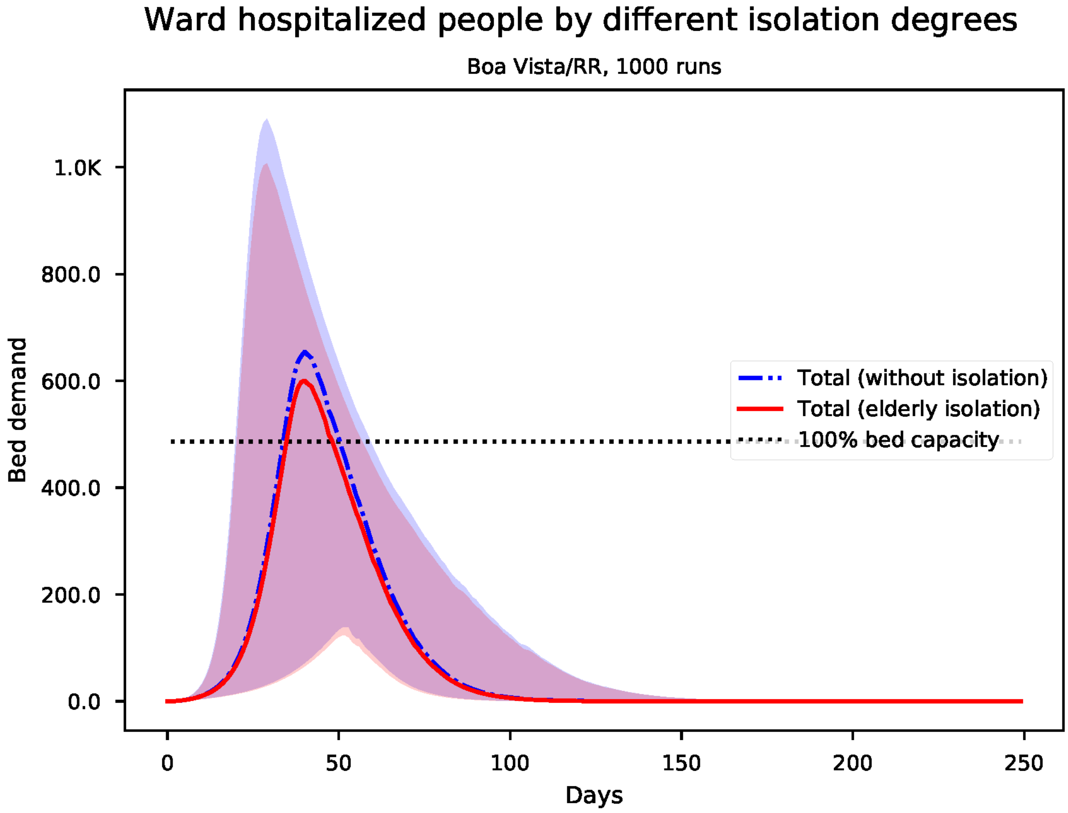


Figure S22: Predicted number of needed ward beds for Boa Vista

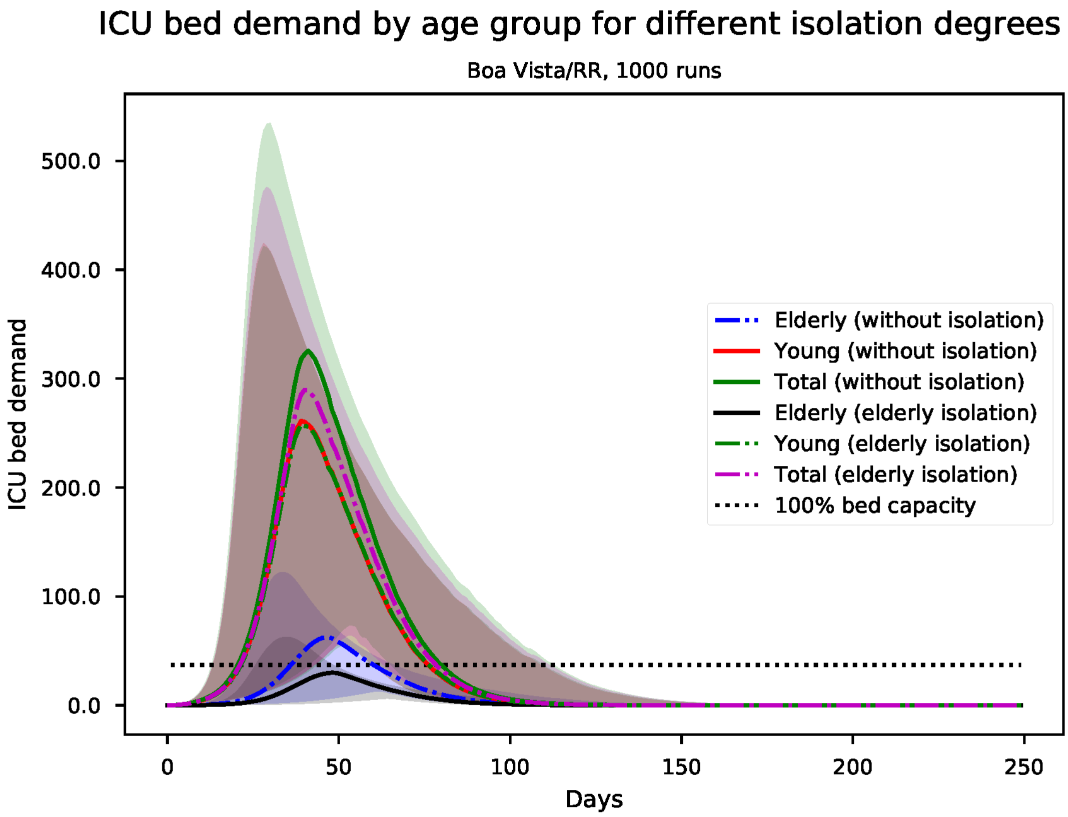


Figure S23: Predicted number of needed ICU beds, by age group for Boa Vista

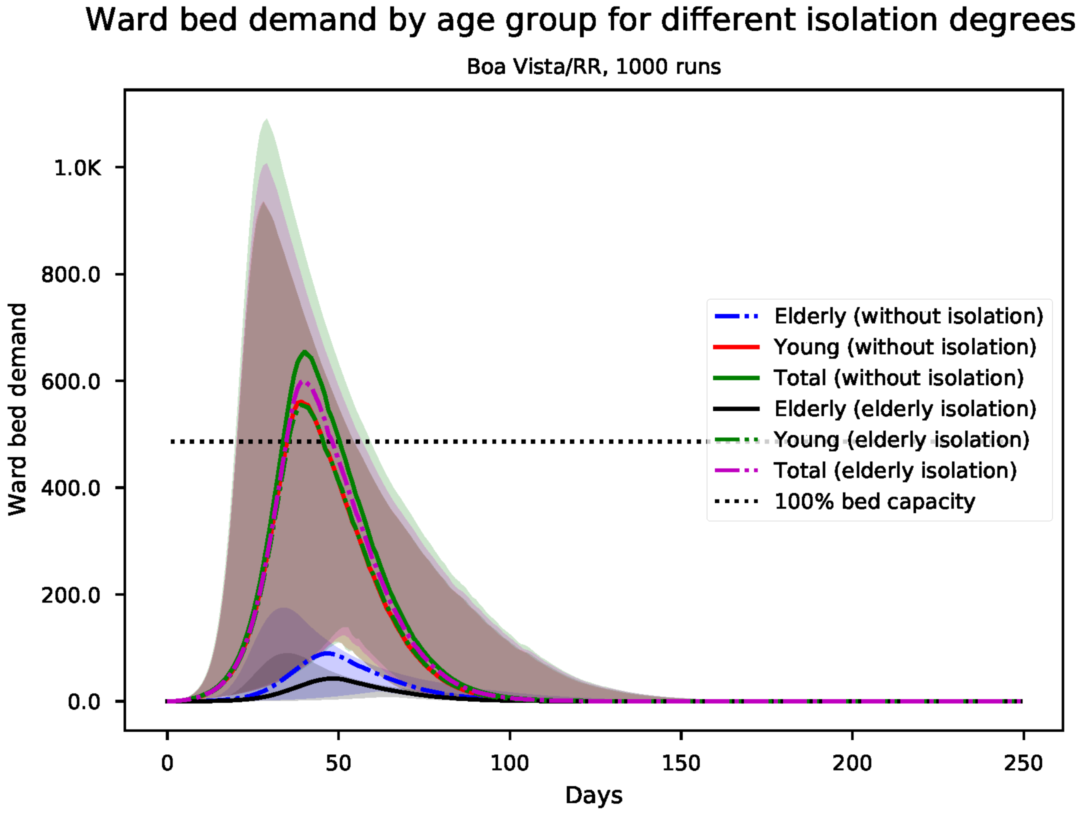


Figure S24: Predicted number of needed ward beds, by age group for Boa Vista

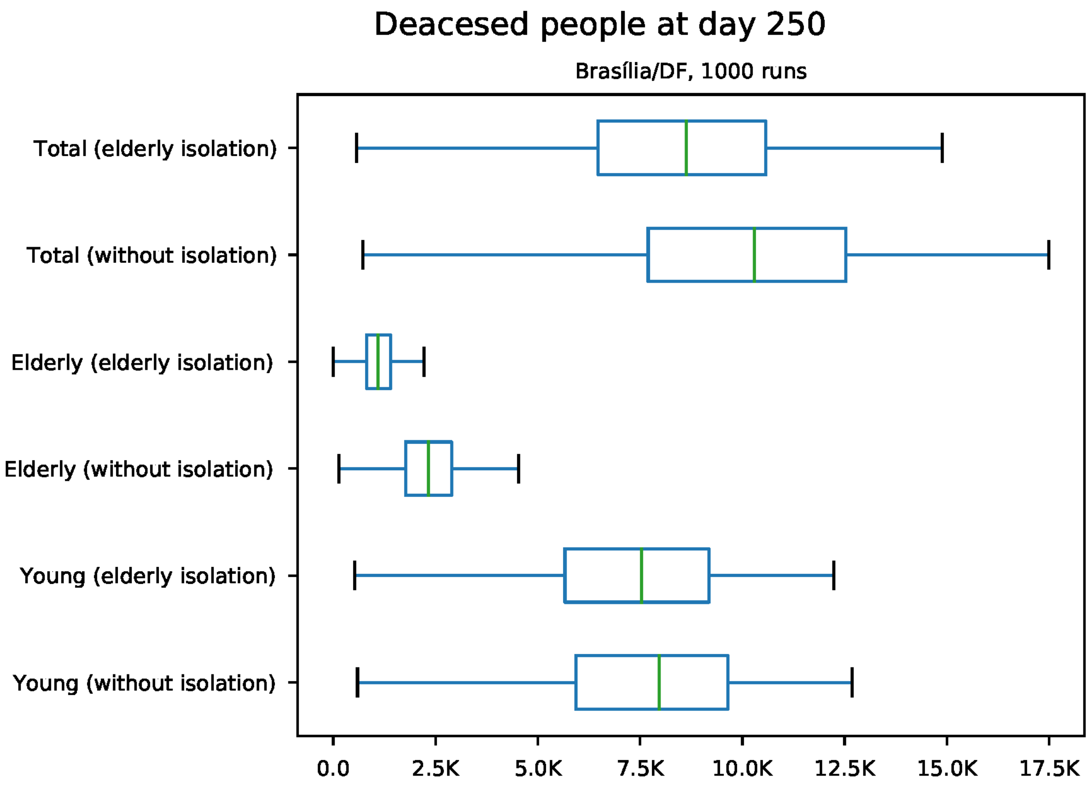


Figure S25: Predicted number of death at day 250 for Brasília

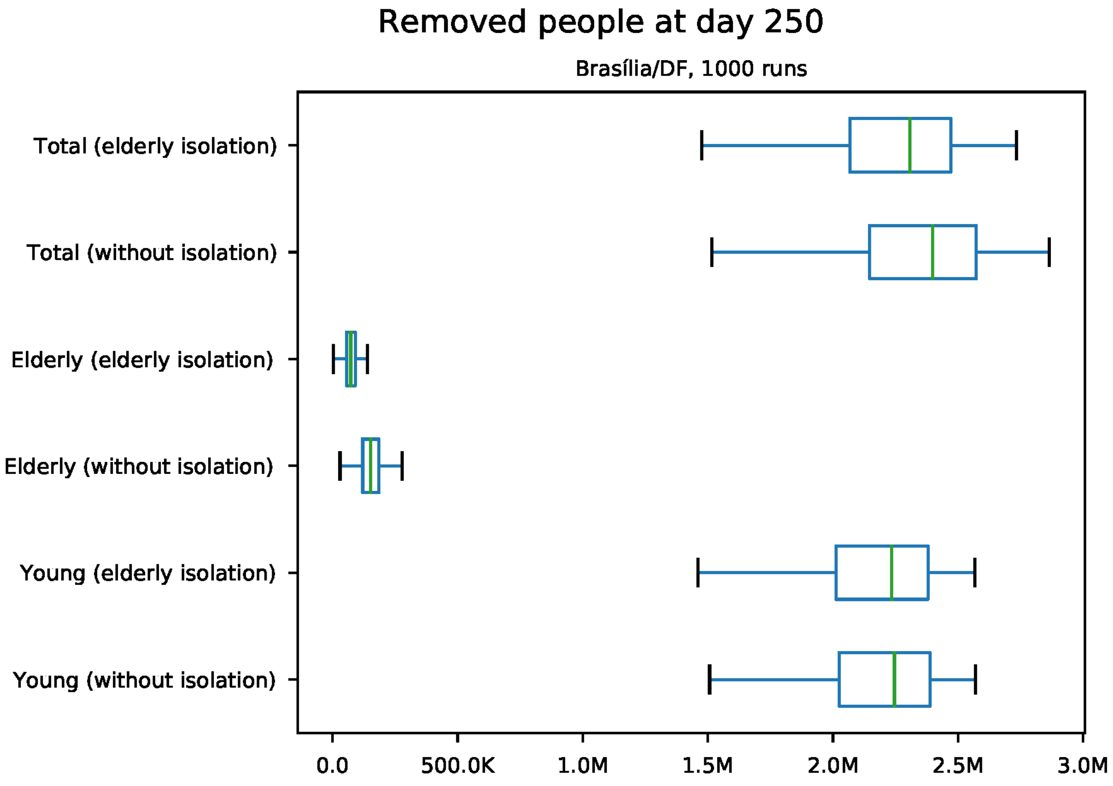


Figure S26: Predicted number of removed individuals at day 250 for Brasília

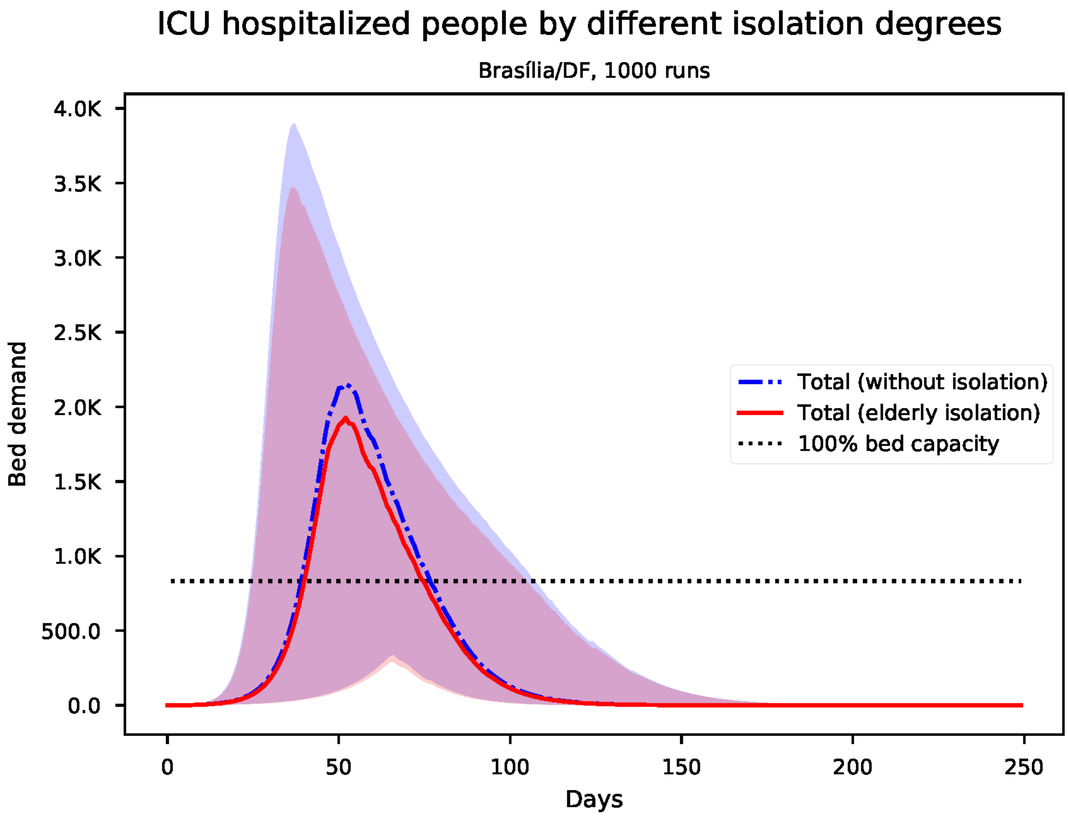


Figure S27: Predicted number of needed ICU beds for Brasília

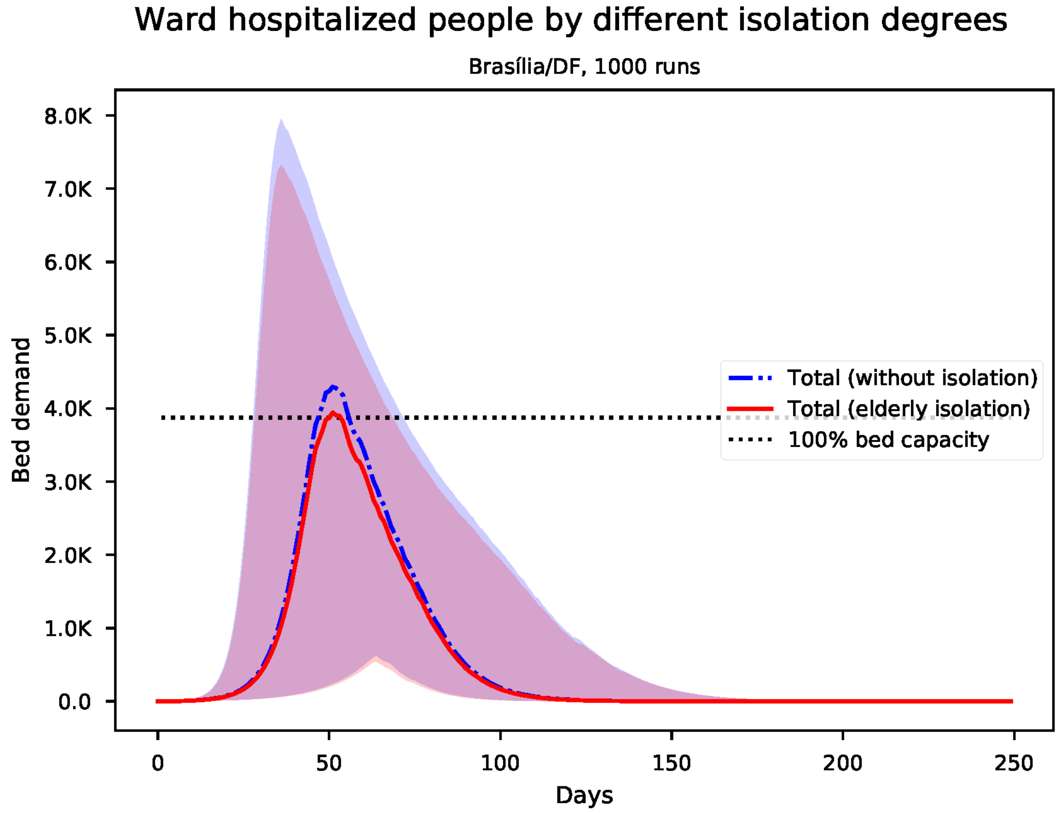


Figure S28: Predicted number of needed ward beds for Brasília

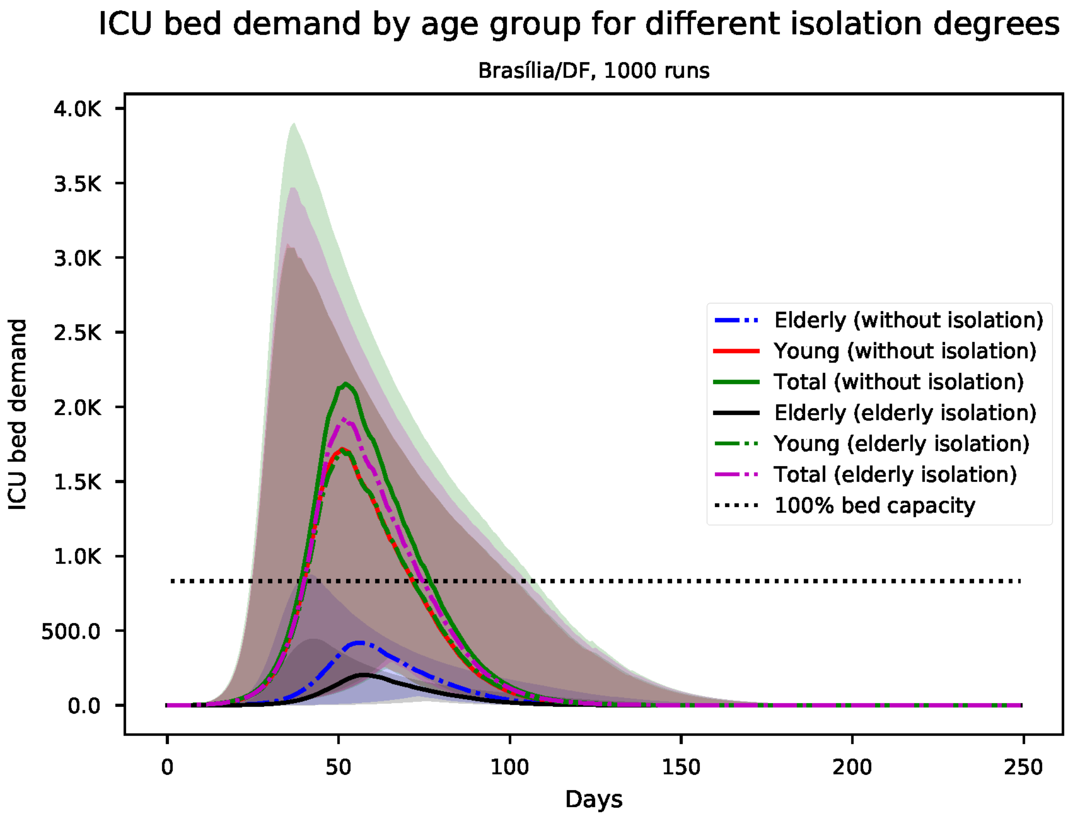


Figure S29: Predicted number of needed ICU beds, by age group for Brasília

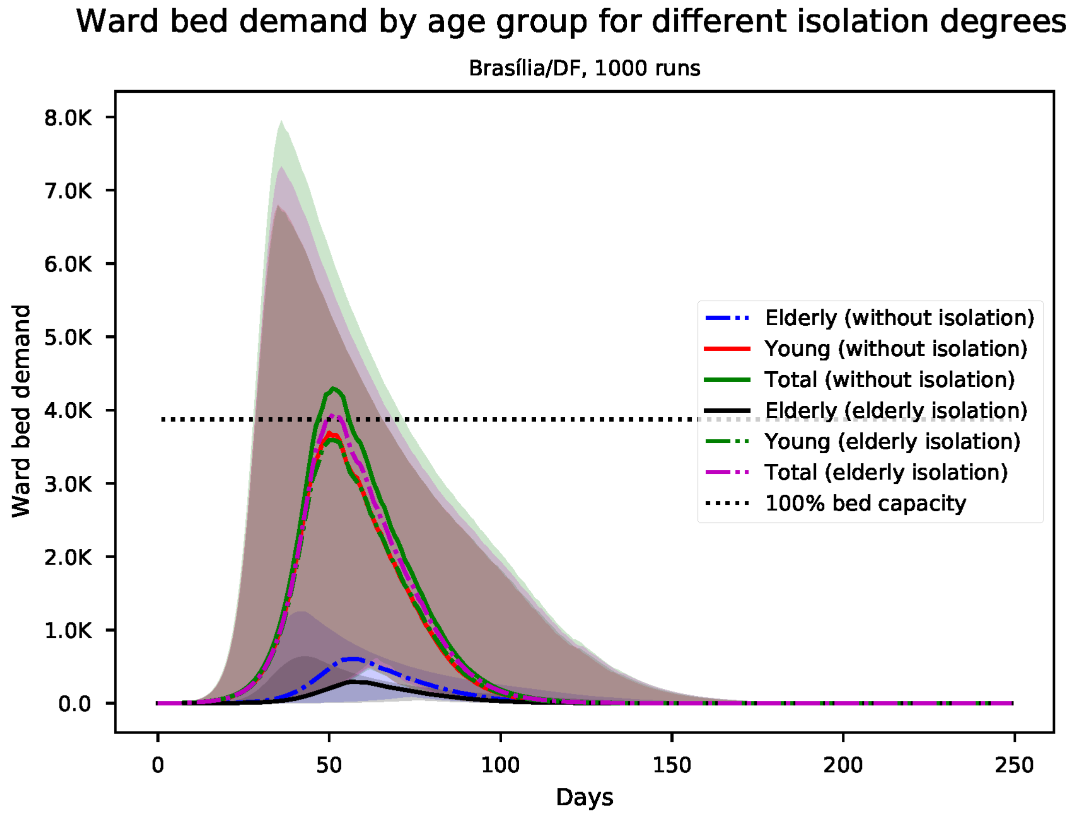


Figure S30: Predicted number of needed ward beds, by age group for Brasília

*S6. Results for Campo Grande*

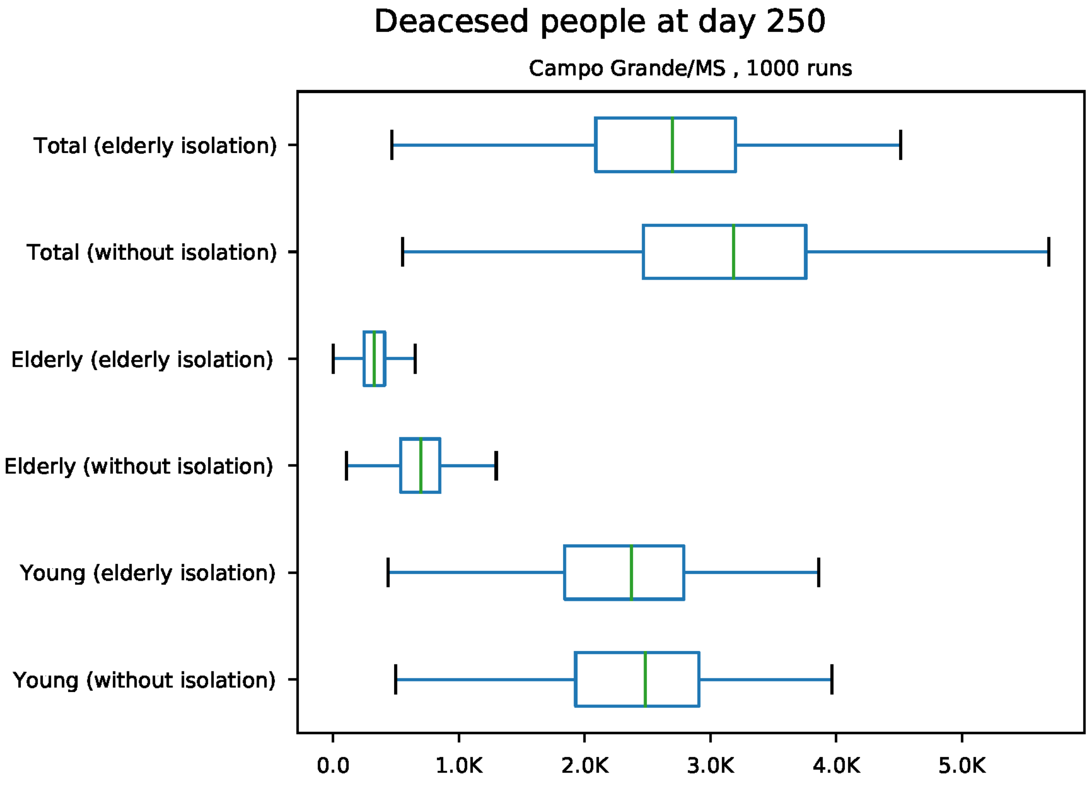


Figure S31: Predicted number of death at day 250 for Campo Grande

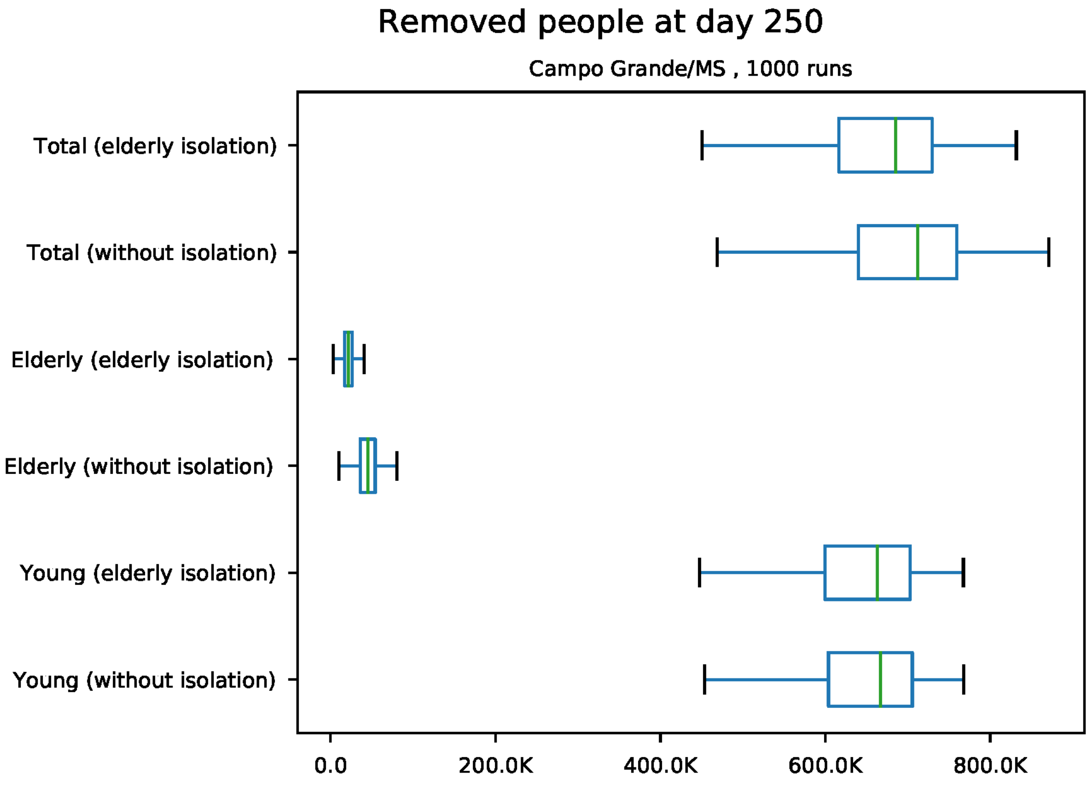


Figure S32: Predicted number of removed individuals at day 250 for Campo Grande

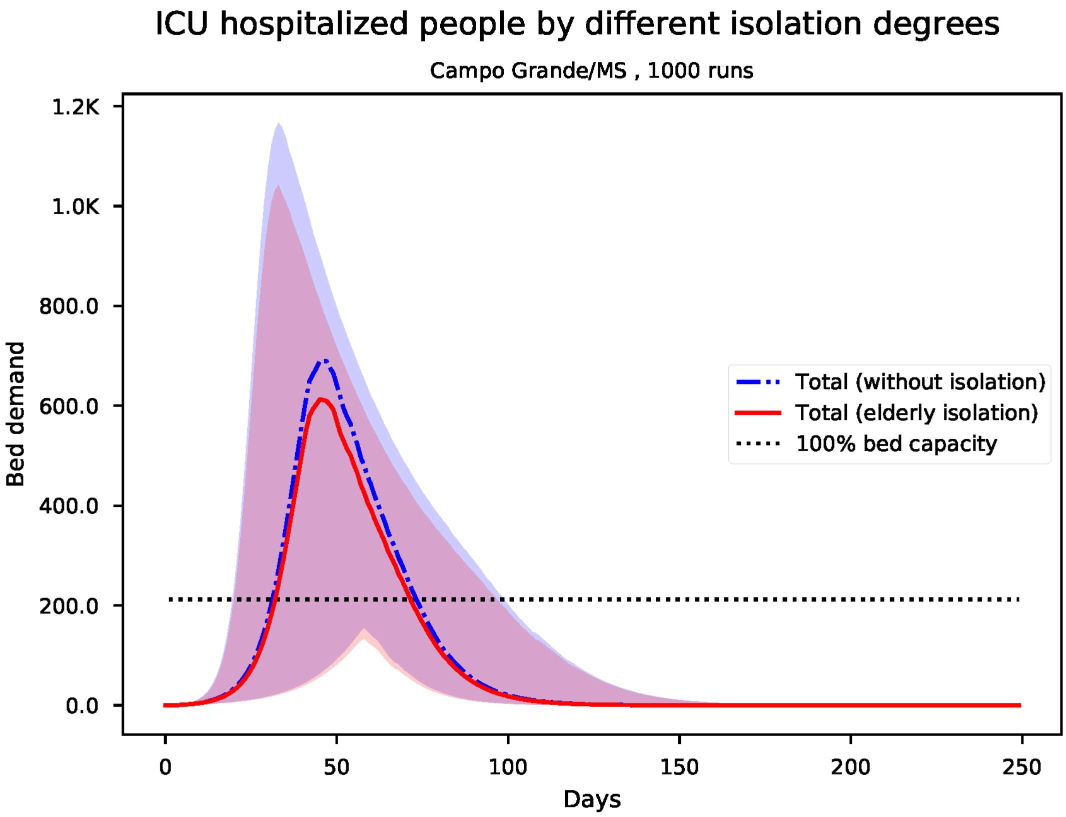


Figure S33: Predicted number of needed ICU beds for Campo Grande

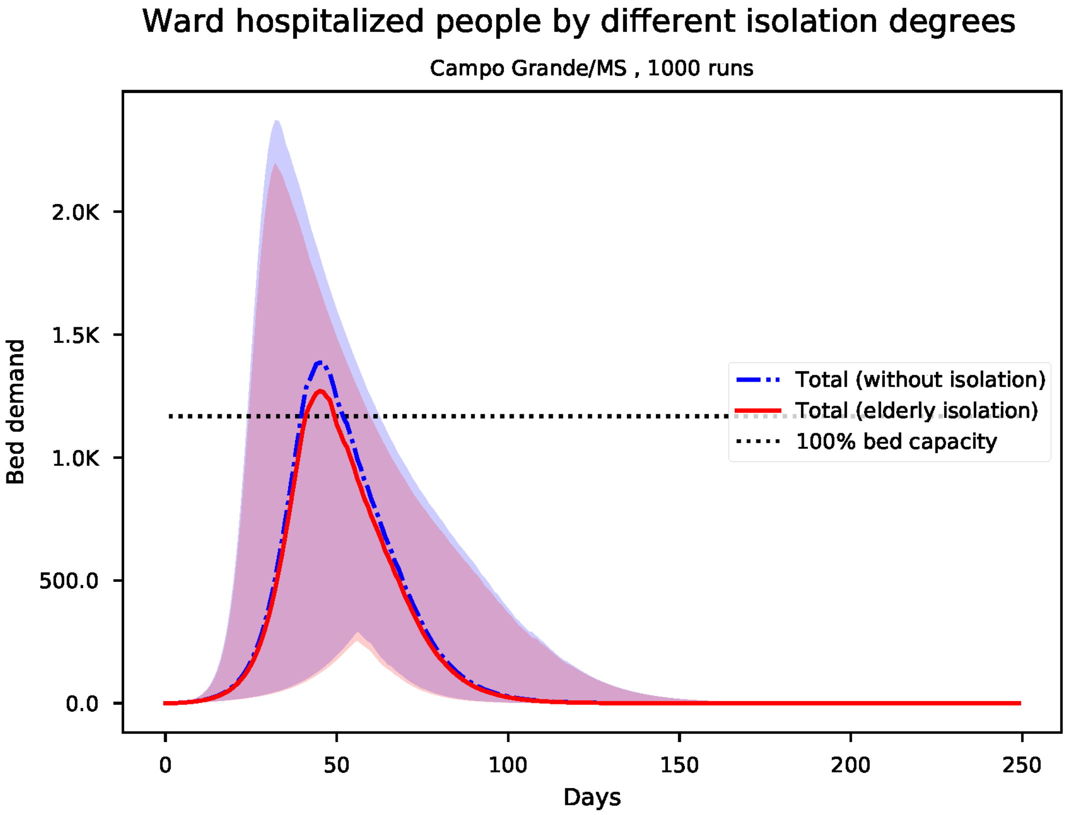


Figure S34: Predicted number of needed ward beds for Campo Grande

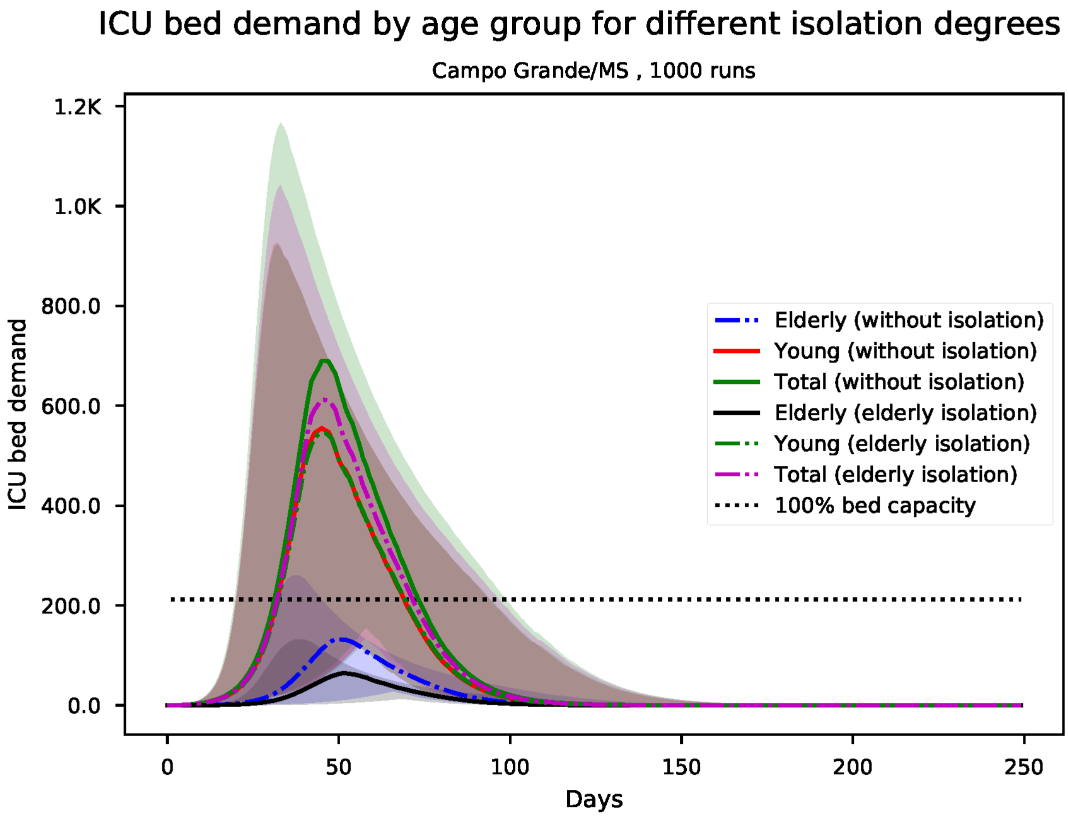


Figure S35: Predicted number of needed ICU beds, by age group for Campo Grande

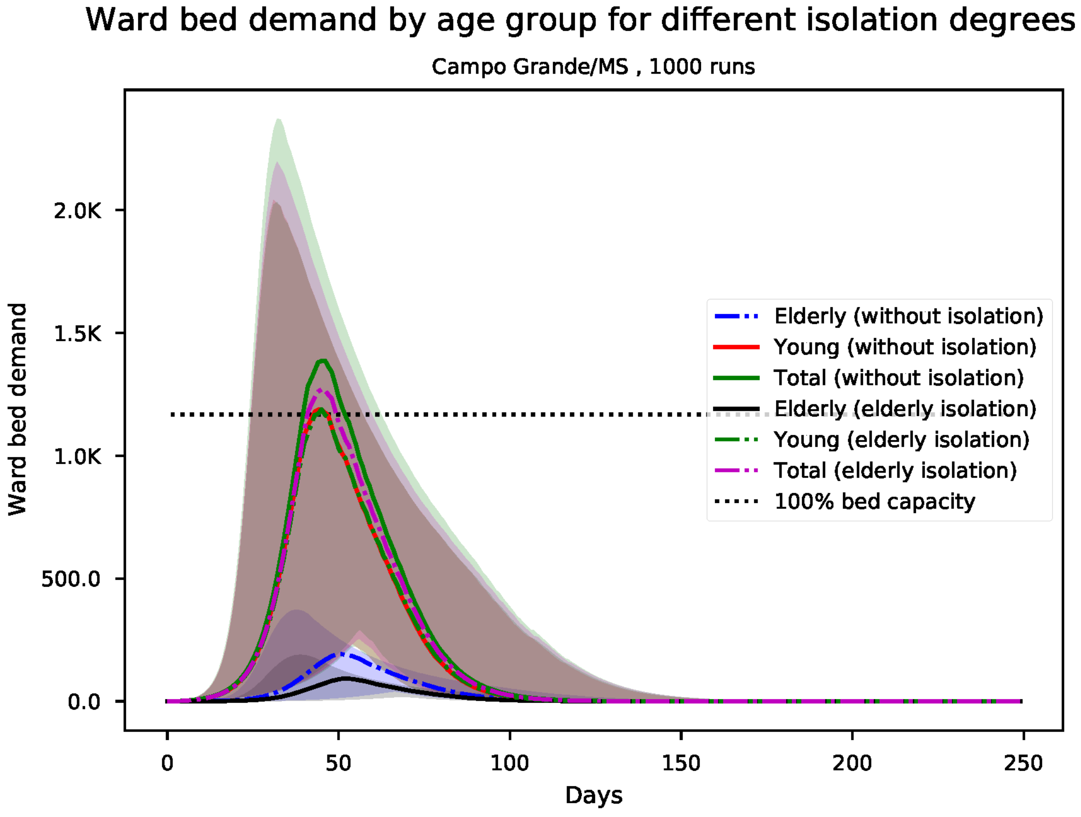


Figure S36: Predicted number of needed ward beds, by age group for Campo Grande

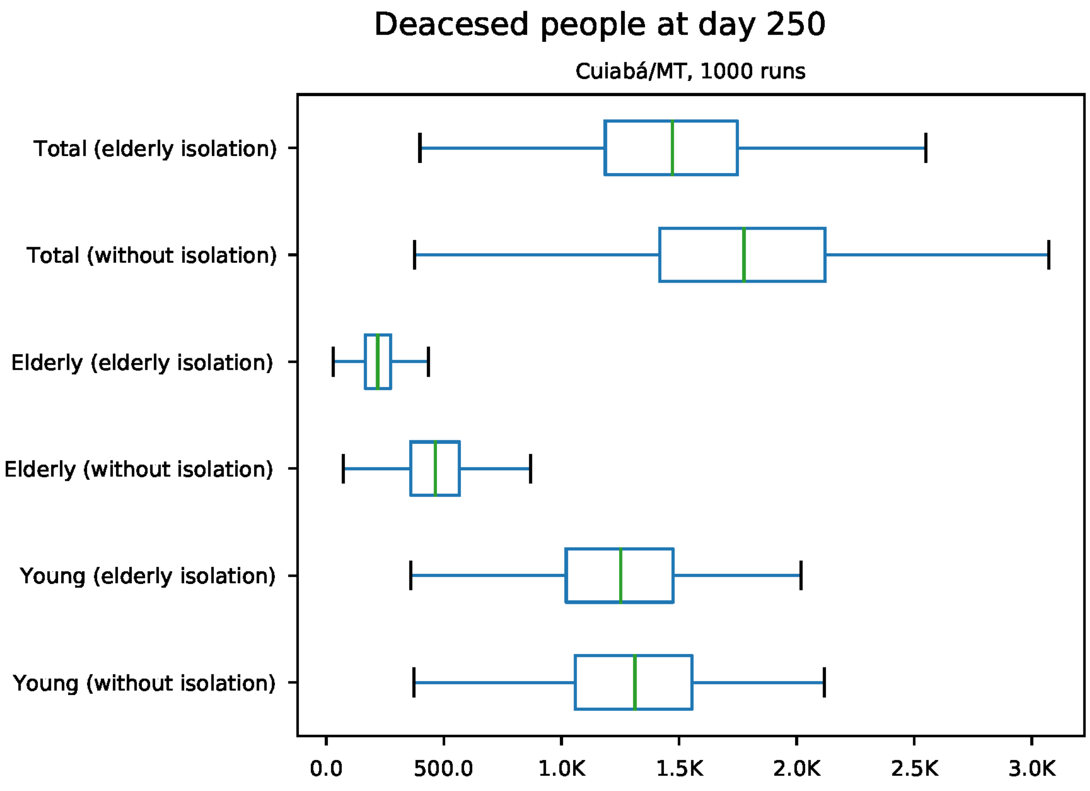


Figure S37: Predicted number of death at day 250 for Cuiabá

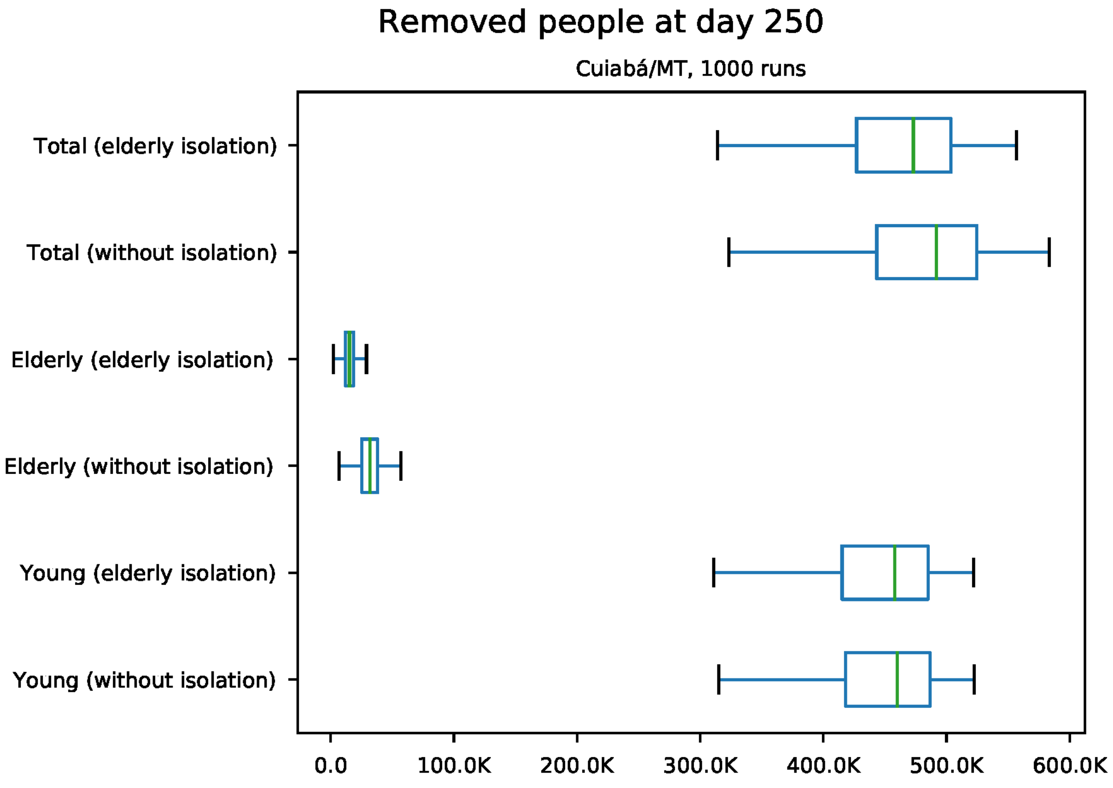


Figure S38: Predicted number of removed individuals at day 250 for Cuiabá

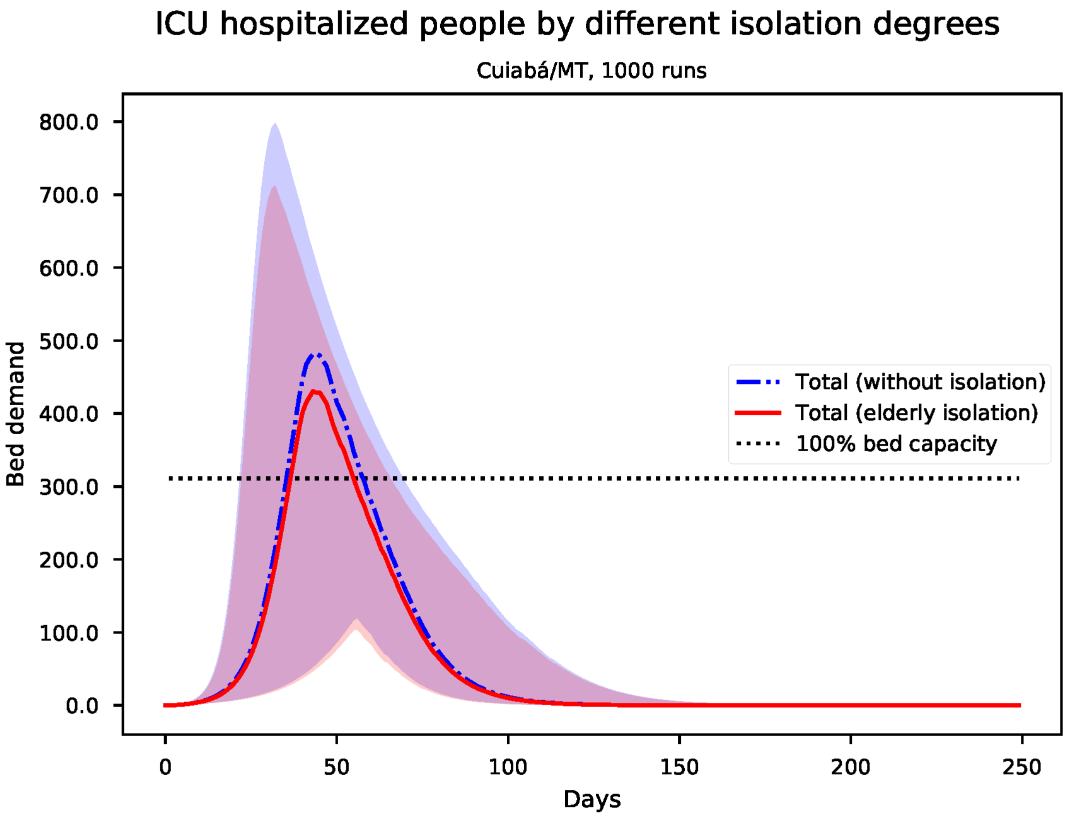


Figure S39: Predicted number of needed ICU beds for Cuiabá

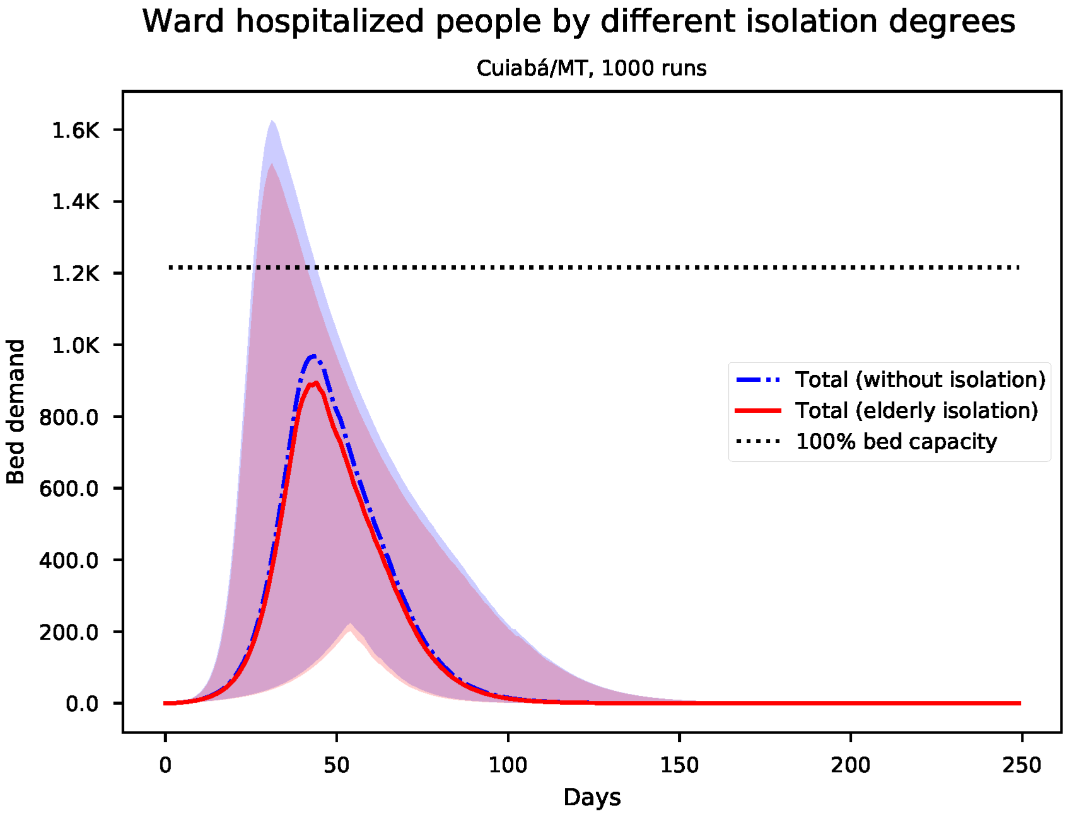


Figure S40: Predicted number of needed ward beds for Cuiabá

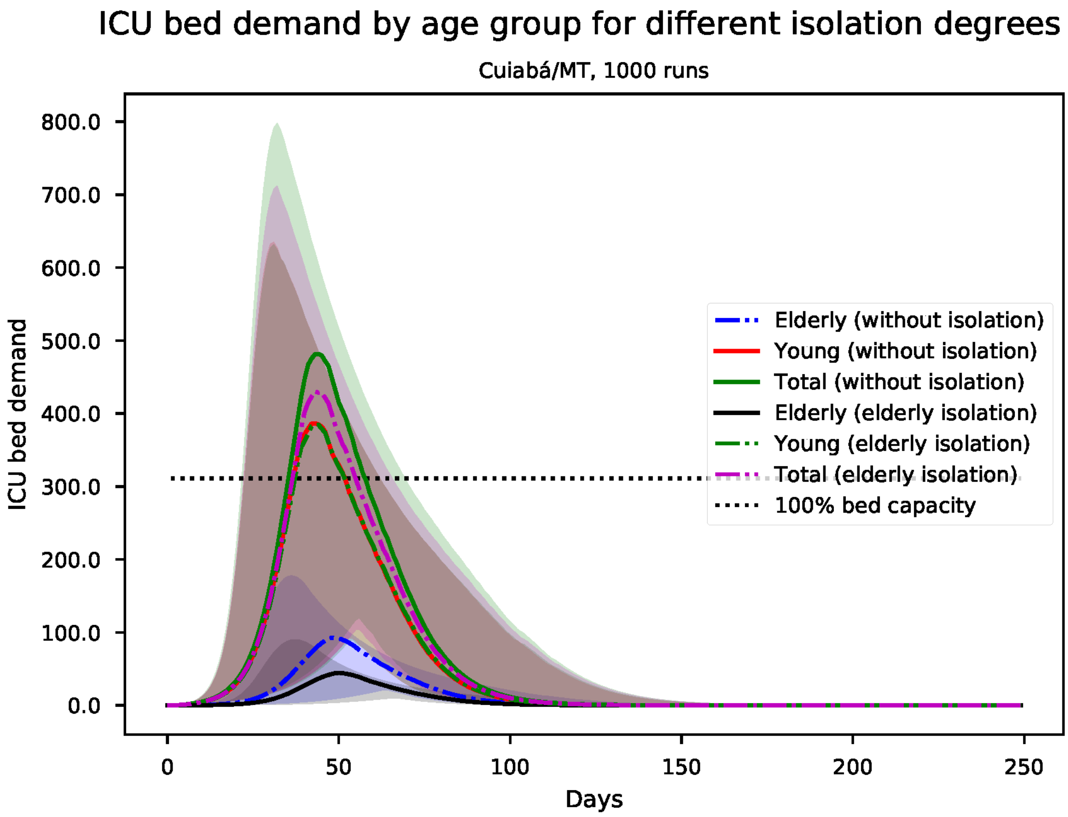


Figure S41: Predicted number of needed ICU beds, by age group for Cuiabá

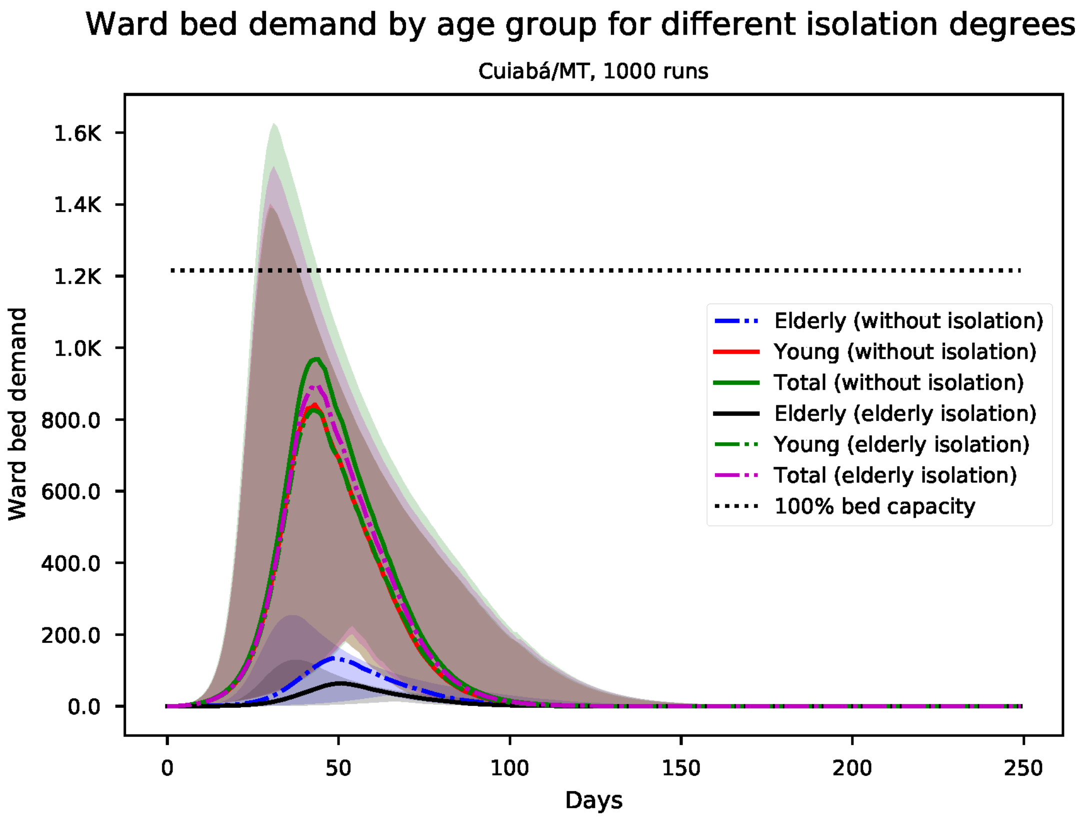


Figure S42: Predicted number of needed ward beds, by age group for Cuiabá

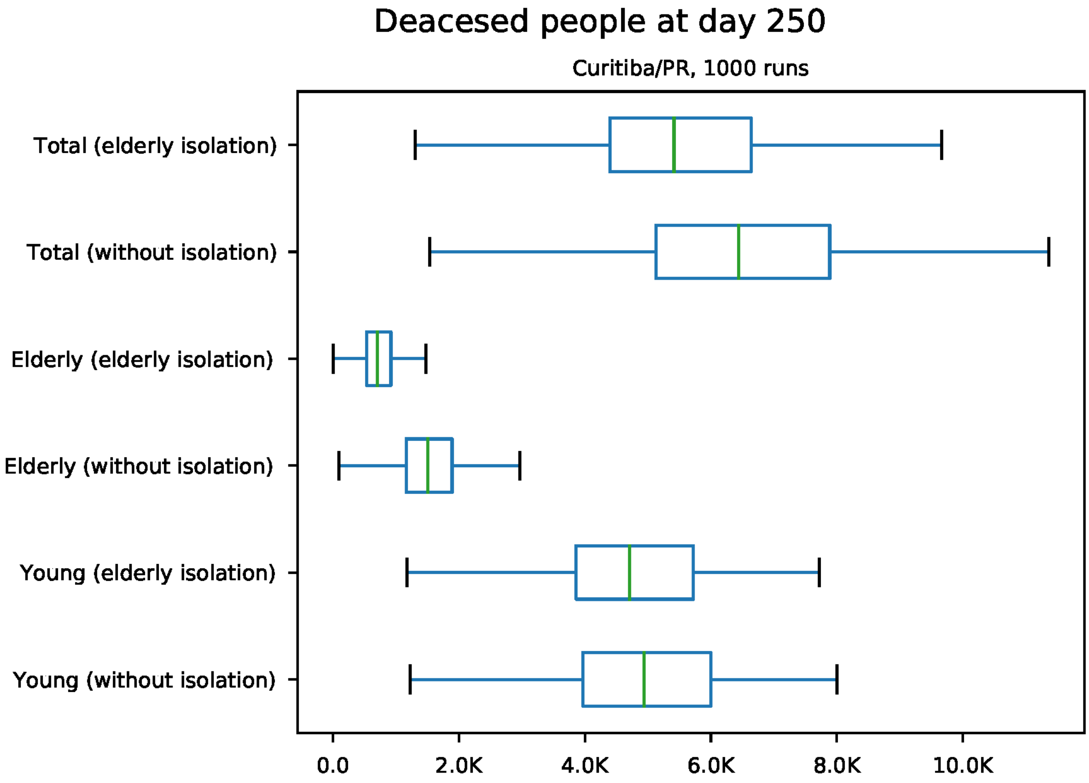


Figure S43: Predicted number of death at day 250 for Curitiba

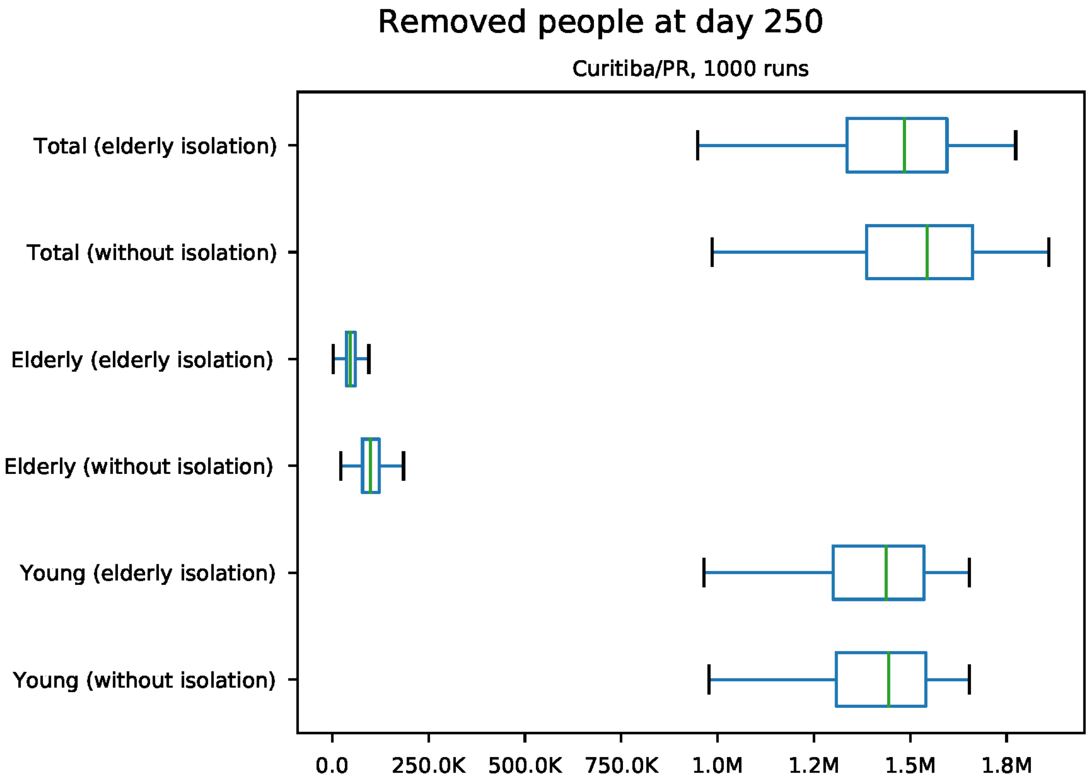


Figure S44: Predicted number of removed individuals at day 250 for Curitiba

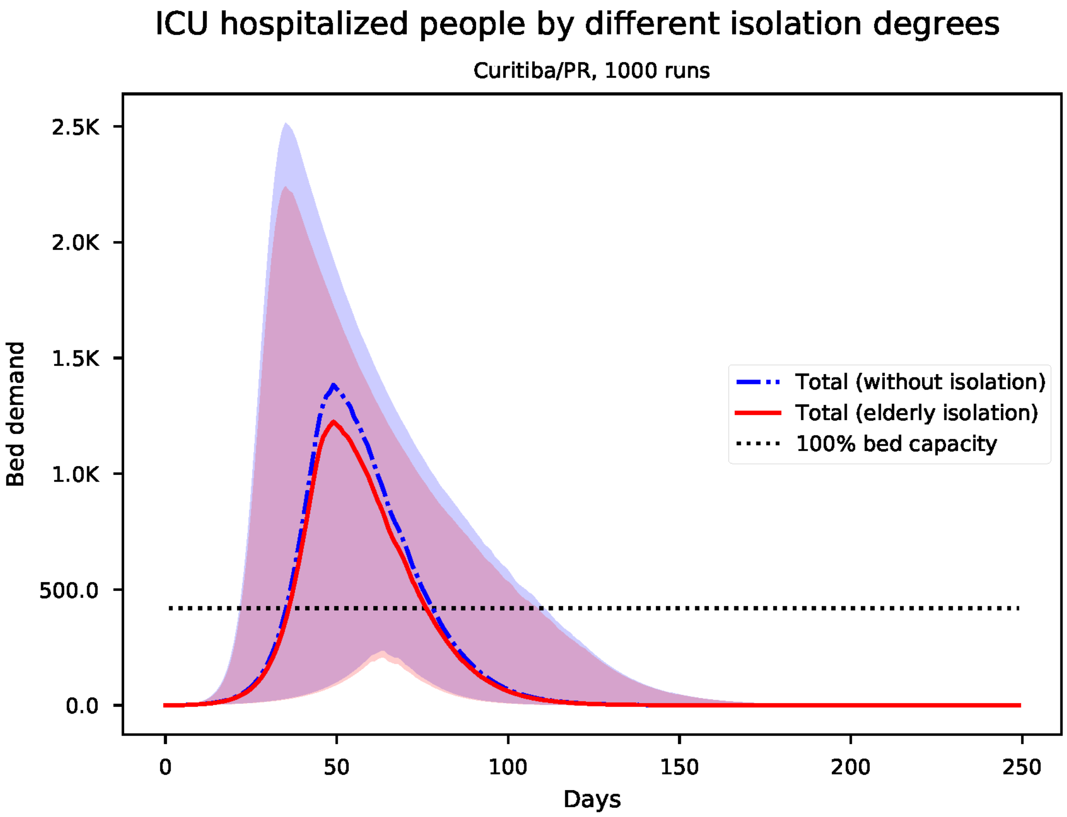


Figure S45: Predicted number of needed ICU beds for Curitiba

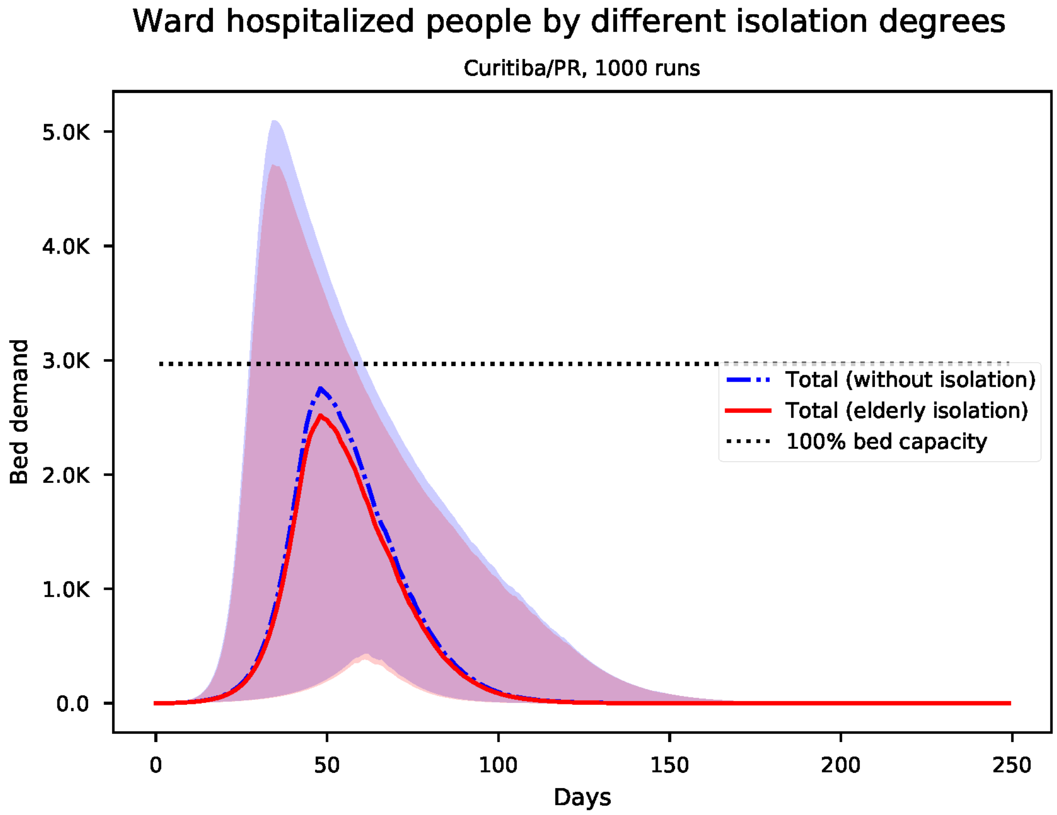


Figure S46: Predicted number of needed ward beds for Curitiba

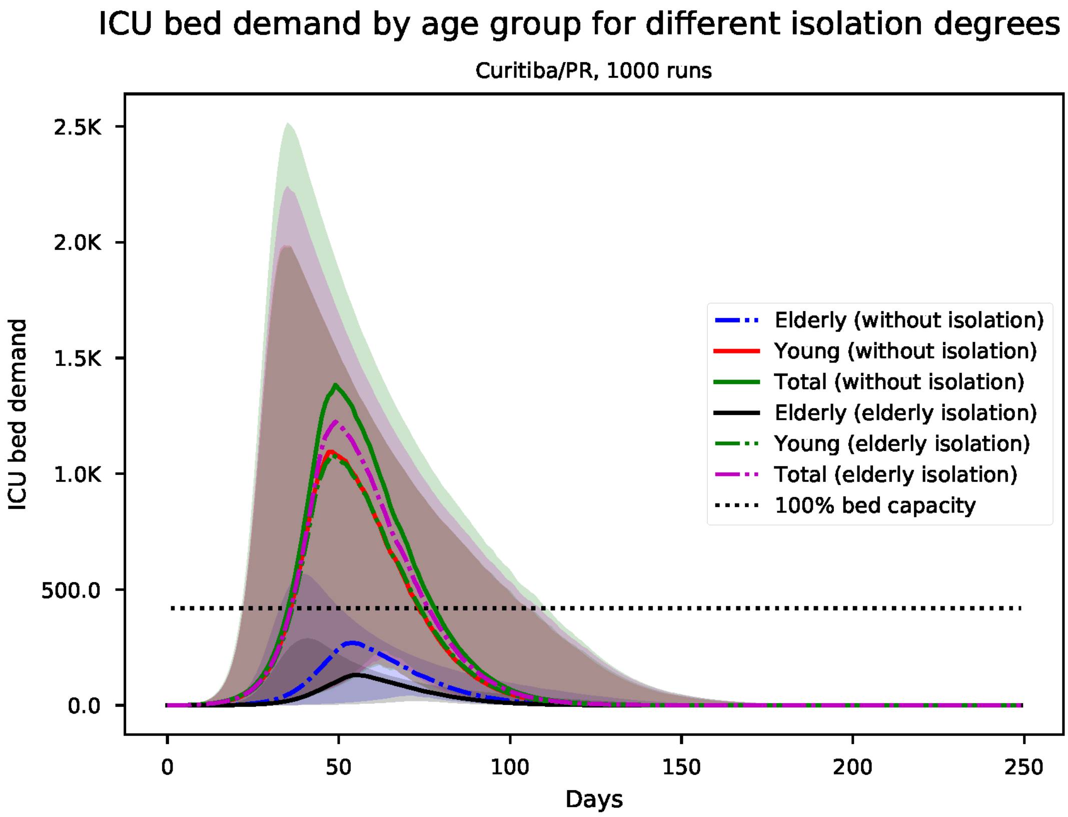


Figure S47: Predicted number of needed ICU beds, by age group for Curitiba

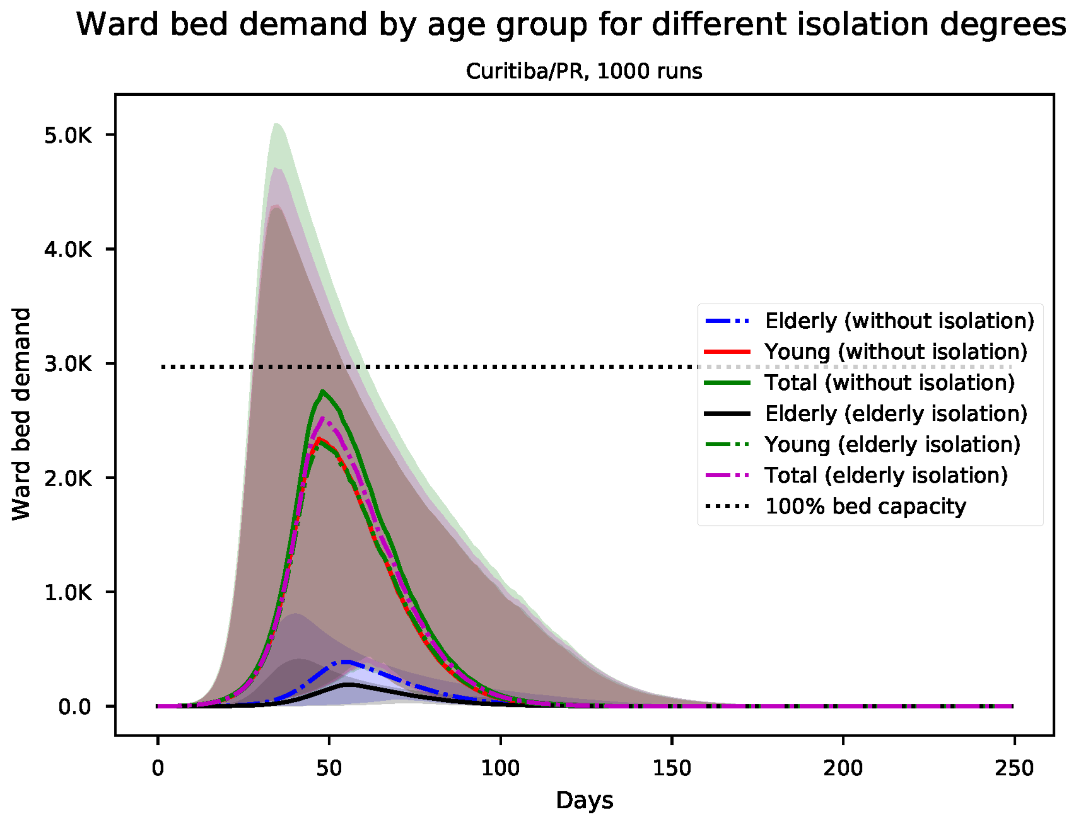


Figure S48: Predicted number of needed ward beds, by age group for Curitiba

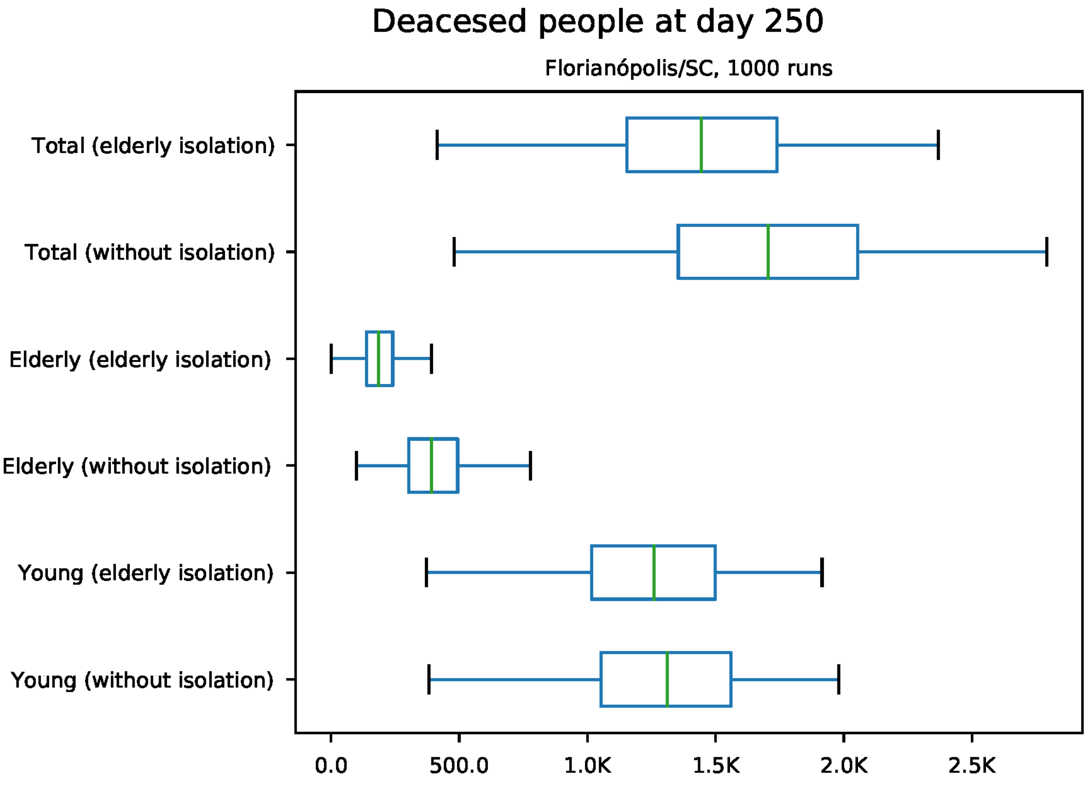


Figure S49: Predicted number of death at day 250 for Florianópolis

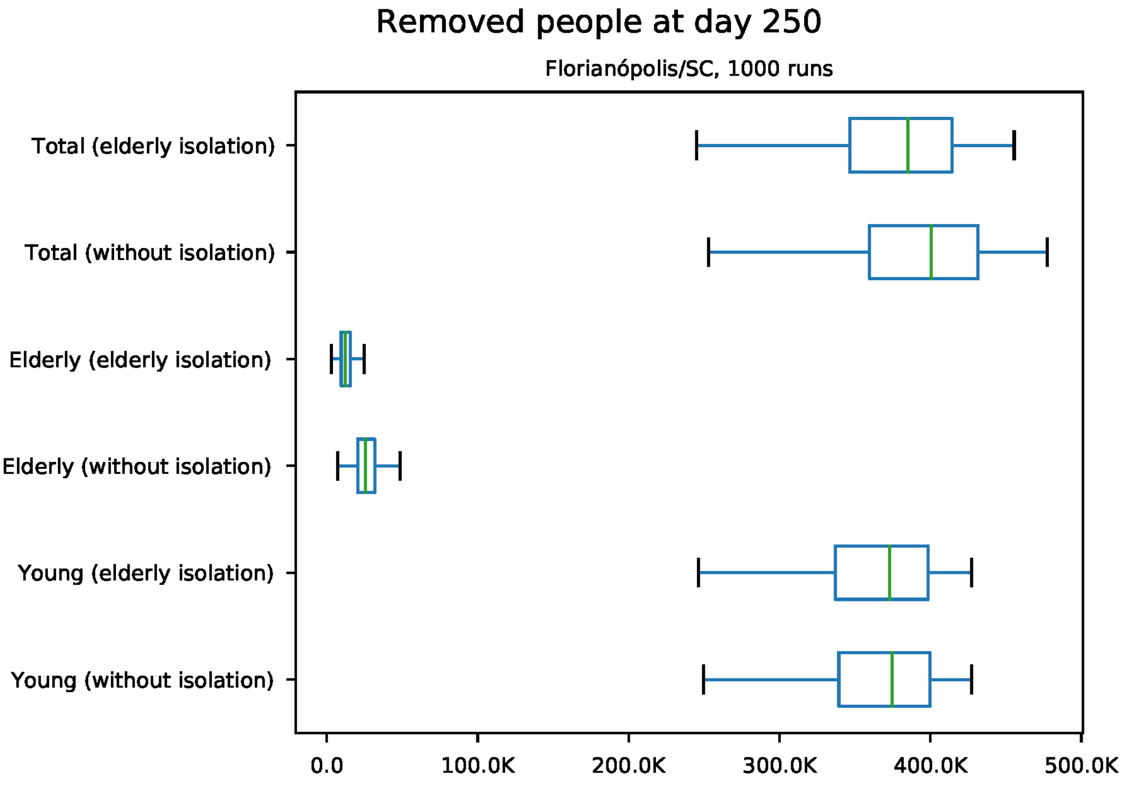


Figure S50: Predicted number of removed individuals at day 250 for Florianópolis

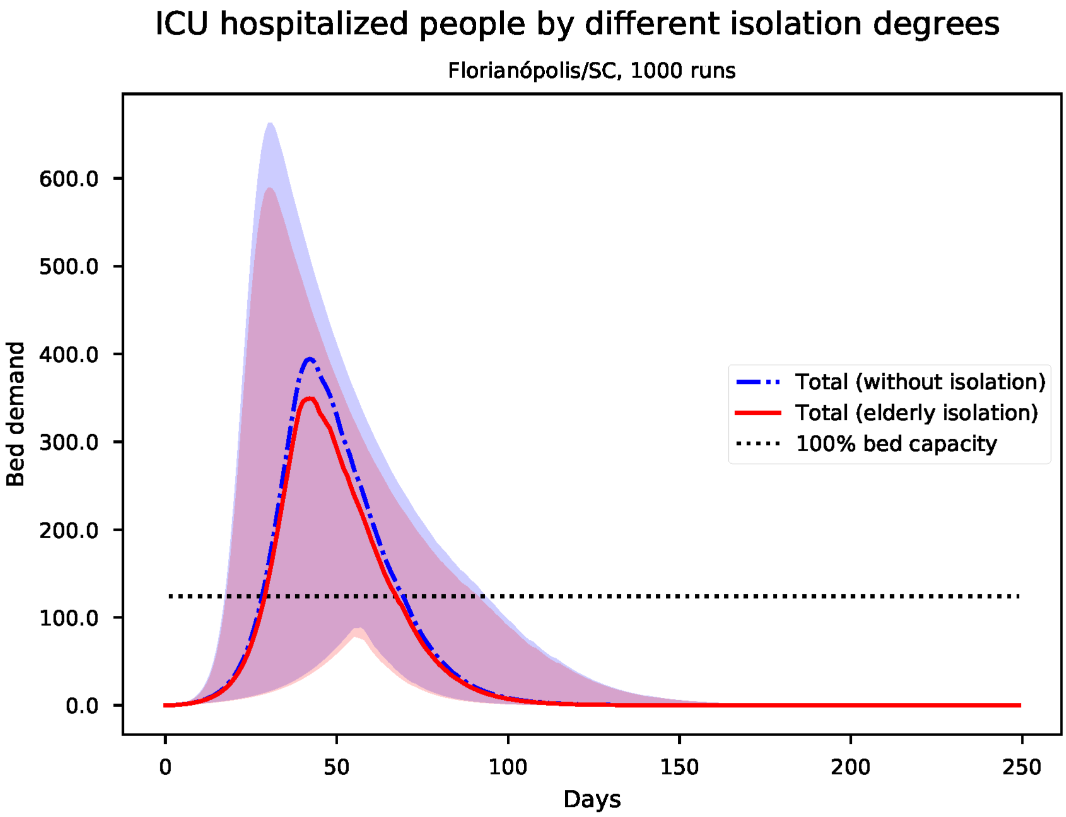


Figure S51: Predicted number of needed ICU beds for Florianópolis

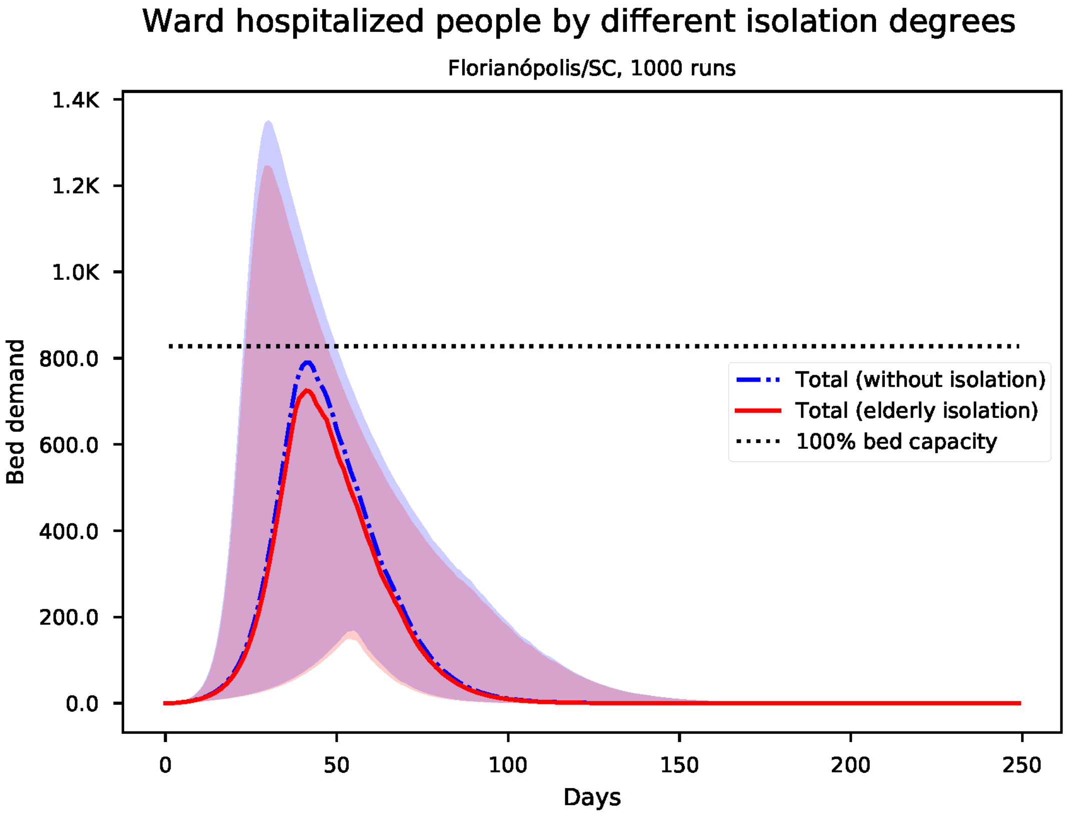


Figure S52: Predicted number of needed ward beds for Florianópolis

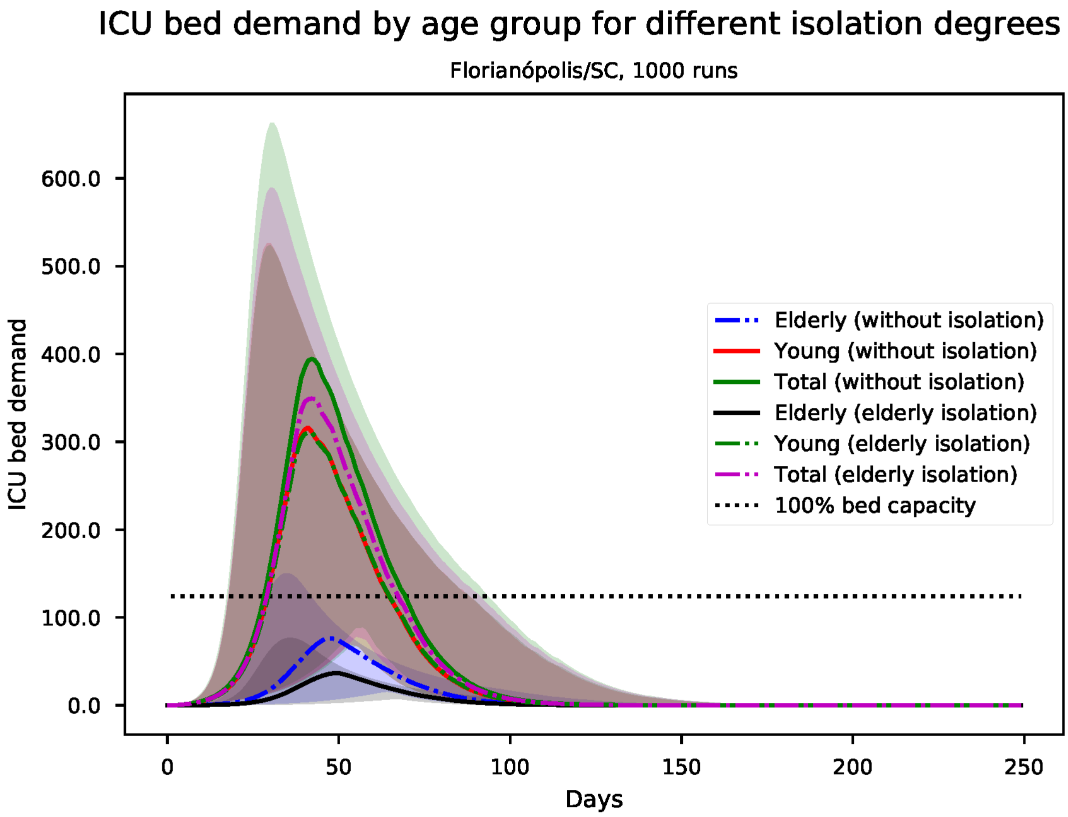


Figure S53: Predicted number of needed ICU beds, by age group for Florianópolis

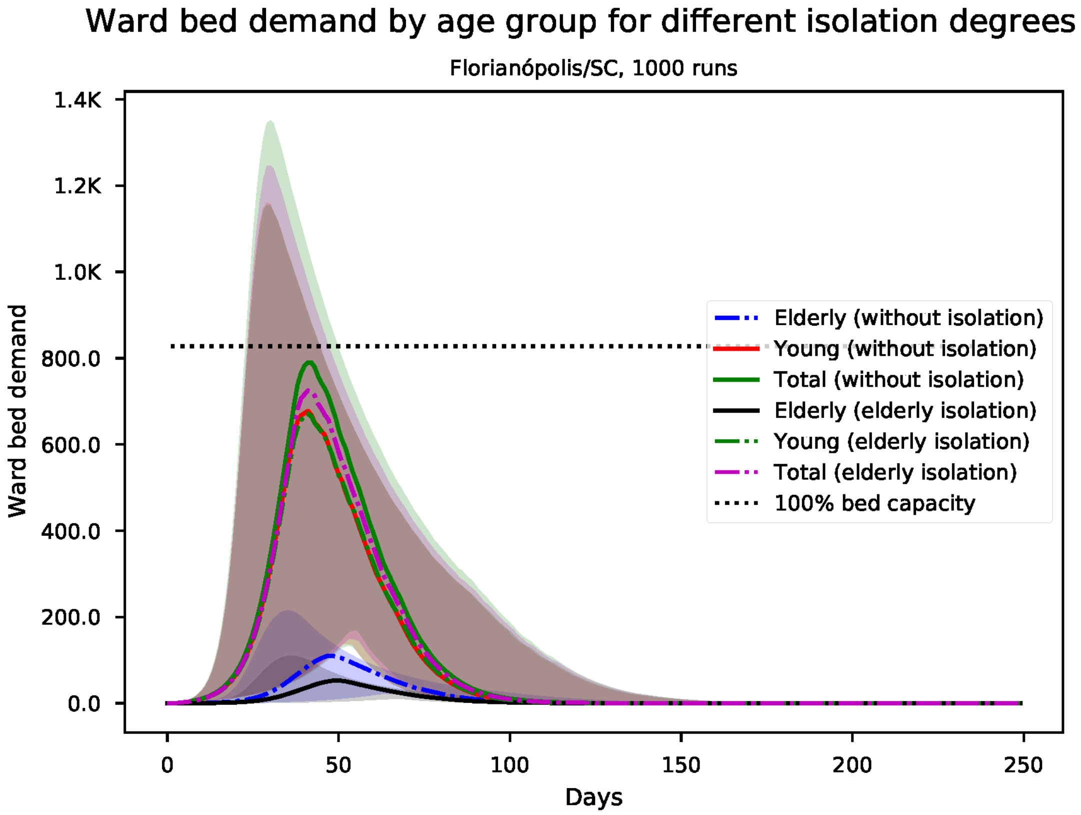


Figure S54: Predicted number of needed ward beds, by age group for Florianópolis

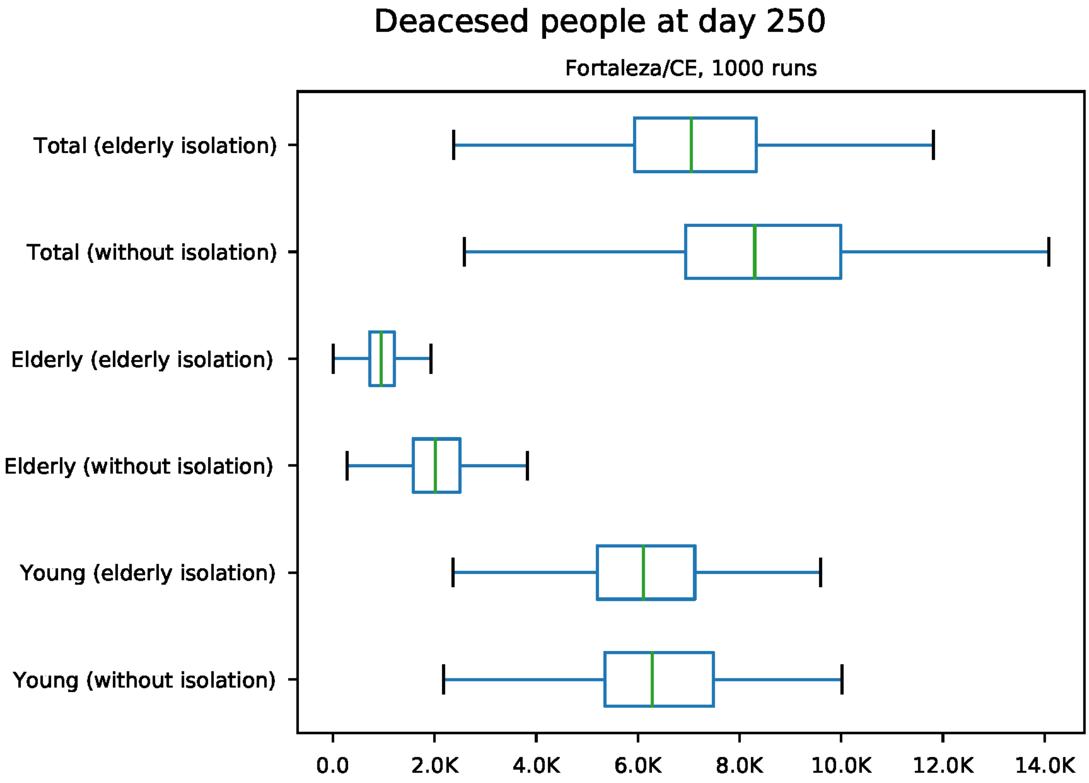


Figure S55: Predicted number of death at day 250 for Fortaleza

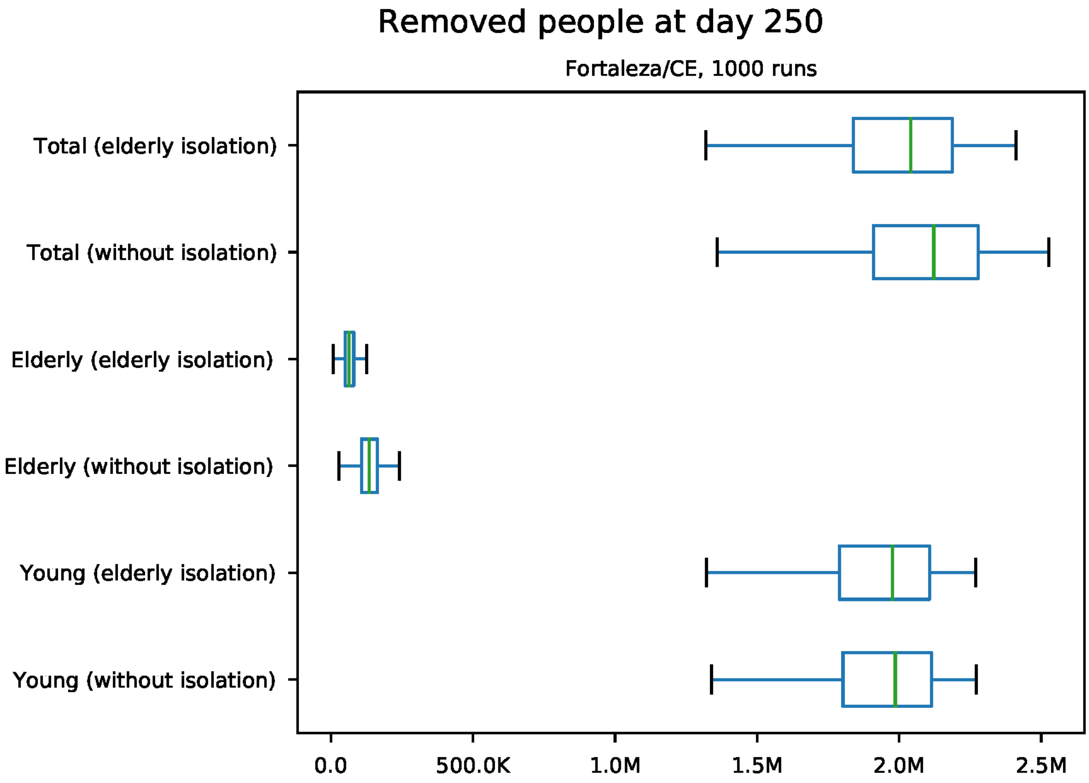


Figure S56: Predicted number of removed individuals at day 250 for Fortaleza

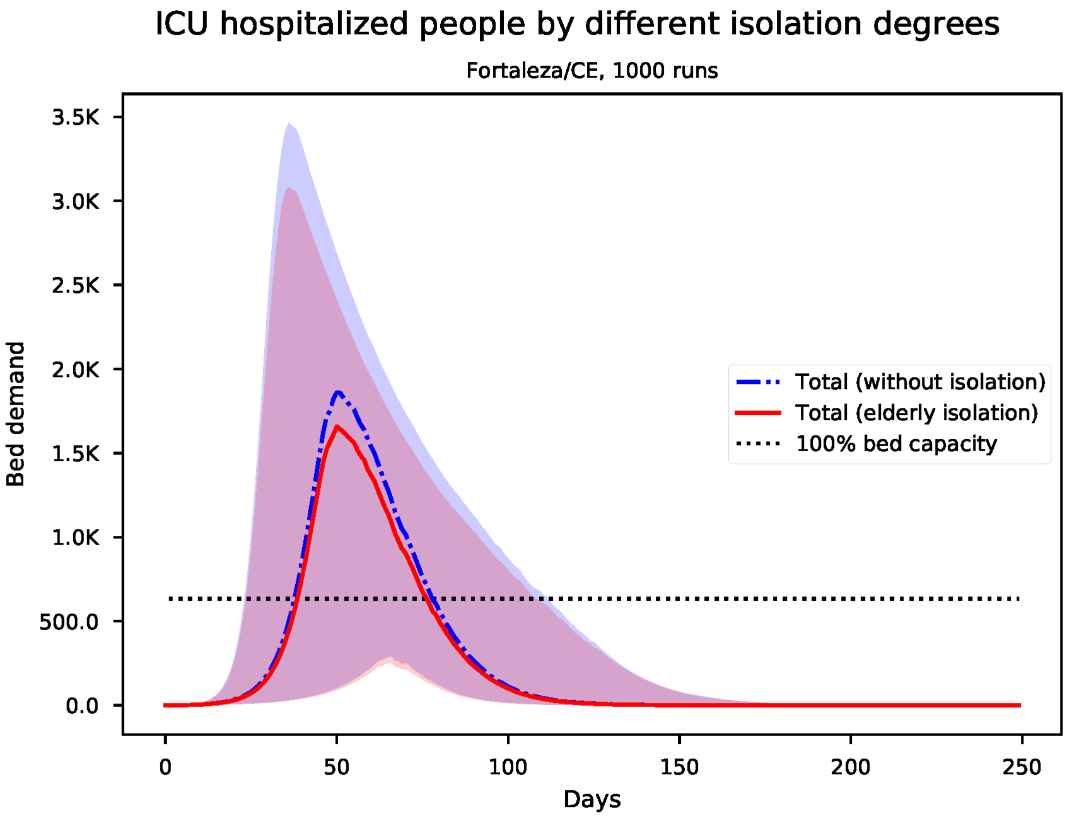


Figure S57: Predicted number of needed ICU beds for Fortaleza

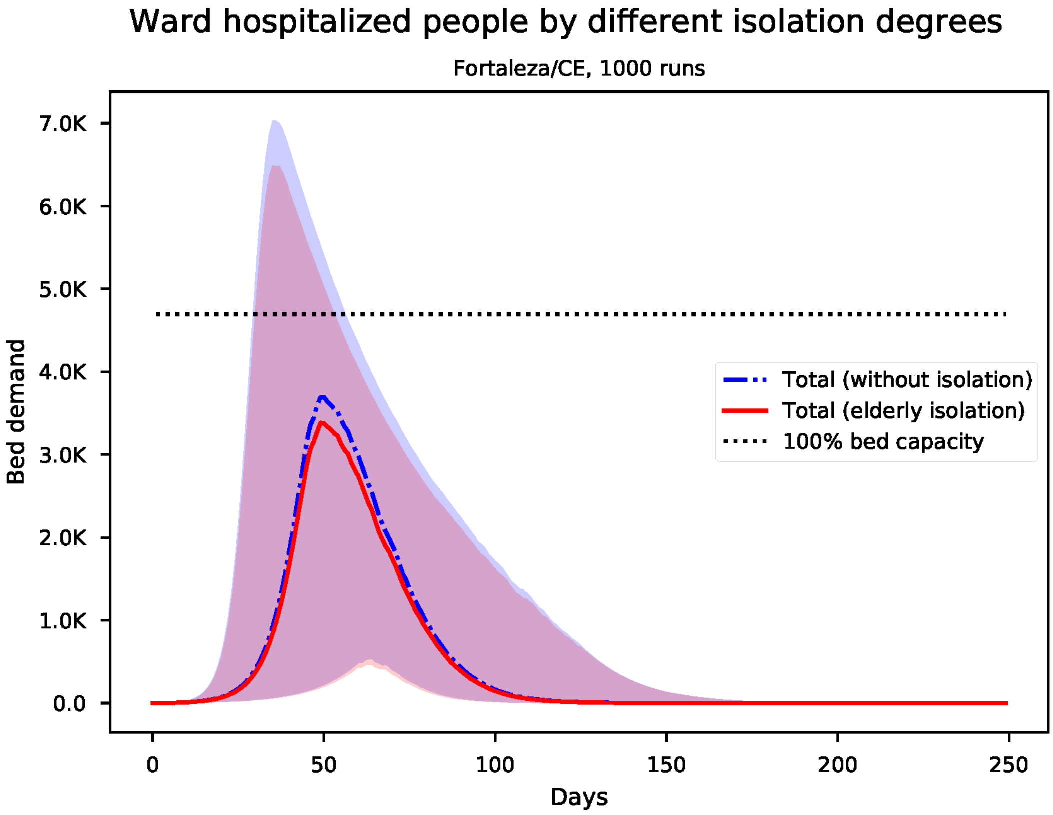


Figure S58: Predicted number of needed ward beds for Fortaleza

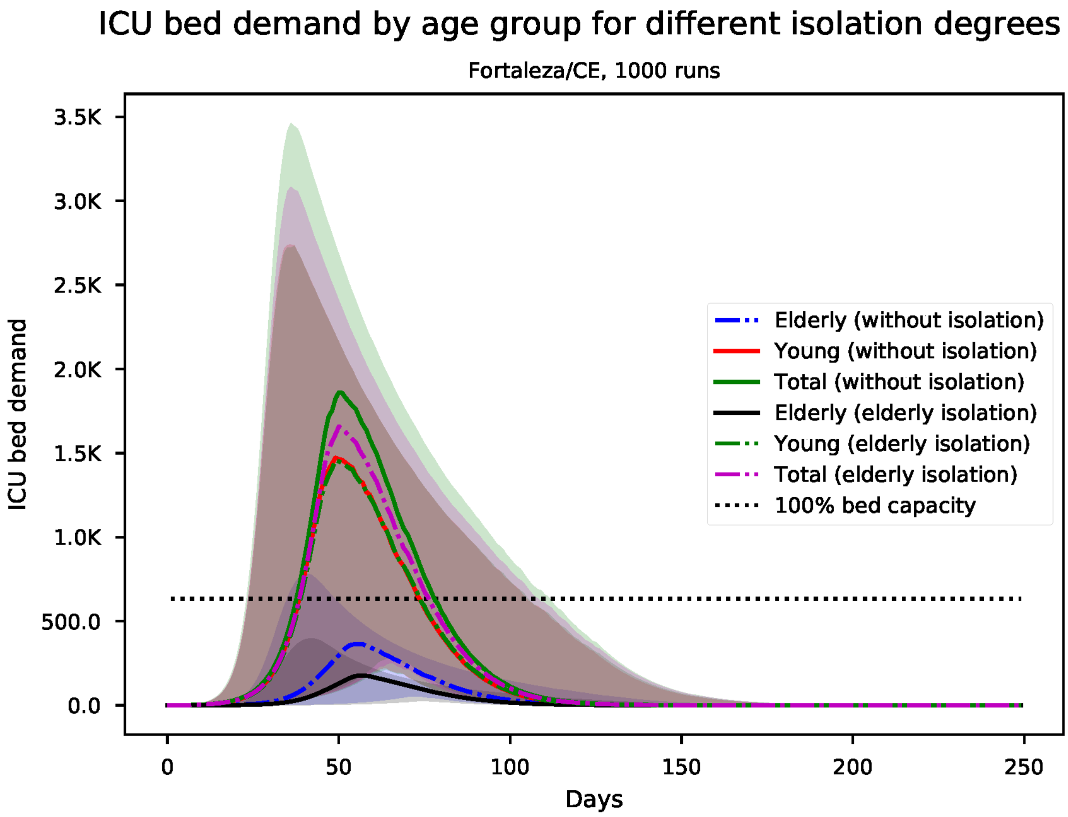


Figure S59: Predicted number of needed ICU beds, by age group for Fortaleza

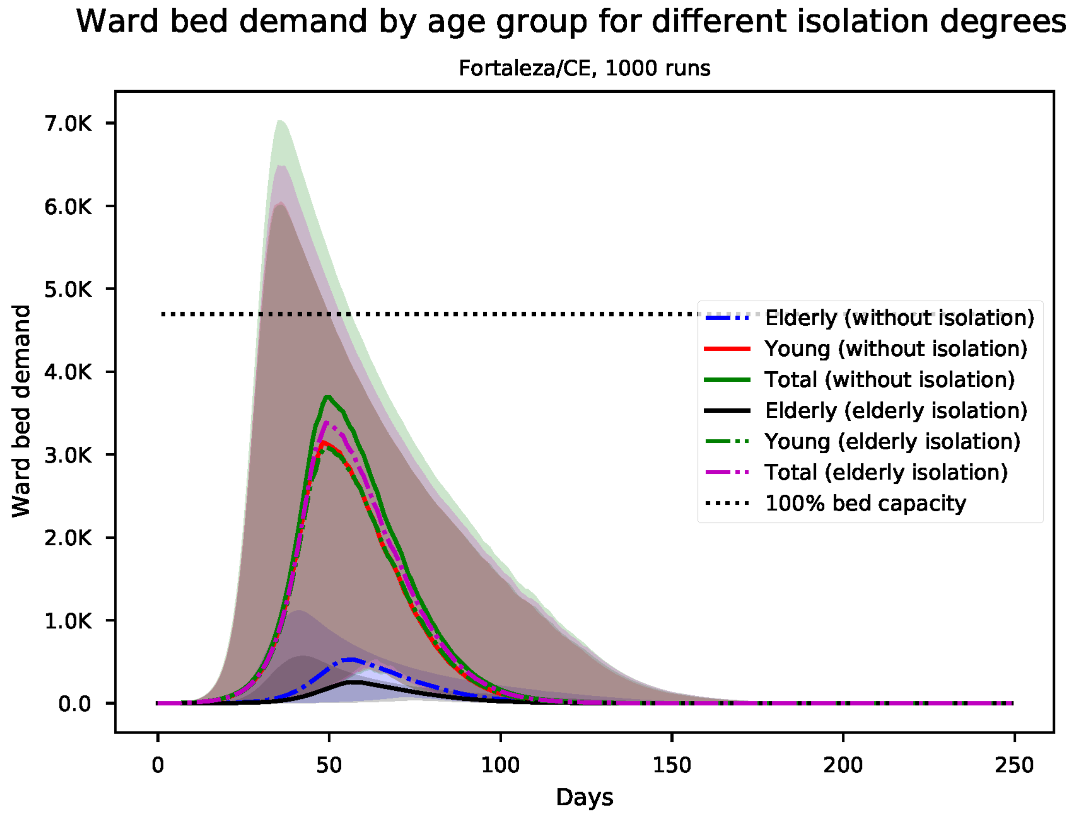


Figure S60: Predicted number of needed ward beds, by age group for Fortaleza

*S11. Results for Goiânia*

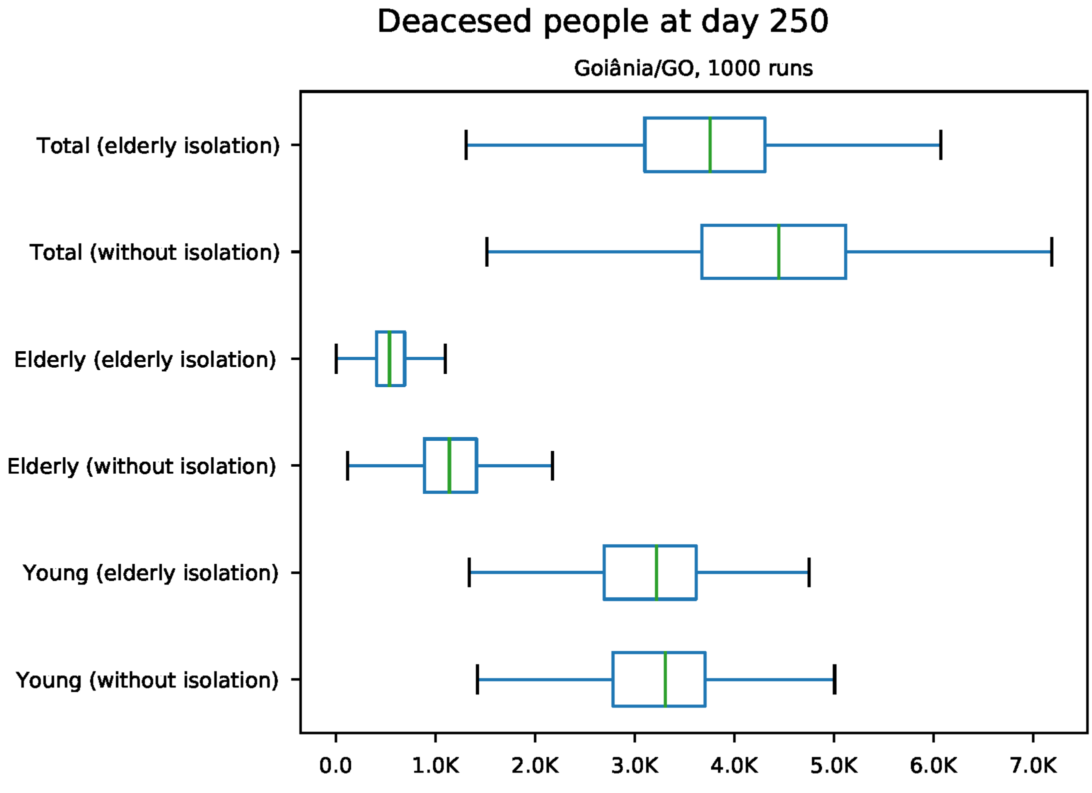


Figure S61: Predicted number of death at day 250 for Goiânia

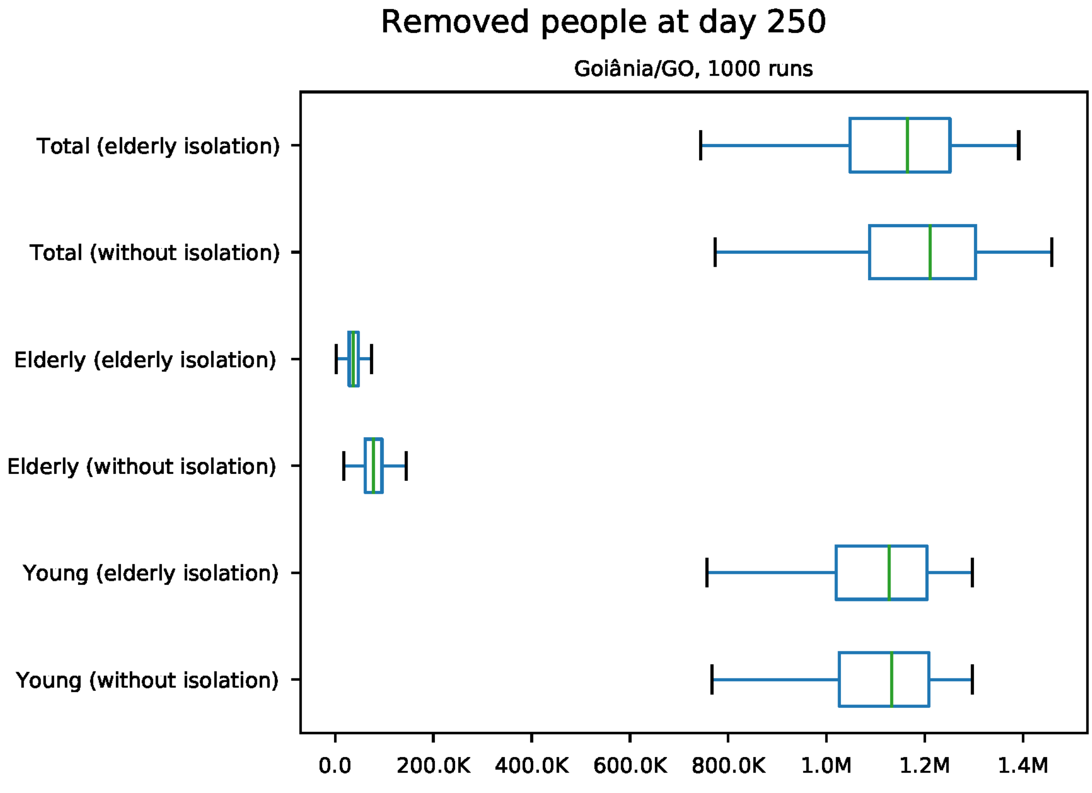


Figure S62: Predicted number of removed individuals at day 250 for Goiânia

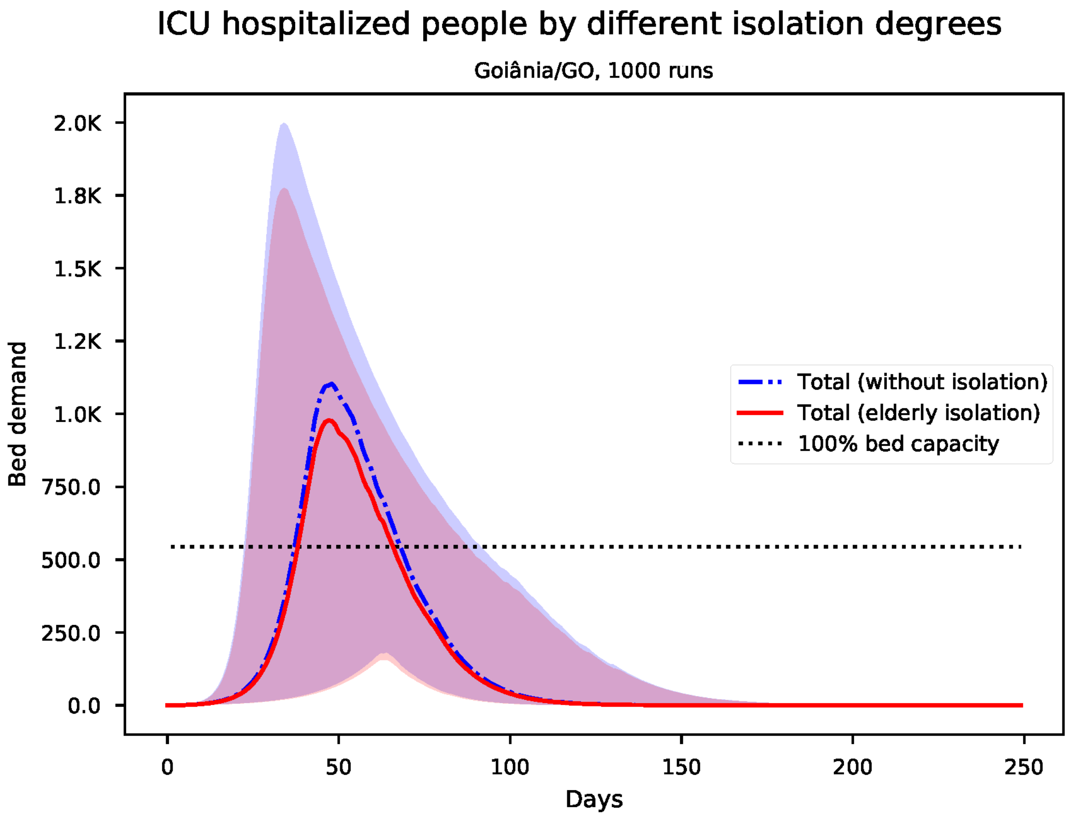


Figure S63: Predicted number of needed ICU beds for Goiânia

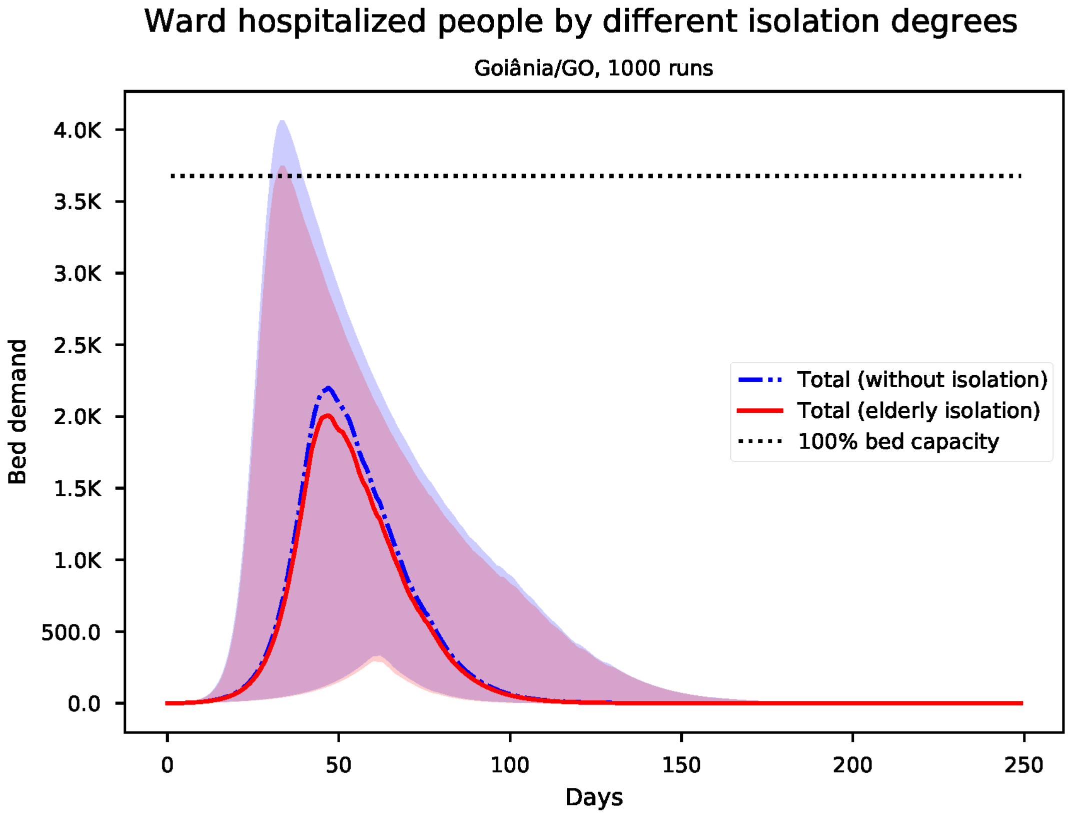


Figure S64: Predicted number of needed ward beds for Goiânia

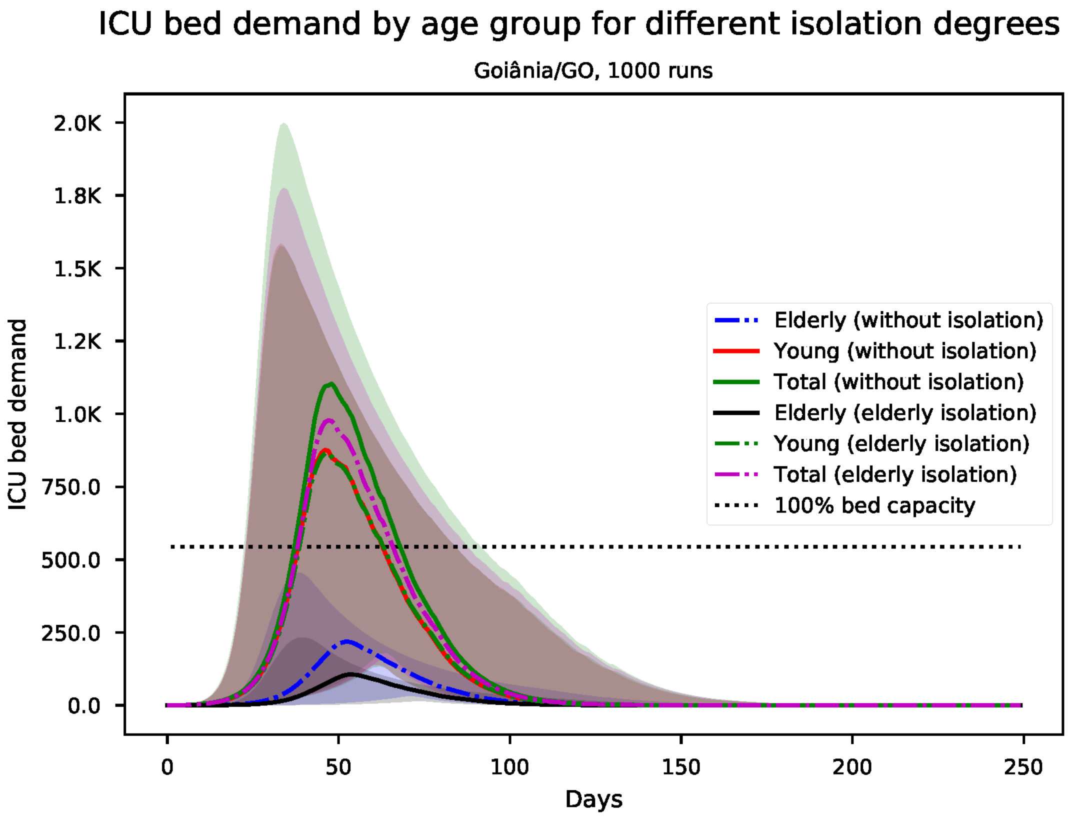


Figure S65: Predicted number of needed ICU beds, by age group for Goiânia

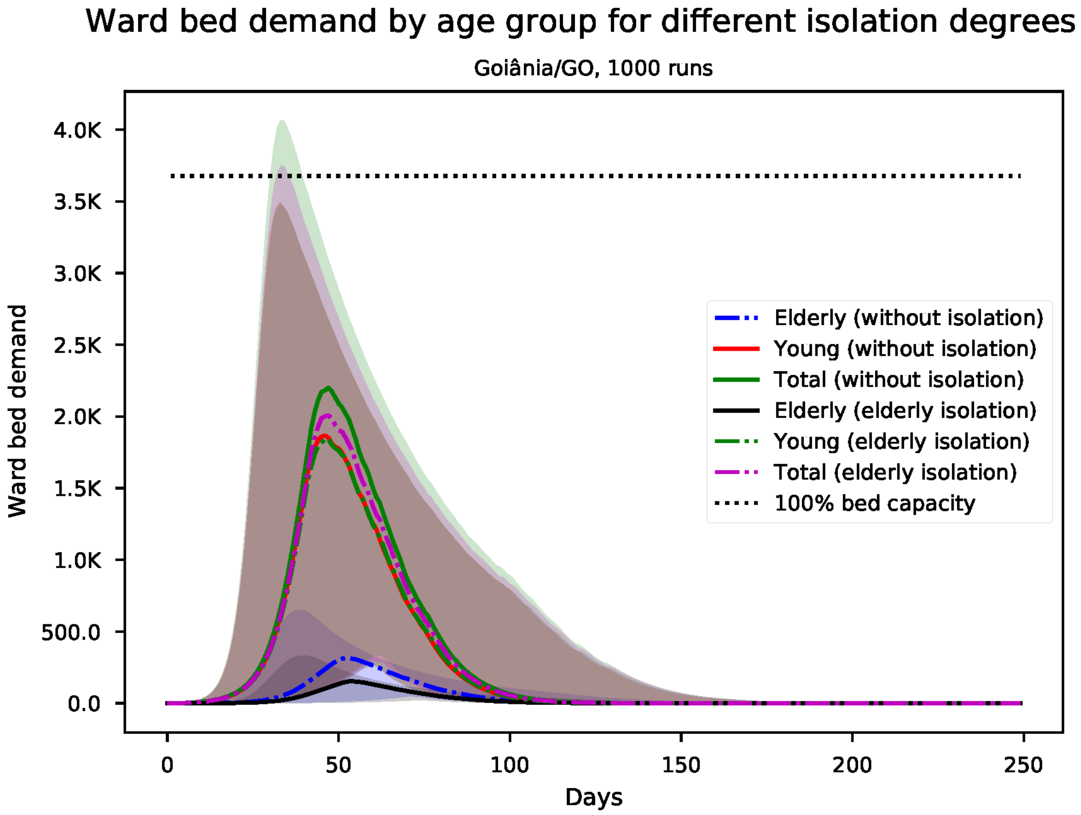


Figure S66: Predicted number of needed ward beds, by age group for Goiânia

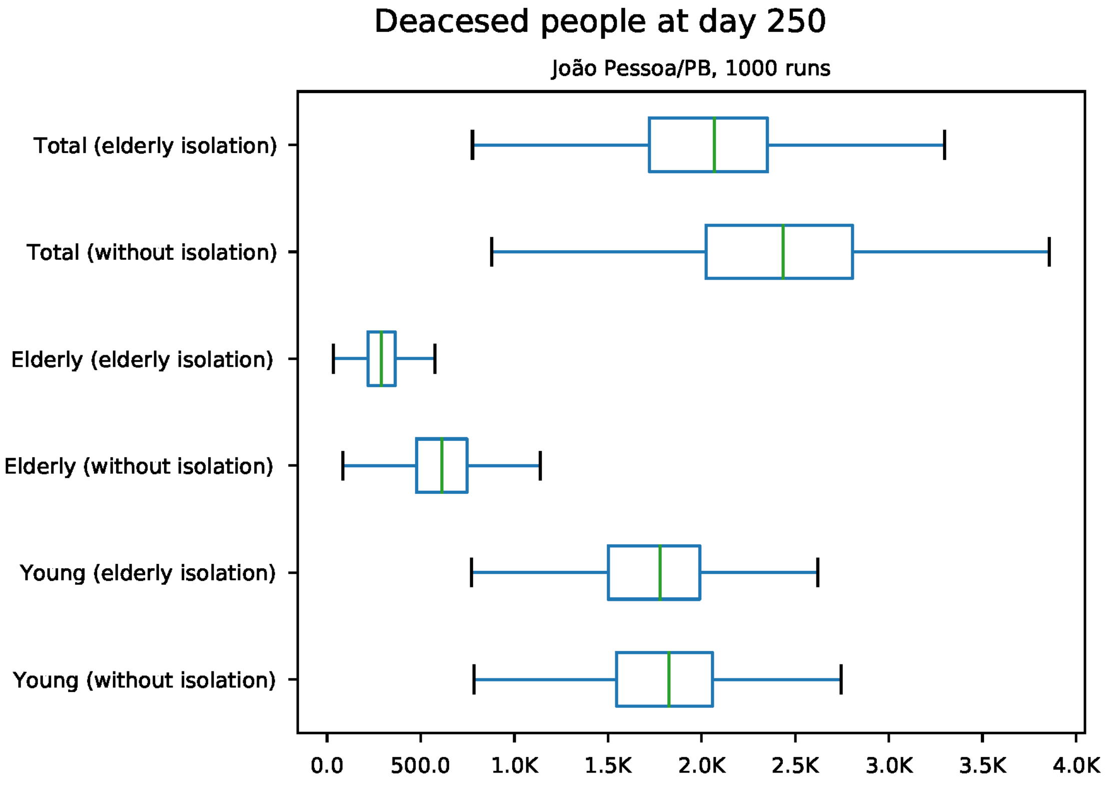


Figure S67: Predicted number of death at day 250 for João Pessoa

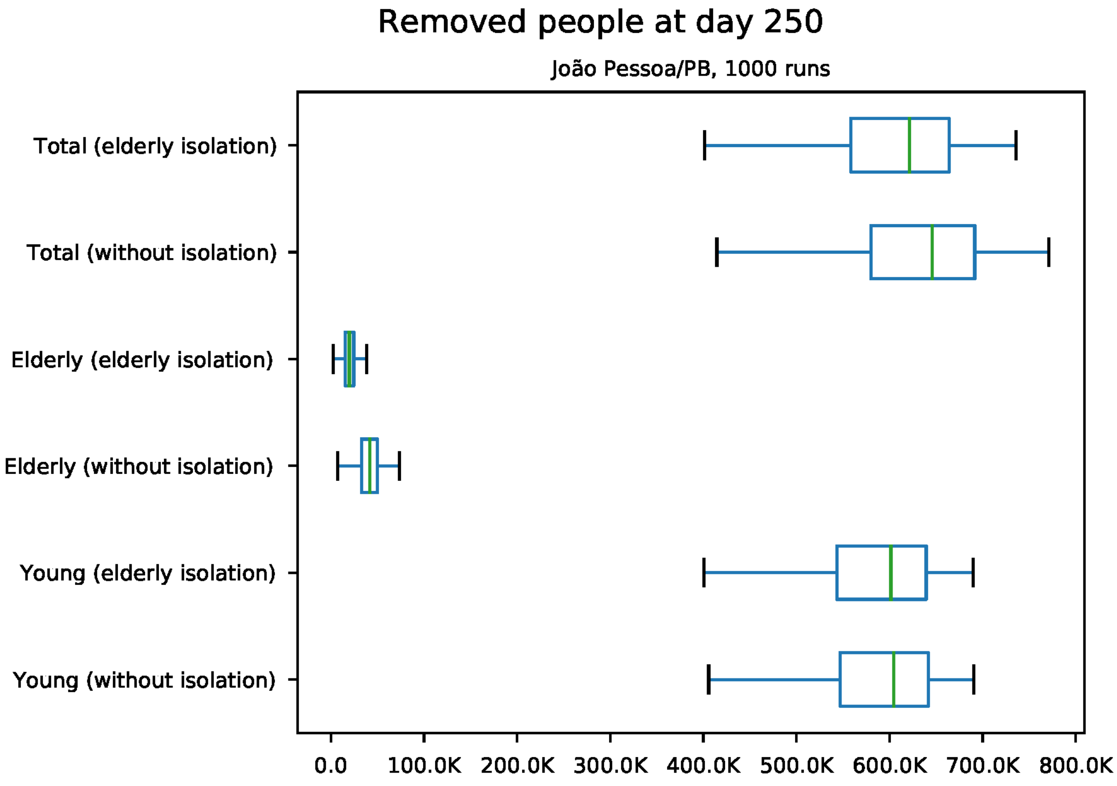


Figure S68: Predicted number of removed individuals at day 250 for João Pessoa

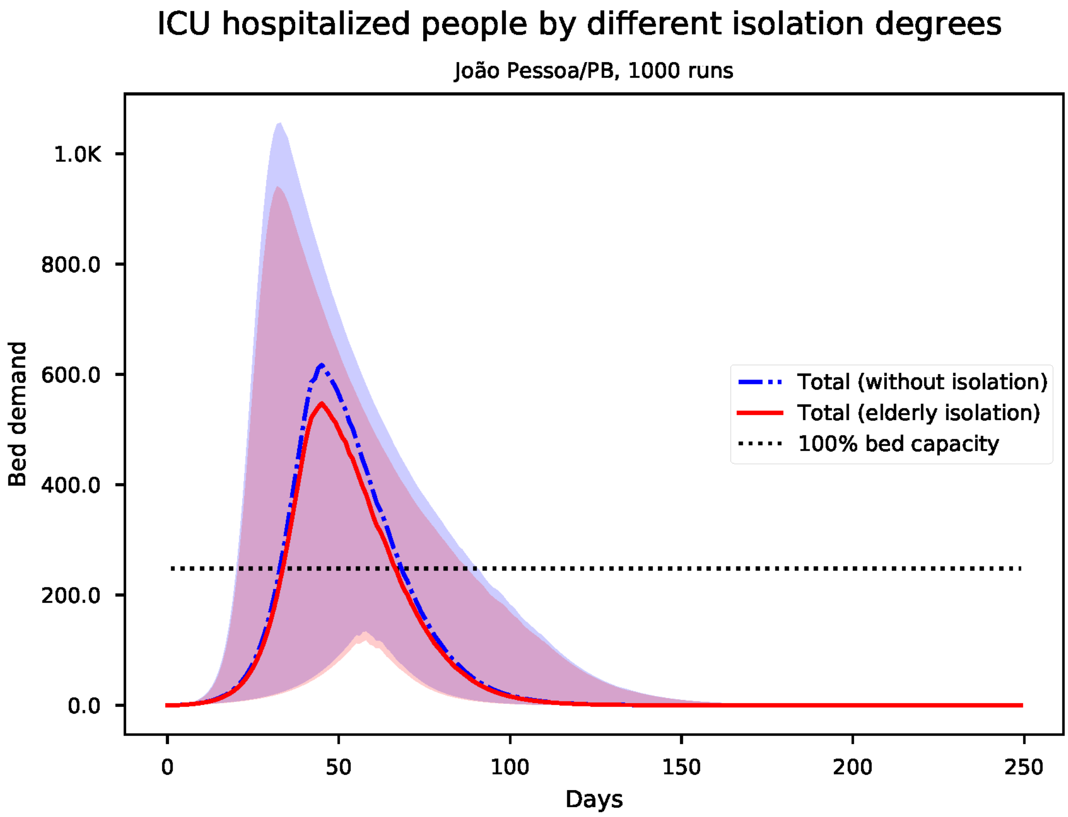


Figure S69: Predicted number of needed ICU beds for João Pessoa

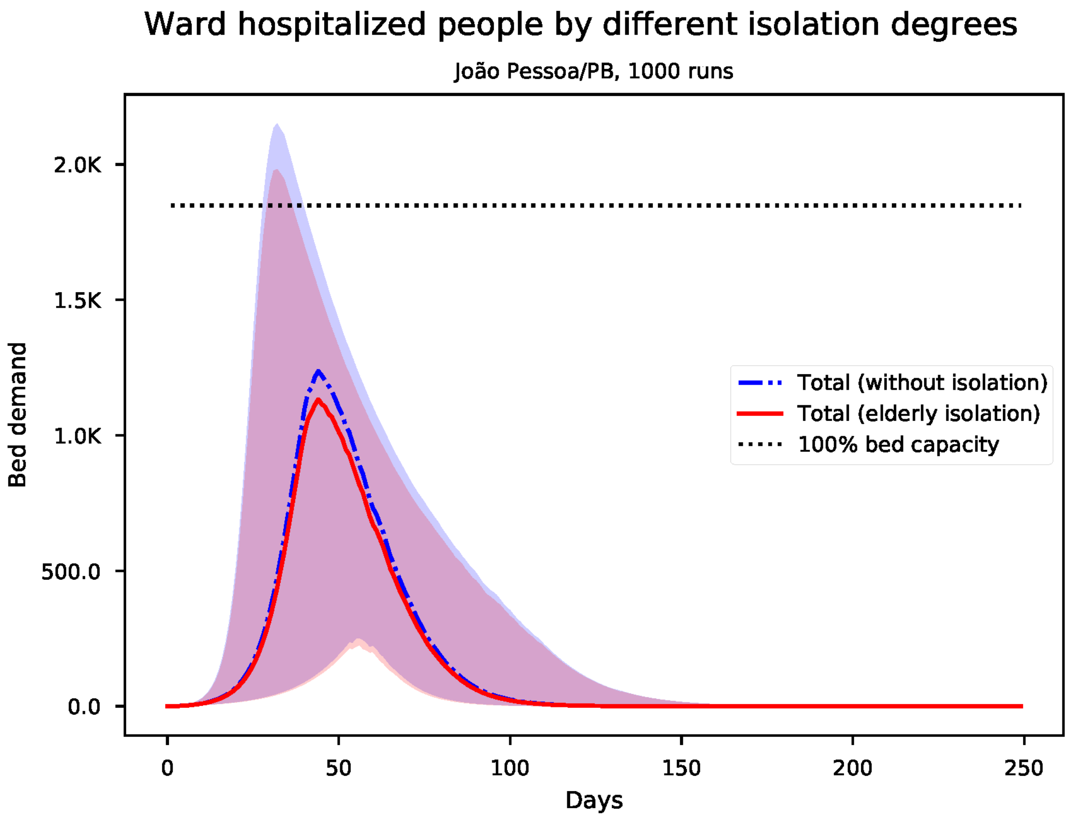


Figure S70: Predicted number of needed ward beds for João Pessoa

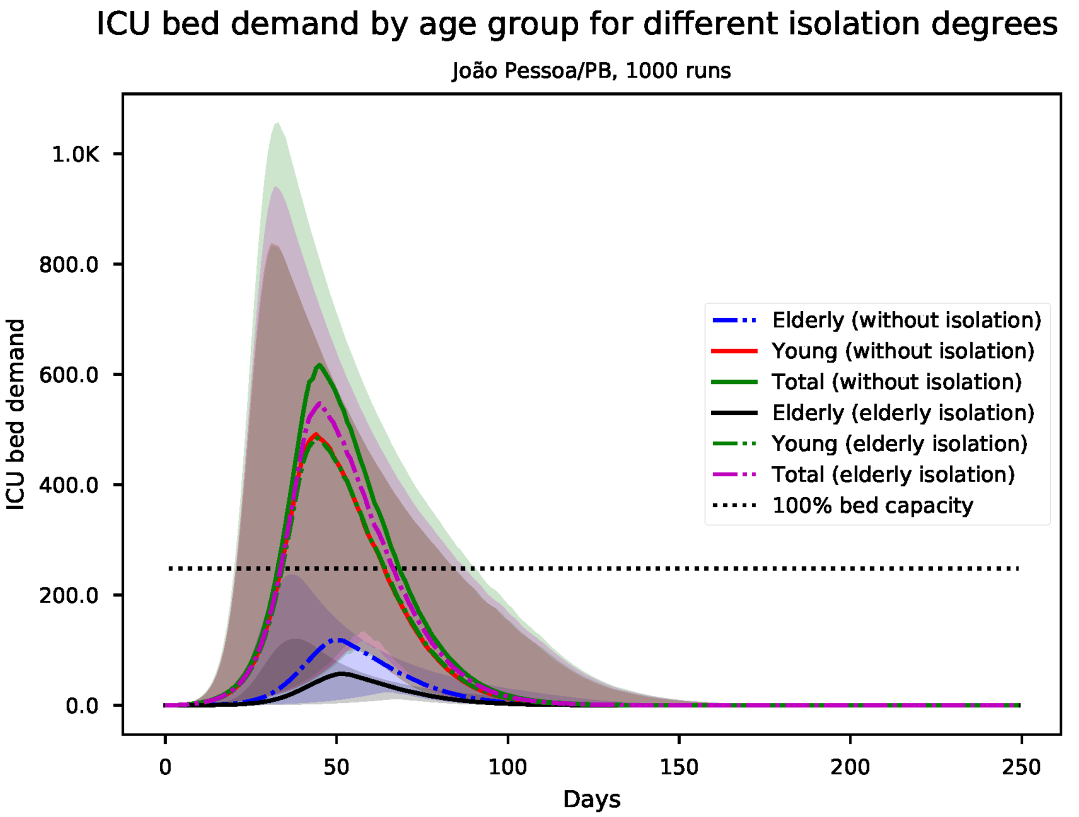


Figure S71: Predicted number of needed ICU beds, by age group for João Pessoa

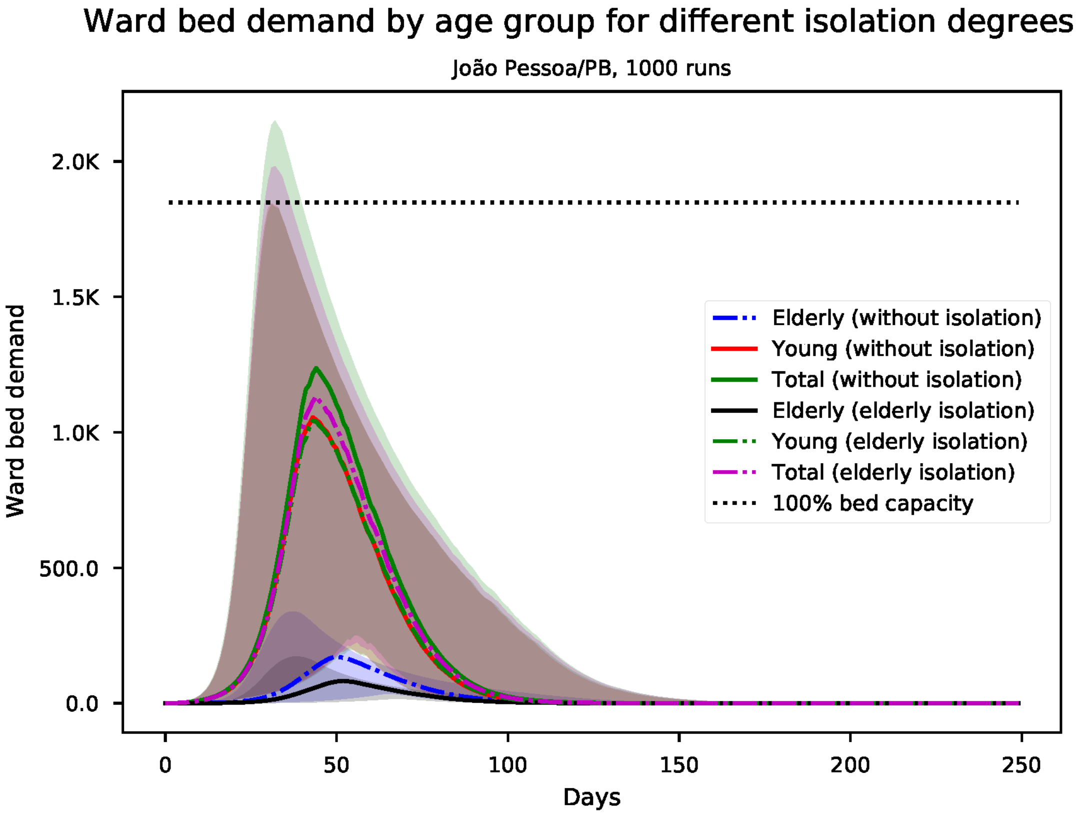


Figure S72: Predicted number of needed ward beds, by age group for João Pessoa

*S13. Results for Macapá*

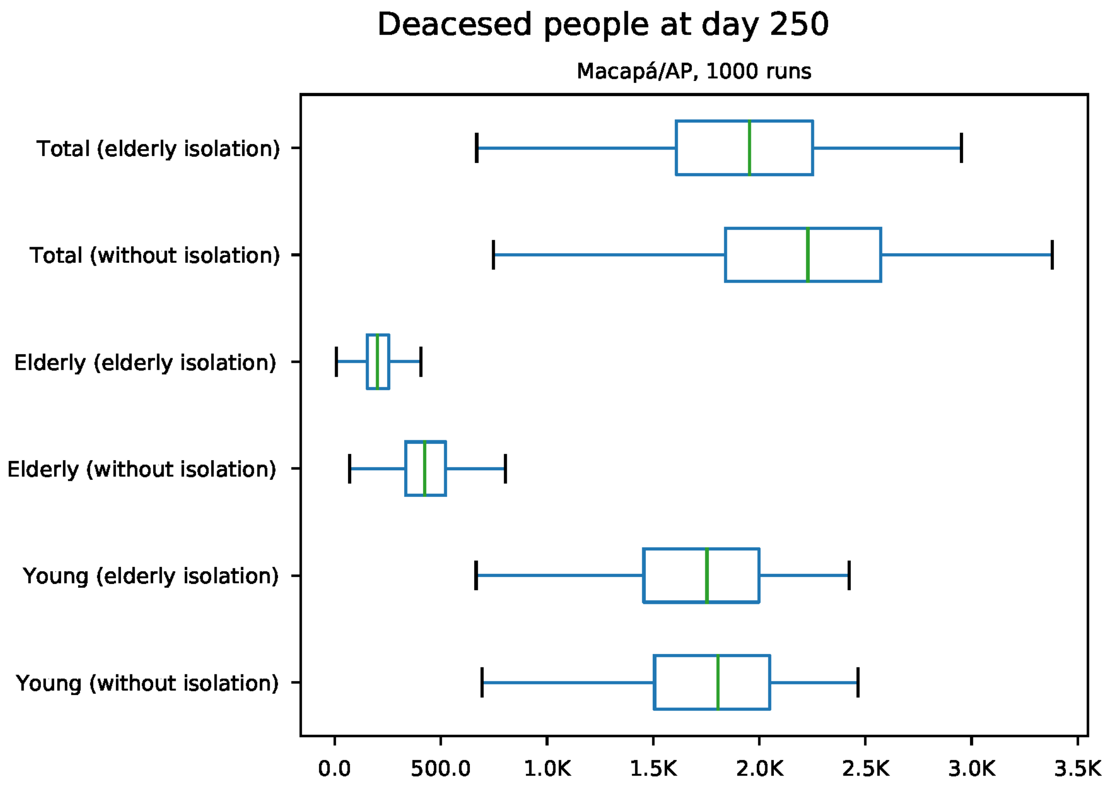


Figure S73: Predicted number of death at day 250 for Macapá

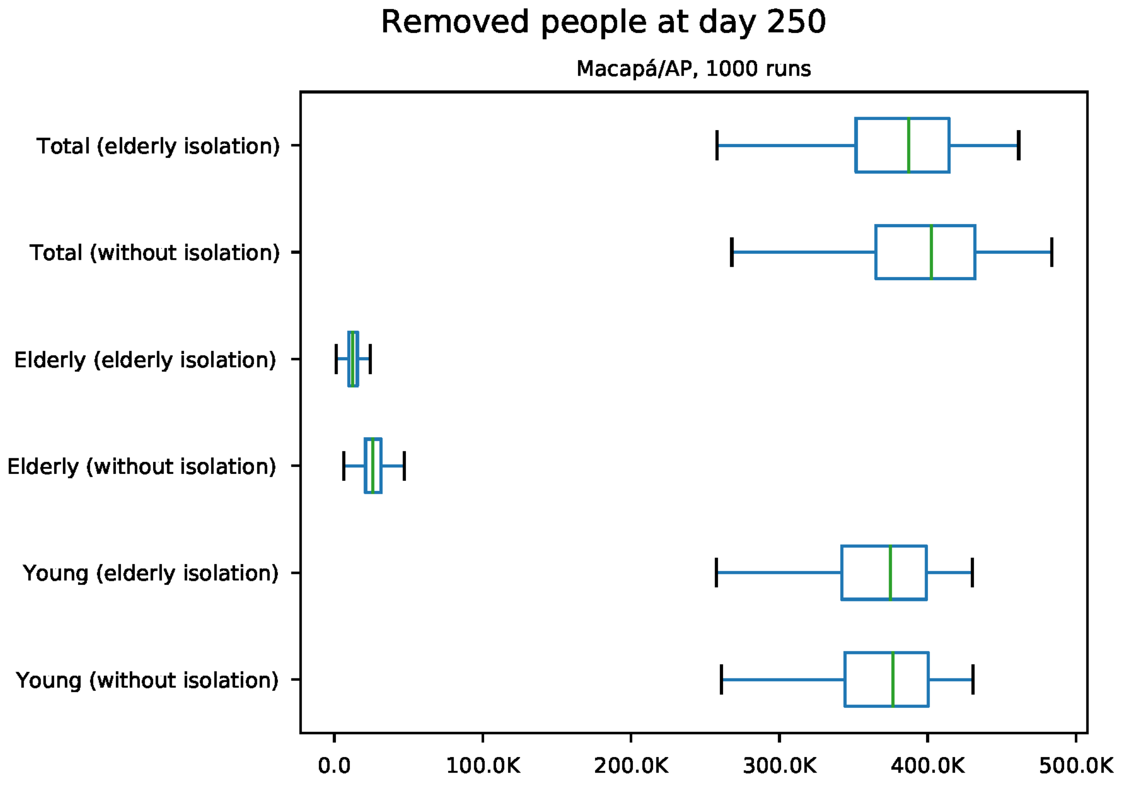


Figure S74: Predicted number of removed individuals at day 250 for Macapá

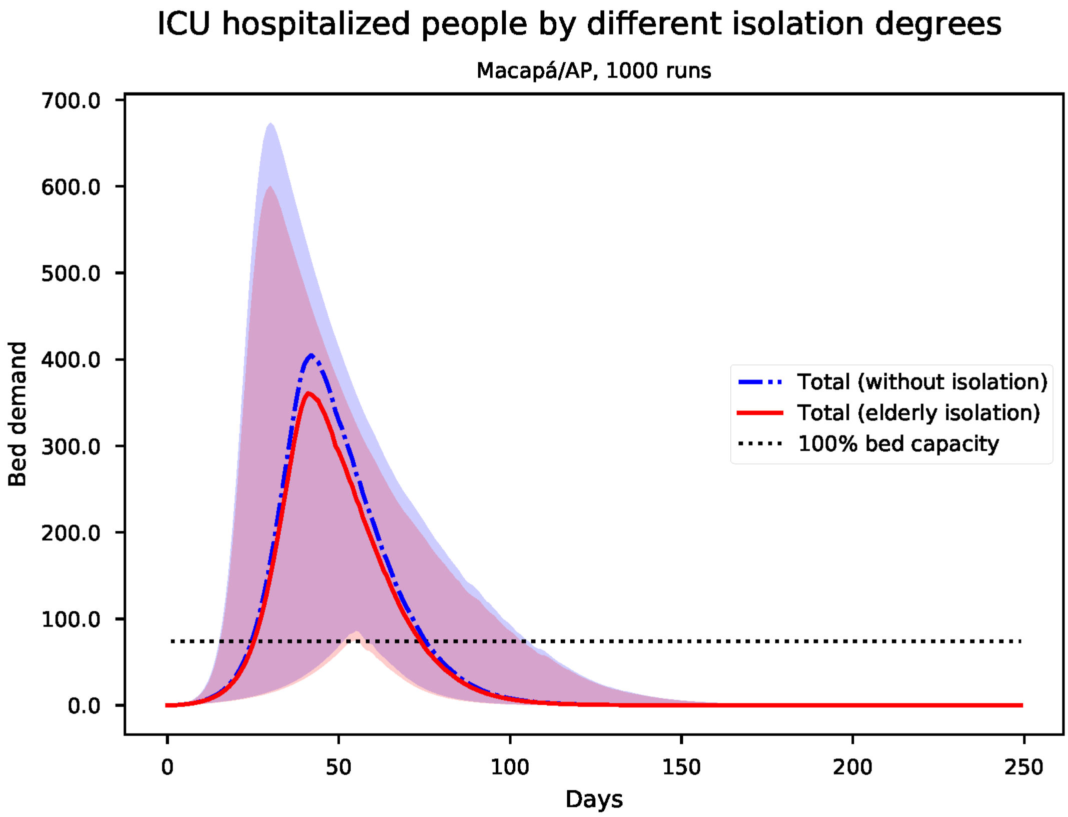


Figure S75: Predicted number of needed ICU beds for Macapá

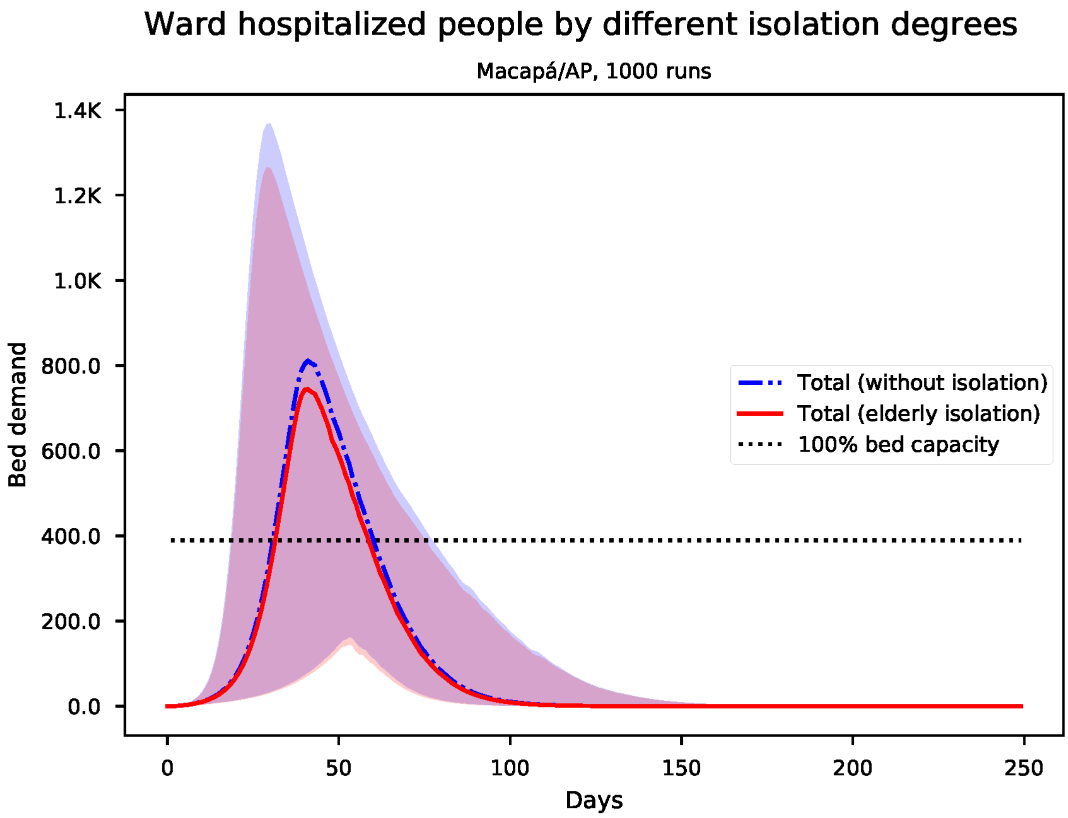


Figure S76: Predicted number of needed ward beds for Macapá

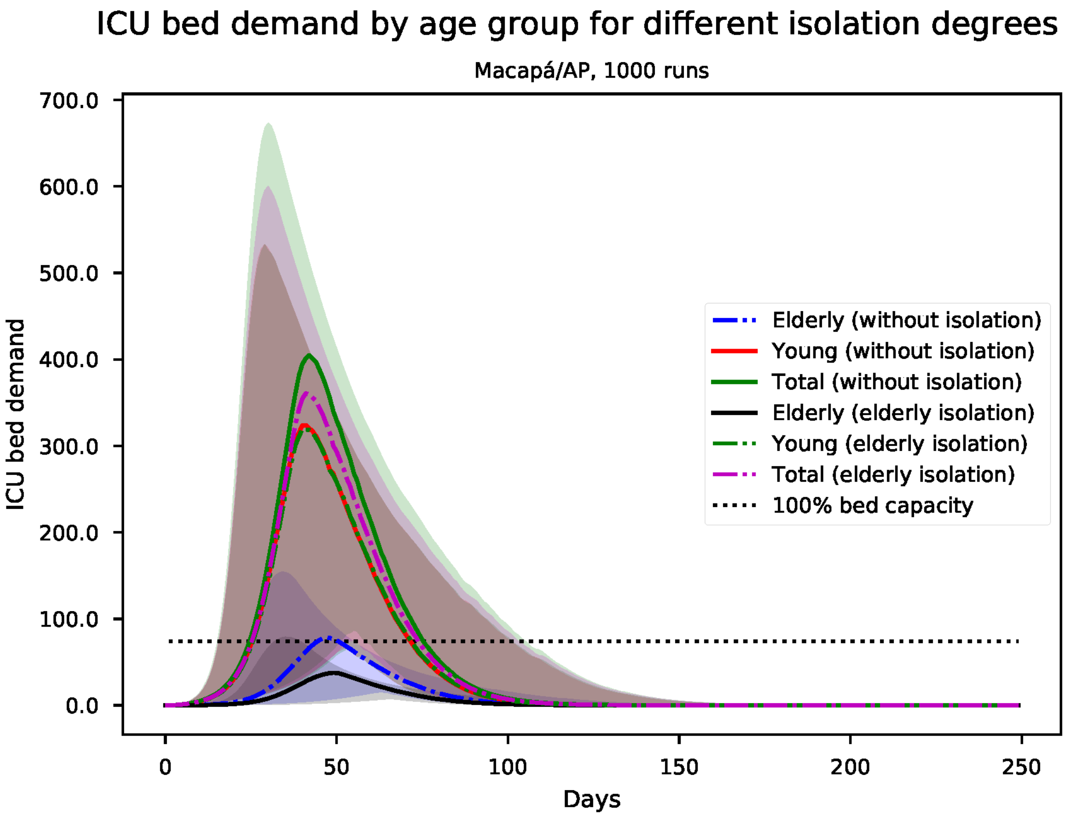


Figure S77: Predicted number of needed ICU beds, by age group for Macapá

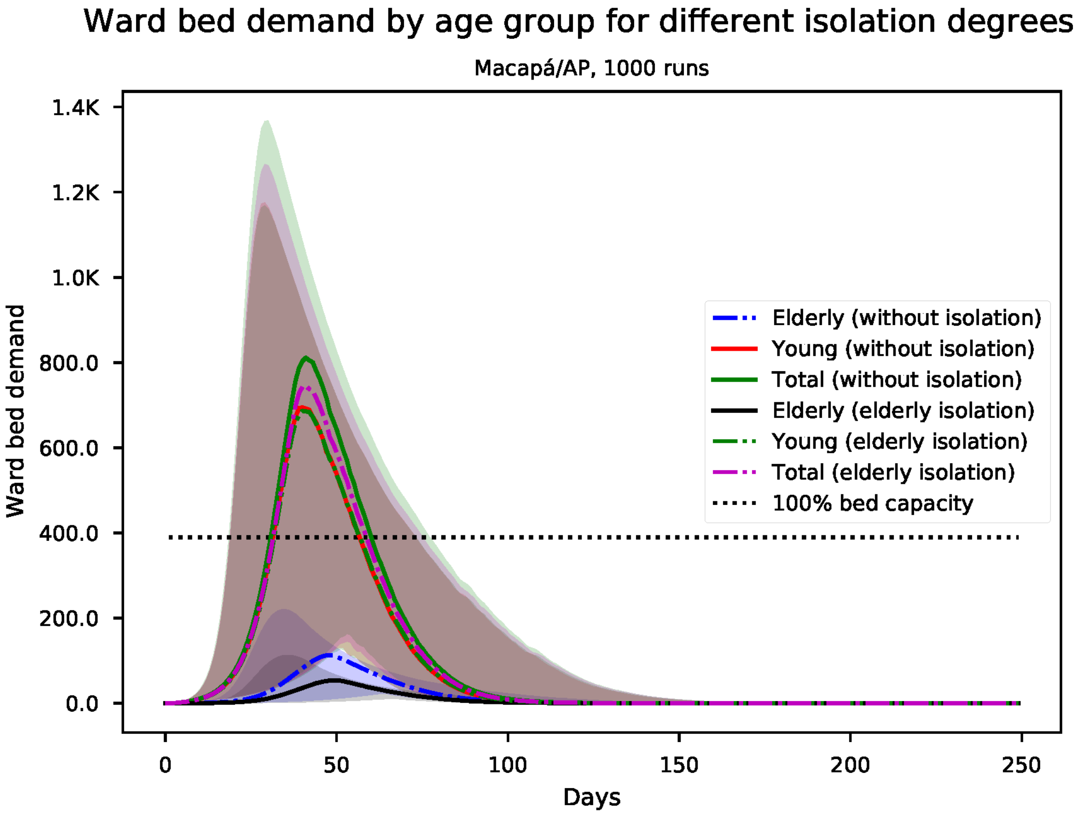


Figure S78: Predicted number of needed ward beds, by age group for Macapá

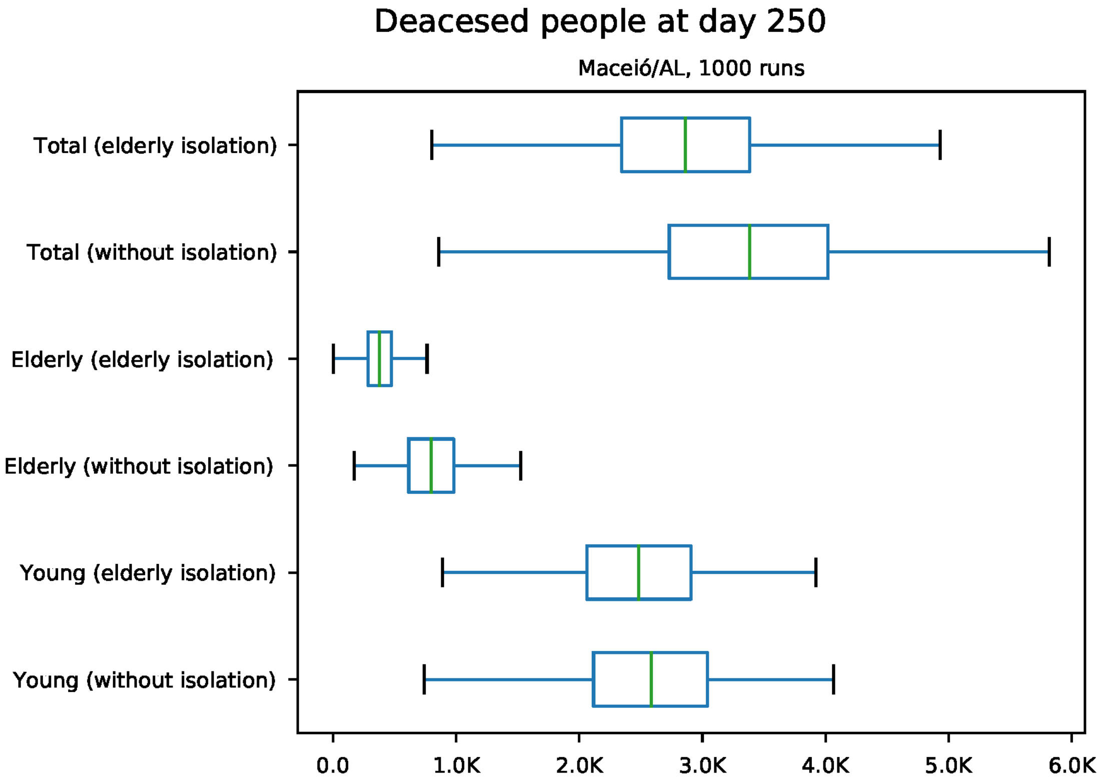


Figure S79: Predicted number of death at day 250 for Maceió

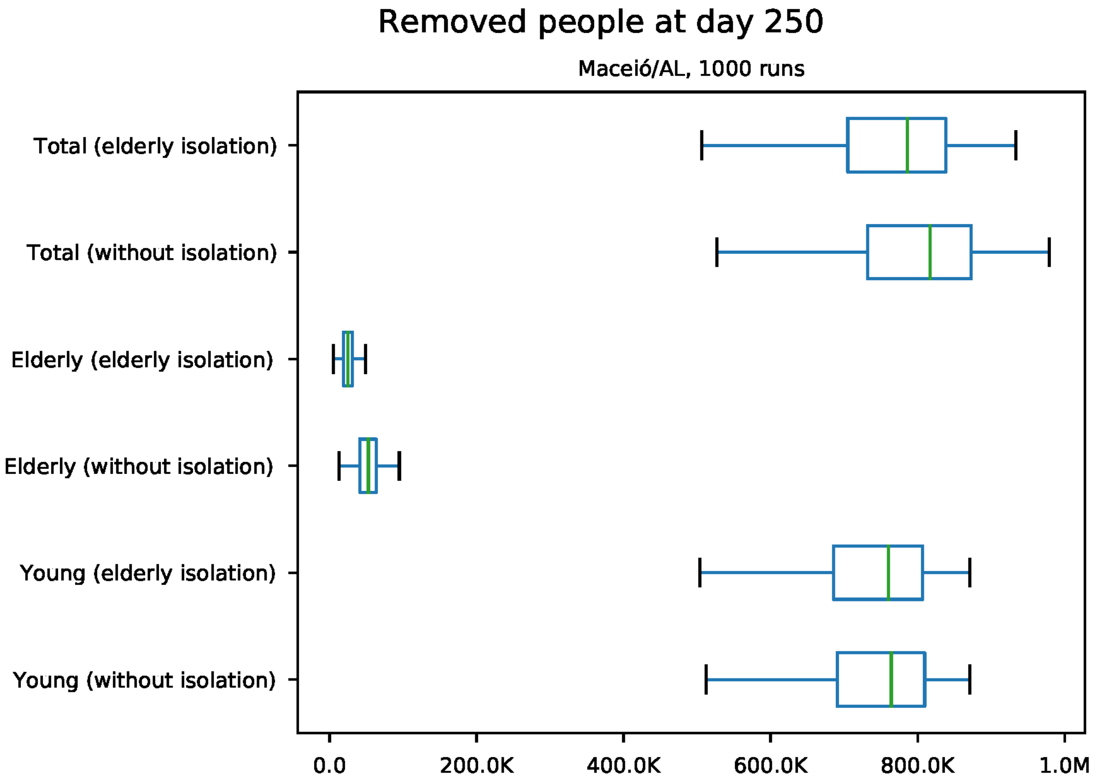


Figure S80: Predicted number of removed individuals at day 250 for Maceió

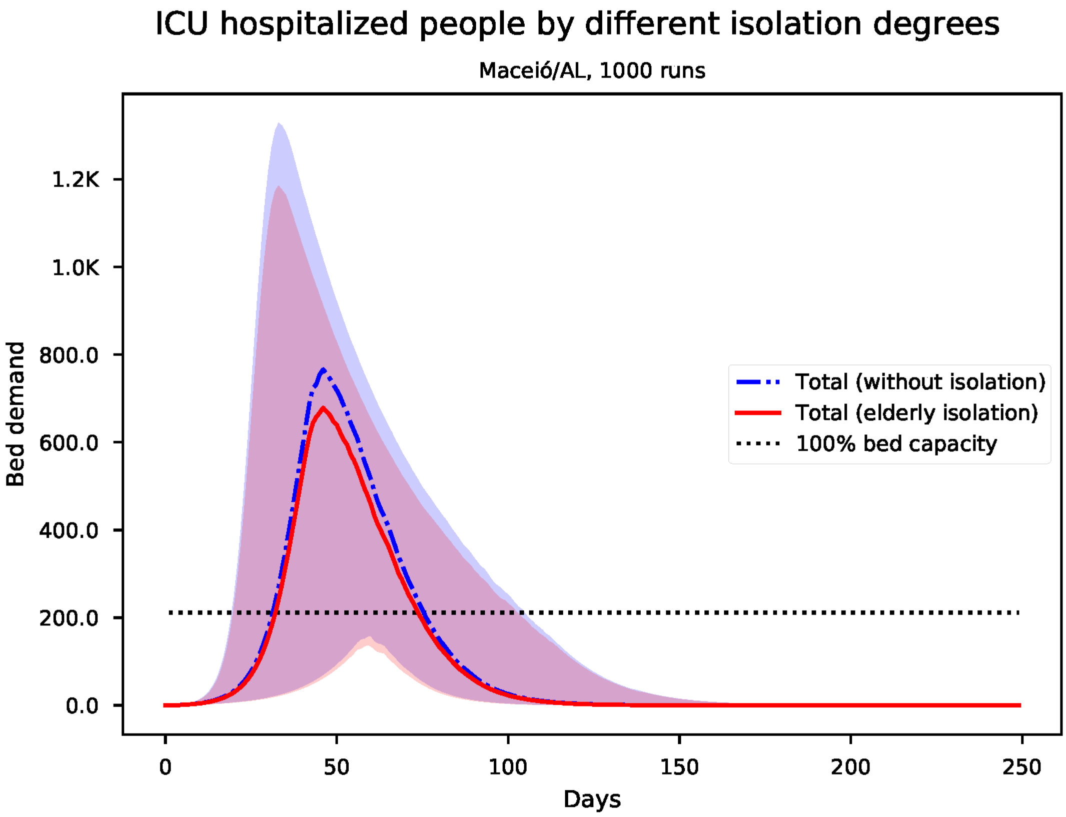


Figure S81: Predicted number of needed ICU beds for Maceió

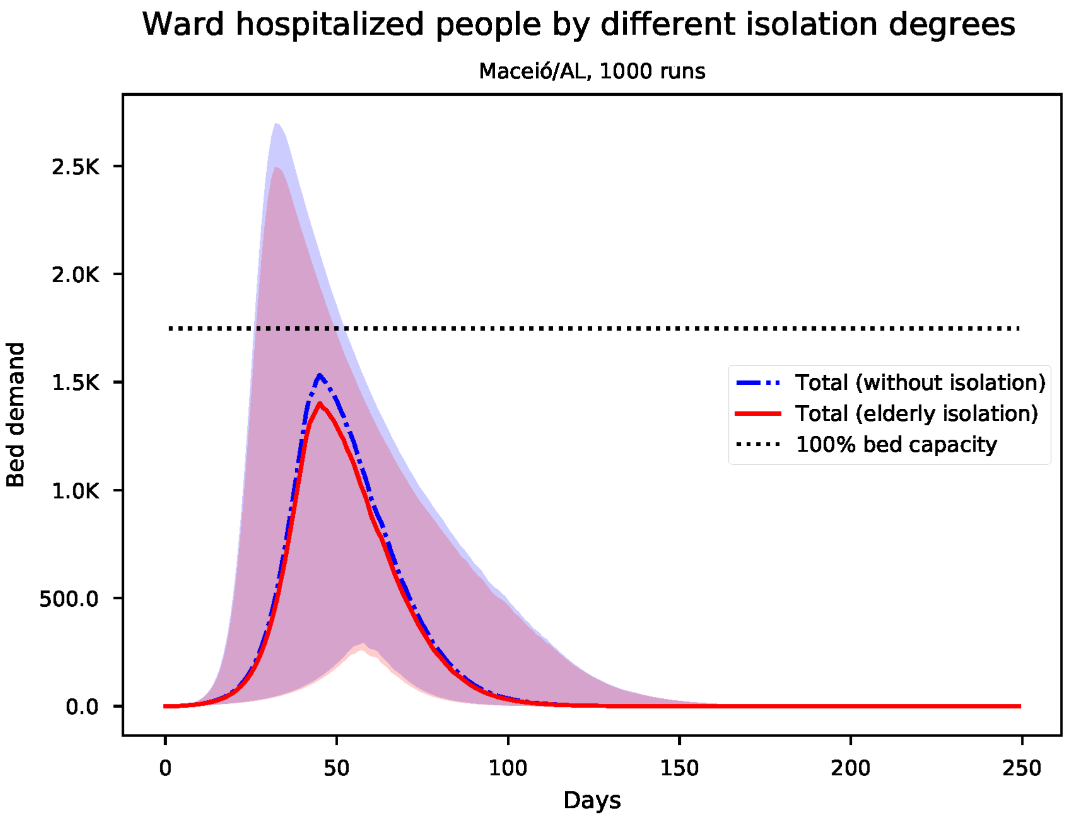


Figure S82: Predicted number of needed ward beds for Maceió

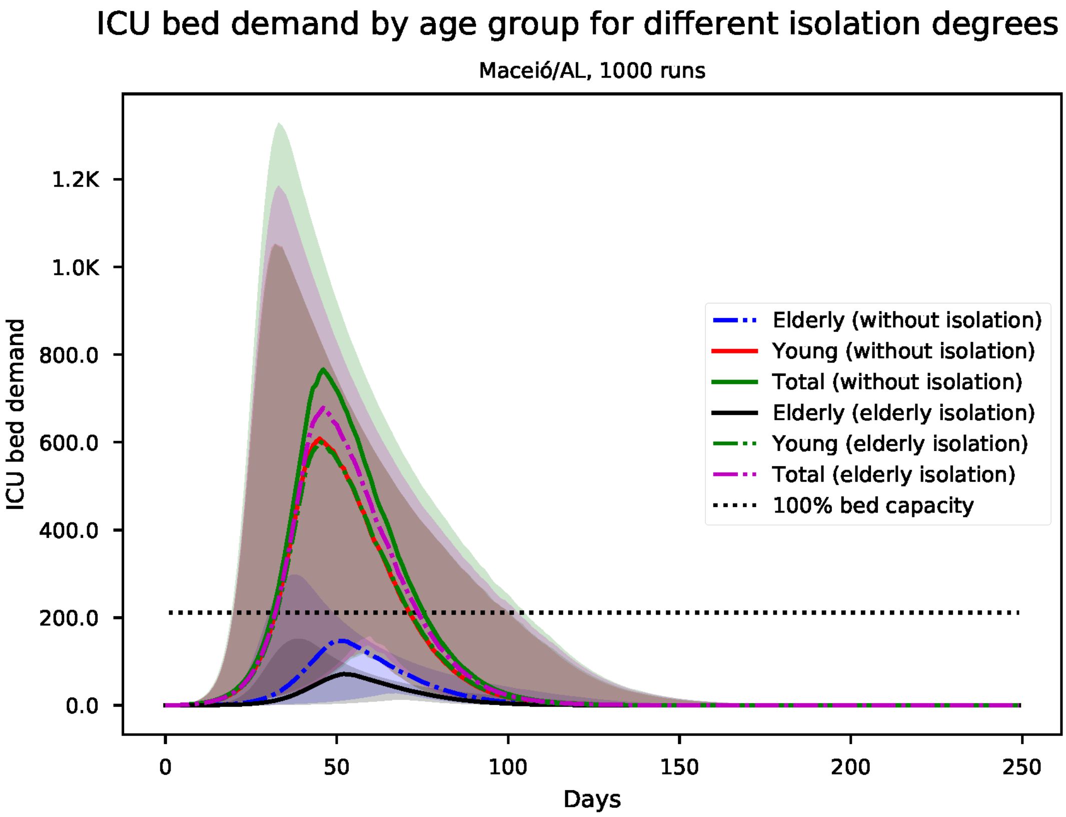


Figure S83: Predicted number of needed ICU beds, by age group for Maceió

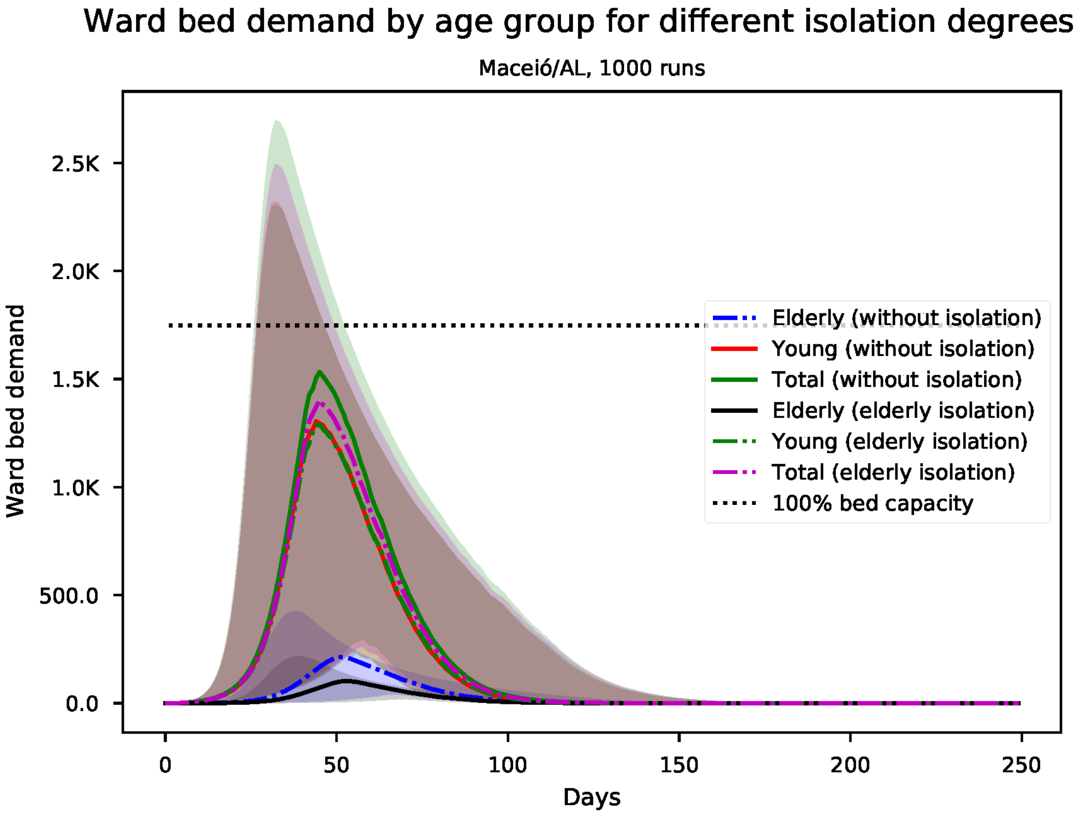


Figure S84: Predicted number of needed ward beds, by age group for Maceió

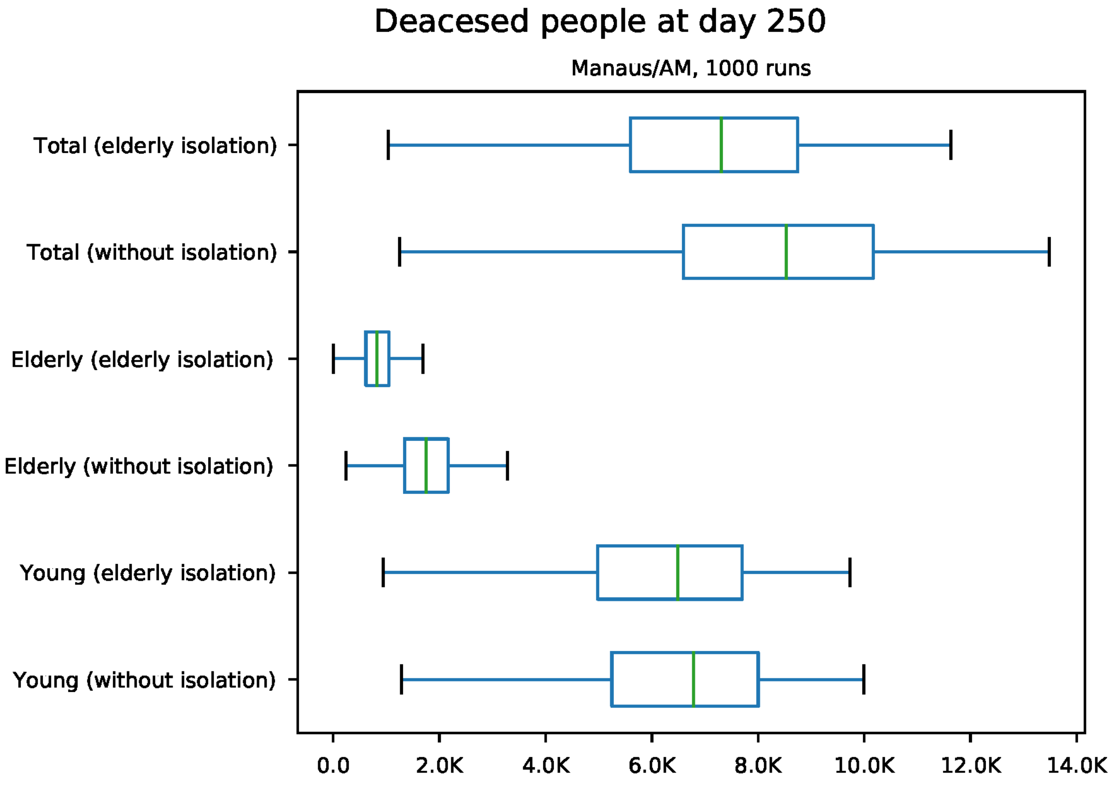


Figure S85: Predicted number of death at day 250 for Manaus

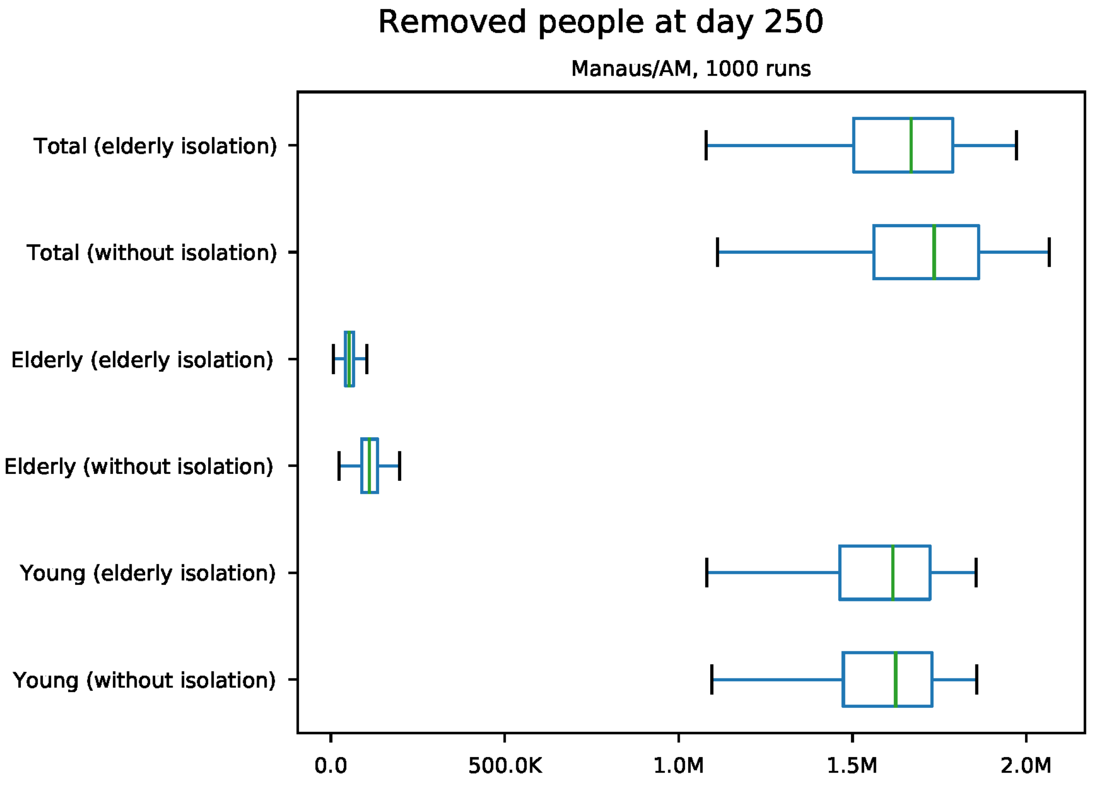


Figure S86: Predicted number of removed individuals at day 250 for Manaus

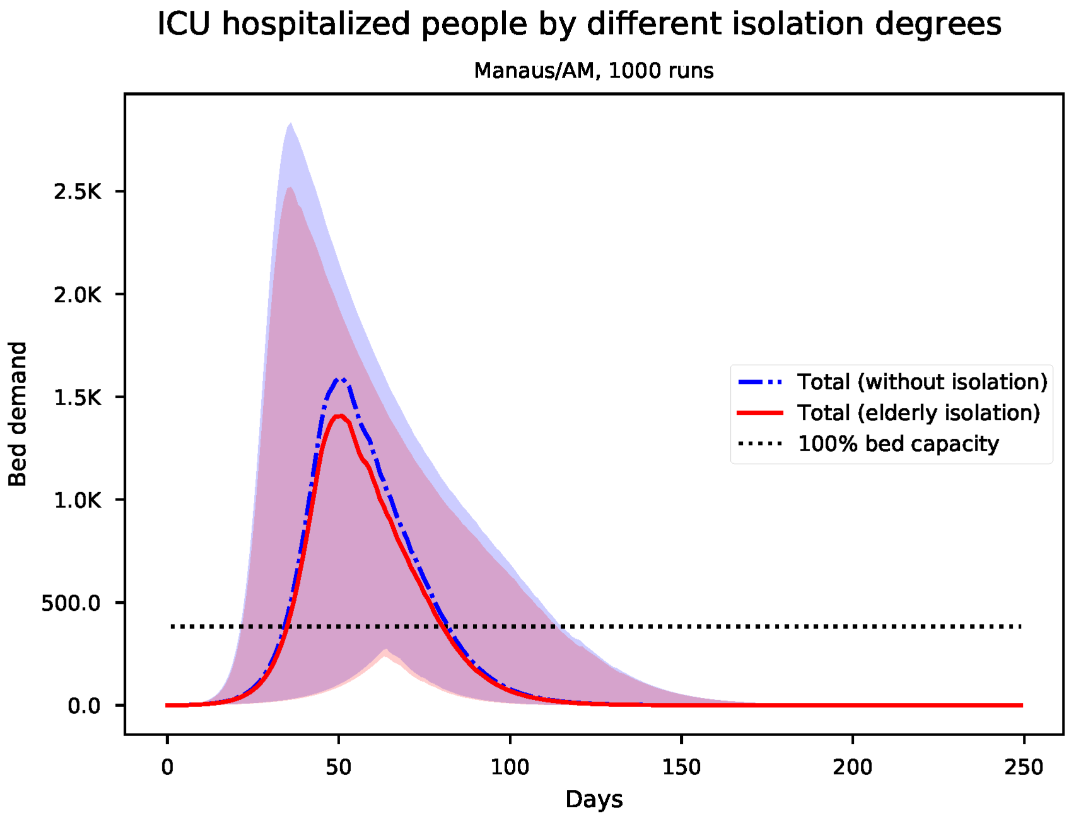


Figure S87: Predicted number of needed ICU beds for Manaus

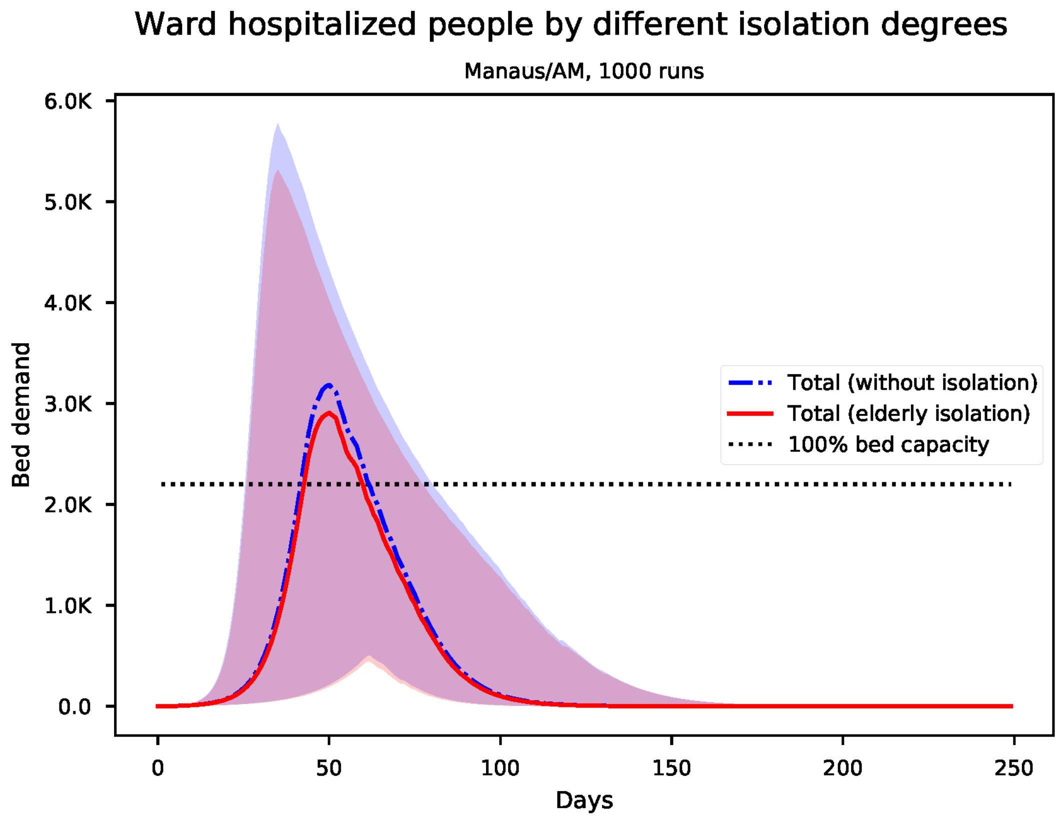


Figure S88: Predicted number of needed ward beds for Manaus

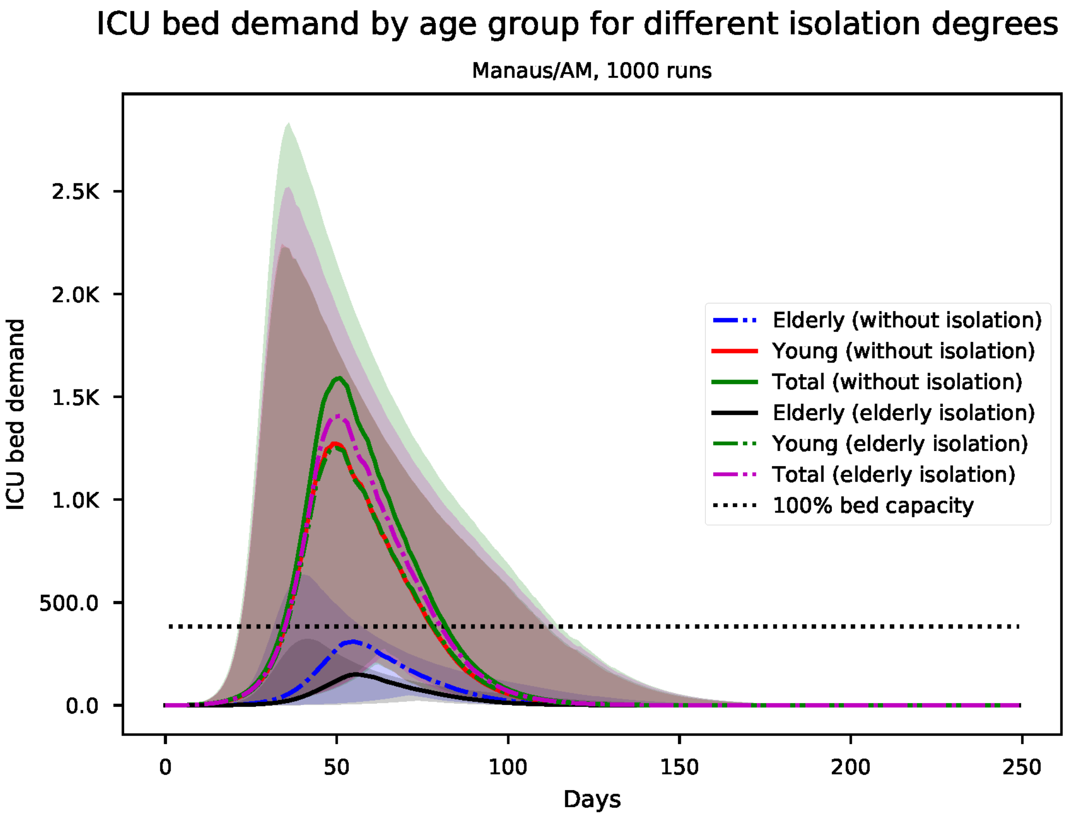


Figure S89: Predicted number of needed ICU beds, by age group for Manaus

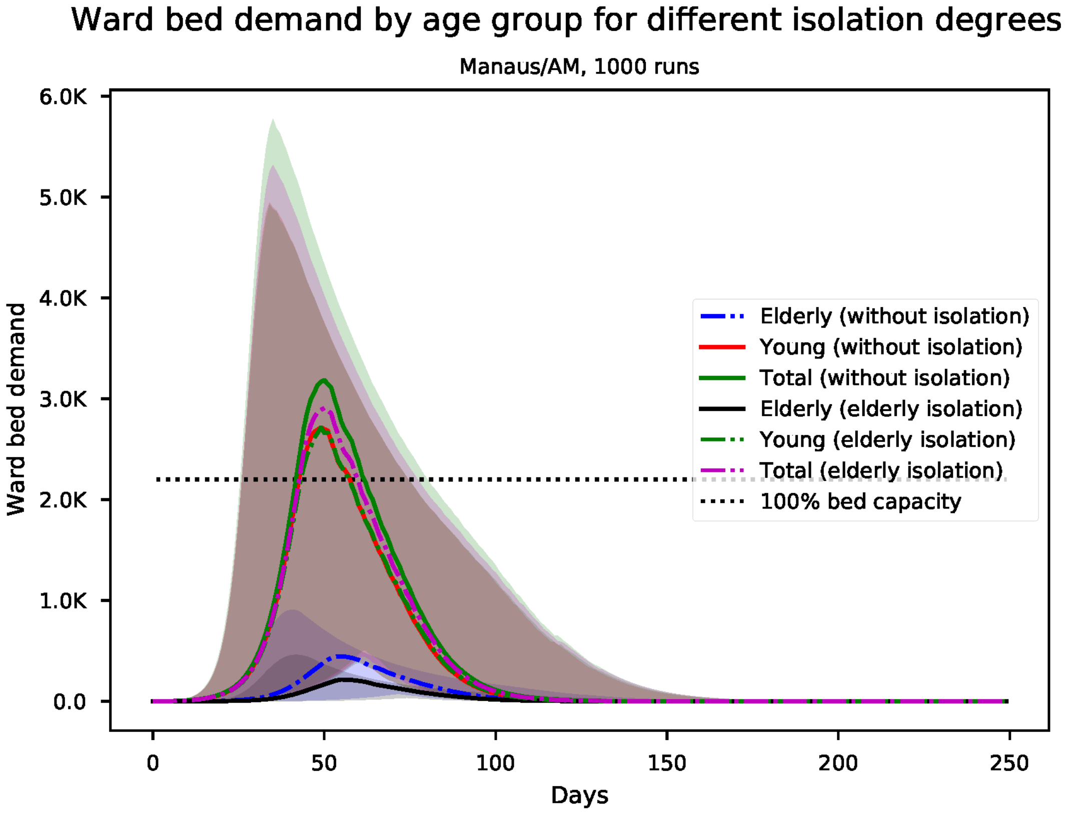


Figure S90: Predicted number of needed ward beds, by age group for Manaus

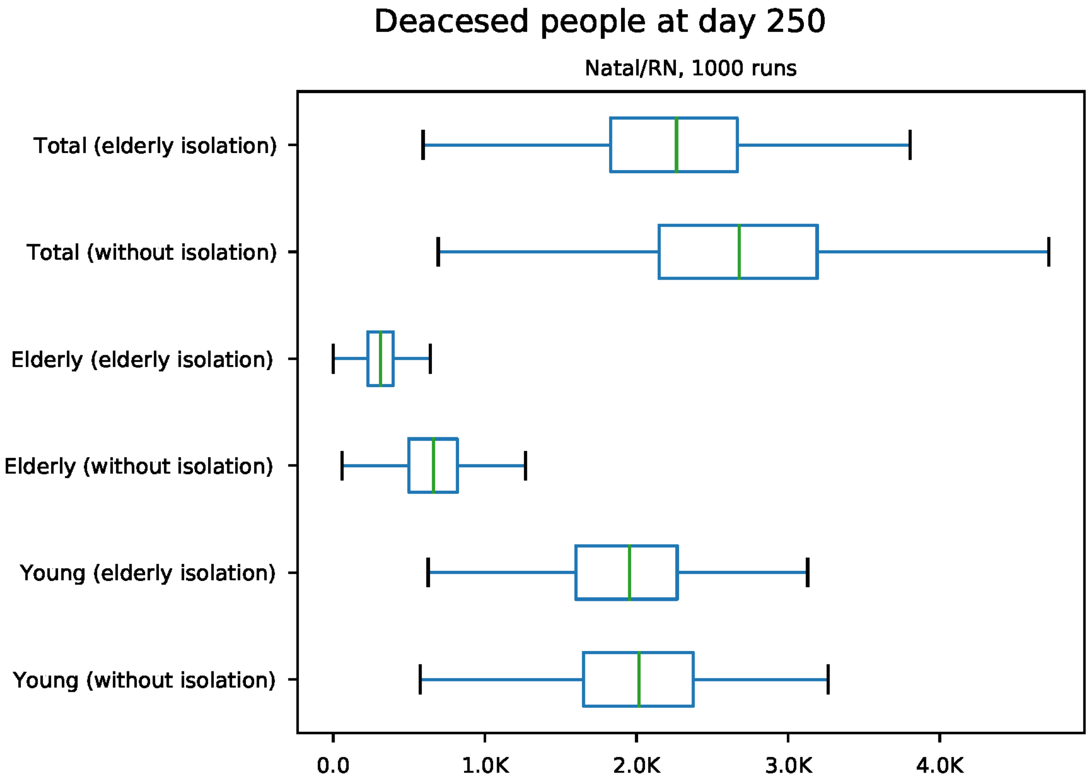


Figure S91: Predicted number of death at day 250 for Natal

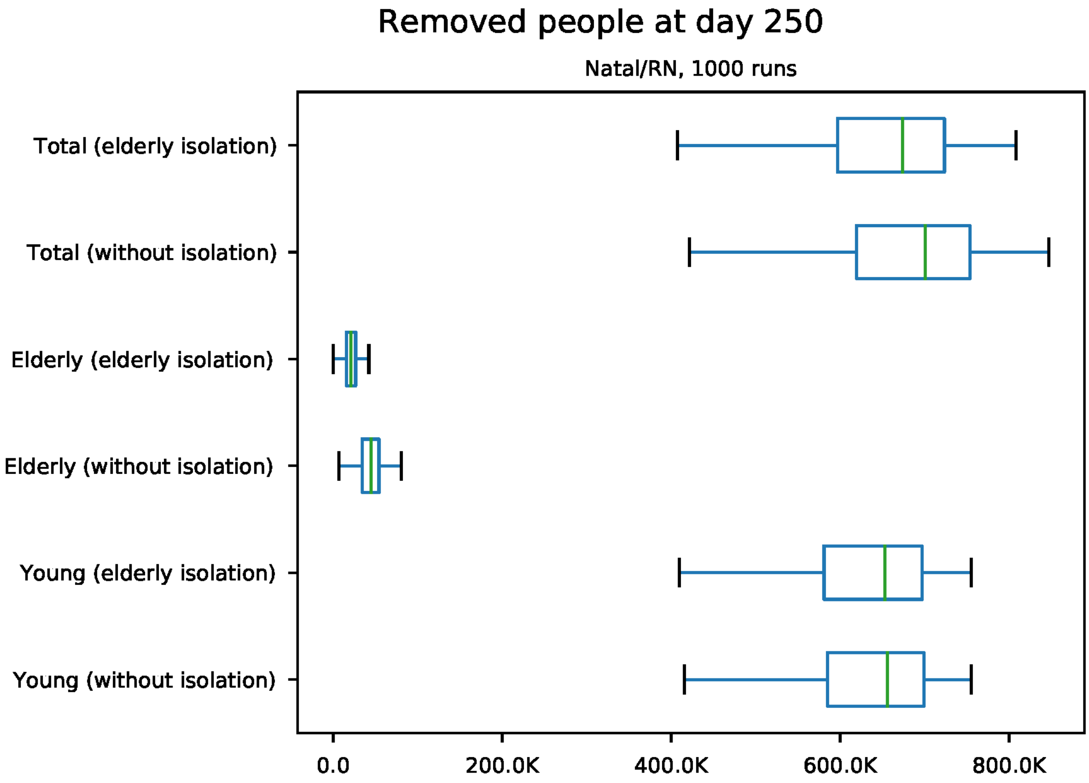


Figure S92: Predicted number of removed individuals at day 250 for Natal

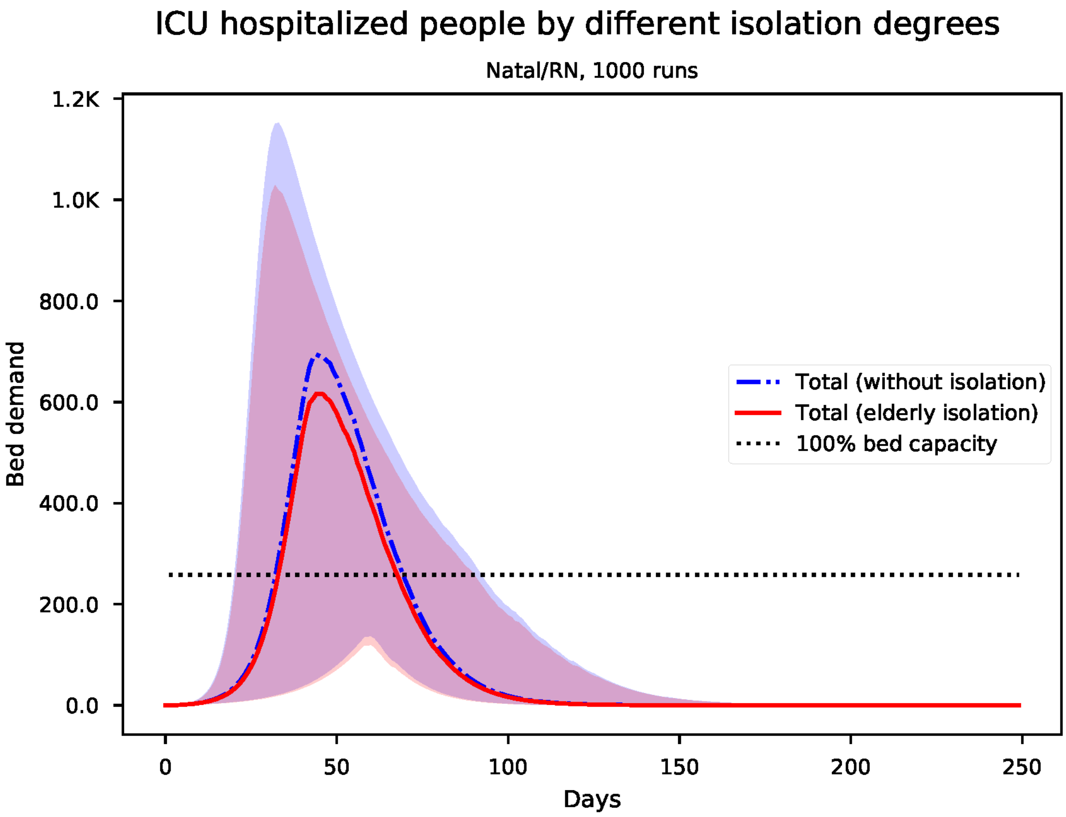


Figure S93: Predicted number of needed ICU beds for Natal

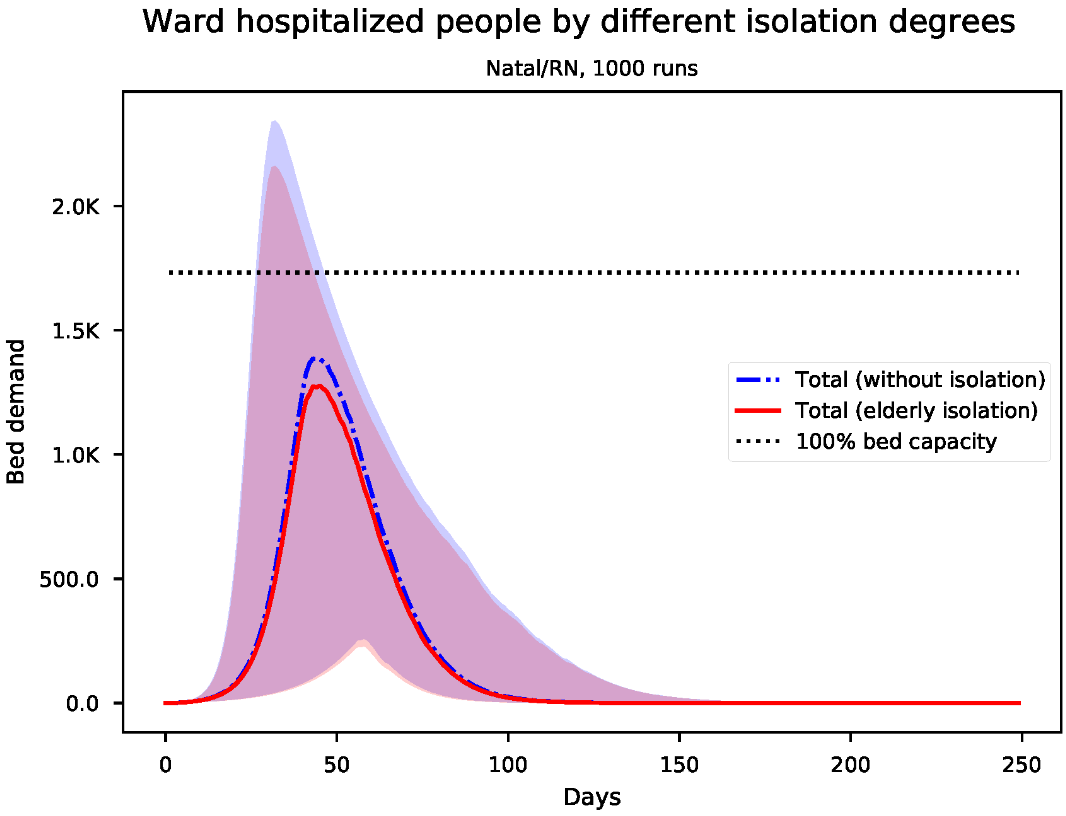


Figure S94: Predicted number of needed ward beds for Natal

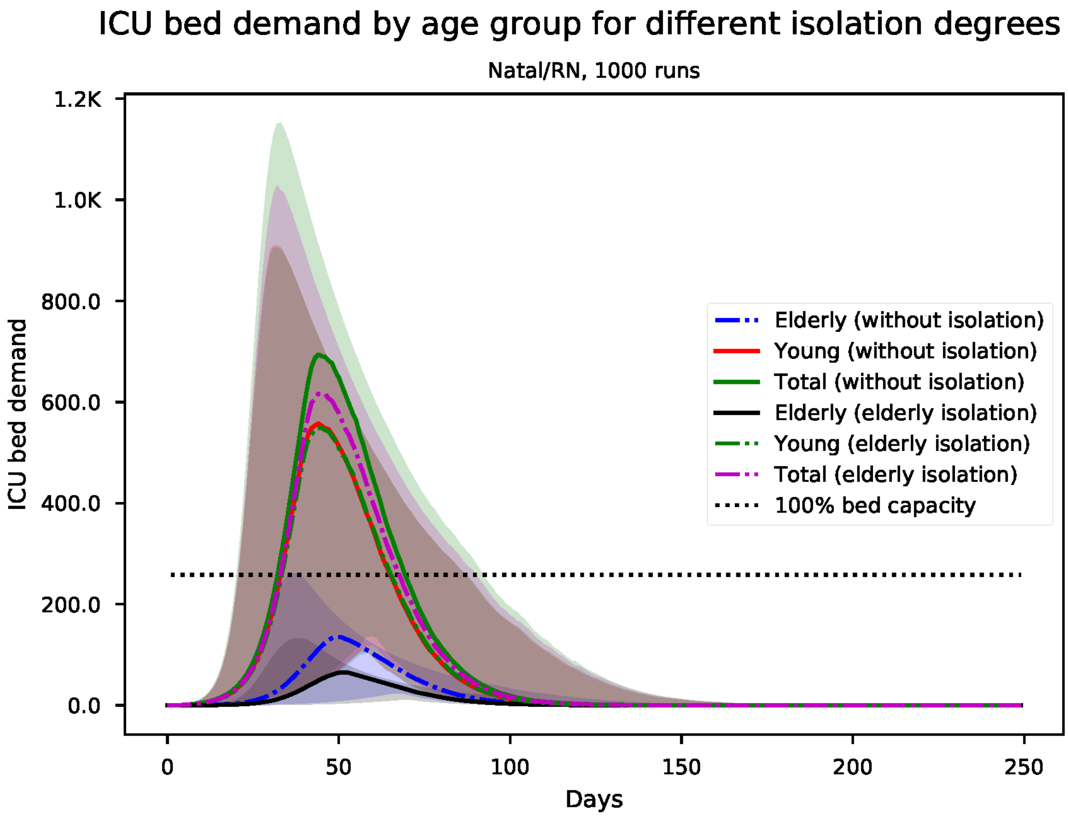


Figure S95: Predicted number of needed ICU beds, by age group for Natal

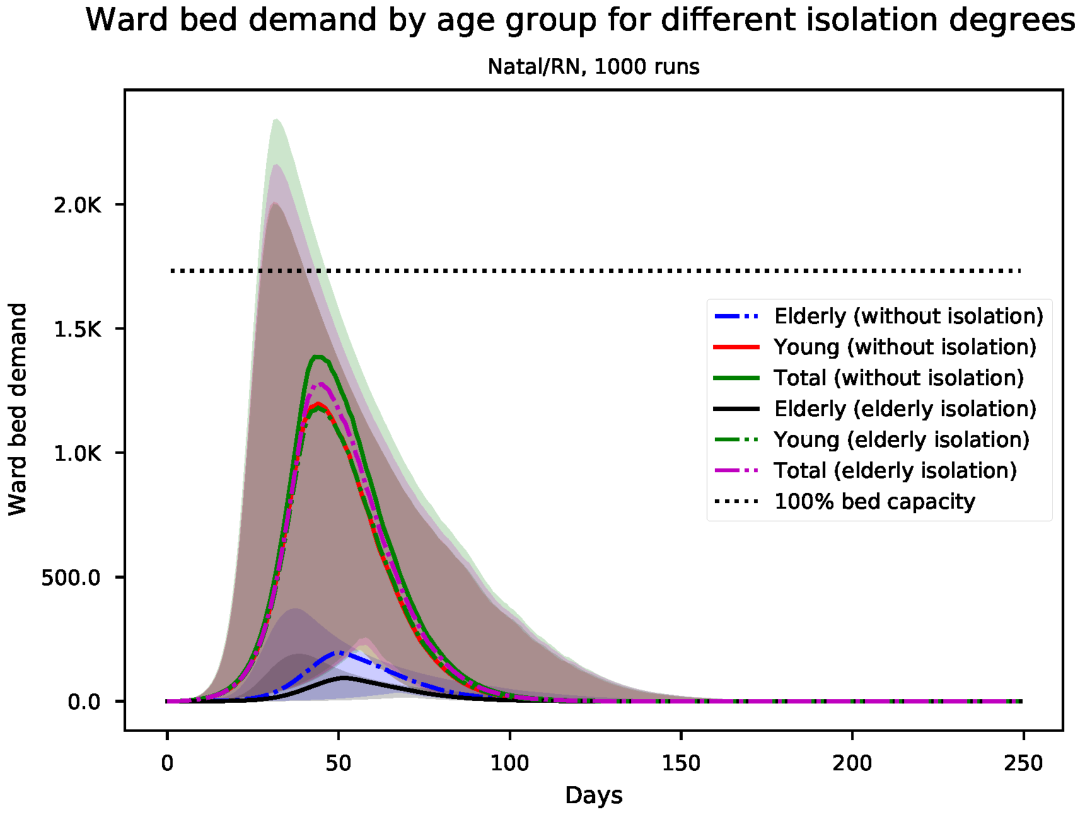


Figure S96: Predicted number of needed ward beds, by age group for Natal

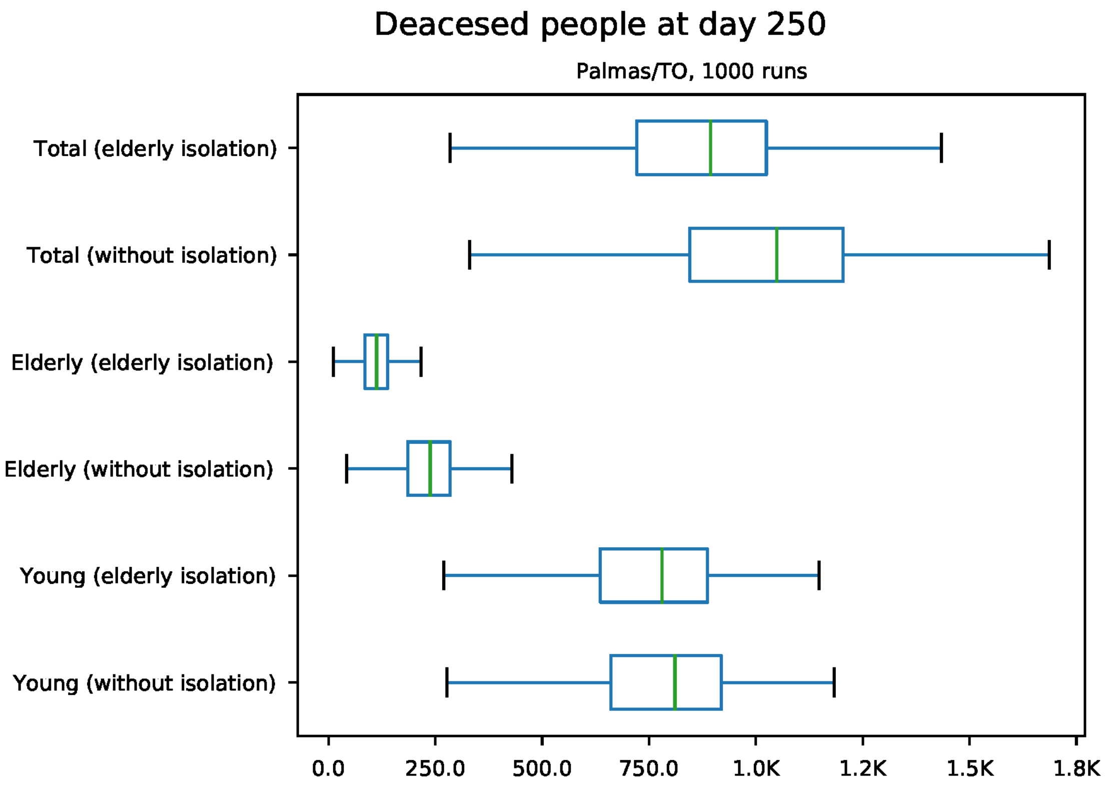


Figure S97: Predicted number of death at day 250 for Palmas

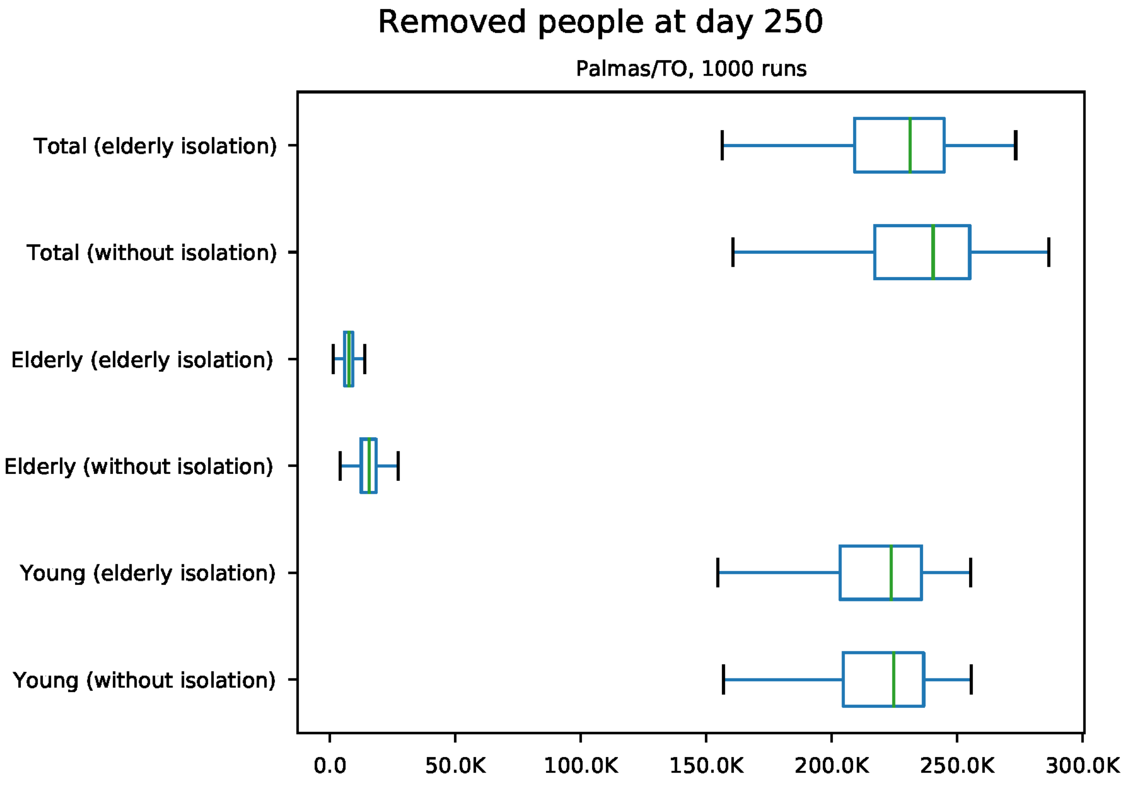


Figure S98: Predicted number of removed individuals at day 250 for Palmas

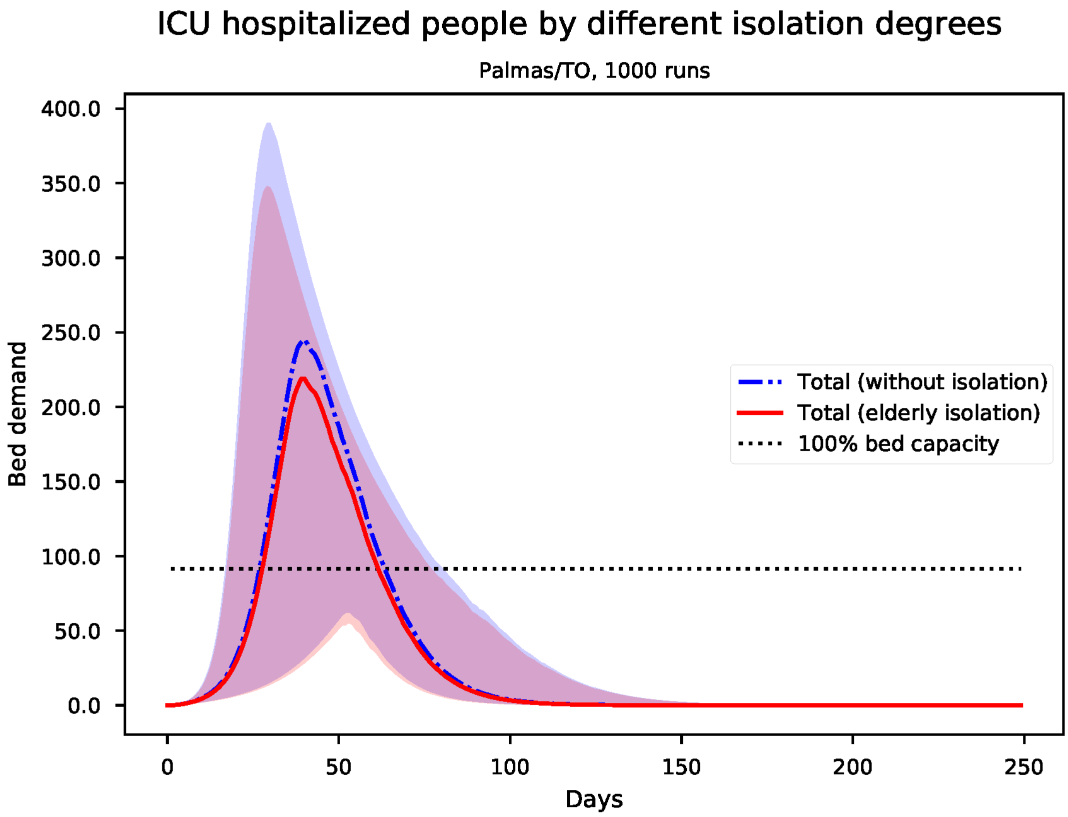


Figure S99: Predicted number of needed ICU beds for Palmas

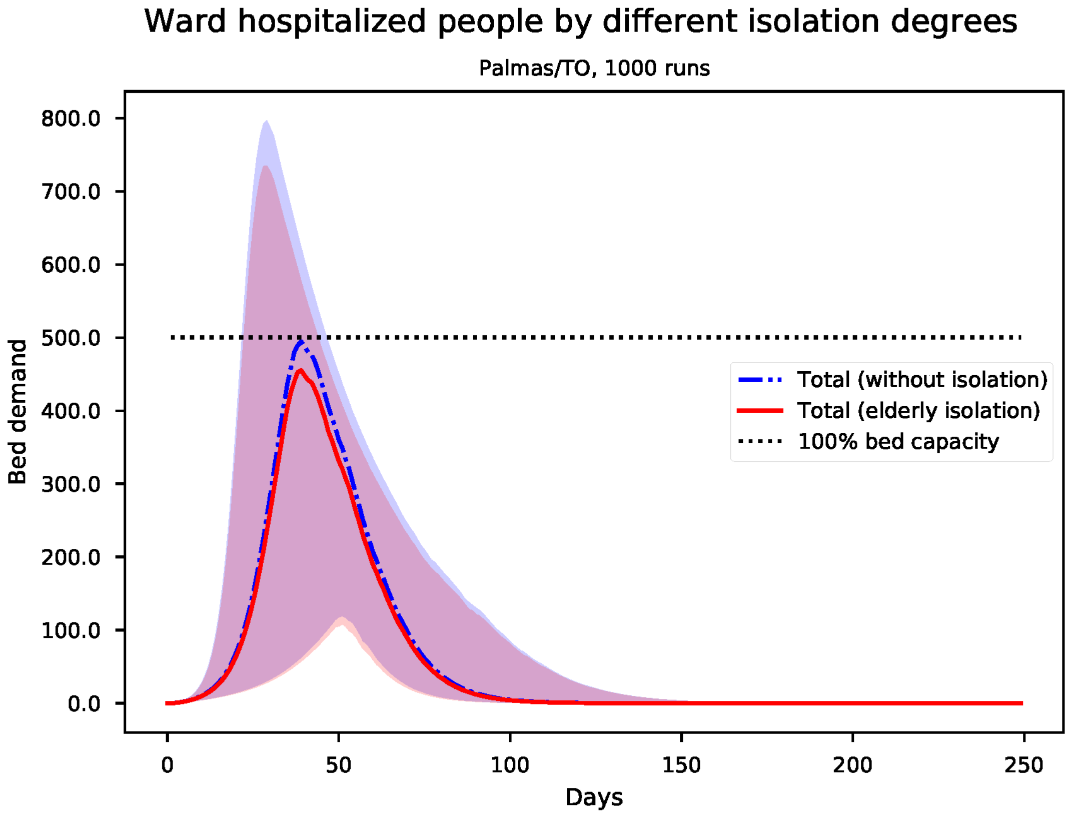


Figure S100: Predicted number of needed ward beds for Palmas

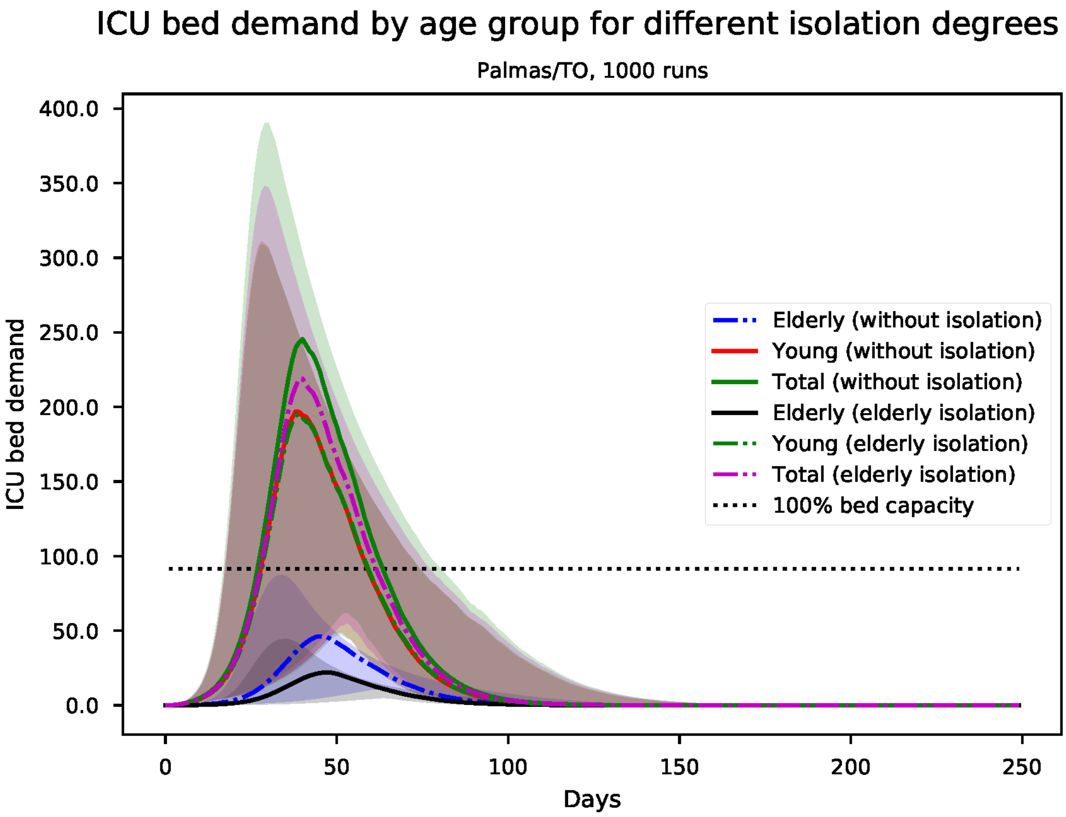


Figure S101: Predicted number of needed ICU beds, by age group for Palmas

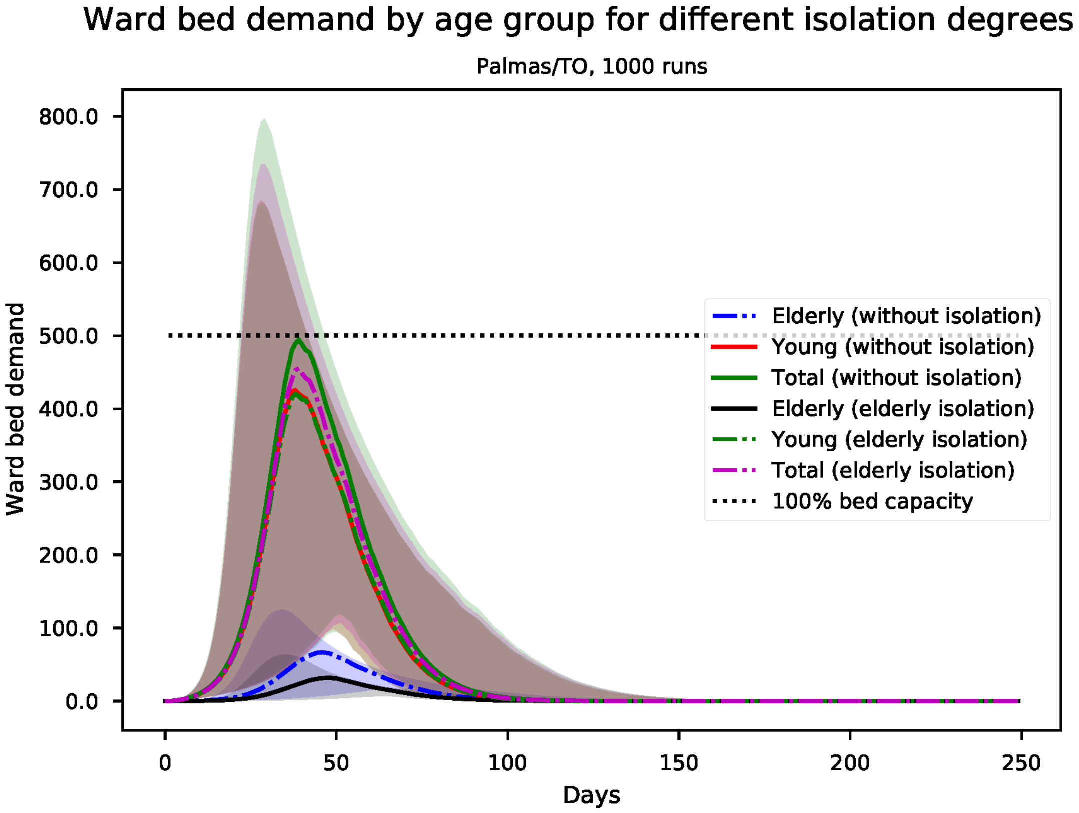


Figure S102: Predicted number of needed ward beds, by age group for Palmas

*S18. Results for Porto Alegre*

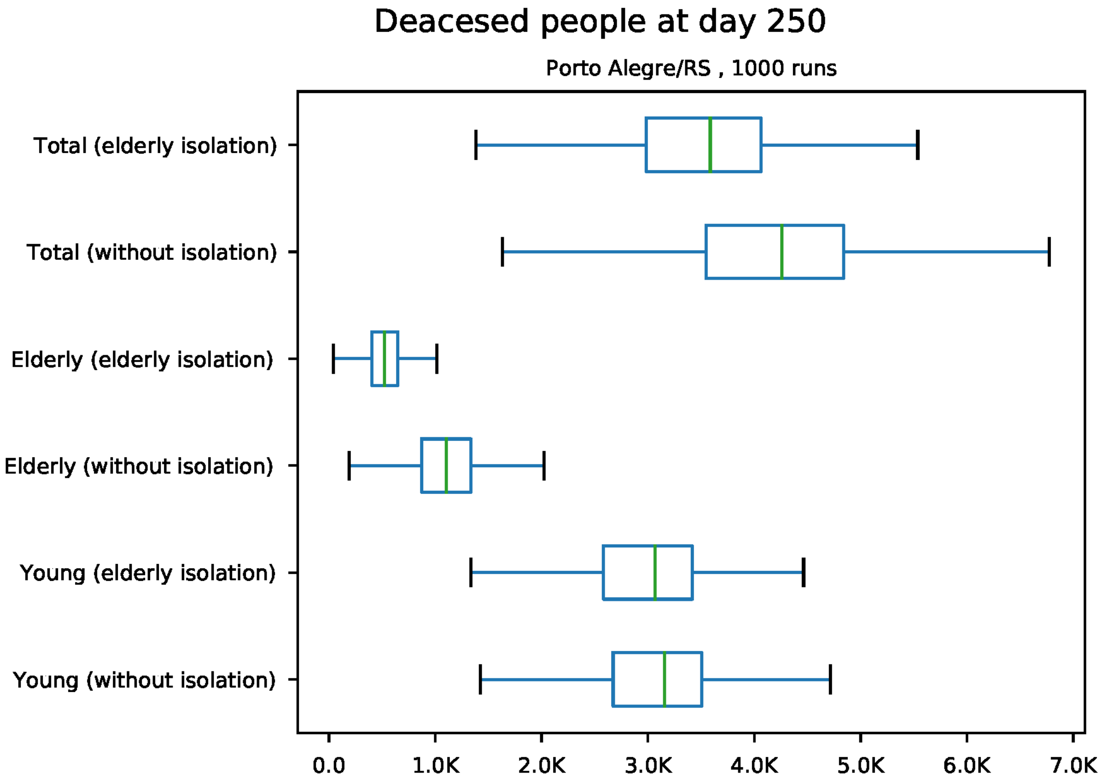


Figure S103: Predicted number of death at day 250 for Porto Alegre

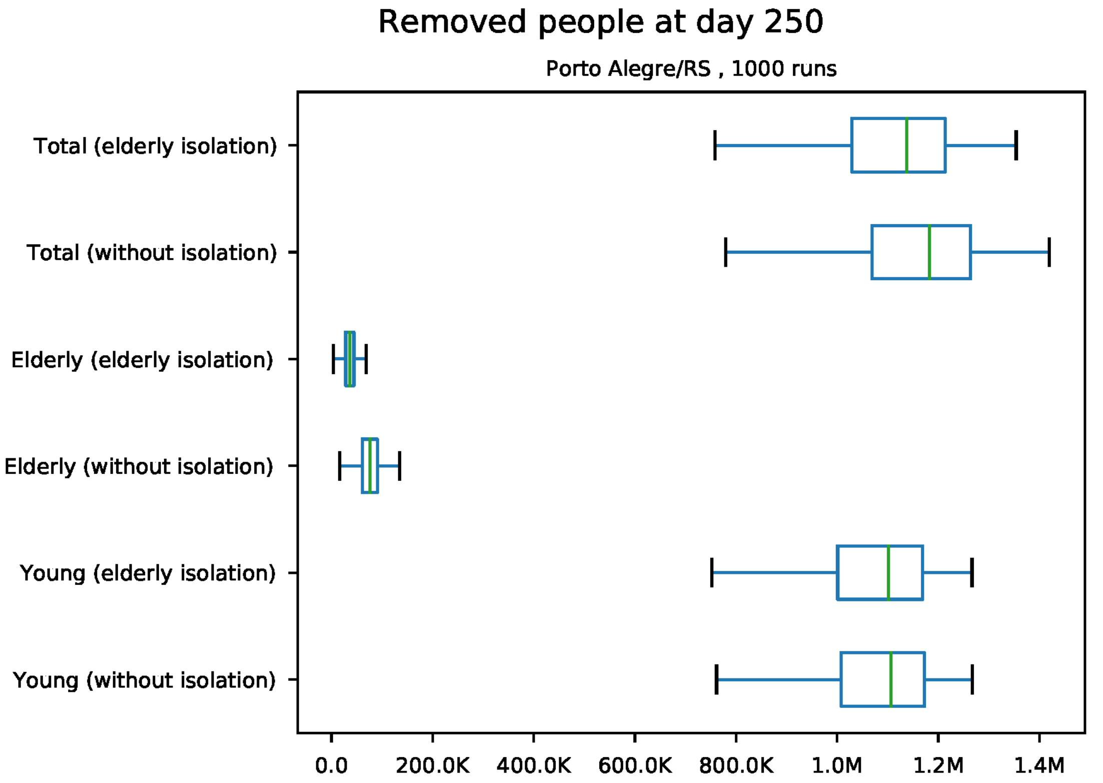


Figure S104: Predicted number of removed individuals at day 250 for Porto Alegre

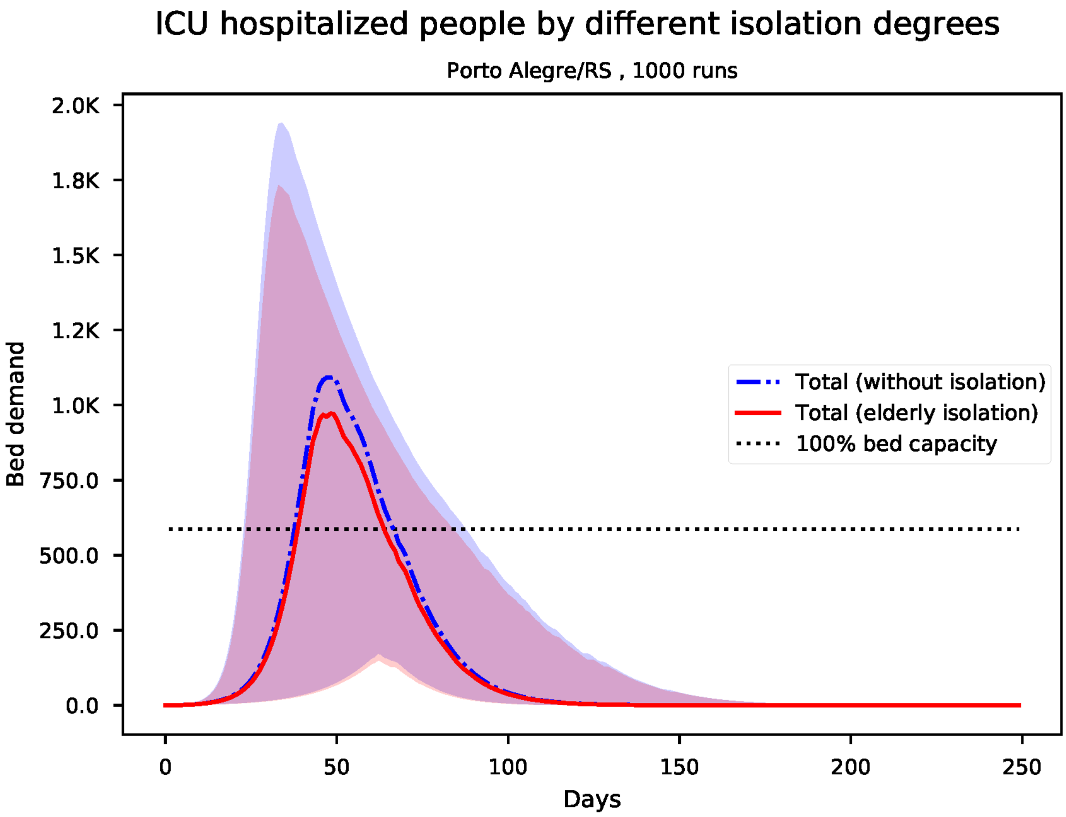


Figure S105: Predicted number of needed ICU beds for Porto Alegre

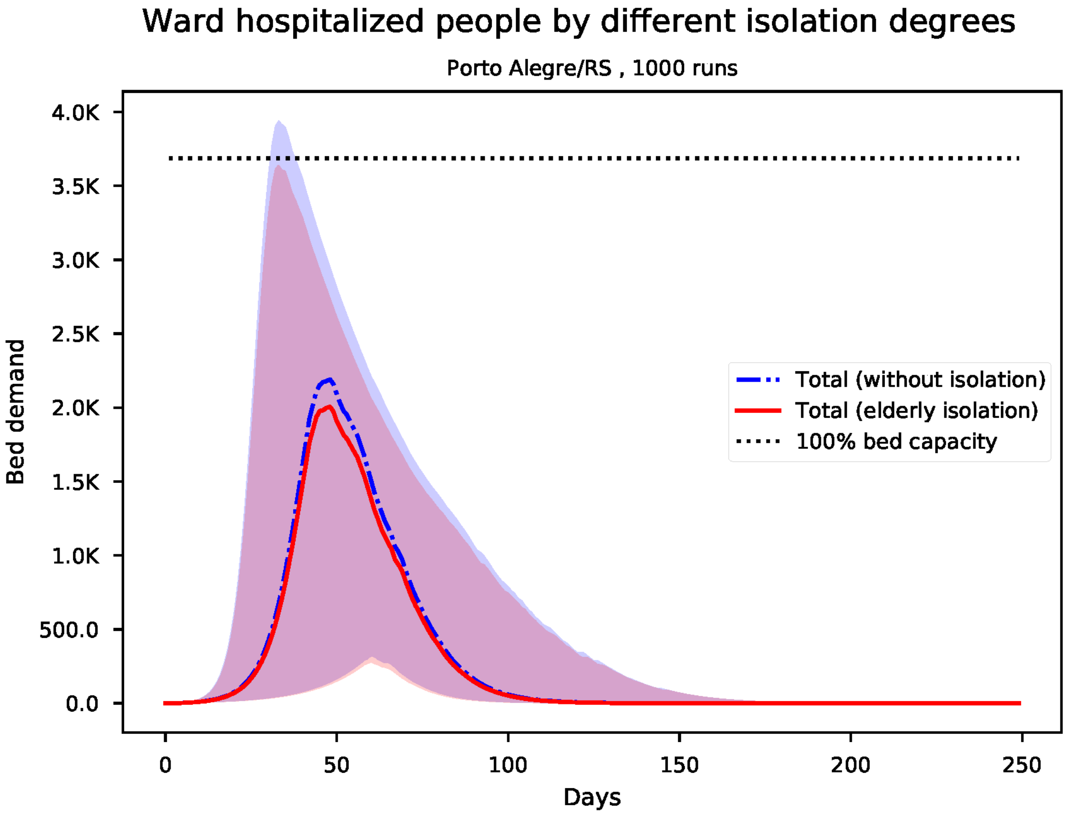


Figure S106: Predicted number of needed ward beds for Porto Alegre

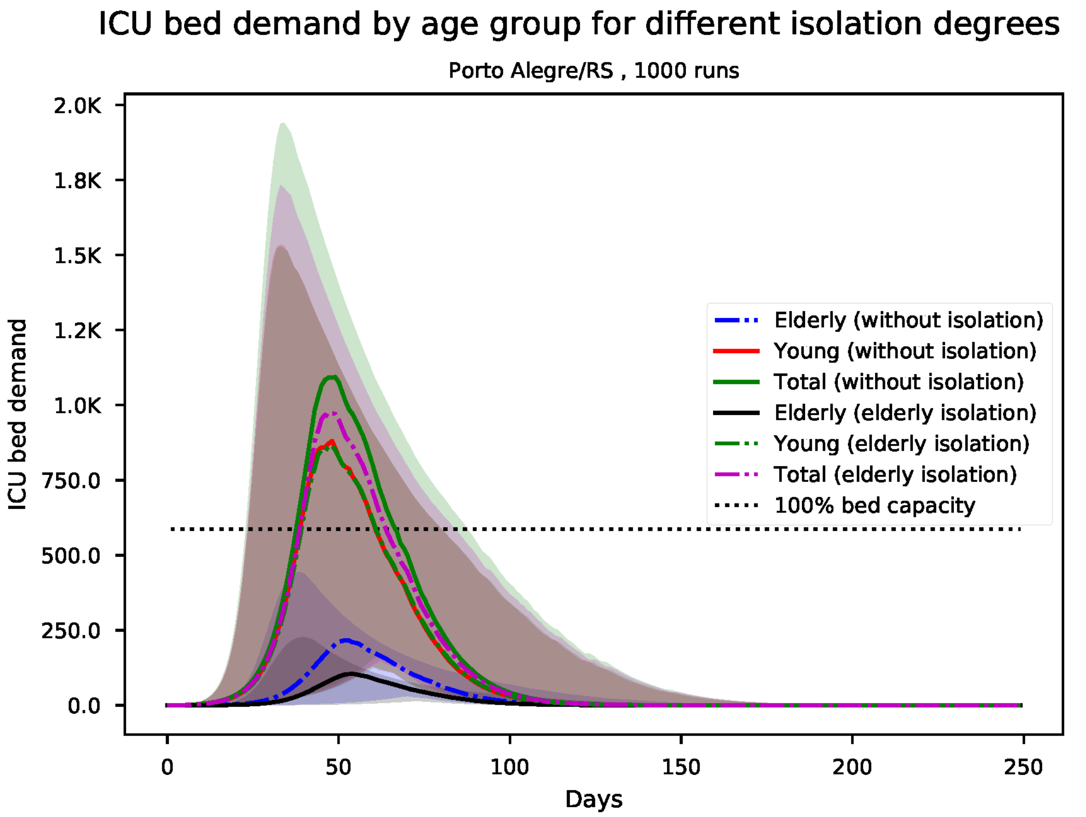


Figure S107: Predicted number of needed ICU beds, by age group for Porto Alegre

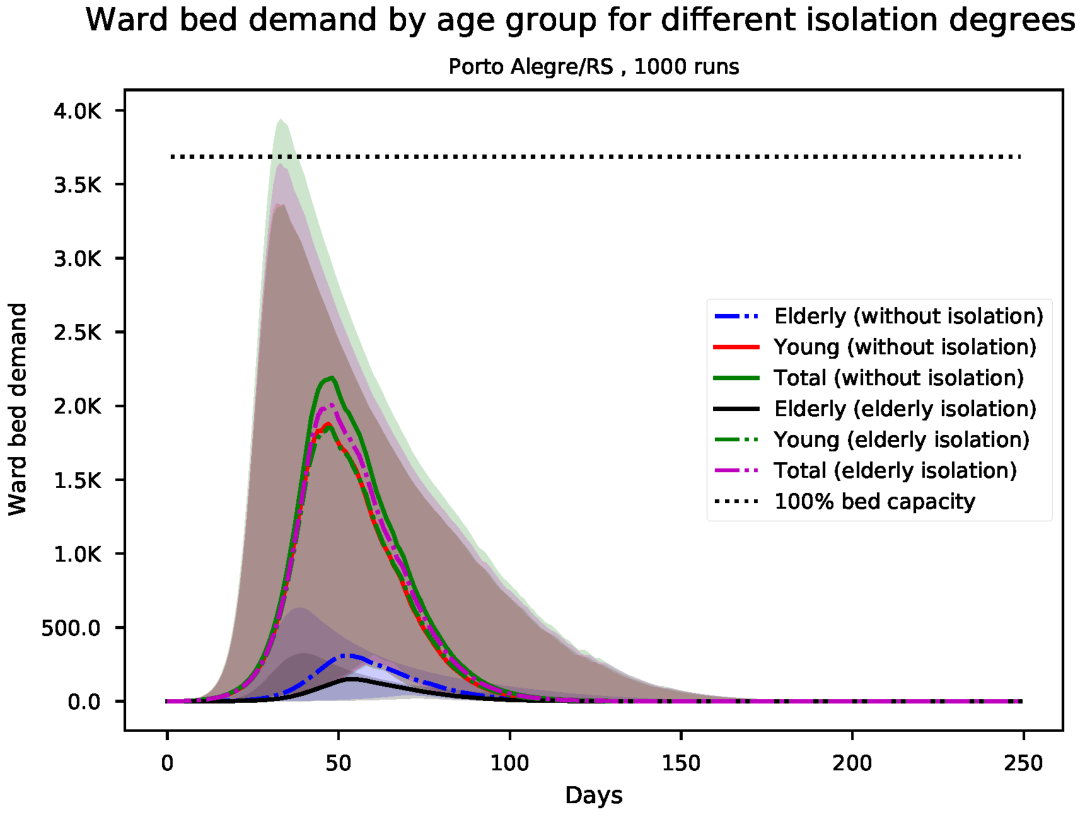


Figure S108: Predicted number of needed ward beds, by age group for Porto Alegre

*S19. Results for Porto Velho*

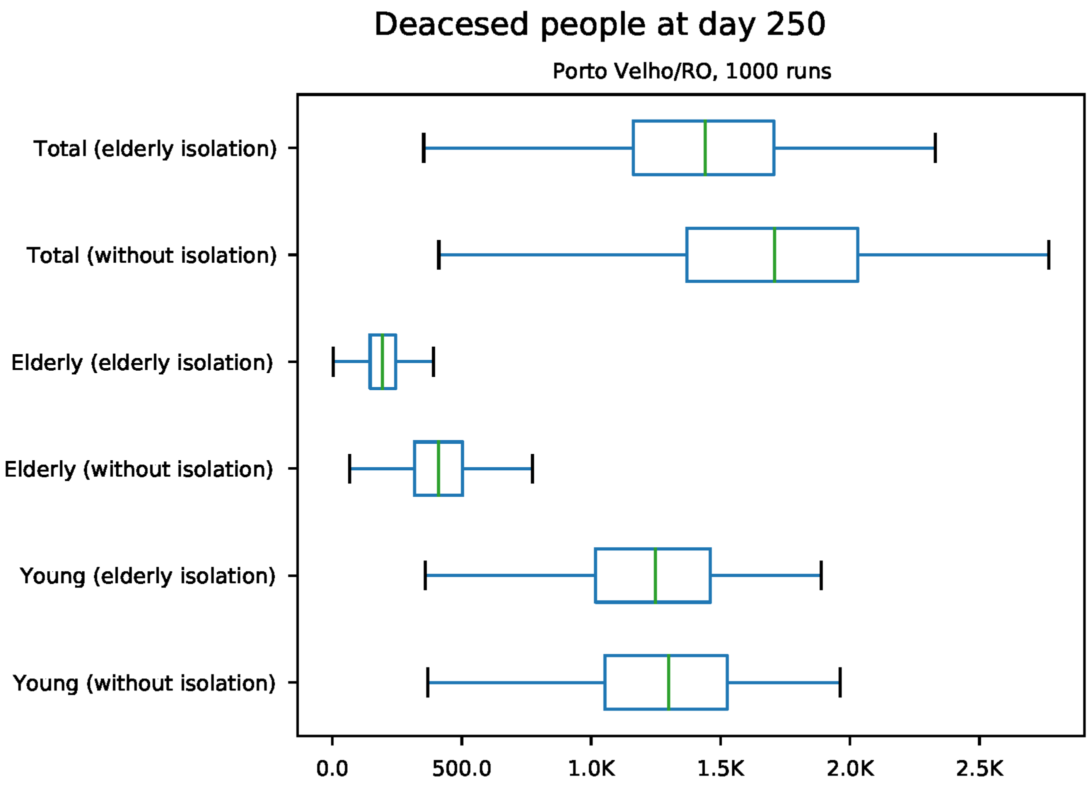


Figure S109: Predicted number of death at day 250 for Porto Velho

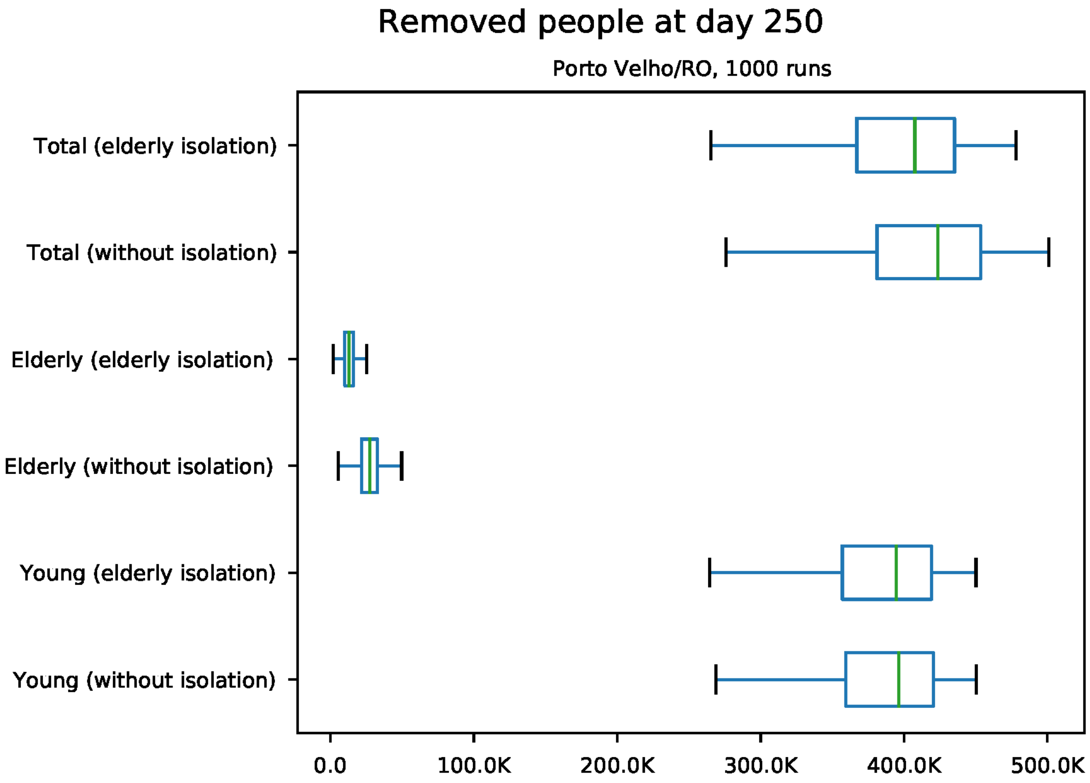


Figure S110: Predicted number of removed individuals at day 250 for Porto Velho

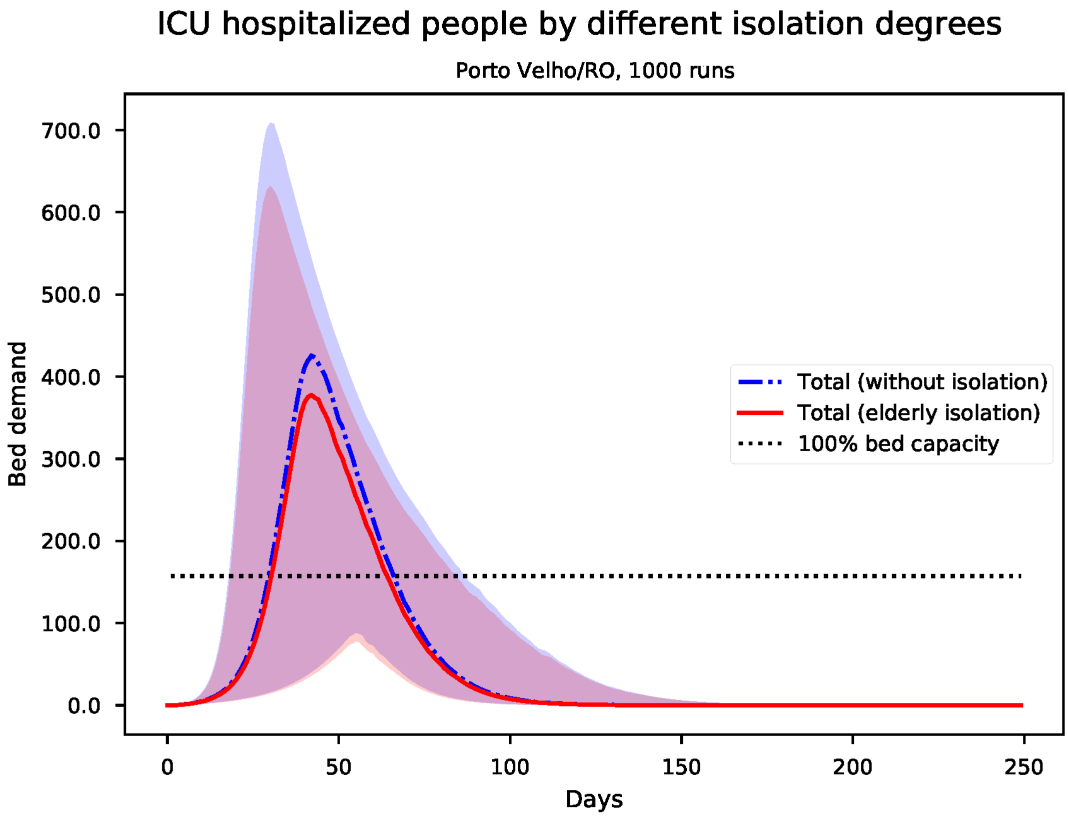


Figure S111: Predicted number of needed ICU beds for Porto Velho

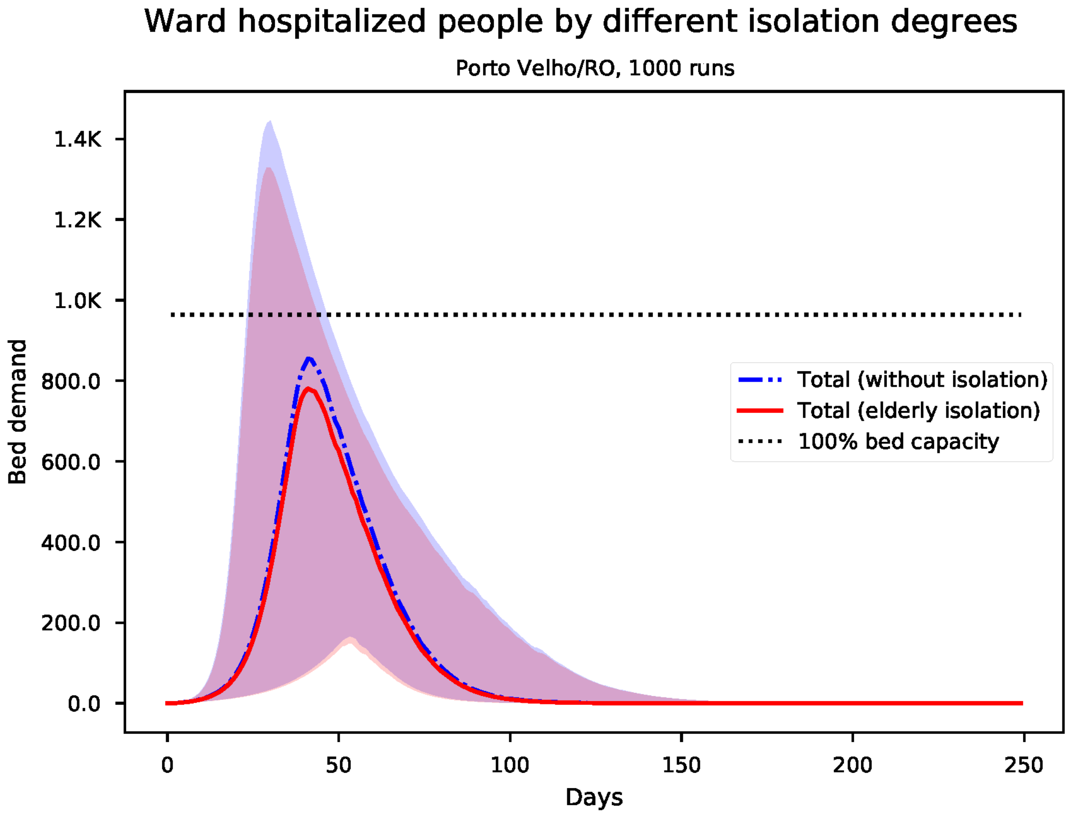


Figure S112: Predicted number of needed ward beds for Porto Velho

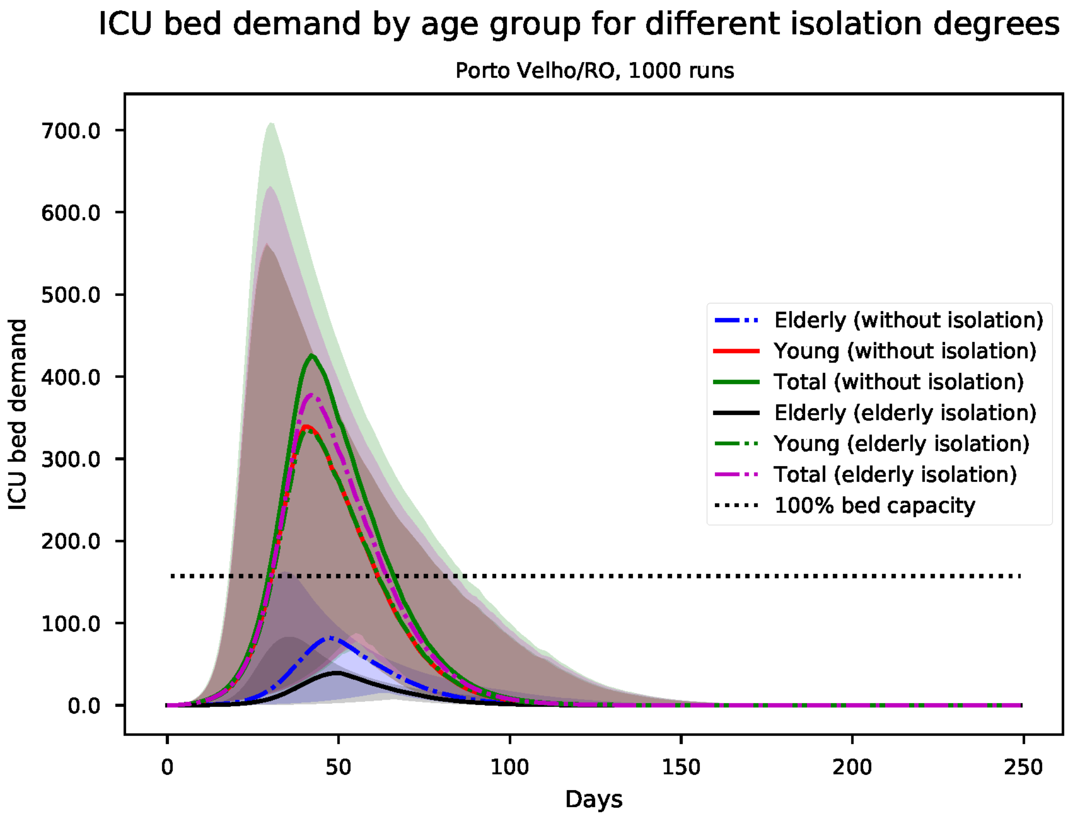


Figure S113: Predicted number of needed ICU beds, by age group for Porto Velho

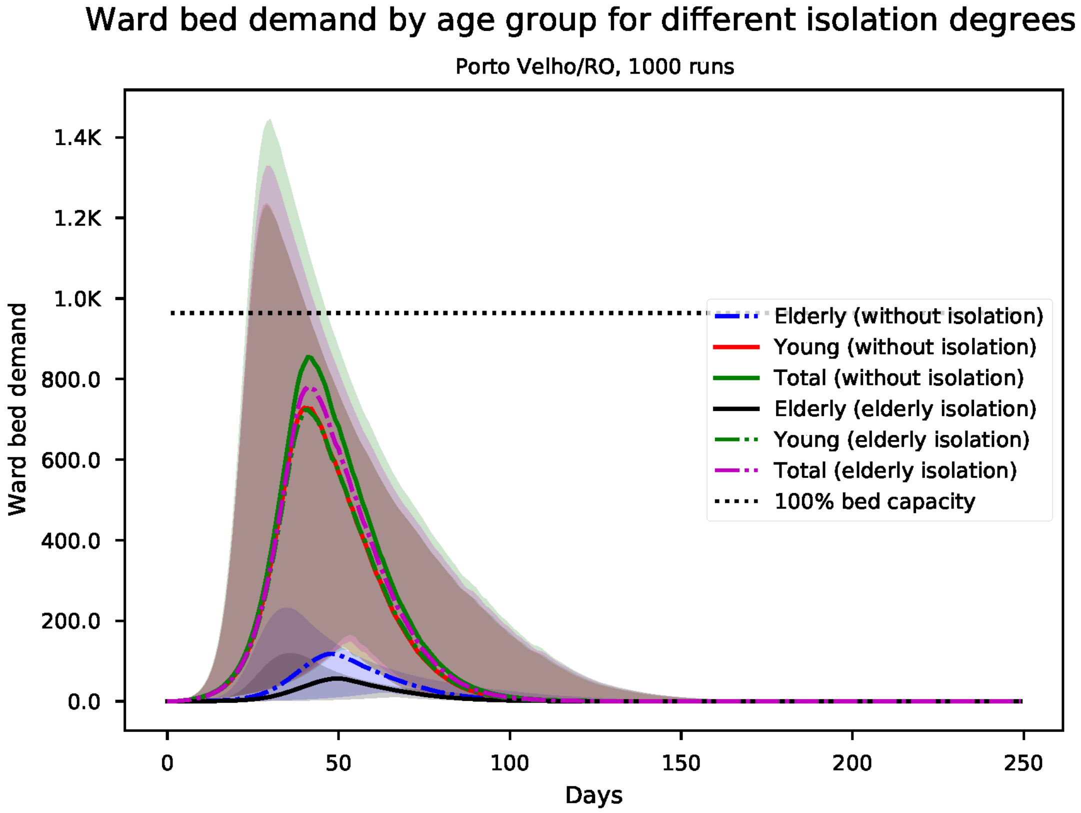


Figure S114: Predicted number of needed ward beds, by age group for Porto Velho

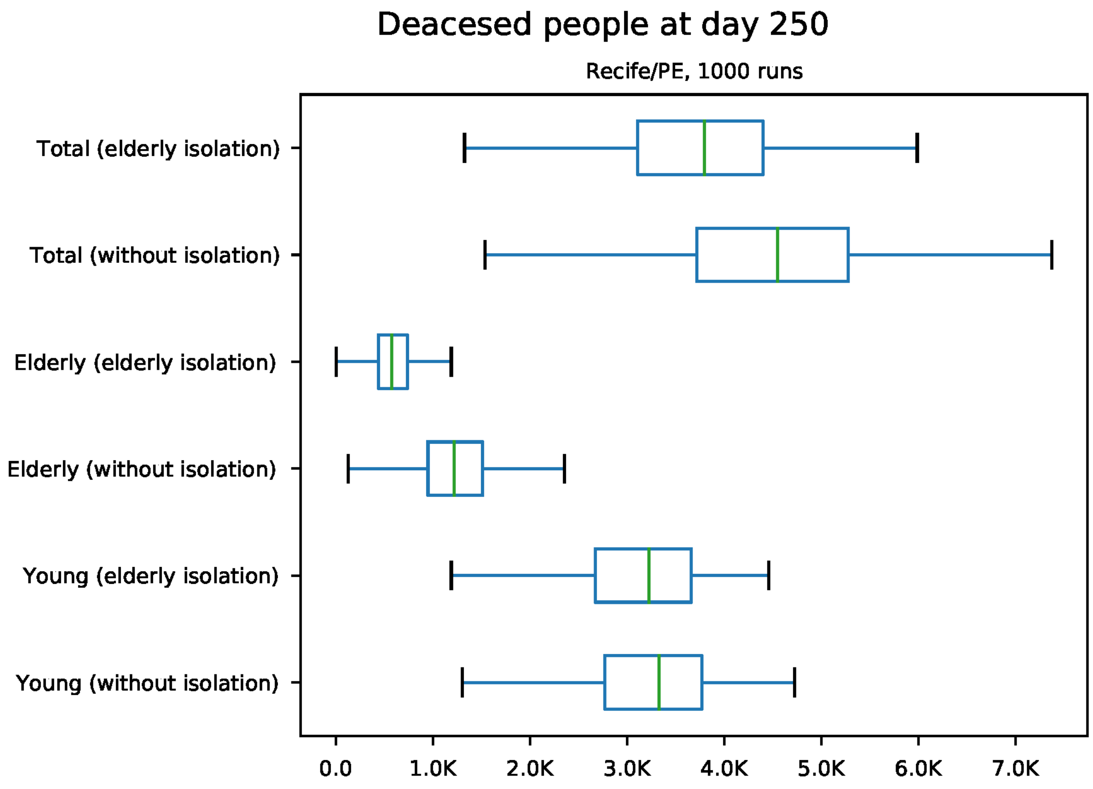


Figure S115: Predicted number of death at day 250 for Recife

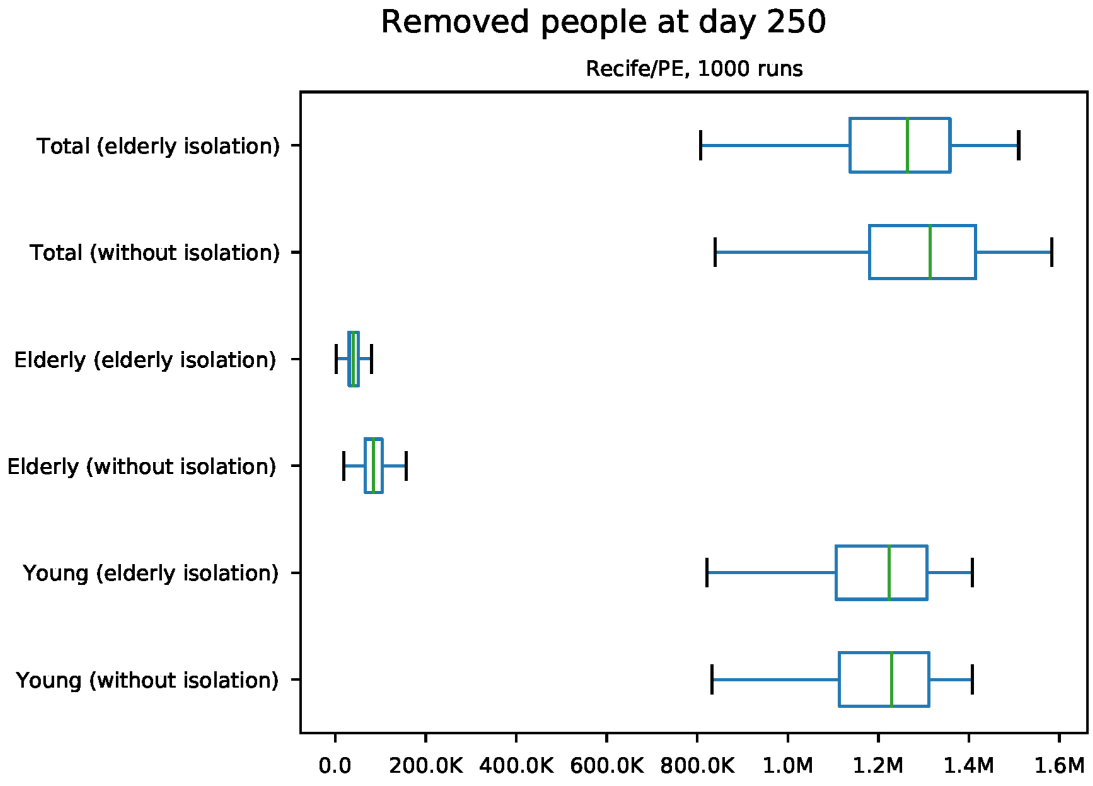


Figure S116: Predicted number of removed individuals at day 250 for Recife

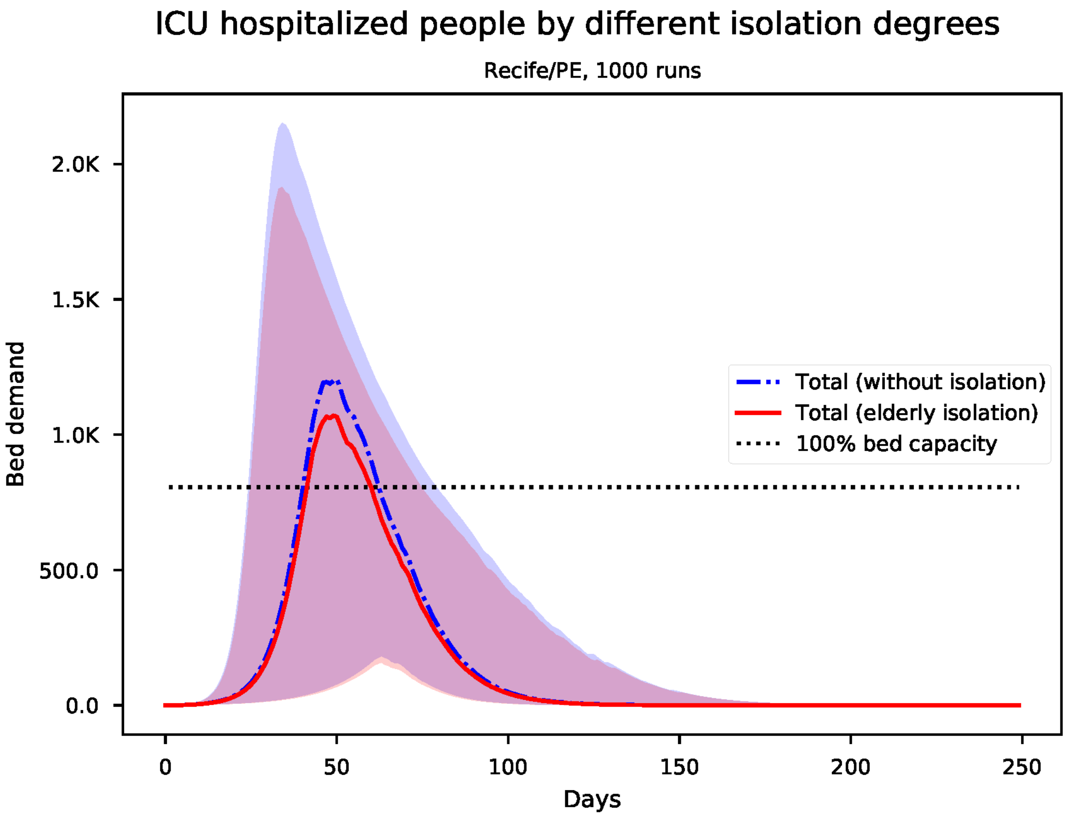


Figure S117: Predicted number of needed ICU beds for Recife

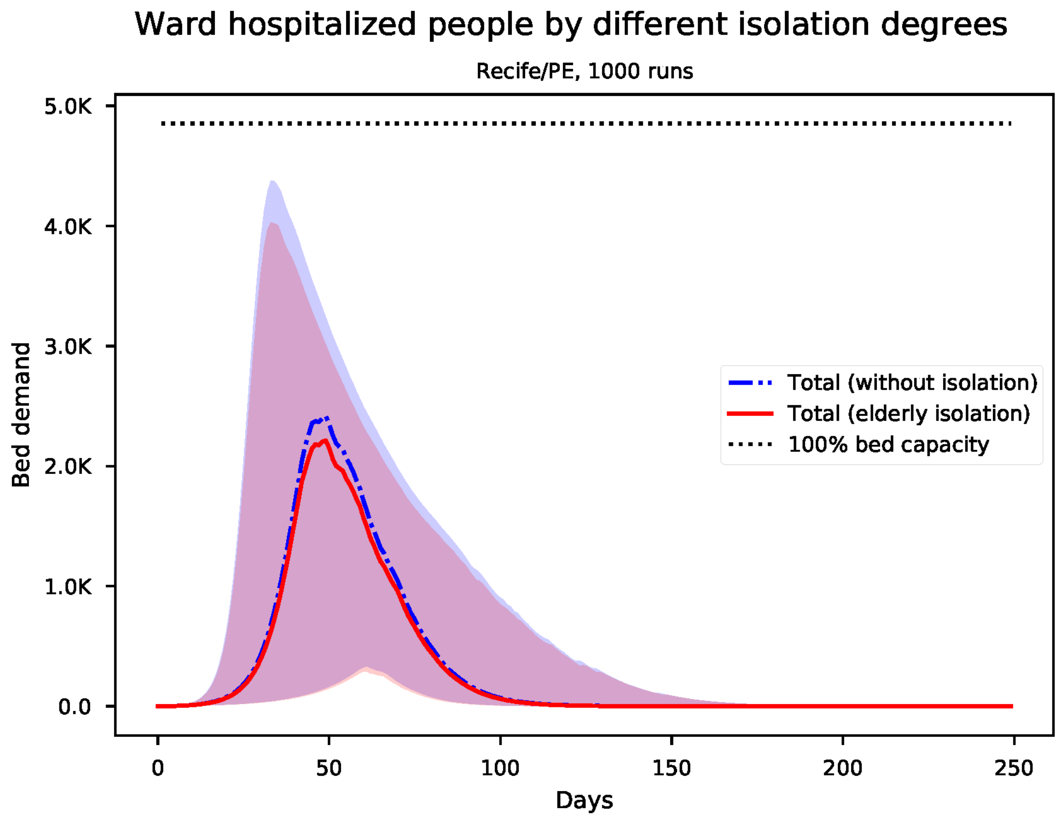


Figure S118: Predicted number of needed ward beds for Recife

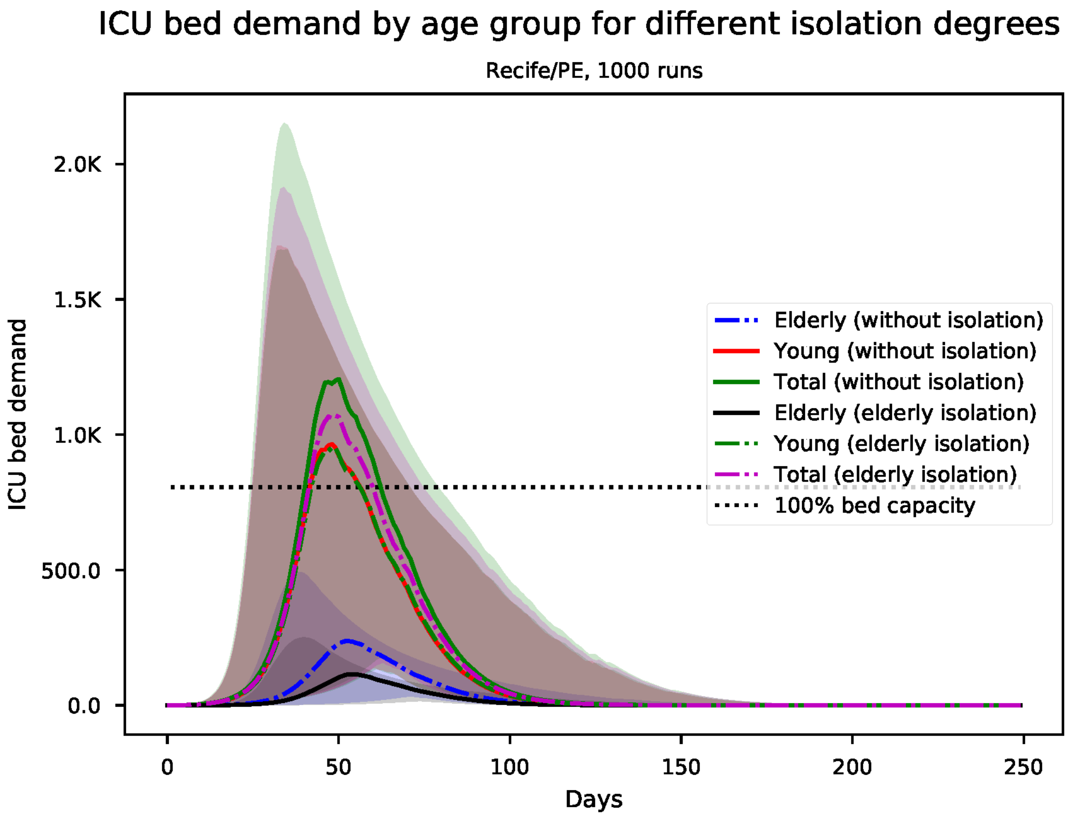


Figure S119: Predicted number of needed ICU beds, by age group for Recife

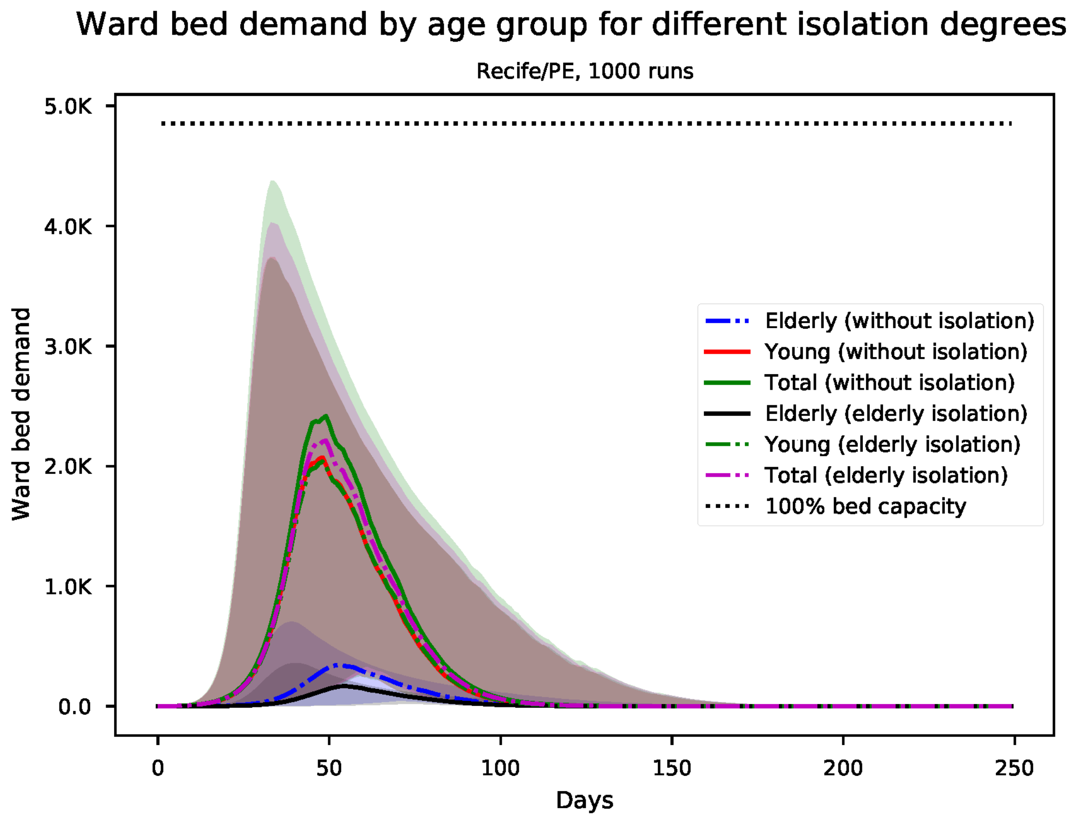


Figure S120: Predicted number of needed ward beds, by age group for Recife

*S21. Results for Rio Branco*

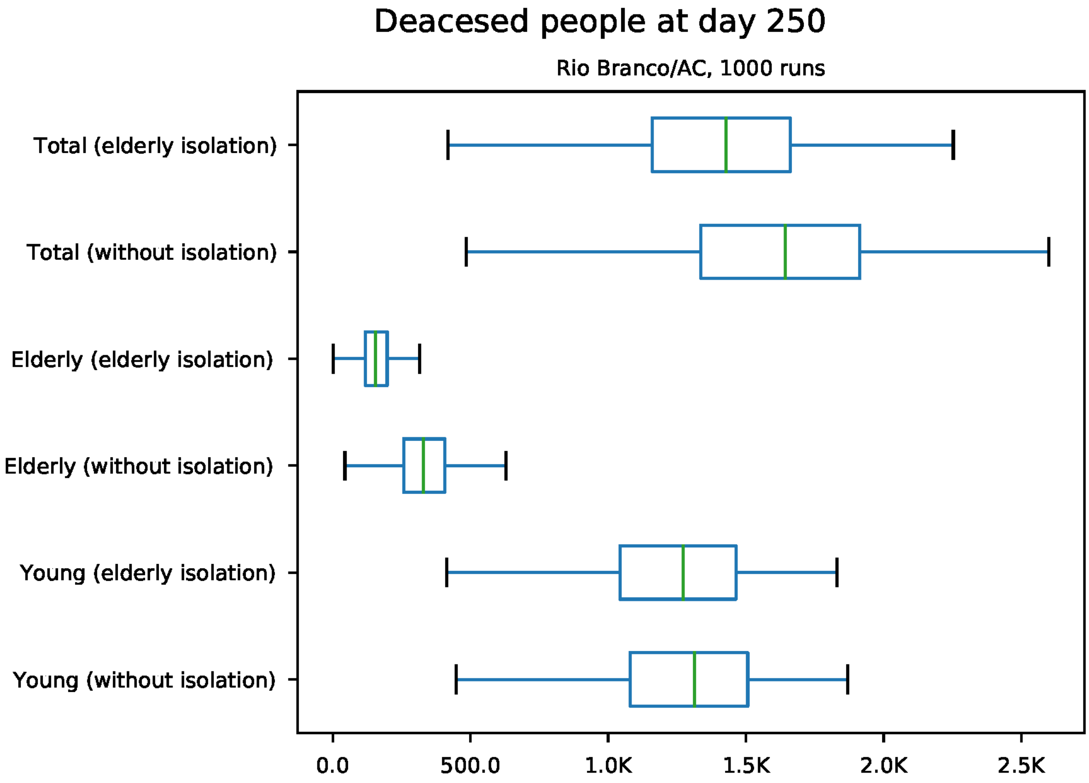


Figure S121: Predicted number of death at day 250 for Rio Branco

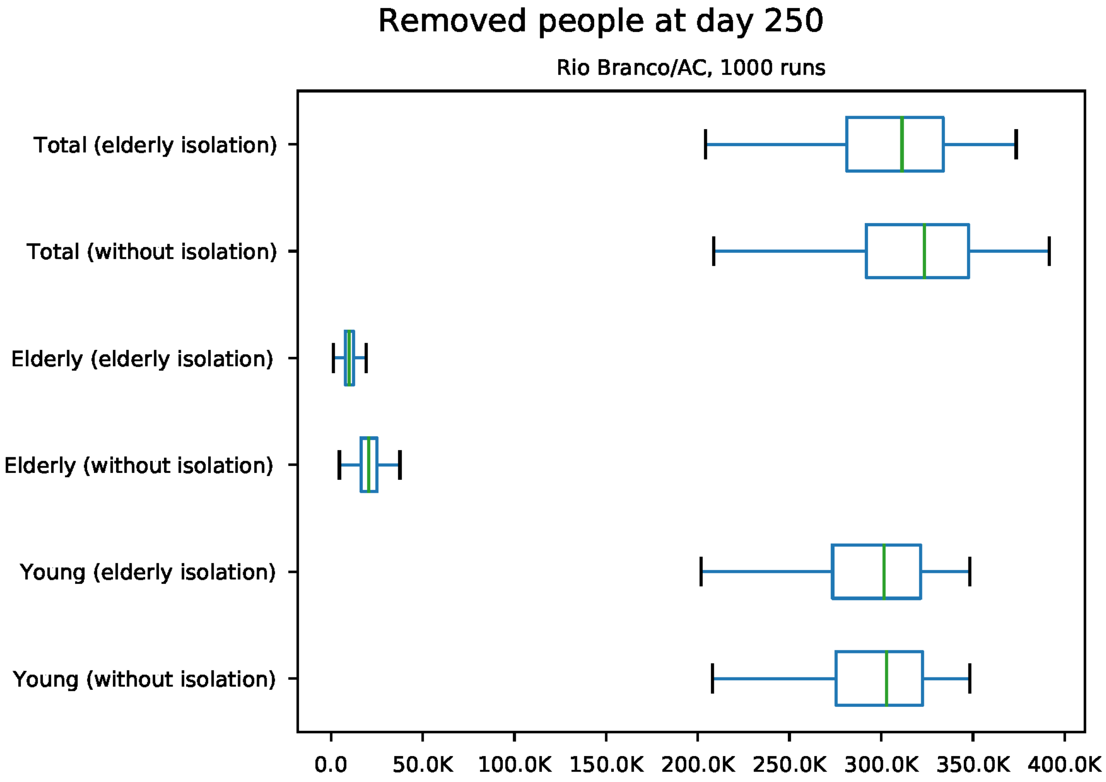


Figure S122: Predicted number of removed individuals at day 250 for Rio Branco

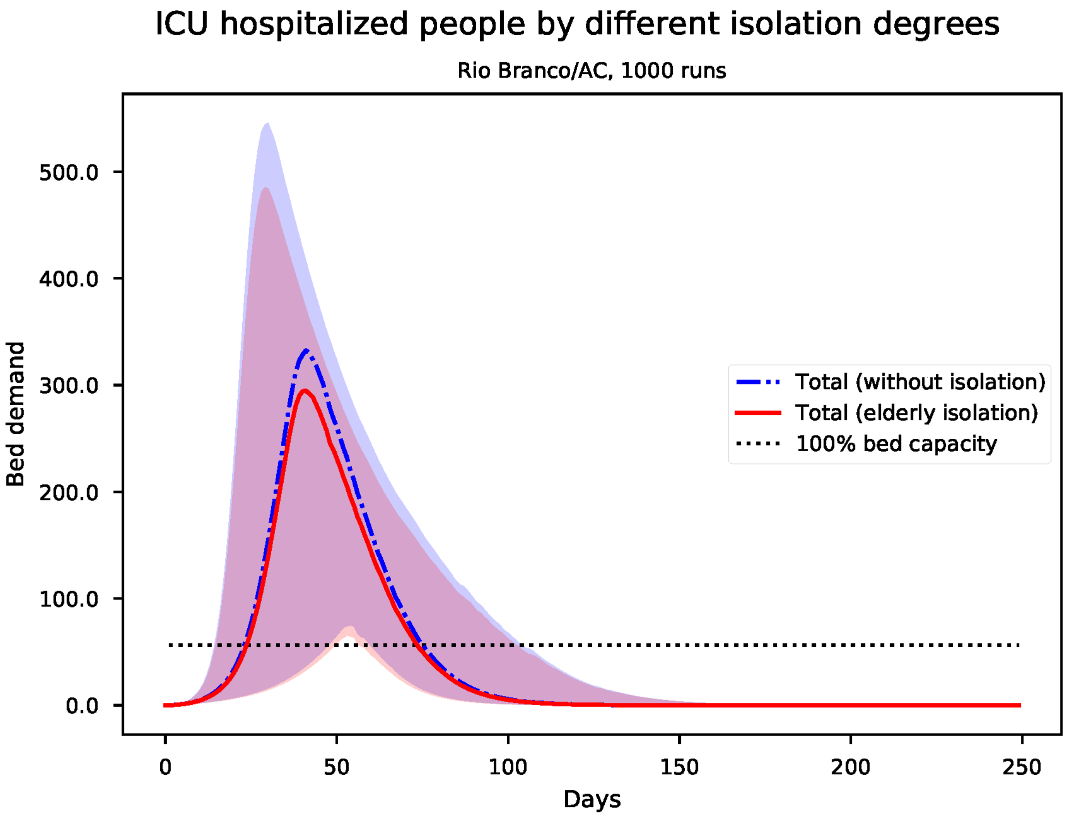


Figure S123: Predicted number of needed ICU beds for Rio Branco

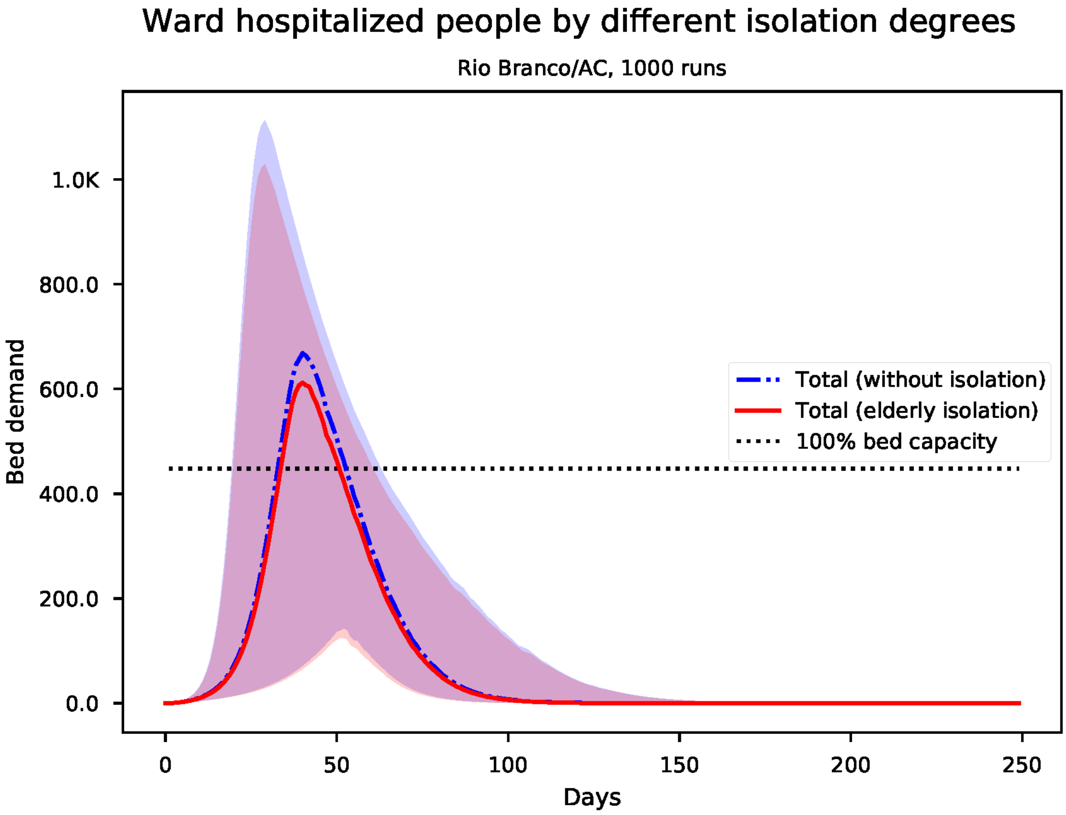


Figure S124: Predicted number of needed ward beds for Rio Branco

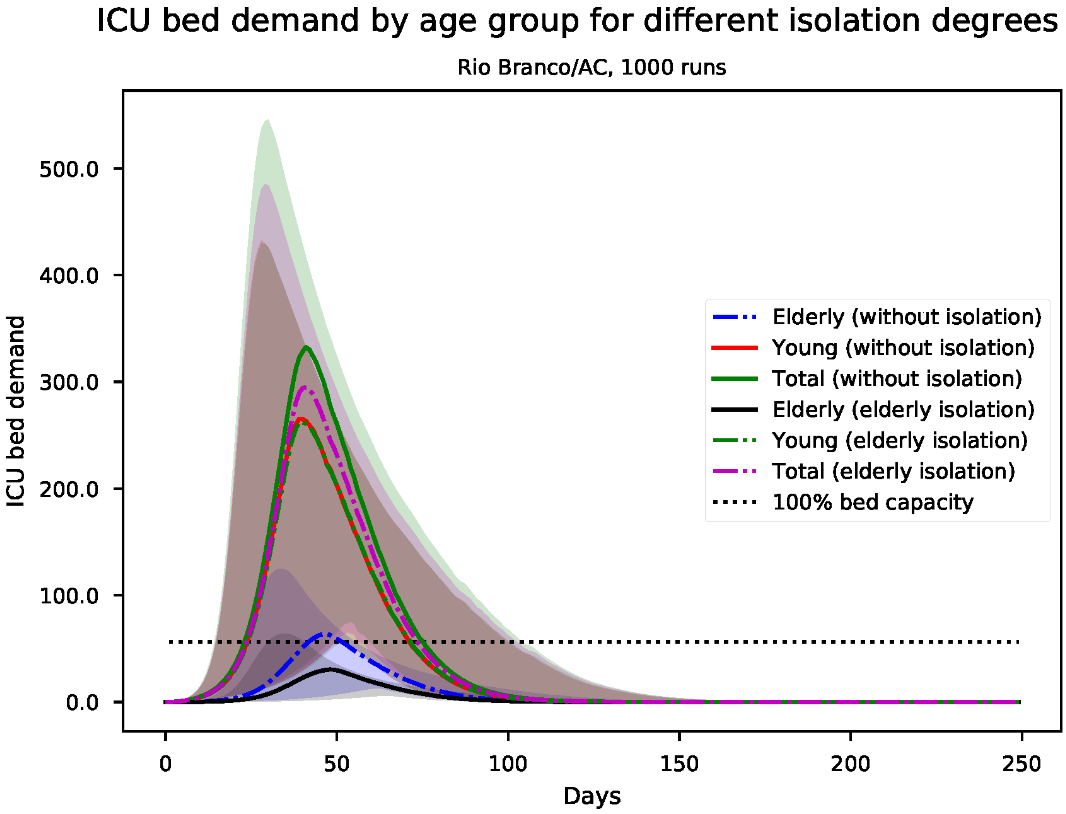


Figure S125: Predicted number of needed ICU beds, by age group for Rio Branco

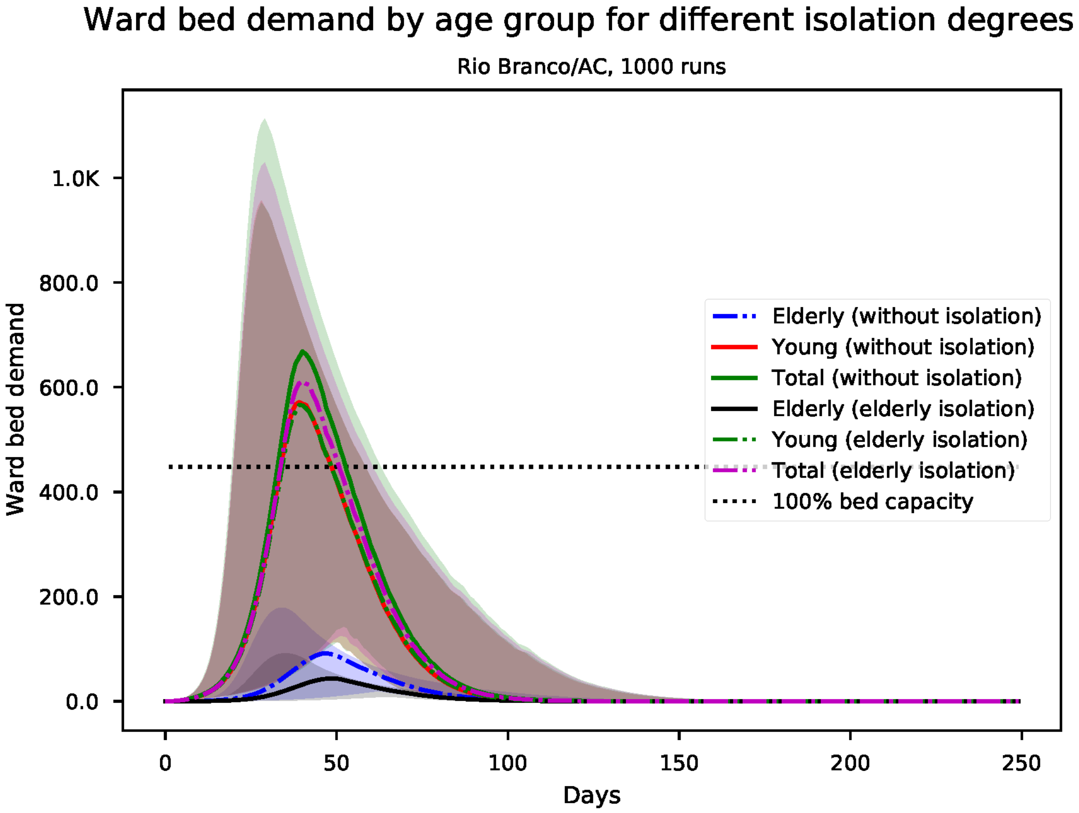


Figure S126: Predicted number of needed ward beds, by age group for Rio Branco

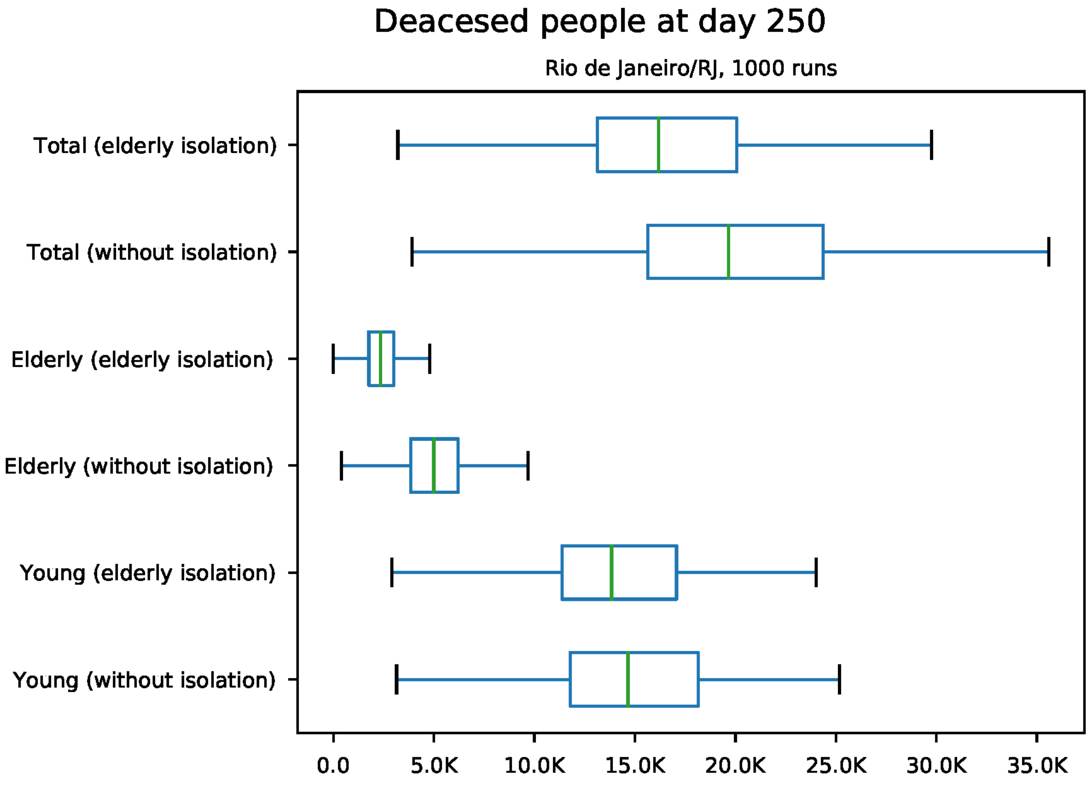


Figure S127: Predicted number of death at day 250 for Rio de Janeiro

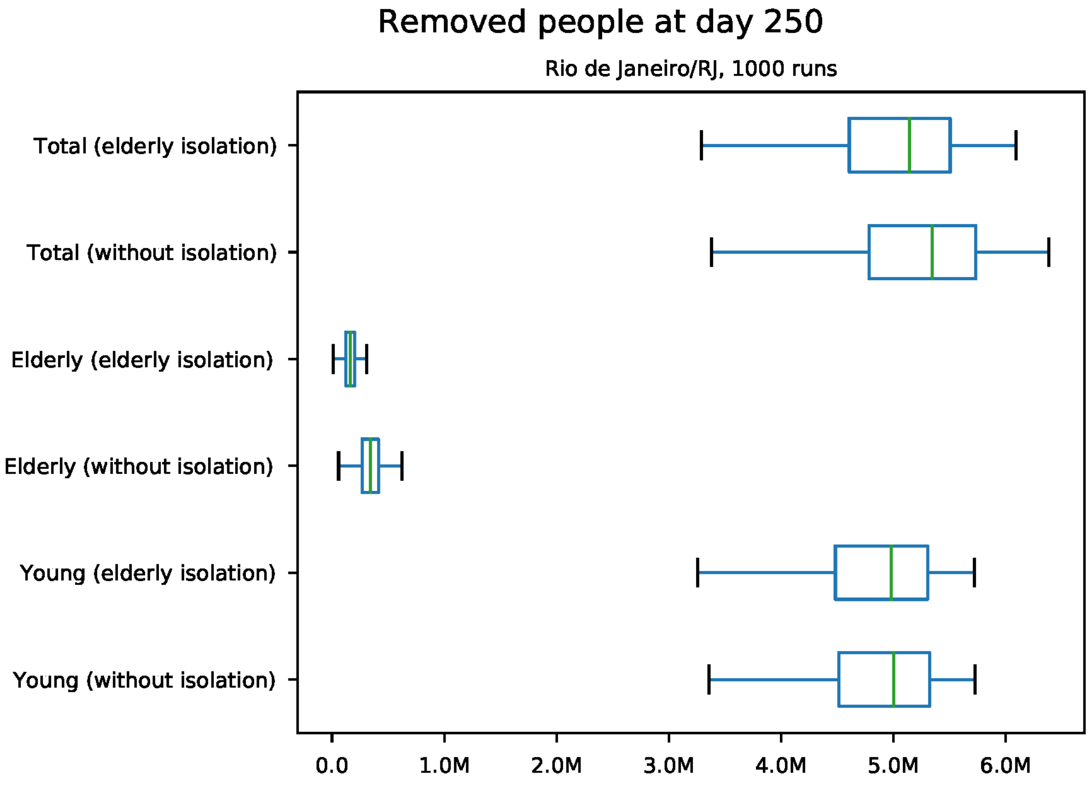


Figure S128: Predicted number of removed individuals at day 250 for Rio de Janeiro

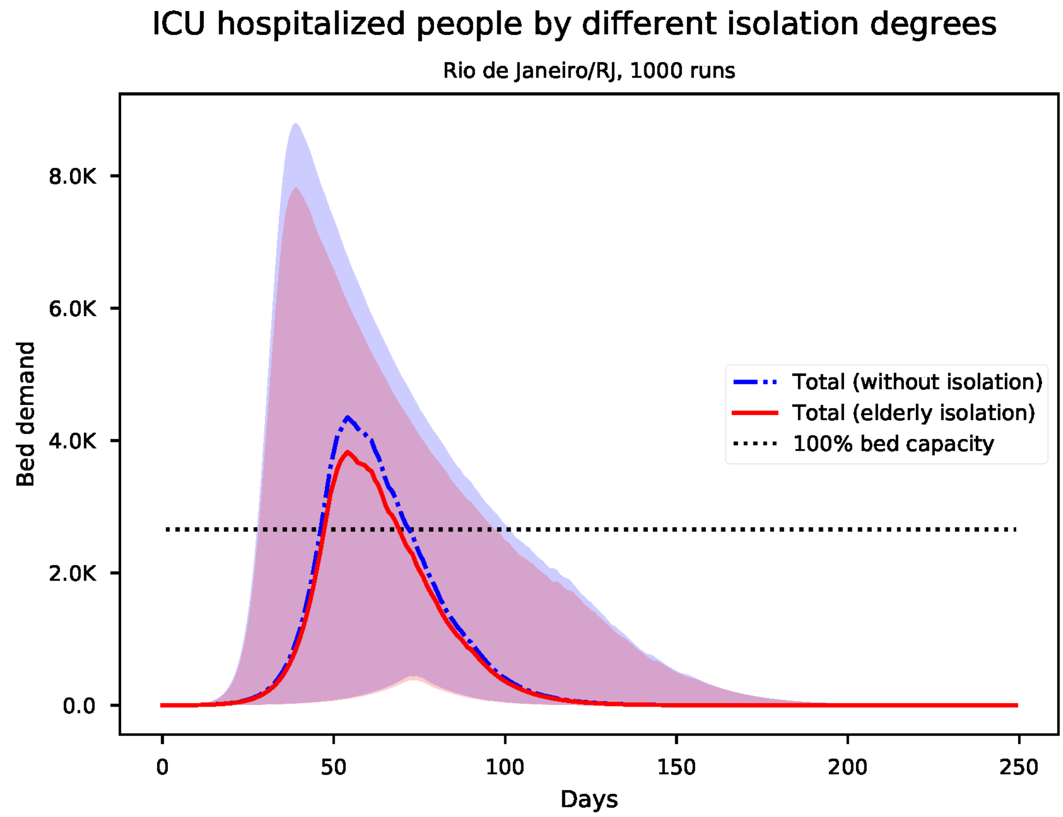


Figure S129: Predicted number of needed ICU beds for Rio de Janeiro

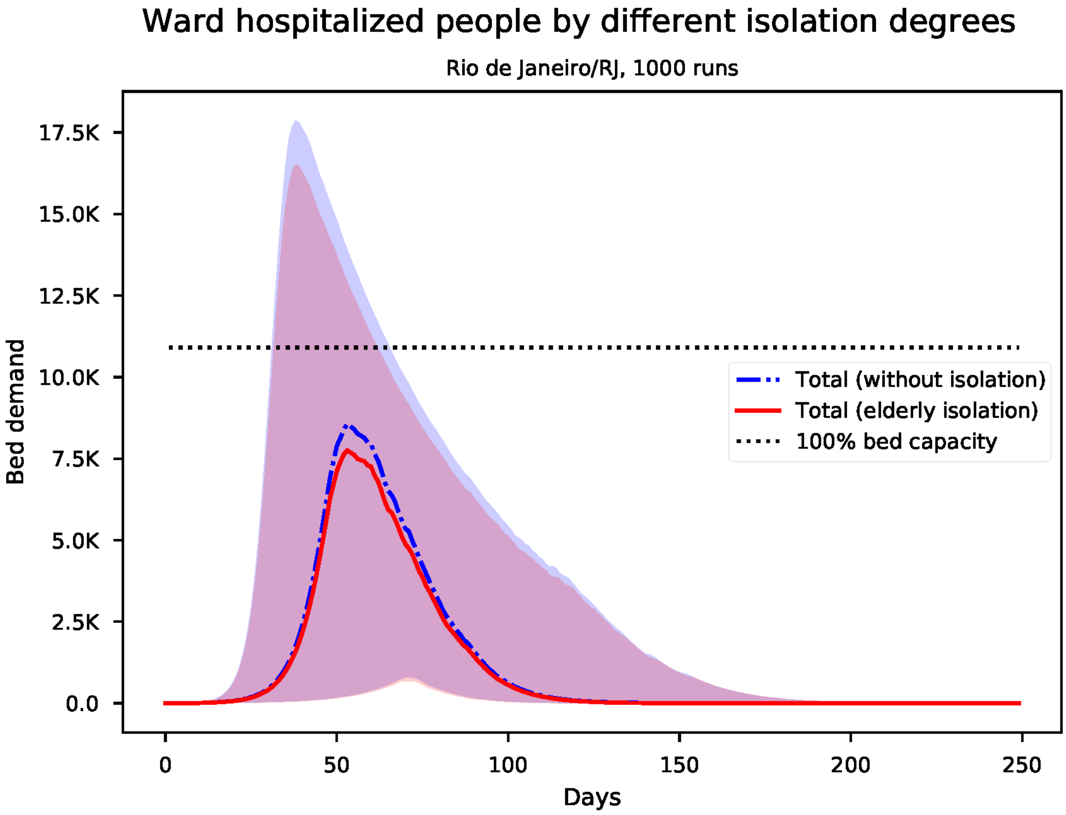


Figure S130: Predicted number of needed ward beds for Rio de Janeiro

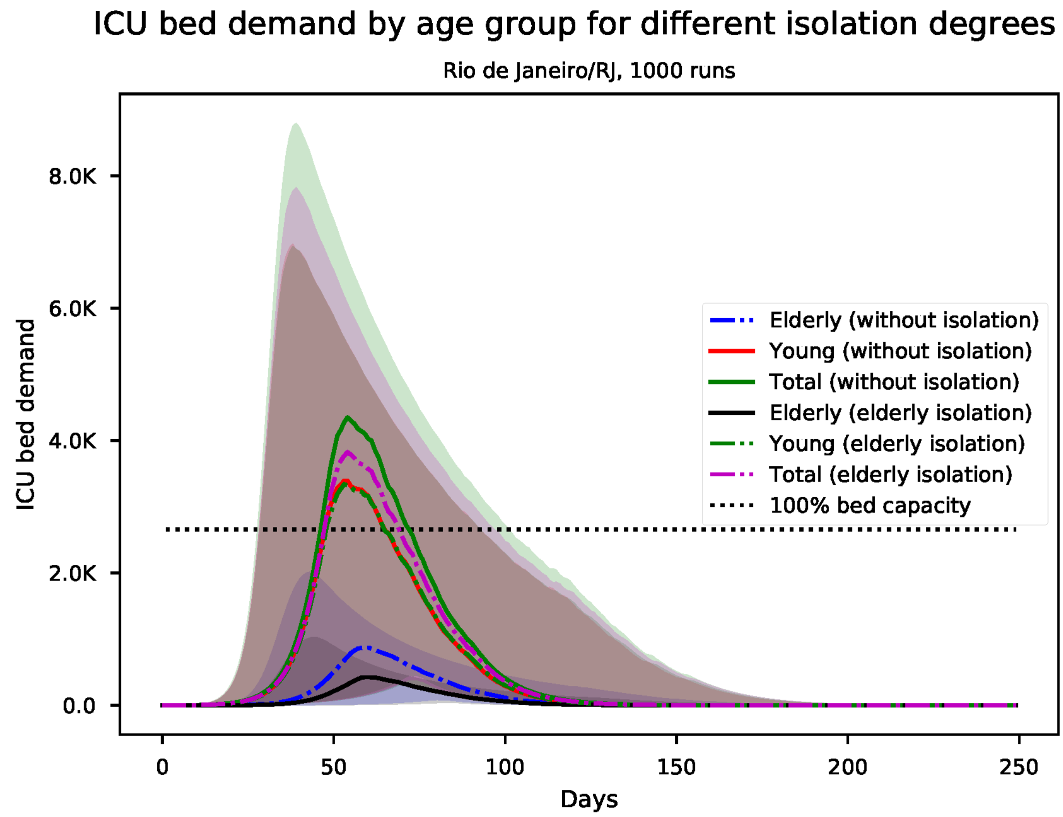


Figure S131: Predicted number of needed ICU beds, by age group for Rio de Janeiro

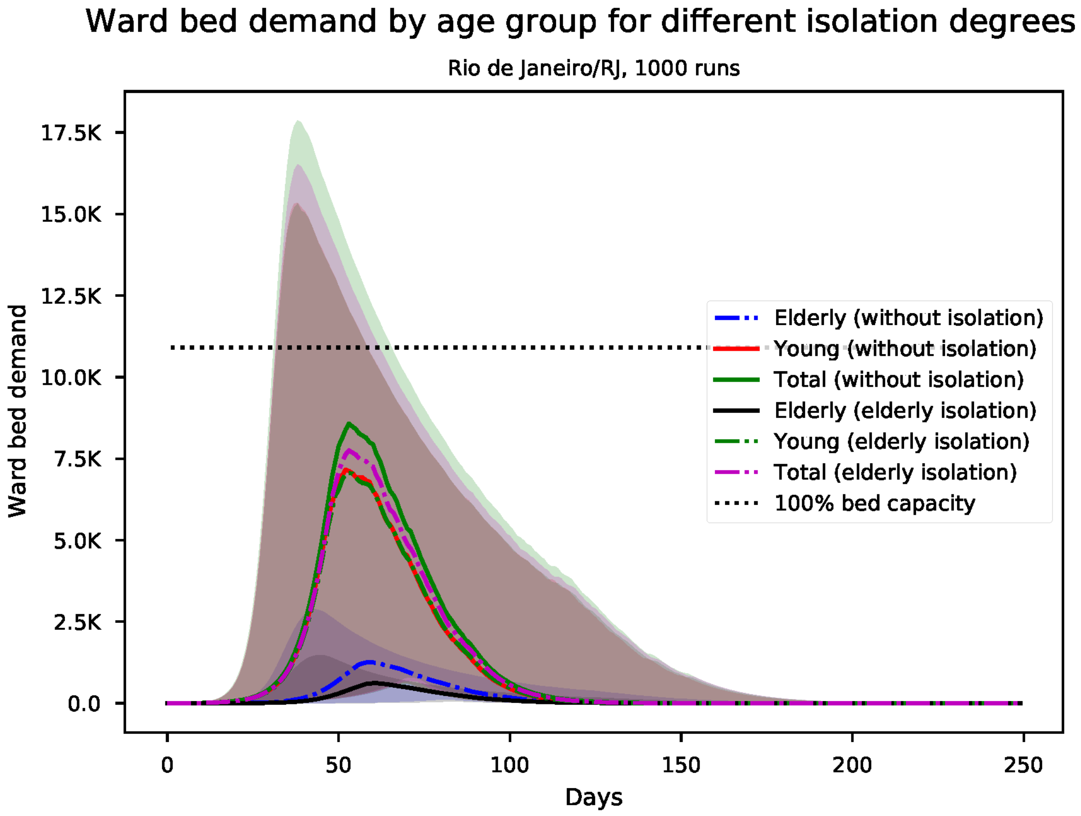


Figure S132: Predicted number of needed ward beds, by age group for Rio de Janeiro

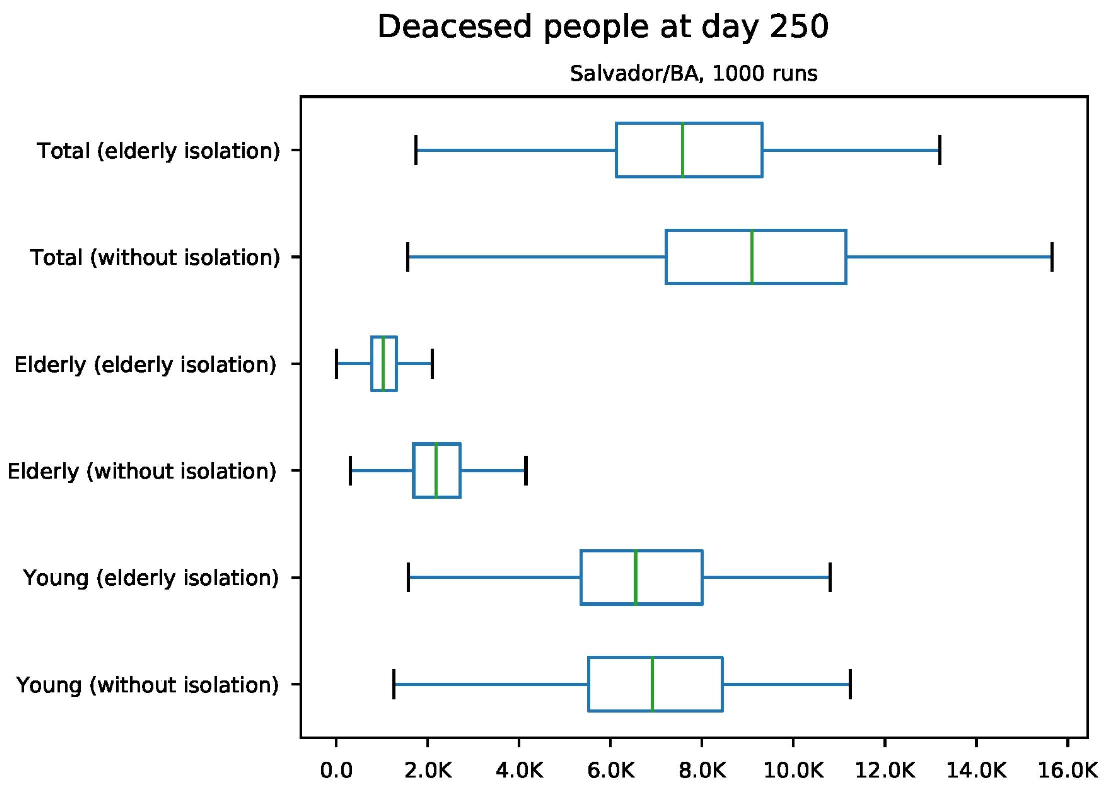


Figure S133: Predicted number of death at day 250 for Salvador

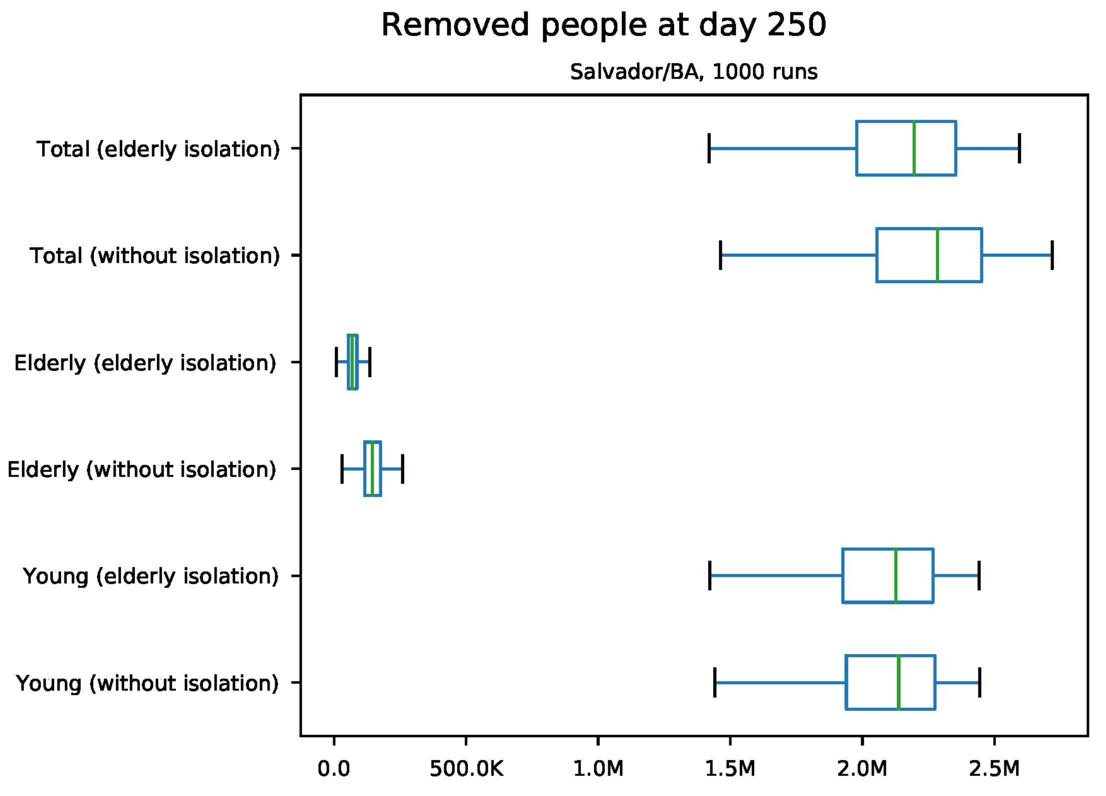


Figure S134: Predicted number of removed individuals at day 250 for Salvador

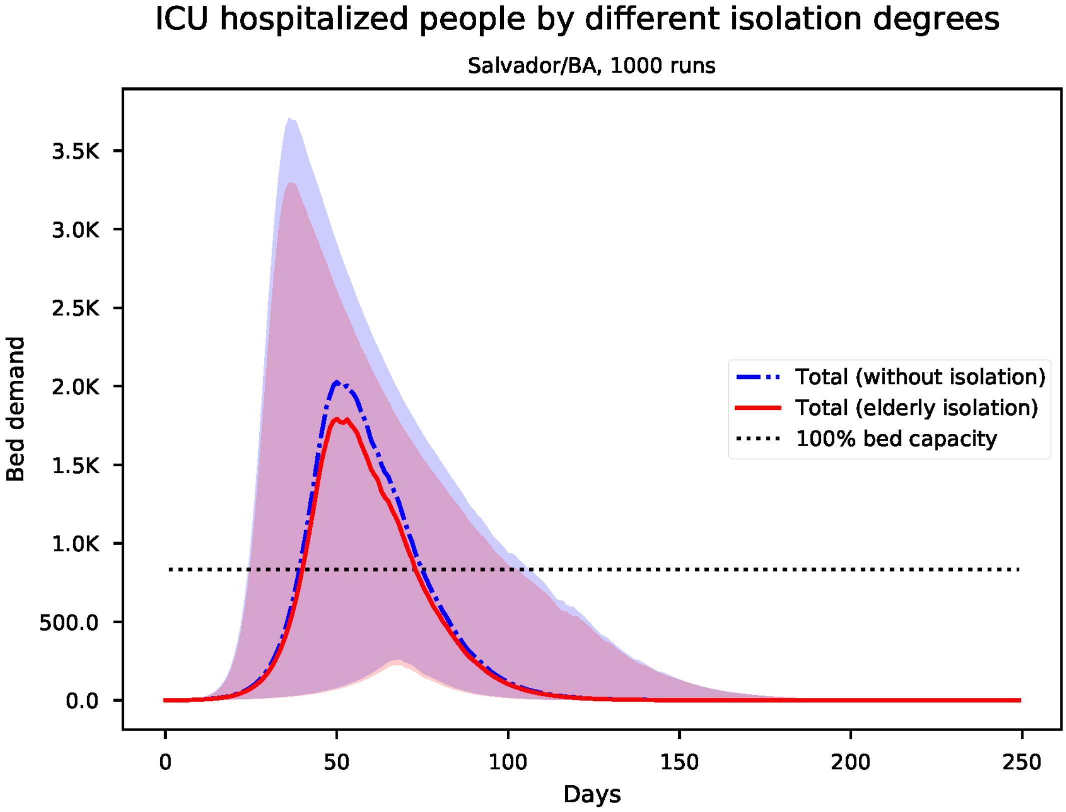


Figure S135: Predicted number of needed ICU beds for Salvador

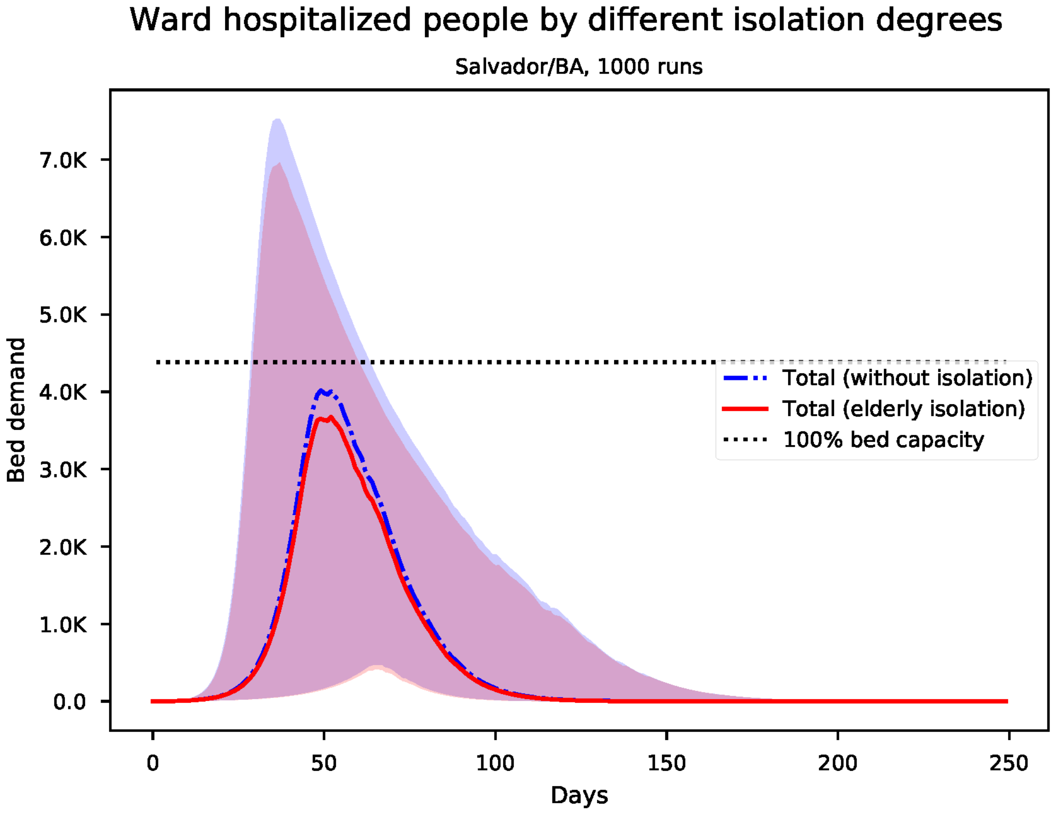


Figure S136: Predicted number of needed ward beds for Salvador

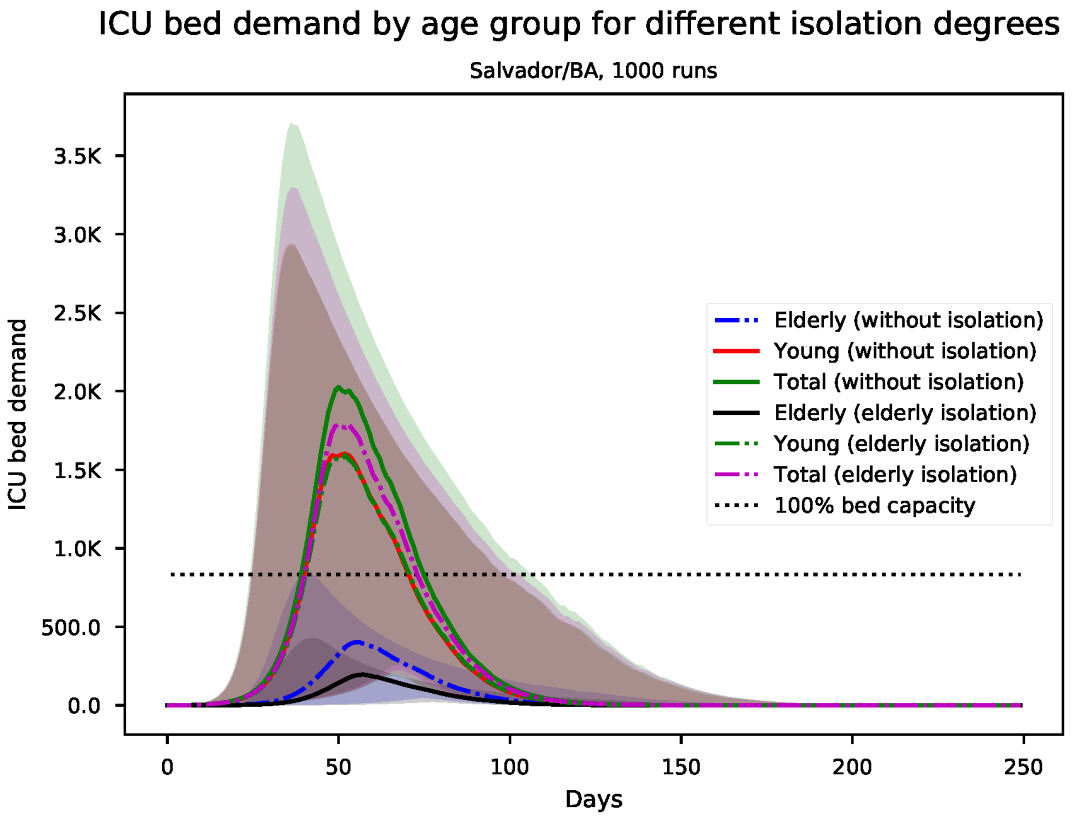


Figure S137: Predicted number of needed ICU beds, by age group for Salvador

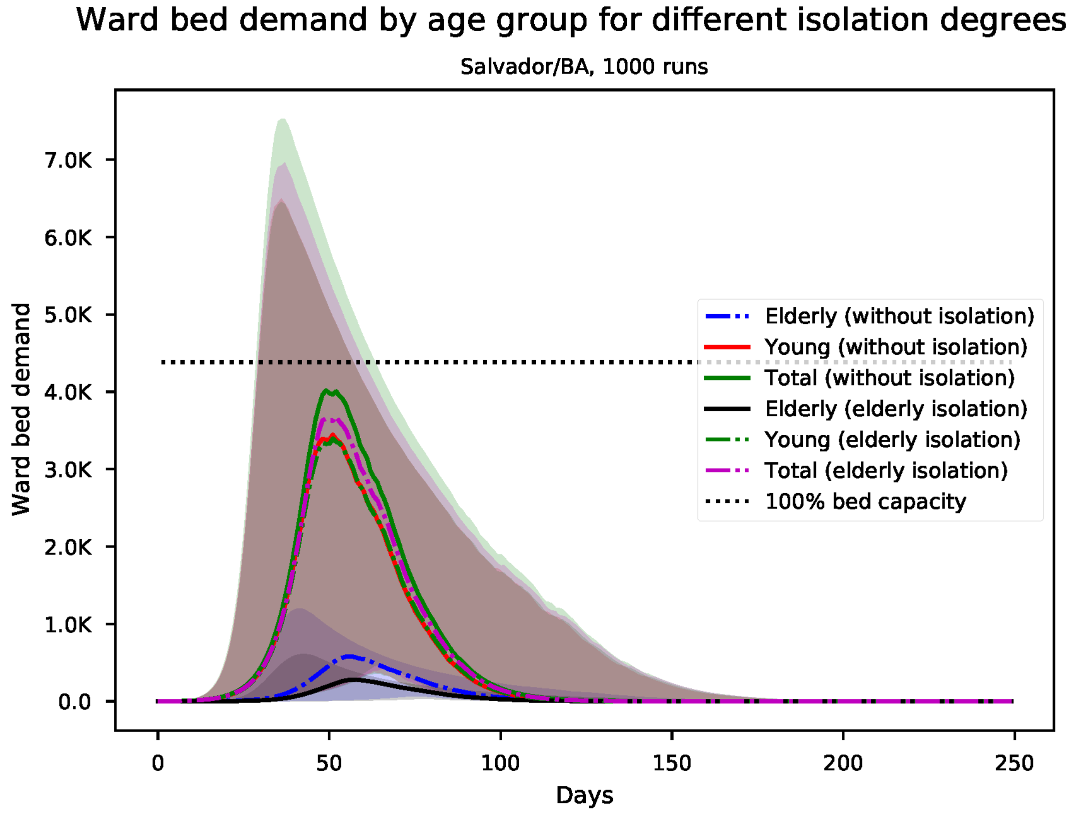


Figure S138: Predicted number of needed ward beds, by age group for Salvador

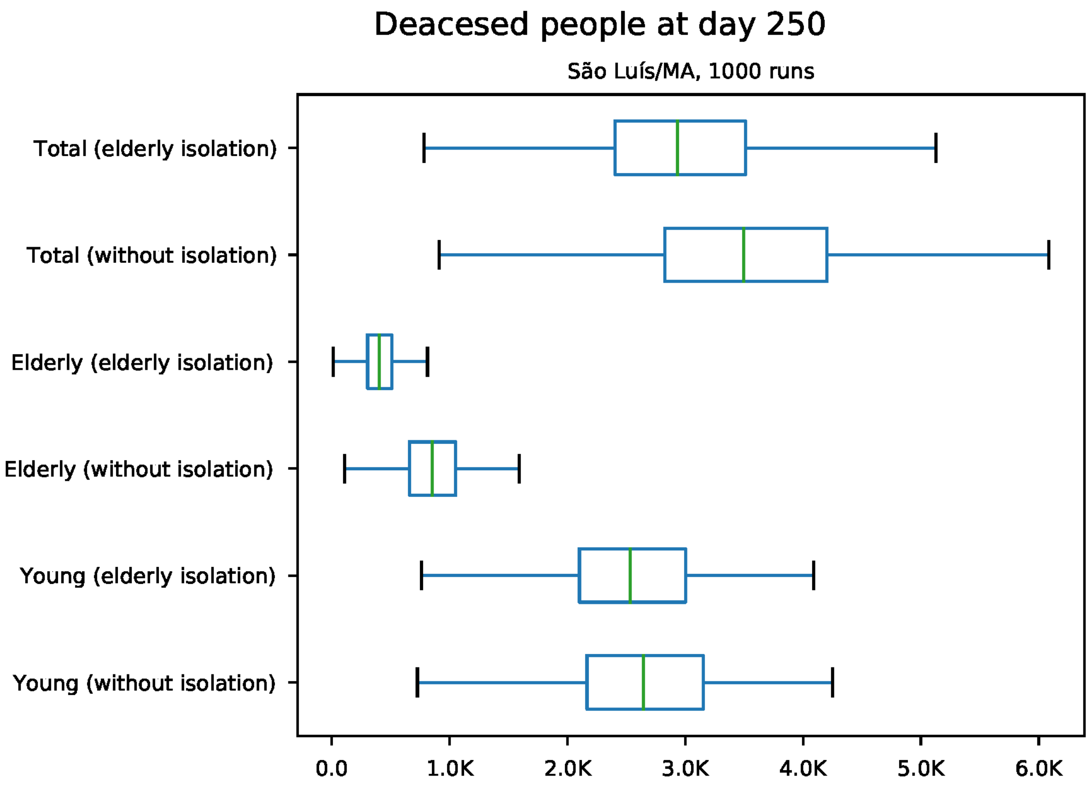


Figure S139: Predicted number of death at day 250 for São Luís

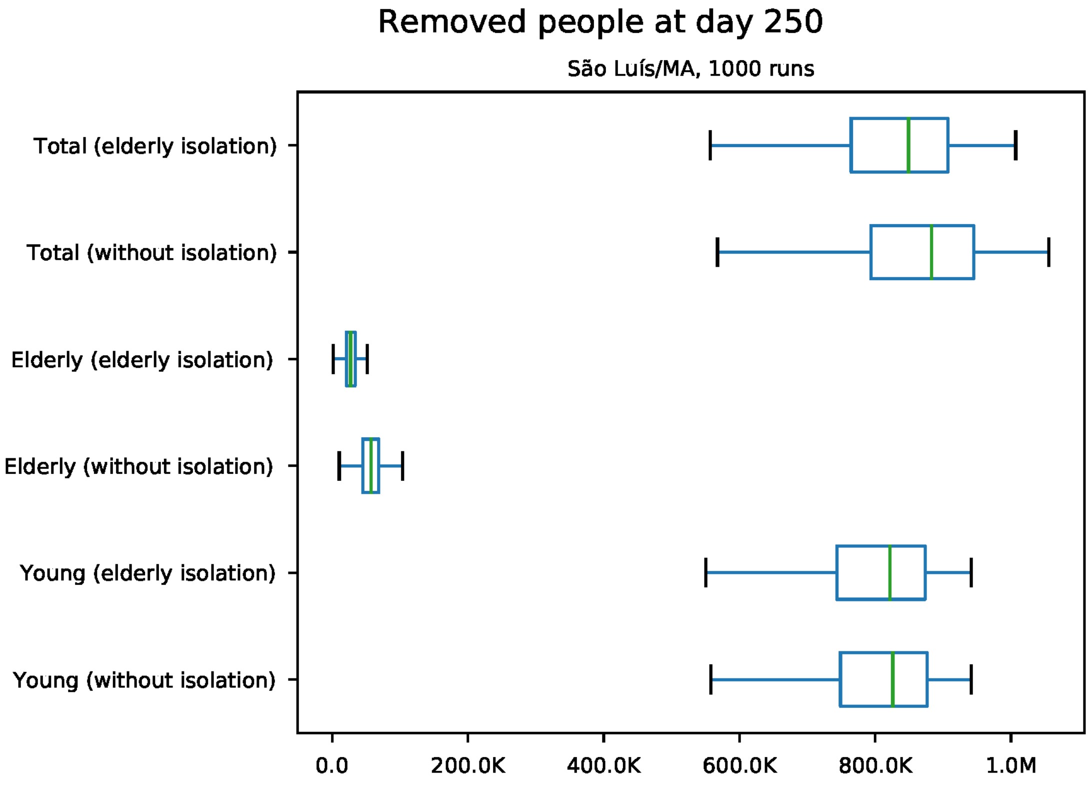


Figure S140: Predicted number of removed individuals at day 250 for São Luís

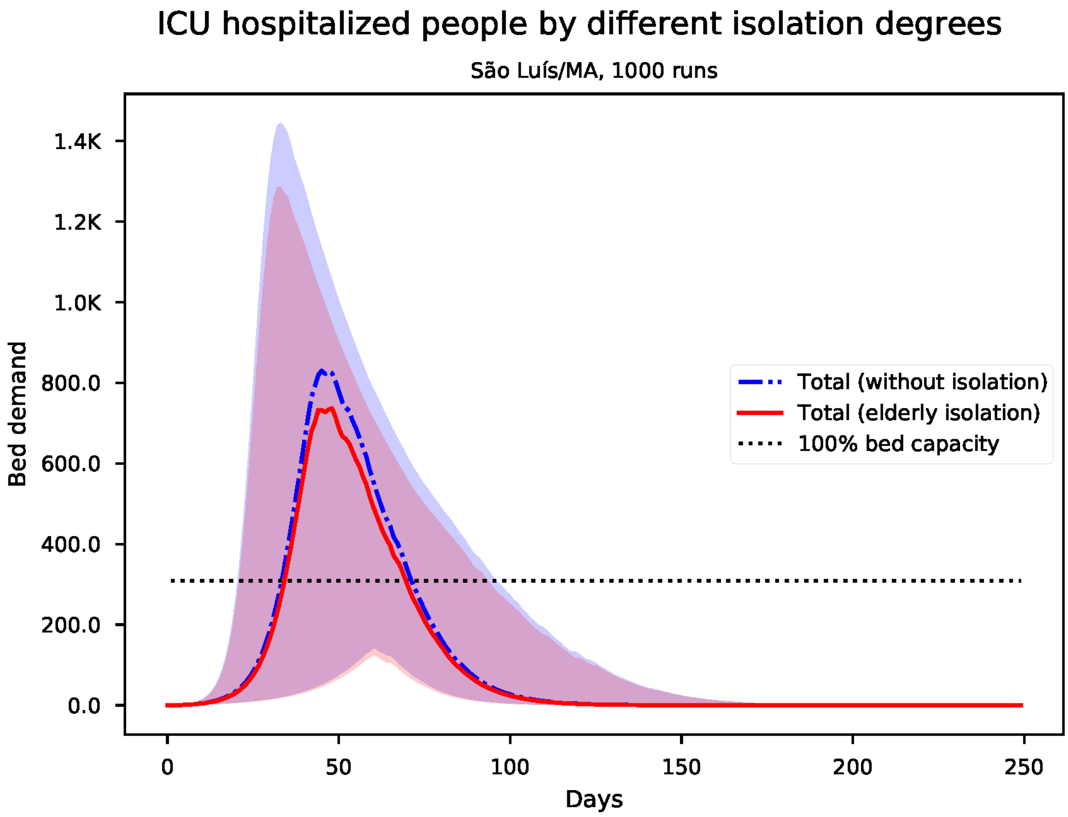


Figure S141: Predicted number of needed ICU beds for São Luís

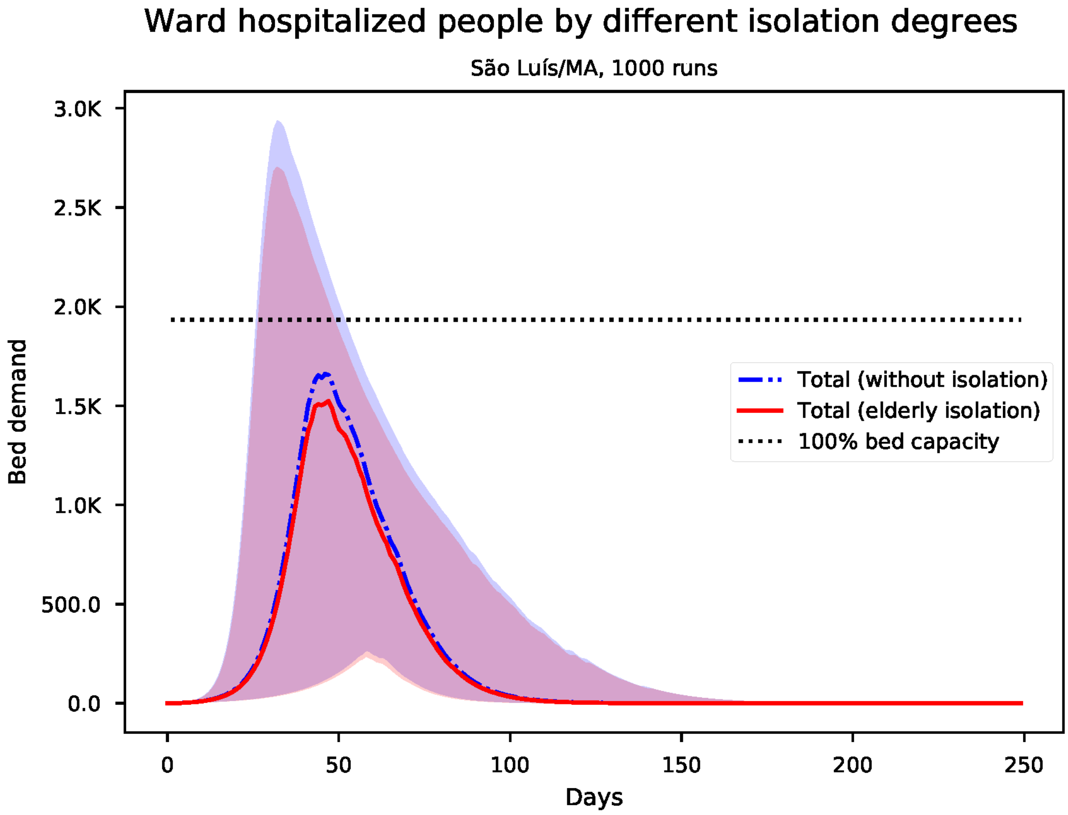


Figure S142: Predicted number of needed ward beds for São Luís

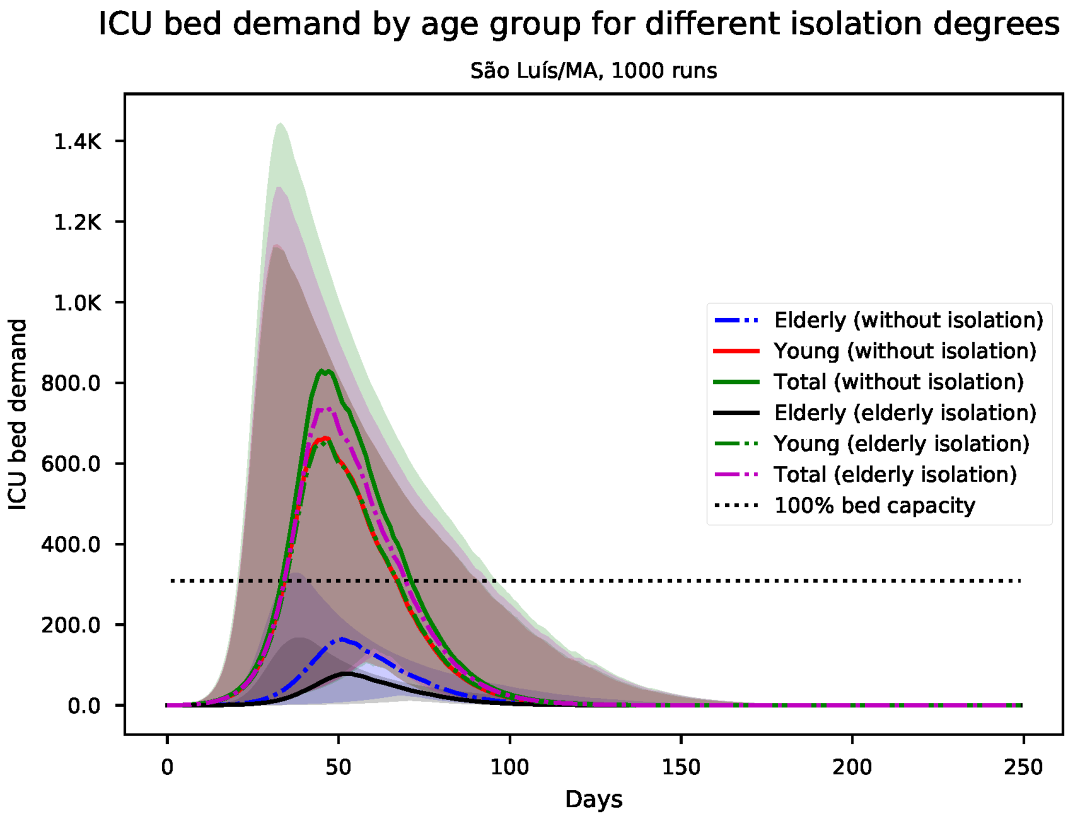


Figure S143: Predicted number of needed ICU beds, by age group for São Luís

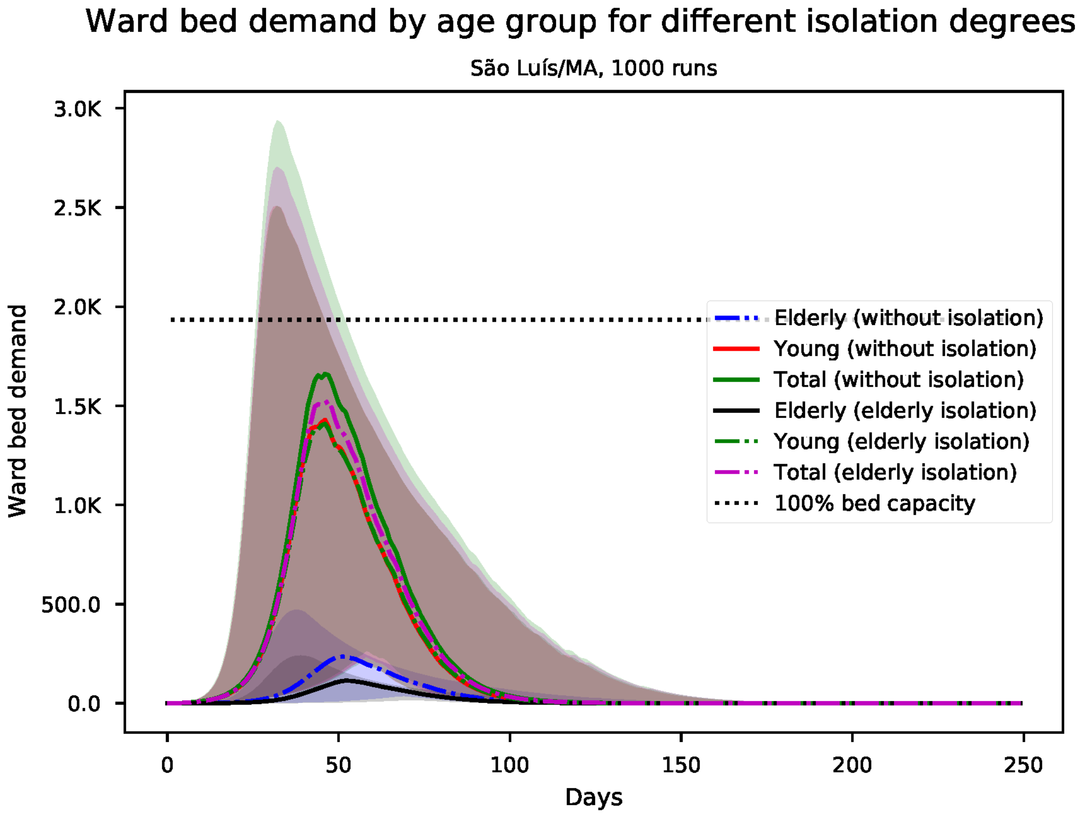


Figure S144: Predicted number of needed ward beds, by age group for São Luís

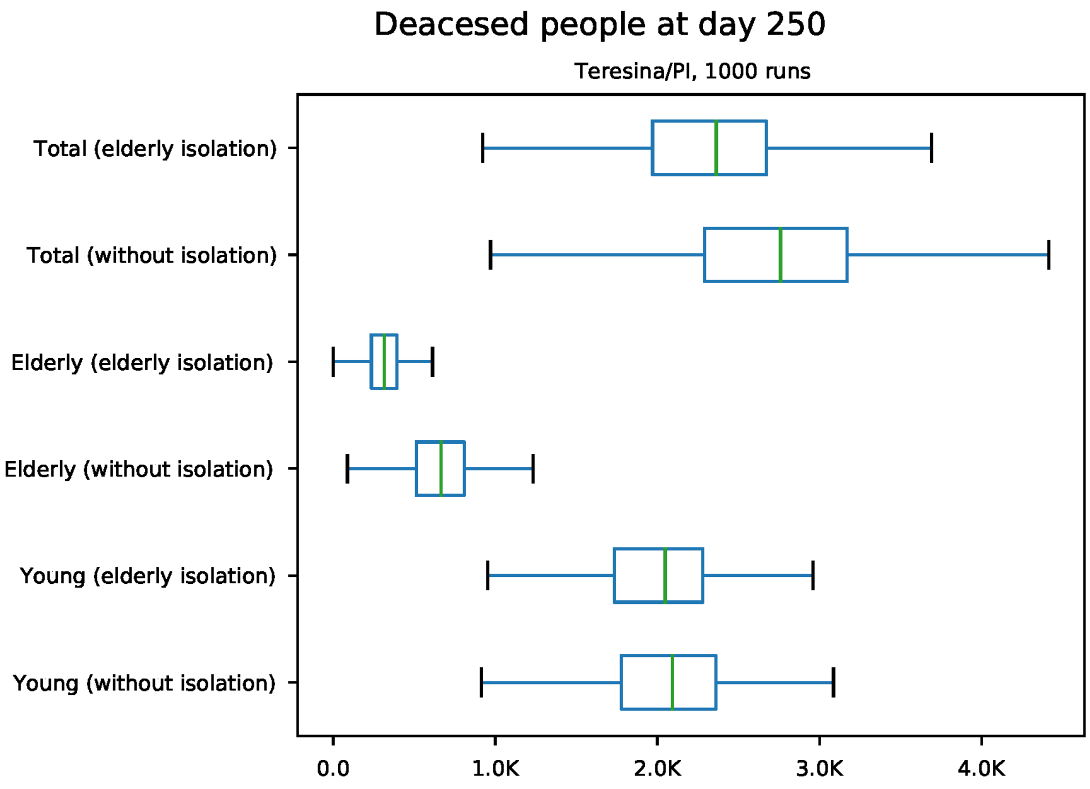


Figure S145: Predicted number of death at day 250 for Teresina

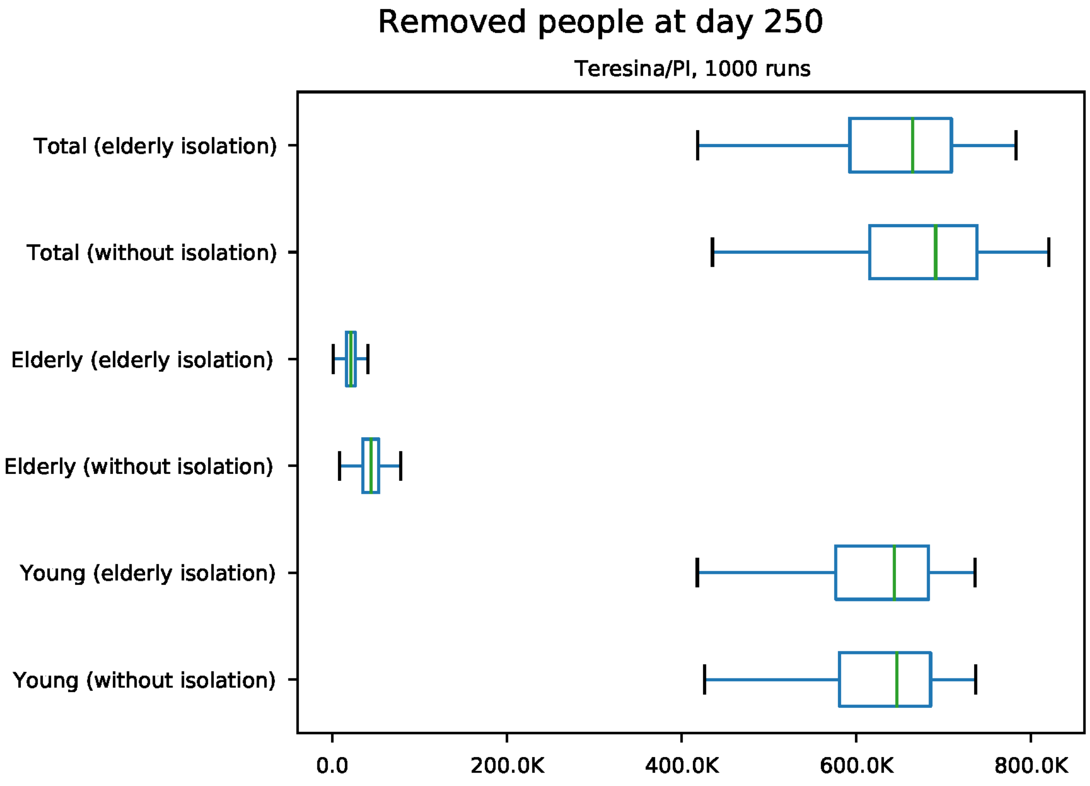


Figure S146: Predicted number of removed individuals at day 250 for Teresina

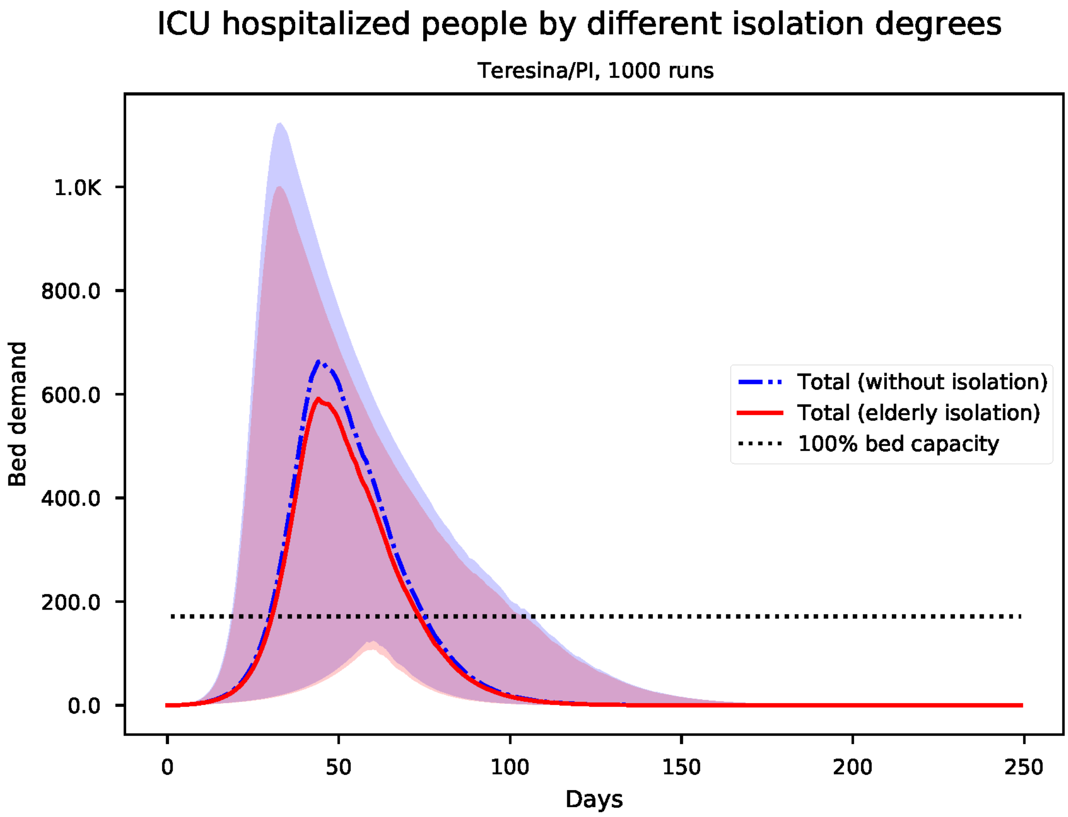


Figure S147: Predicted number of needed ICU beds for Teresina

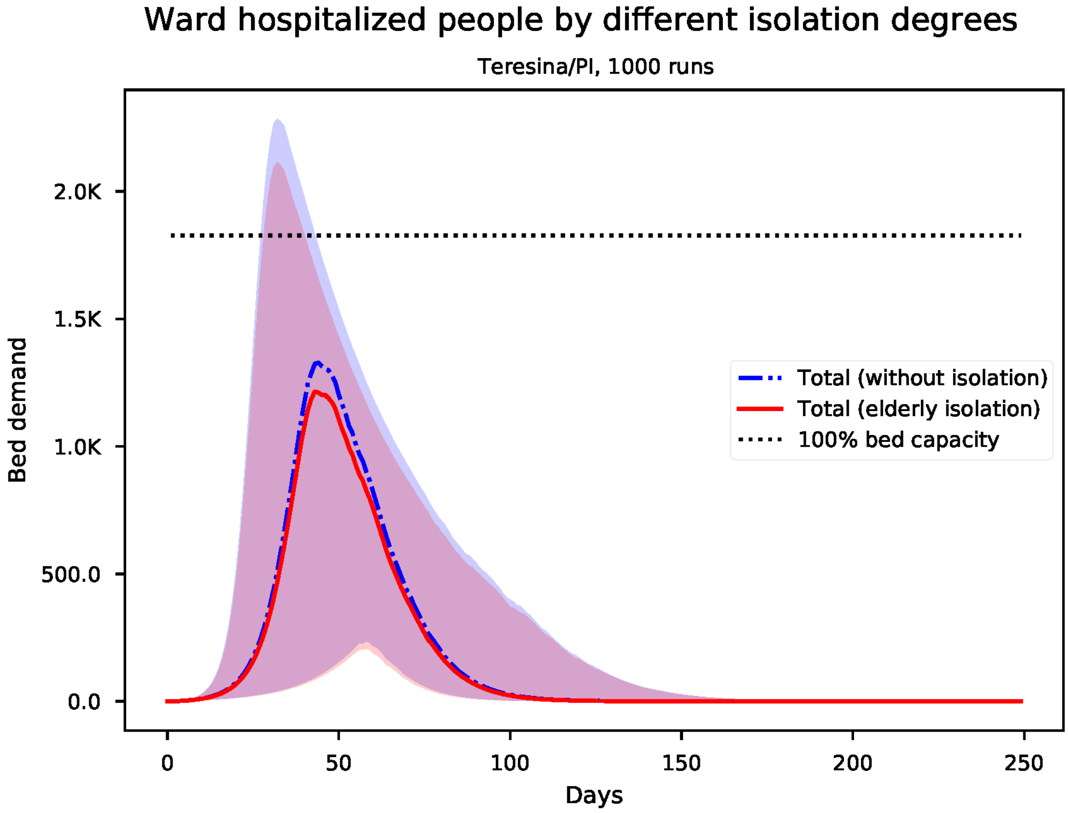


Figure S148: Predicted number of needed ward beds for Teresina

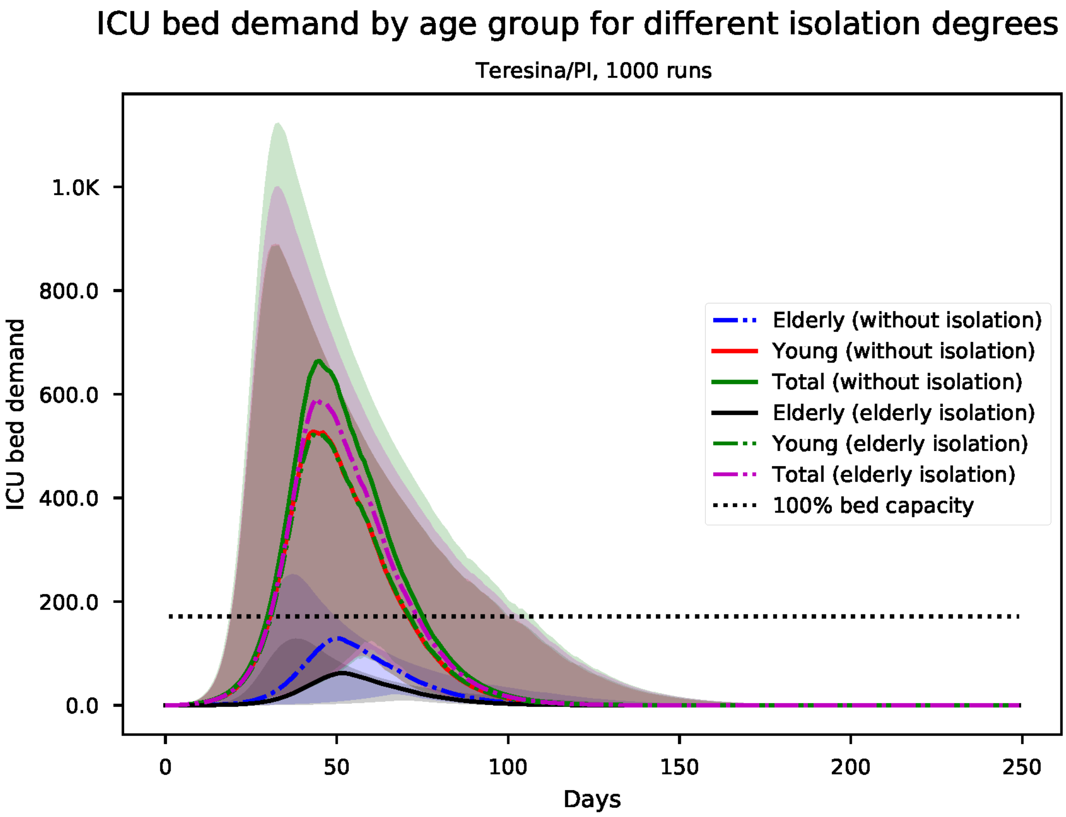


Figure S149: Predicted number of needed ICU beds, by age group for Teresina

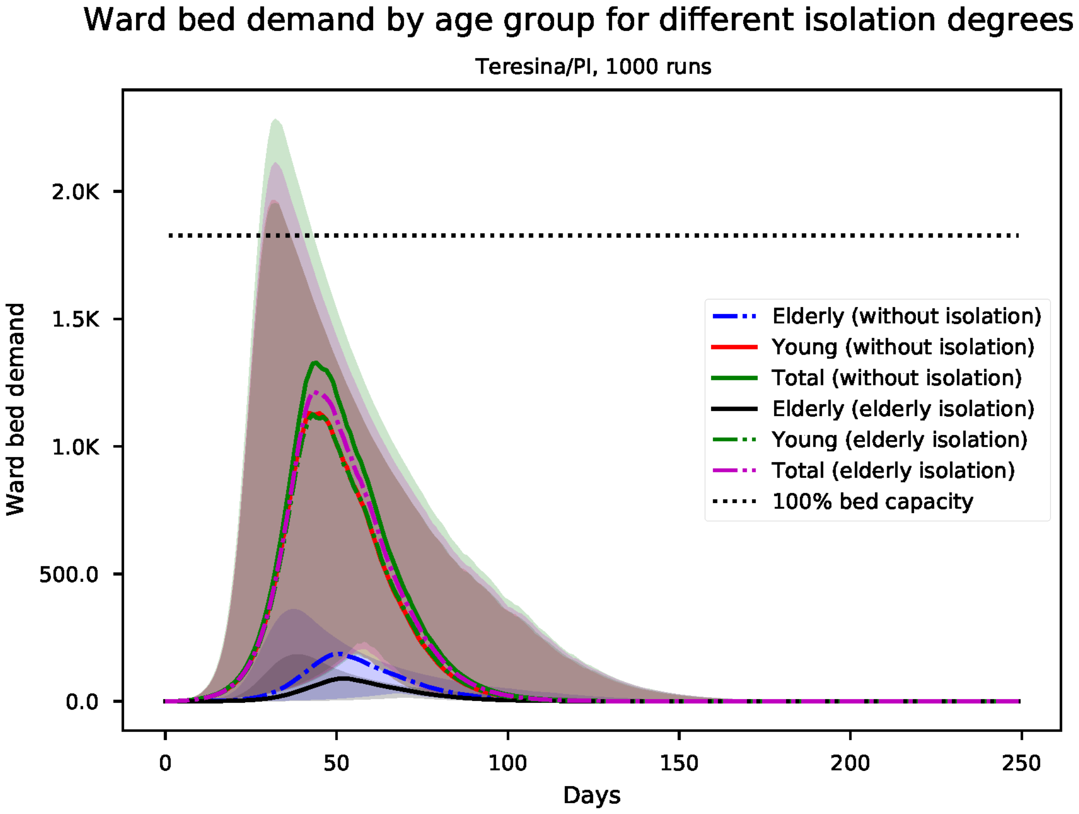


Figure S150: Predicted number of needed ward beds, by age group for Teresina

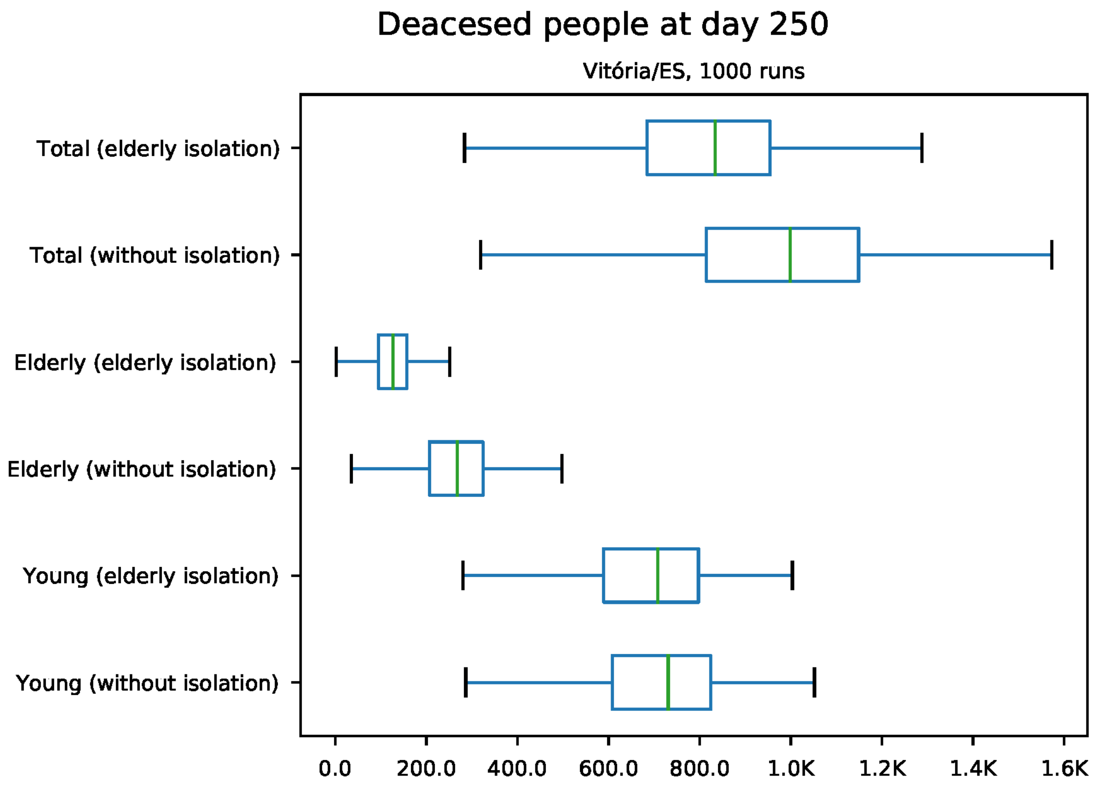


Figure S151: Predicted number of death at day 250 for Vitória

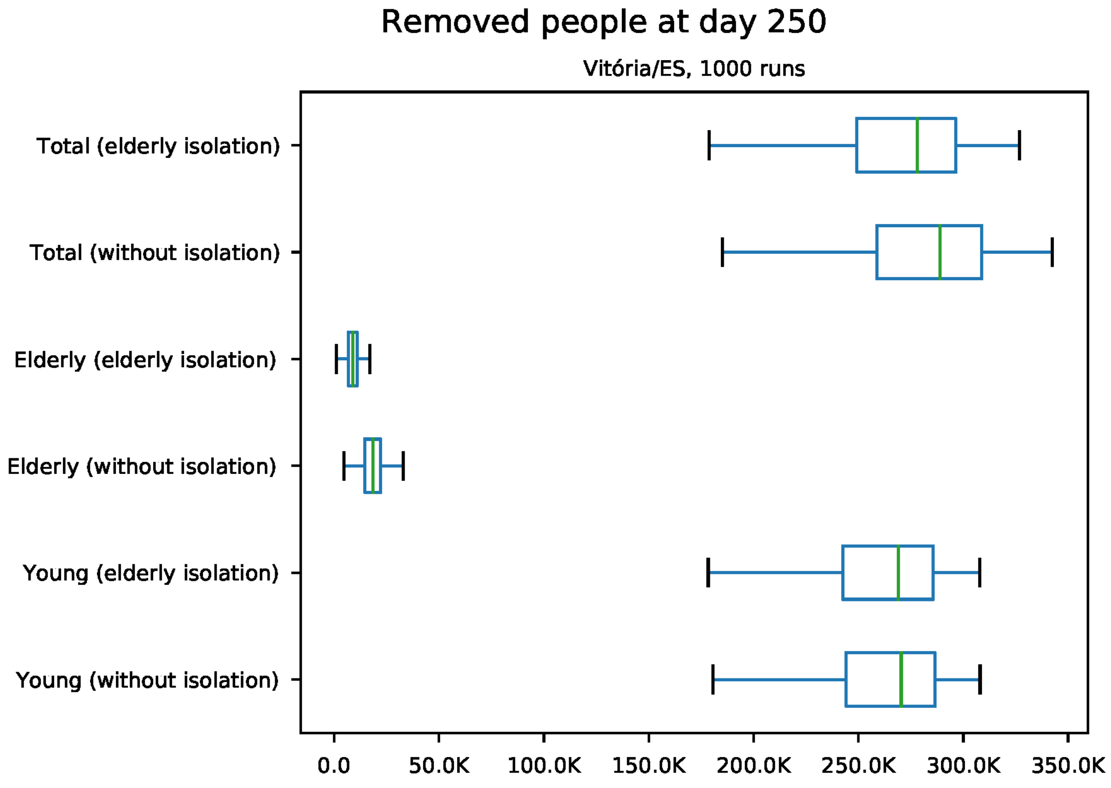


Figure S152: Predicted number of removed individuals at day 250 for Vitória

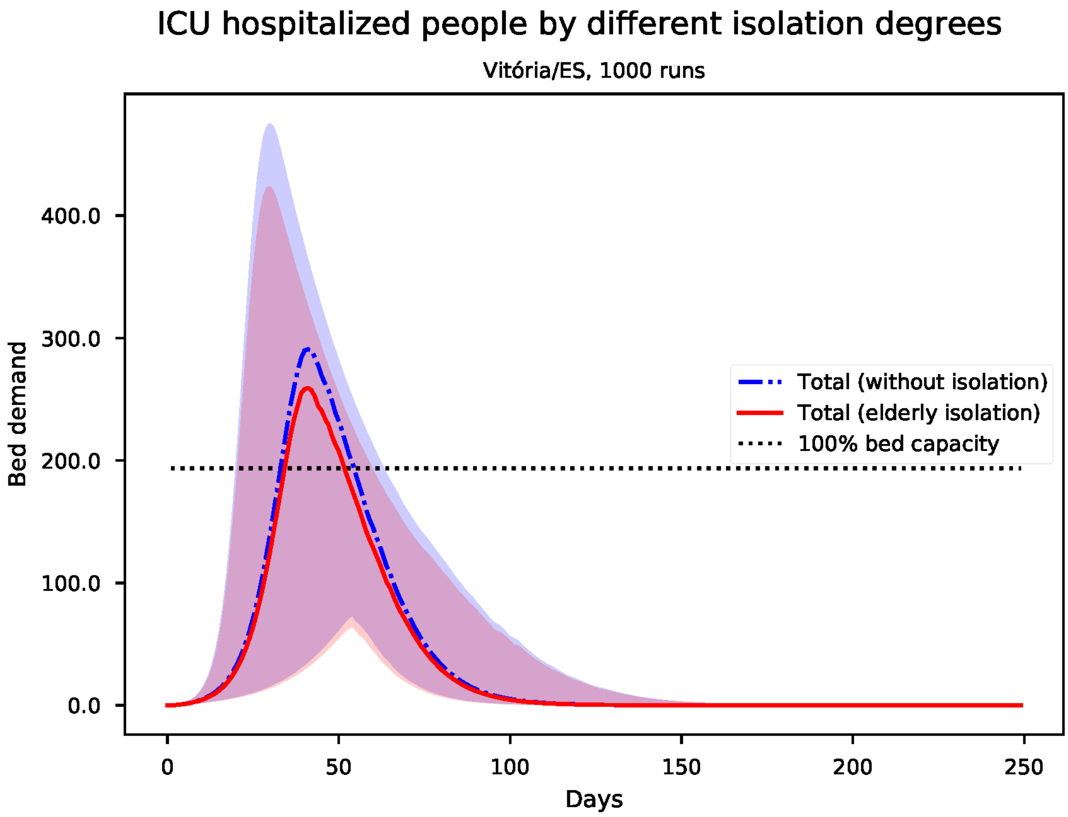


Figure S153: Predicted number of needed ICU beds for Vitória

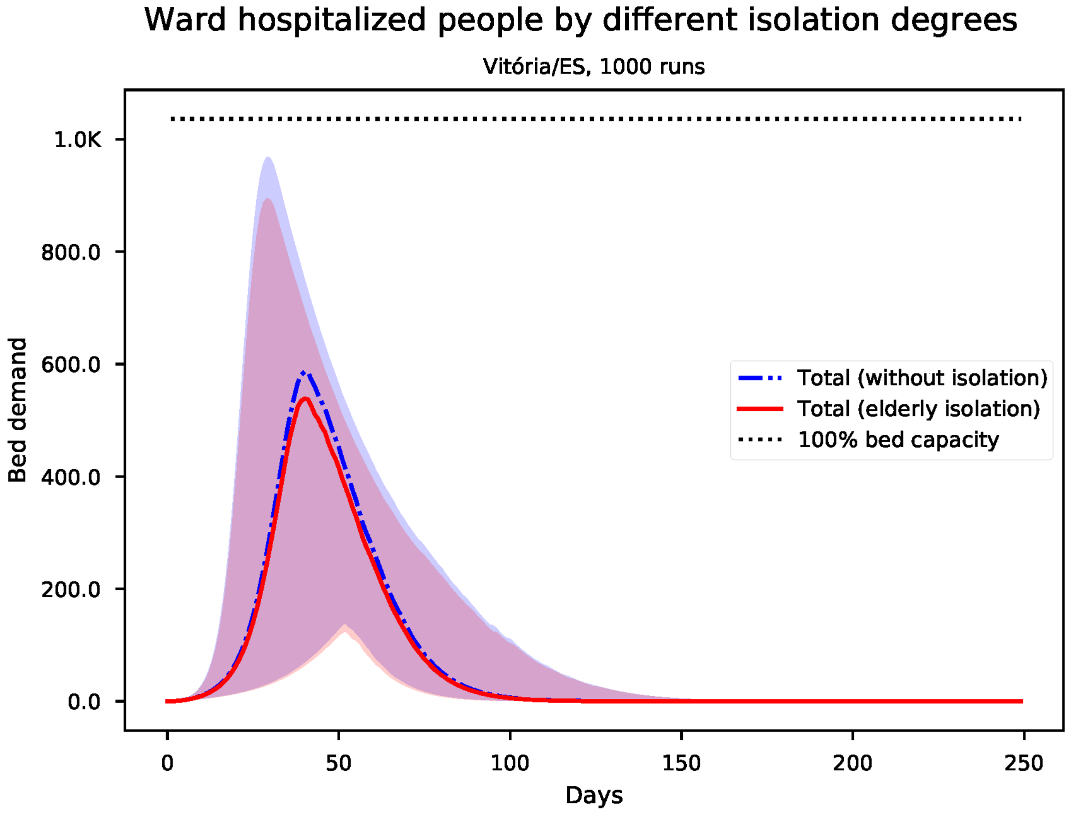


Figure S154: Predicted number of needed ward beds for Vitória

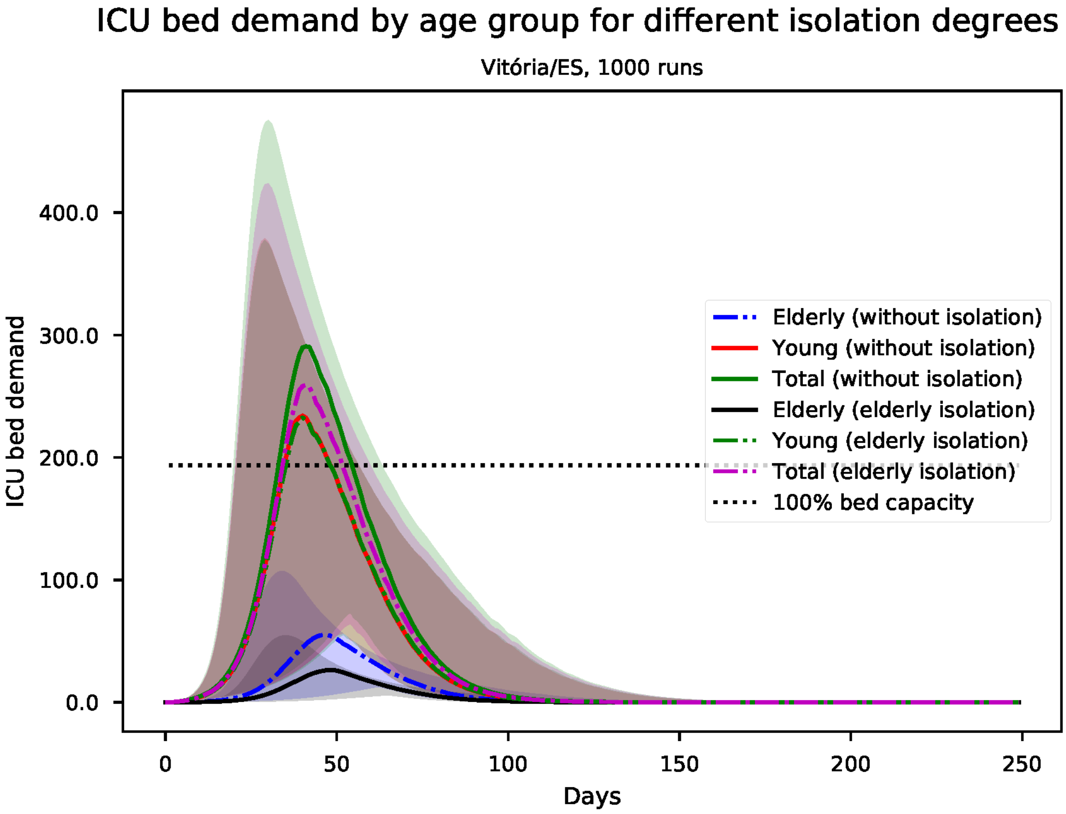


Figure S155: Predicted number of needed ICU beds, by age group for Vitória

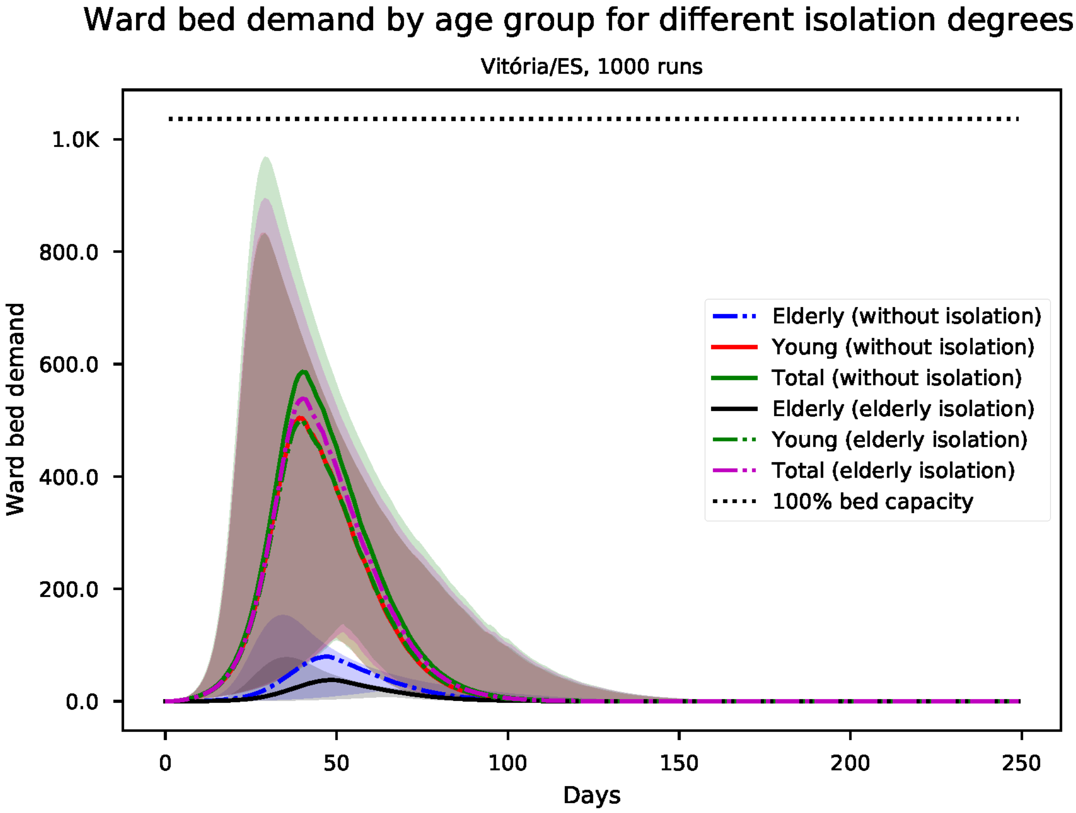


Figure S156: Predicted number of needed ward beds, by age group for Vitória
